# Approaches to improve mental health care for autistic children and young people: a systematic review and meta-analysis

**DOI:** 10.1101/2023.11.30.23299246

**Authors:** Tamara Pemovska, Sofia Loizou, Rebecca Appleton, Debbie Spain, Theodora Stefanidou, Ariana Kular, Ruth Cooper, Anna Greenburgh, Jessica Griffiths, Phoebe Barnett, Una Foye, Helen Baldwin, Matilda Minchin, Gráinne Brady, Katherine R. K. Saunders, Nafiso Ahmed, Robin Jackson, Rachel Rowan Olive, Jennie Parker, Amanda Timmerman, Suzi Sapiets, Eva Driskell, Beverley Chipp, Bethany Parsons, Vaso Totsika, Will Mandy, Richard Pender, Philippa Clery, Brynmor Lloyd-Evans, Alan Simpson, Sonia Johnson

## Abstract

Autistic children and young people (CYP) experience mental health difficulties but face many barriers to accessing and benefiting from mental health care. There is a need to explore strategies in mental health care for autistic CYP to guide clinical practice and future research and support their mental health needs. Our aim was to identify strategies used to improve mental health care for autistic CYP and examine evidence on their acceptability, feasibility, and effectiveness. A systematic review and meta-analysis were carried out. All study designs reporting acceptability/feasibility outcomes and empirical quantitative studies reporting effectiveness outcomes for strategies tested within mental health care were eligible. We conducted a narrative synthesis and separate meta-analyses by informant (self, parent, clinician). Fifty-seven papers were included, with most investigating cognitive behavioural therapy (CBT)-based interventions for anxiety and several exploring service-level strategies, such as autism screening tools, clinician training, adaptations regarding organisation of services. Most papers described caregiver involvement in therapy and reported adaptations to communication and intervention content; a few reported environmental adjustments. In the meta-analyses, parent- and clinician-reported outcomes, but not self-reported outcomes, showed with moderate certainty that CBT for anxiety was an effective treatment compared to any comparison condition in reducing anxiety symptoms in autistic individuals. The certainty of evidence for effectiveness, synthesised narratively, ranged from low to moderate. Evidence for feasibility and acceptability tended to be positive. Many identified strategies are simple, reasonable adjustments that can be implemented in services to enhance mental health care for autistic individuals. Notable research gaps persist, however.

## Introduction

Autism is clinically defined as a neurodevelopmental condition characterised by social communication differences, sensory sensitivities, and difficulties with behavioural and cognitive flexibility (APA, 2013). It is also conceptualised as a form of neurodivergence, representing natural differences in human minds (Chapman & Botha, 2023). Autism prevalence is estimated to be 1-2% of the global population (Lai, Lombardo, & Baron-Cohen, 2014; Roman-Urrestarazu et al., 2021). However, complex referral pathways and lengthy waits for diagnostic assessment often translate into untimely or incorrect diagnosis (NHSE, 2023), probably impacting the accuracy of prevalence estimates.

Autistic children and young people (CYP) experience high rates of co-occurring mental health difficulties (Simonoff et al., 2008), contributing to considerable long-term negative effects on health and quality of life (Lai et al., 2019). An increasing body of research is highlighting the impact mental health difficulties can have on various aspects of life, including education, quality of life, behaviour, family, work, and independence beyond what is linked to autism (Adams, Clark, & Keen, 2019; Adams & Emerson, 2020, 2021; Adams, Young, Simpson, & Keen, 2019; Den Houting, Adams, Roberts, & Keen, 2020; Robertson et al., 2018). Disentangling mental health difficulties from autistic traits can be difficult due to poor clinician knowledge of autism, diagnostic overshadowing, and a lack of validated measures, resulting in challenges and delays to diagnosis and, subsequently, a lack of or ineffective mental health support (Adams & Young, 2021; Brede et al., 2022; Hus & Segal, 2021; Maddox et al., 2020). There is preliminary evidence for the feasibility and effectiveness of standard and adapted psychological interventions for anxiety and mood-related outcomes for autistic CYP (Linden et al., 2023). Meanwhile, pharmacological interventions trialled in this population have obtained mixed results when prescribed for mental health symptoms (Deb et al., 2021), and clinical guidelines have recommended caution when prescribing them for CYP, especially without concurrent psychological interventions (NICE, 2021).

Mental health care requires tailoring for autistic CYP, as standard care can fail to meet their preferences and needs (Dickson et al., 2021; Lickel, MacLean, Blakeley-Smith, & Hepburn, 2012; NICE, 2021). Mental health services may attempt to address autistic people’s needs through implementing bespoke interventions specifically developed for this population, adapted standard interventions, and/or changes to service delivery overall. Adaptations are needed to make the overall experience of contact with services more accessible and acceptable, as well as to ensure that the structure, delivery, and content of interventions is appropriate for autistic young people. These adaptations should also be in line with the person’s developmental age and stage (NICE, 2021). Adaptations that have been recommended include offering shorter or longer appointments, incorporating visual means to facilitate discussion, and changing the physical environment to accommodate sensory preferences (National Autistic Society, 2021). However, parents often report lack of clinician knowledge/expertise regarding autism and an inability of mental health services and clinicians to tailor their support to autistic CYP (Adams & Young, 2021). Failure to embed adaptations can result in distress, disengagement from services and reduced help-seeking (Benevides et al., 2020; Brede et al., 2022; Crane, Adams, Harper, Welch, & Pellicano, 2018). This can negatively impact the wellbeing of families as well as of CYP, increasing carer stress (Read & Schofield, 2010).

More research is needed to explore strategies used in mental health care settings for autistic CYP to guide clinical practice and future research in this area so that needs for effective mental health care can be better met. Thus, this systematic review aimed to identify and examine strategies used to improve mental health care for autistic CYP and, if possible, conduct a meta-analysis, addressing the following research questions:

1. What strategies, including service adaptations, initiatives to detect autism, and bespoke and adapted interventions, have been used to improve mental health care for autistic CYP?
2. What is the acceptability and feasibility of strategies to improve mental health care for autistic CYP?
3. What is the effectiveness of strategies to improve mental health care for autistic CYP?

## Methods

This systematic review was conducted by the National Institute for Health and Care Research (NIHR) Mental Health Policy Research Unit, as part of their research programme aimed at building evidence to inform policy (MHPRU, n.d.). The protocol, developed in collaboration with a working group, comprising lived experience researchers, academics, clinicians, and policy experts with personal/professional expertise of autism and/or review methodology, was pre-registered on PROSPERO (CRD42022347690). We followed the PRISMA guidelines (Page et al., 2021). See Table S1 for a PRISMA checklist.

This systematic review reports the findings regarding autistic CYP and mixed samples of adults and CYP when only combined outcomes were available. A separate systematic review was conducted regarding autistic adults (Loizou et al., 2023).

### Search strategy

A systematic literature search using keywords and subject headings relating to autism and mental health problems and services/treatments was conducted in three electronic databases (Medline, PsycINFO, CINHAL) and two pre-print servers (medRxiv and PsyArXiv) for papers published between 1994-July 2022. The date range was chosen to cover the Diagnostic and Statistical Manual of Mental Disorders fourth (DSM-IV) and fifth (DSM-5) edition periods, in line with International Classification of Diseases 10^th^ and 11^th^ edition (ICD-10/11). We searched for additional eligible papers through checking the reference lists of identified relevant systematic reviews and a call for evidence from experts including academics and lived experience networks. Tables S2-S4 present the full search strategy.

### Screening

The selection strategy was piloted, and reviewers conducted the title and abstract screening, using Rayyan (Ouzzani, Hammady, Fedorowicz, & Elmagarmid, 2016), with a random 10% of records independently reviewed in duplicate (97.98% agreement). Full texts were screened independently in duplicate in line with Cochrane guidance (J. P. T. Higgins & Thomas, 2023). Discrepancies were resolved by discussion with a third reviewer and the working group.

### Eligibility criteria

#### Population

Papers eligible for inclusion included CYP or mixed samples of CYP and adults (aged 18+) where data from autistic CYP could not be disentangled. Participants with an autism diagnosis or who suspected they were autistic or were identified by clinicians as potentially autistic were eligible. Views of carers and clinicians about mental health interventions for autistic CYP were also eligible. Papers with samples including both autistic and non-autistic people were excluded, unless data from autistic people could be isolated, or papers explored detection of autism.

#### Strategies

We included papers that assessed any strategies/adaptations to improve mental health care for autistic CYP, including: 1) bespoke mental health interventions originally developed for autistic people, 2) adaptations to existing mental health interventions, and 3) service-level strategies (e.g., strategies to detect autism) within mental health services and/or in mental health care delivered in primary care. Authors were contacted if the setting or the intervention’s eligibility and classification as adapted/bespoke were unclear. Papers were eligible regardless of the presence and type of comparison group.

#### Outcomes

Eligible outcomes were any quantitative or qualitative measure of feasibility (e.g., recruitment adherence, retention rates), service use (e.g., engagement), acceptability of care, experience of and satisfaction with care, and/or quantitative measure of mental health, detection of autism, quality of life, service use, and social outcomes (e.g., social functioning) at end of treatment or follow-up. Papers measuring only physical health outcomes were excluded.

#### Study types

All study designs and service evaluations were eligible for the first and second research question, and only empirical quantitative studies were eligible for the third research question. Reviews, case studies without group analysis, commentaries, book chapters, editorials, letters, and conference abstracts were excluded.

### Data extraction

Reviewers extracted data including study design and aims, setting, sample size, participant characteristics (e.g., age, ethnicity, diagnosis), outcome measures, strategies and adaptations (e.g., type, brief description, parent/carer involvement), and relevant findings (feasibility, acceptability, effectiveness). The data extraction form was first piloted on 10% of the eligible papers, discussed with the working group and updated accordingly. The extracted data were checked by at least one other reviewer, thus at least two reviewers reached consensus of the extracted information. Two researchers independently double-extracted raw end-of-treatment (EOT) outcome data (mean, standard deviation, sample size per group) for the meta-analyses.

### Autism-Inclusive Research Assessment

Lived experience researchers in the working group observed that relevant studies might not have been sufficiently inclusive of autistic experiences (e.g., allowing non-verbal communication, using straightforward language, using measures valid for autistic people). Therefore, a lived experience researcher (RRO) developed criteria derived from existing literature and personal experience, labelled the Autism-Inclusive Research Assessment (AIRA), to measure the extent of autism-inclusive practices in research. The criteria were first used in our systematic review regarding autistic adults (Loizou et al., 2023) but were also piloted on papers with CYP in the present review to determine applicability. The five assessment criteria for the AIRA are: 1) reported lived experience involvement in the design, conduct, or write-up of the paper; 2) reported adjustments made to data collection process for papers with qualitative elements (Benford & Standen, 2011); 3) reported adjustments made to data collection tools for papers with quantitative elements (Nicolaidis et al., 2020); 4) reported adaptations or validity of relevant outcome measures for autistic people for papers with quantitative elements; 5) for papers with quantitative elements, perceived inappropriate focus of the tested intervention/strategy on masking/changing autistic traits, which might have not inherently caused distress or worsened quality of life (Chapman & Botha, 2023). Two researchers extracted all relevant data, and a lived experience researcher was involved as second assessor of the final criterion.

### Quality and certainty of available evidence

Reviewers assessed study quality using the Mixed Methods Appraisal Tool (MMAT) (Hong et al., 2018). All scores were checked by a second reviewer and consensus was reached. Reviewers independently double-evaluated the strength of evidence about effectiveness of cognitive behavioural therapy (CBT) for anxiety synthesised via meta-analyses using the Grading of Recommendations Assessment, Development and Evaluation (GRADE) system (Guyatt et al., 2008). The strength of the narratively synthesised effectiveness evidence of all interventions/strategies was double-evaluated using GRADE adapted for narrative synthesis (Murad, Mustafa, Schünemann, Sultan, & Santesso, 2017).

### Evidence synthesis

We conducted a narrative synthesis following Economic and Social Research Council guidelines (Popay et al., 2006). With the input of lived experience researchers, the identified intervention-level and service-level adaptations were grouped into categories and sub-categories according to shared commonalities. This was informed by our previous review relating to autistic adults (Loizou et al., 2023) and refined based on the current included studies.

The included papers were grouped in service-level strategies or interventions to synthesise the extracted outcome data. Service-level strategies were categorised based on their focus. Different interventions were characterised based on type, format, bespoke/adapted therapy, and focus. To distinguish between bespoke and adapted interventions, we relied on authors’ descriptions in the papers or their responses when more clarification was needed. We considered interventions to be bespoke (e.g., Facing Your Fears - FYF) if authors reported they were originally designed for autistic people. Authors themselves were primarily involved in developing these interventions/manuals for their study and they were used unmodified. These were considered bespoke interventions regardless of whether they had been based on mainstream CBT or mindfulness principles. We considered the interventions as adapted if authors reported testing adapted existing interventions not originally designed specifically for autistic people. The same approach was used to classify modified versions of interventions originally designed for autistic people, e.g., changed original mode of delivery for FYF to telehealth delivery or developmentally modified version of FYF for use with adolescents.

The extracted data for the AIRA were synthesised descriptively. The feasibility/acceptability findings were synthesised from all contributing study types. We synthesised the effectiveness findings, placing greater importance on randomised controlled trials (RCTs) and non-randomised controlled trials making contemporaneous comparisons rather than before-and-after comparisons. Upon inspection of the included papers, a meta-analysis was deemed appropriate, as a large subset of pilot RCTs and RCTs appeared to be sufficiently homogenous in outcome, intervention, and population. Three meta-analyses were conducted for ratings respectively by children/care recipients, parents/carers, and clinicians to examine whether bespoke/adapted CBT for anxiety is superior to any control condition (active and non-active) in reducing anxiety symptoms at EOT. Separate analyses were performed, as previous meta-analyses have found differences across raters (Sharma, Hucker, Matthews, Grohmann, & Laws, 2021; Sukhodolsky, Bloch, Panza, & Reichow, 2013).

The R-package ‘metafor’ (Viechtbauer, 2010) was used to calculate the standardised mean difference (SMD), correcting for small sample sizes (Hedges’ *g)* between groups at EOT (42). Effect sizes were significant if *p <* .05, and were tentatively interpreted as small (0.2), medium (0.5) and large (0.8) (Cohen, 1988). Random-effects models were used to account for variability in the average effect size across papers (Hedges, 1992). Heterogeneity was assessed using Cochran’s *Q* (significant if *p <* .05) (Cochran, 1954) and Higgins’ *I* (25% = *low*, 50% = *moderate*, 75% = *high*) (J. P. T. Higgins, Thompson, Deeks, & Altman, 2003).

Sensitivity analysis was performed by removing outliers from the models. Where there were sufficient studies (k>10), meta-regression analyses were conducted to examine the moderating effects of type (adapted, bespoke) and format (individual, group, combined) of CBT on effectiveness. Funnel plots were visually inspected, and Egger’s test (significant if p < .05) (Egger, Smith, Schneider, & Minder, 1997) was conducted to test for publication bias.

## Results

### Study selection

Figure 1 shows the PRISMA flow diagram. In total, 57 papers were eligible for inclusion and a full list of studies excluded at full-text screening with reasons is presented in Table S5.

**Figure 1.**
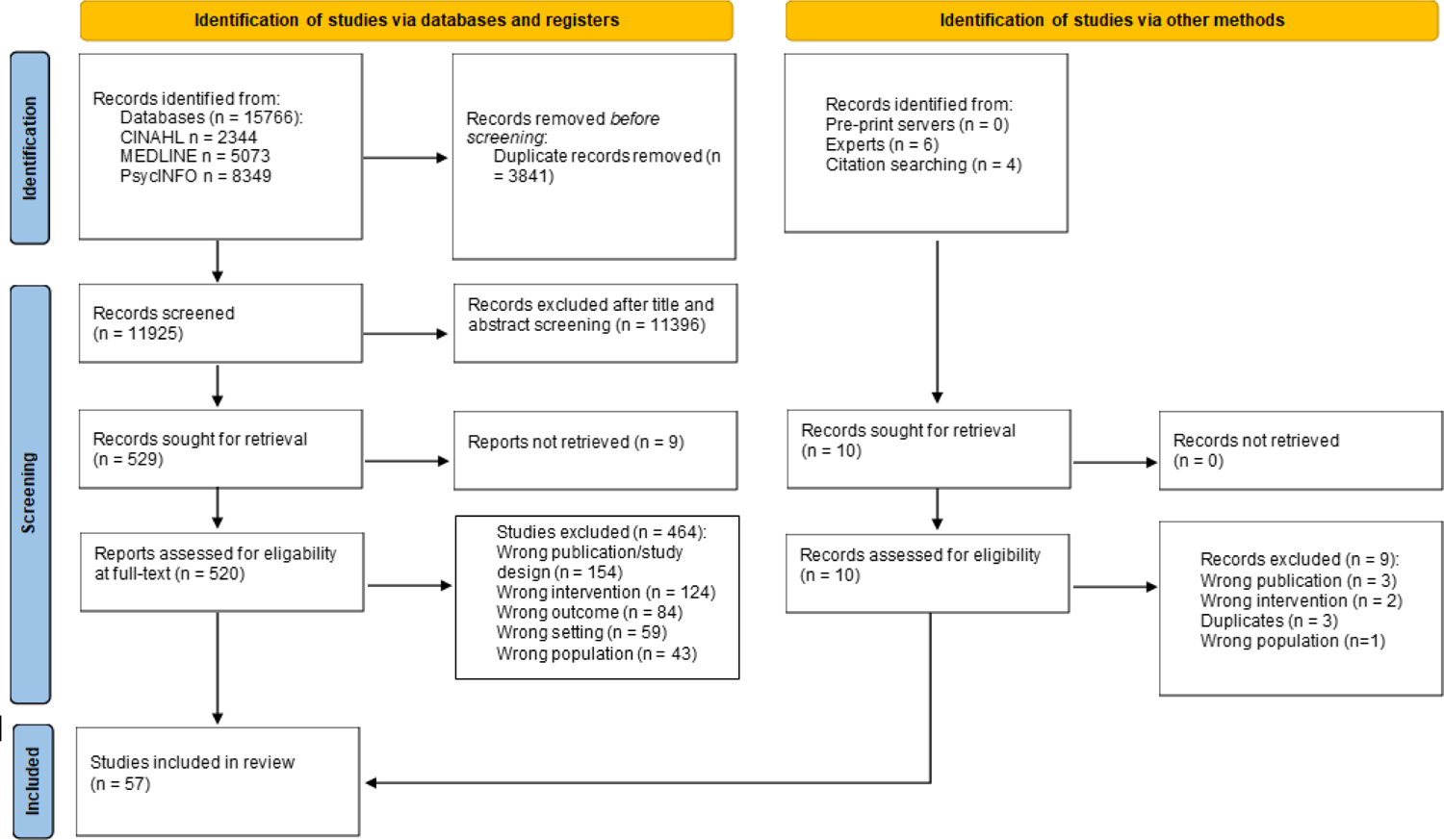
PRISMA flowchart.

### Study characteristics

Of the 57 papers, 23 were RCTs (Chalfant, Rapee, & Carroll, 2007; Cook, Donovan, & Garnett, 2017; Factor et al., 2019; Fujii et al., 2013; Kilburn et al., 2020; Langdon et al., 2016; Maskey, Rodgers, et al., 2019; McConachie et al., 2014; Murphy et al., 2017; J. Reaven, Blakeley-Smith, Culhane-Shelburne, & Hepburn, 2012; J. Reaven et al., 2018; A. J. Russell et al., 2013; Santomauro, Sheffield, & Sofronoff, 2016; Scarpa & Reyes, 2011; Sofronoff, Attwood, & Hinton, 2005; Storch et al., 2013, 2015, 2020; Sung et al., 2011; Walsh et al., 2018; White et al., 2013; White, Schry, Miyazaki, Ollendick, & Scahill, 2015; Wood et al., 2015), of which 11 were pilot RCTs (Cook et al., 2017; Fujii et al., 2013; Langdon et al., 2016; Maskey, Rodgers, et al., 2019; McConachie et al., 2014; Murphy et al., 2017; Santomauro et al., 2016; Scarpa & Reyes, 2011; Storch et al., 2020; White et al., 2013, 2015), three were non-randomised controlled trials (Hepburn, Blakeley-Smith, Wolff, & Reaven, 2016; McGillivray & Evert, 2014; J. A. Reaven et al., 2009), 20 were before-after comparisons (Bemmer et al., 2021; Burke, Prendeville, & Veale, 2017; Dreiling, Cook, Lamarche, & Klinger, 2022; Driscoll, Schonberg, Stark, Carter, & Hirshfeld-Becker, 2020; Drüsedau et al., 2022; Ehrenreich-May et al., 2014; Ekman, Hiltunen, Ekman, & Hiltunen, 2015; Helverschou et al., 2021; E. Higgins, Slattery, Perry, & O’Shea, 2019; Keefer et al., 2017; Kilburn et al., 2019; Maskey, McConachie, et al., 2019; Oerbeck, Overgaard, Attwood, & Bjaastad, 2021; Ollendick, Muskett, Radtke, & Smith, 2021; J. Reaven et al., 2015; J. Reaven, Blakeley-Smith, Leuthe, Moody, & Hepburn, 2012; Sofronoff, Silva, & Beaumont, 2017; Solish et al., 2020; Swain, Murphy, Hassenfeldt, Lorenzi, & Scarpa, 2019; Wise et al., 2019), two papers compared different samples before and after implementation of a new care pathway (Cervantes et al., 2019; Kuriakose et al., 2018), seven were surveys (Cooper, Loades, & Russell, 2018; Fisher, van Diest, Leoni, & Spain, 2023; Ford et al., 2019; Hollocks et al., 2019; Jones, Gangadharan, Brigham, Smith, & Shankar Background, 2021; Pickard et al., 2020; Stadnick, Brookman-Frazee, Nguyen Williams, Cerda, & Akshoomoff, 2015), and two were qualitative (Petty, Bergenheim, Mahoney, & Chamberlain, 2021; Spain et al., 2017). There were multiple papers that were from the same trials (Cervantes et al., 2019; Keefer et al., 2017; Kuriakose et al., 2018; Pickard et al., 2020; J. Reaven et al., 2018; Walsh et al., 2018; White et al., 2013, 2015), thus 57 papers reported on 52 studies. All studies were conducted in high-income countries, mainly in the United Kingdom and United States. Study characteristics are reported in Tables 1 and S6.

**Table 1.** Study characteristics.

| Author (Year) | Country | Study design | Population | Age mean (SD), range | Strategy | Caregiver involvement in therapy | Setting | Condition targeted |
| --- | --- | --- | --- | --- | --- | --- | --- | --- |
| **Detection of autism in mental health care** |  |  |  |  |  |  |  |  |
| Ford et al. (2019) | UK | Prospective cohort study | Autistic CYP | 5-11 years | Detection of autism using the DAWBA | Not applicable | Child and Adolescent Mental Health Services | Autism detection |
| Hollocks et al. (2019) | UK | Survey (Service evaluation) | Autistic CYP | 12.8 years (3.6), 6-19 | Detection of autism using the SCQ | Not applicable | Child and Adolescent Mental Health Services | Autism detection |
| Stadnick et al. (2015) | USA | Cross-sectional survey (service evaluation) | Autistic CYP | 10.69 years (3.48), 5-18 | Detection of autism using the ADOS | Not applicable | Outpatient community-based mental health clinics | Autism detection |
| **Strategies for improving clinician skills and autism knowledge** |  |  |  |  |  |  |  |  |
| Cervantes et al. (2019) | USA | Before-and-after comparison (service implementation) | Autistic CYP | Pre-implementation 35.3% age 4-12 years and 64.7% age 13-17 years; post-implementation 60% age 4-12 years and 40% age 13-17 years; follow-up: 66.7% age 4-12 years, 33.3% age 13-17 years | ASD-CP consisting of a modular staff training, toolkit and prescribed practices to be utilised with the person. | Not applicable | Psychiatric emergency care | Mental health care |
| Dreiling et al. (2022) | USA | Non-randomised service evaluation | Staff working with autistic people | 42.22 years (10.6), 25-66 | Project ECHO, a tele-mentoring platform to support mental health professionals | Not applicable | Community services | Mental health care |
| Helverschou et al. (2021) | Norway | Before-and-after comparison (service evaluation) | Autistic CYP and adults; ID | 28.6 years (10.6), 16-66 | AUP network consisting of meetings and seminars to guide mental health professionals in providing specialised mental health care | Not applicable | Specialist hospital-level mental health services | Psychiatric problems |
| Kuriakose et al. (2018) - linked to Cervantes et al. (2019) | USA | Before-and-after comparison (service implementation) | Autistic CYP | Pre-implementation 35.3% age 4-12 years and 64.7% age 13-17 years; post-implementation: 60% age 4-12 years and 40% age range 13-17 years | ASD-CP consisting of a modular staff training, toolkit and prescribed practices to be utilised with the person | Not applicable | Psychiatric emergency care | Mental health care |
| **General adaptations to standard practice** |  |  |  |  |  |  |  |  |
| Jones et al. (2021) | UK | Cross-sectional survey | Staff working with autistic people | - | Evaluation of strategies and adaptations to inpatient care | Not applicable | Inpatient psychiatric services | Mental health care |
| Petty et al. (2021) | UK | Qualitative (ethnographic technique) | Staff working with autistic people | 25-44 years | Evaluation of adaptations to improve mental health care. | Not applicable | Specialist autism service | Mental health care |
| Spain et al. (2017) | UK | Qualitative (thematic analysis) | Staff working with autistic people | - | Evaluation of general adaptations to standard practice | Not applicable | Inpatient and outpatient services | Mental health care |
| **Group CBT for anxiety** |  |  |  |  |  |  |  |  |
| Bemmer et al. (2021) | Australia | Before-and-after comparison | Autistic CYP and adults | 22.77 years (5.31), 16-38 | Adapted group CBT for social anxiety. | Yes | Research clinic | Social anxiety |
| Burke et al. (2017) | Ireland | Mixed methods (quantitative: before-and-after comparison; | Autistic CYP | 10 – 11 years | Adapted group CBT for anxiety ('FRIENDS for life' programme) | No information | Specialist autism service | Anxiety |
|  |  | qualitative: thematic analysis) |  |  |  |  |  |  |
| Chalfant et al. (2007) | Australia | RCT | Autistic CYP | 10.8 years (1.35), range 8-13 | Adapted group CBT for anxiety ('Cool Kids' programme) (n = 28) vs waitlist (n = 19). | Yes | School Outreach Service | Anxiety |
| Cook et al. (2017) | Australia | Pilot RCT | Autistic CYP | 5 years (0.83). CBT 5.5 years (0.88); waitlist 5.42 years (0.81) | Adapted group parent-mediated CBT for anxiety ('Fun with Feelings' programme) for children aged 4-6 years (n = 14) vs waitlist (n = 17) | Yes | University Psychology Clinic | Anxiety |
| Hepburn et al. (2016) | USA | Non-randomised controlled trial | Autistic CYP | CBT 11.5 years (2.67); waitlist 12.1 years (1.96) | Adapted group CBT for anxiety (telehealth 'FYF' programme) (n = 17) vs waitlist (n = 16) | Yes | Specialist autism clinic | Anxiety |
| E. Higgins et al. (2019) | Ireland | Mixed methods (quantitative: before-and-after comparison; qualitative: thematic analysis) | Autistic CYP | 10.25 years (1.26), 9-12 | Adapted group CBT for anxiety ('Special FRIENDS' programme) | Yes | Children disability service | Anxiety |
| Keefer et al. (2017) - linked to Reaven et al. (2018) | USA | Before-and-after comparison | Autistic CYP | 11.18 years (2.02) | Bespoke group CBT for anxiety ('FYF' programme) | Yes | University outpatient clinic | Anxiety |
| Kilburn et al. (2019) | Denmark | Before-and-after comparison | Autistic CYP | 9-13 years | Adapted group CBT for anxiety ('Cool Kids' programme) | Yes | Outpatient clinic at the Centre of Child and Adolescent Psychiatry | Anxiety |
| Kilburn et al. (2020) | Denmark | RCT | Autistic CYP | 11.34 years (1.77). CBT: 11.99 years (1.70); waitlist: 10.68 (1.60) | Adapted group CBT for fears and phobias ('Cool Kids' programme) (n = 25) vs waitlist (n = 24) | Yes | Outpatient clinic in a general child psychiatric hospital | Fears and phobias |
| Langdon et al. (2016) | UK | Pilot single-blind RCT | Autistic CYP and adults | 35.9 years (14.6), 17-65. CBT 33.1 years (14.6), 20-64; waitlist 38.7 years (14.3), 17-65. | Bespoke group CBT for anxiety + TAU (n = 26) vs TAU (n = 26) | No information | Community-based settings | Anxiety |
| McConachie et al. (2014) | UK | Pilot RCT | Autistic CYP | CBT 11.7 years (1.4); delayed therapy 11.8 years (1.3) | Bespoke group CBT for anxiety ('Exploring Feelings' programme) (n = 17) vs delayed therapy (n = 15) | Yes | Child and Adolescent Mental Health Services | Anxiety |
| Pickard et al. (2020) - linked to Reaven et al. (2018) | USA | Mixed methods survey | Staff working with autistic people | - | Bespoke group CBT for anxiety for youth 8-14 years ('FYF' programme) | Yes | University-based clinics | Anxiety |
| Reaven et al. (2009) | USA | Non-randomised controlled trial | Autistic CYP | 132 months (22.80), range 97-177 months | Bespoke group CBT for anxiety ('FYF' programme) (n = 10) vs waitlist (n = 23) | Yes | Community-based services | Anxiety |
| J. Reaven, Blakeley-Smith, Culhane-Shelburne et al. (2012) | USA | RCT | Autistic CYP | CBT 125.75 months (21.47); TAU 125 months (20.45) | Bespoke group CBT for anxiety ('FYF' programme) (n = 24) vs TAU (n = 26) | Yes | University outpatient clinic | Anxiety |
| J. Reaven, Blakeley-Smith, Leuthe et al. (2012) | USA | Before-and-after comparison study | Autistic CYP | 15.5 years (13.4), 13.4-18 | Adapted group CBT for anxiety ('FYF' programme - adolescent version) | Yes | Research clinic | Anxiety |
| Reaven et al. (2015) | USA and Canada | Before and after comparison study | Autistic CYP | 10.4 years (1.5), 8-13 | Bespoke group CBT for anxiety ('FYF' programme) | Yes | Tertiary paediatric health centre | Anxiety |
| Reaven et al. (2018) | USA | RCT (3-group parallel design) | Autistic CYP | Manual 132.9 months (20.4); workshop only 130.8 months (27.6); workshop plus 135.5 months (19.5) | Bespoke group CBT for anxiety ('FYF' programme) | Yes | University-affiliated outpatient clinics | Anxiety |
| Sofronoff et al. (2005) | Australia | RCT | Autistic CYP | CBT child only 10.56 years (0.99), 9-12; CBT child + parent 10.54 years (1.26), 9-12; waitlist 10.75 years (1.04) 9-12 | Bespoke group CBT for anxiety: child only (n = 23) vs child + parent (n = 25) vs waitlist (n = 23) | Yes | University psychology clinic | Anxiety |
| Solish et al. (2020) | Canada | Before-and-after comparison (service evaluation) | Autistic CYP | Specialised hospital 10.08 years (1.71); community 10.87 (1.72) | Bespoke group CBT for anxiety ('FYF' programme) | Yes | Community clinics and specialised hospital | Anxiety |
| Sung et al. (2011) | Singapore | RCT | Autistic CYP | 9-16 years. CBT 11.33 years (2.03); Social recreational 11.09 years (1.53) | Adapted group CBT for anxiety (n = 36) | No | Outpatient mental health clinic for children and adolescents | Anxiety |
| Walsh et al. (2018) - linked to Reaven et al. (2018) | USA | RCT (3 group parallel design) | Autistic CYP | 133.35 (23.59) months | Bespoke group CBT for anxiety ('FYF' programme) | Yes | University-affiliated outpatient clinics | Anxiety |
| **Individual CBT for anxiety** |  |  |  |  |  |  |  |  |
| Driscoll et al. (2020) | USA | Before-and-after comparison | Autistic CYP | 5.7 years (1.4), 3-7 | Adapted individual CBT for anxiety ('Being Brave' programme) | Yes | Outpatient clinics of two urban hospitals affiliated with a medical school | Anxiety |
| Ehrenreich-May et al. (2014) | USA | Before-and-after comparison | Autistic CYP | 12.2 years (1.11), range 11-14 | Adapted individual CBT for anxiety ('BIACA' programme) | Yes | University treatment centres | Anxiety |
| Ekman et al. (2015) | Sweden | Before-and-after comparison | Autistic CYP and adults | Teens 14.9 years (1.5), 13-17; adults 29.8 years (4.4), 23-36 | Adapted individual CBT for anxiety | No information | Private clinic, child and adolescent psychiatric clinic, treatment centre for youth | Anxiety |
| Fujii et al. (2013) | USA | Pilot RCT | Autistic CYP | 8.80 years (1.60), 7-11. CBT 8.7 years (1.8); TAU 9 years (1.6) | Adapted individual CBT for anxiety ('BIACA' programme) (n = 7) vs TAU (n = 5) | Yes | University clinic and an associated autism community clinic | Anxiety |
| Maskey, McConachie, et al. (2019) | UK | Before-and-after comparison | Autistic CYP | 8-12 years | Bespoke individual CBT for fears and phobias using flat screen computer delivery of images ('Blue Room') | Can't tell | NHS/university setting | Fears and phobias |
| Maskey, Rodgers et al. (2019) | UK | Feasibility RCT | Autistic CYP | CBT 130.13 months (28.38), 89-174; delayed therapy 129 months (21.51), 90-157 | Bespoke individual CBT for anxiety with virtual reality ('Blue Room') (n = 16) vs delayed therapy (n = 16) | Can't tell | Recruited from mental health services but delivered in a university setting | Anxiety |
| Oerbeck et al. (2021) | Norway | Before-and-after comparison study | Autistic CYP | 9.5 years, range 8-12; | Adapted individual CBT for anxiety ('Less Stress' programme) | Yes | Child and Adolescent Mental Health Clinics | Anxiety |
| Ollendick et al. (2021) | USA | Before-and-after comparison study | Autistic CYP | 6-14 years | Adapted individual CBT for specific phobia ('OST') | Yes | Clinic | Phobias |
| A. J. Russell et al. (2013) | UK | Single-blind RCT | Autistic CYP and adults | 26.9 years, range 14-65. CBT 28.6 years (11.3), 14-49; anxiety management 25.2 (13.5), 14-65 | Adapted individual CBT for OCD (n = 23) vs adapted anxiety management (n = 23) | No information | Specialist autism, OCD clinics and mental health services | OCD |
| Storch et al. (2013) | USA | RCT | Autistic CYP | 8.89 years (1.3), 7-11. CBT 8.83 years (1.31); TAU 8.95 years (1.40) | Adapted individual CBT for anxiety ('BIACA' programme) (n = 24) vs TAU (n = 11) | Yes | University based mental health clinic | Anxiety |
| Storch et al. (2015) | USA | RCT | Autistic CYP | 2.74 years (1.34), 11-16. CBT 12.75 years (1.24); TAU 12.73 years (1.49) | Adapted individual CBT for anxiety ('BIACA' programme) (n = 16) vs TAU (n = 15) | Yes | University based multidisciplinary behavioural health clinic specialising in the treatment of paediatric anxiety | Anxiety |
| Storch et al. (2020) | USA | Pilot RCT | Autistic CYP | 10.03 years (2.81), 6-17. FET 10.07 years (2.89); TAU 10 years (SD = 2.83) | Adapted individual FET for anxiety (n = 14) | Yes | Tertiary care clinic specialising in paediatric obsessive compulsive and anxiety disorders | Anxiety |
| Wise et al. (2019) | USA | Before-and-after comparison | Autistic CYP and adults | 17.14 years (1.68), range 16-20 | Adapted individual CBT for anxiety | Yes | University-based health clinic specialising in the treatment of anxiety | Anxiety |
| Wood et al. (2015) | USA | RCT | Autistic CYP | 12.3 years (1.14), 11-15. CBT 12.4 years (1.3); waitlist 12.2 (0.98) | Adapted individual CBT for anxiety 'BIACA' programme) (n = 19) vs waitlist (n = 14) | Yes | University clinic and associated autism community clinic | Anxiety |
| **Individual and group CBT for anxiety** |  |  |  |  |  |  |  |  |
| Murphy et al. (2017) | UK | Pilot RCT | Autistic CYP | CBT 14.94, (1.63); waitlist 15.56 (1.91) | Bespoke individual and group CBT for anxiety ('MASSI' programme) (n = 17) vs counselling (n = 19) | Yes | Child and Adolescent Mental Health Services | Anxiety |
| White et al. (2013) | USA | Pilot RCT | Autistic CYP | 175 months (15 years). CBT 170 months (14 years); waitlist 180 months (15 years) | Bespoke individual and group CBT for anxiety ('MASSI' programme) (n = 15) vs waitlist (n = 11) | Yes | University-affiliated clinic specialising in autism treatment | Anxiety |
| White et al. (2015) - linked to White et al. (2013) | USA | Pilot RCT | Autistic CYP | 174.05 months (18.66) | Bespoke individual and group CBT for anxiety ('MASSI' programme) (n = 11) vs waitlist (n = 11) | Yes | University-affiliated clinic specialising in autism treatment | Anxiety |
| **Interventions targeting emotional regulation** |  |  |  |  |  |  |  |  |
| Drusedau et al. (2022) | Germany | Before-and-after comparison | Autistic CYP | 10.08 years (1.32), range 7-12 | Bespoke group mindfulness-based intervention ('TütASS' programme) | Yes | Outpatient unit at University Hospital of Psychiatry and Psychotherapy | Mental health difficulties; emotion regulation |
| Factor et al. (2019) | USA | RCT | Autistic CYP | 5.46 (1.01), 4-7. CBT 5.54 years (0.94); waitlist 5.36 years (1.12) | Adapted group CBT for emotion regulation ('STAMP' programme) for younger children (4-7 years) (n = 12) vs delayed therapy (n = 11) | Yes | Autism clinic | Mental health difficulties; emotion regulation |
| Scarpa et al. (2011) | USA | Pilot RCT | Autistic CYP | 5-7 years | Adapted group CBT for emotion regulation (n = 5) | Yes | Autism clinic | Mental health difficulties; emotion regulation |
| Sofronoff et al. (2017) | Australia | Before-and-after comparison | Autistic CYP | 9.56 years, 7 years and 11 months-12 | Adapted group cognitive behavioural emotional and social skills intervention ('SAS' programme). | Yes | University clinic | Mental health difficulties; emotion regulation |
| Swain et al. (2019) | USA | Before-and-after comparison study | Autistic CYP | 3.89 months (11.87), 53-90 months | Adapted group CBT for young children (aged 4-8) ('STAMP' programme) | Yes | University associated community clinic and a hospital | Mental health difficulties; emotion regulation |
| **CBT for various mental health needs** |  |  |  |  |  |  |  |  |
| Cooper et al. (2018) | UK | Cross-sectional survey | Staff working with autistic people | - | Adapted individual CBT | Yes | IAPT and secondary mental health services | Mental health difficulties |
| McGillivray et al. (2014) | Australia | Non-randomised controlled trial | Autistic CYP and adults | 20.6 years (4.1), 15-25. CBT 20.27 years (4.39); waitlist 20.50 (3.4) | Bespoke group CBT for anxiety, stress and depression ('think well, feel well and be well' programme) (n = 26) vs waitlist (n = 16) | No information | Disability service agency | Mental health difficulties |
| Santomauro et al. (2016) | Australia | Pilot RCT | Autistic CYP | 15.75 years (1.37). CBT 16 years (1.33); waitlist 15.5 years (1.43). | Bespoke group CBT for depression ('Exploring depression' programme) (n = 11) vs waitlist (n = 12) | Can't tell | University clinic | Depression |
| **EMDR for PTSD** |  |  |  |  |  |  |  |  |
| Fisher et al. (2023) | Netherlands; UK | Delphi survey (3 rounds) | Staff working with autistic people | - | Adapted EMDR | Can't tell | Psychological therapies, community mental health, ID, forensic and tertiary services, independent practice, education, military, voluntary organisations | PTSD |

*Note.* Where age characteristics are not listed in the table this means that they were not reported in the paper. **ADHD** = Attention Deficit Hyperactivity Disorder, **ADOS** = Autism Diagnostic Observation Schedule, **ASC** = Autism Spectrum Conditions, **ASD-CP** = Autism Spectrum Disorder Care, **AUP** = Pathway Autism Intellectual Disability and Psychiatric Disorder, **BIACA** = Behavioral Interventions for Anxiety in Children with Autism, **CBT** = Cognitive Behavioural Therapy, **CYP** = Children and Young People, **DAWBA** = Development and Well-Being Assessment, **ECHO** = Extension for Community Healthcare Outcomes, **EMDR** = Eye Movement Desensitisation and Reprocessing, **FET** = Family-based exposure-focused treatment, **FYF** = Facing Your Fears, **IAPT** = Improving Access to Psychological Therapies, **ID** = Intellectual Disability, **MASSI** = Multimodal Anxiety and Social Skills Intervention, **OCD** = Obsessive Compulsive Disorder, **OST** = One-Session Treatment, **PTSD** = Post Traumatic Stress Disorder, **RCT** = Randomised Controlled Trial, **SAS** = Secret Agent Society, **SCQ** = Social Communication Questionnaire, **STAMP** = Stress and Anger Management Programme, **TAU** = Treatment as Usual, **TütASS** = Tübinger Training for Autism Spectrum Disorders.

### Quality appraisal and publication bias

According to appraisal using the MMAT, for RCTs, 13 papers were of high (≥4 criteria met), five papers were of moderate (3 criteria met), and three papers were of low quality (≤2 criteria met). Appropriate randomisation and blind outcome assessors were the main areas of concern for RCTs. For non-randomised studies, six were of high, 15 of moderate, and two of low quality. These studies often did not meet the criteria for representativeness and confounder adjustment. For quantitative descriptive studies, three were of high, one of moderate and one of low quality. Nonresponse bias was the main area of concern for these studies. For mixed-method studies, five were of high (of which two combined RCT with qualitative methods), and one of low quality. The two qualitative studies were of high quality. All MMAT ratings are shown in Table S7. Visual inspection of the funnel plots showed outliers (Figure S1). Egger’s test was significant (child/self *z* = 2.13, *p* = .033; parent *z* = 4.70, *p* < .001; clinician *z* = 3.99, *p* < .001), suggesting the presence of publication bias.

### Autism-Inclusive Research Assessment

Four out of 57 papers (7%) reported involvement of autistic people in study design or delivery. One of the 10 papers (10%) with a qualitative element reported adjustments to the data collection process (e.g., allowing non-verbal communication). Five out of 55 papers (9%) with a quantitative element reported making some adjustments to the data collection tools (e.g., defining key terms, using straightforward language, adapting Likert scales for greater precision, using visual tools). Thirteen out of 55 papers (24%) with a quantitative element reported using at least one valid or adapted measure for autistic individuals relevant to the review. For 12 of the 50 papers (24%) with a quantitative element that measured outcomes in autistic mental health service users, the intervention/strategy was identified to involve some focus on masking people’s autistic traits. However, 36 of the 50 papers (72%) did not include any evidence to suggest such a focus, and this was unclear for two of the 50 papers (4%). All extracted data from the AIRA are shown in Table S8.

### Sample characteristics

Sample sizes at baseline in the papers ranged from 7 to 132 autistic participants (median 32, n=43 studies), 62-302 participants (median 77, n=3 studies) for studies of strategies to improve the detection of autism, 11-105 parents (median 33, n=18 studies), and 15-103 clinicians (median 42, n=8 studies). Fifty papers included CYP, all of whom were given an autism diagnosis, except for three papers regarding initiatives to improve the detection of autism. Two papers included participants with co-occurring intellectual disability (ID). Forty-seven papers described co-occurring mental health difficulties at baseline. Forty-three papers included CYP with an age range 3-18 years, and seven papers reported on combined outcomes of CYP and adults with an age range 13-66 years. Ten papers included clinicians as participants. Detailed sample characteristics are in Table S6.

### Data synthesis

#### Strategies used to improve mental health care in autism

Identified strategies included service-level strategies (n=10) and adapted/bespoke mental health interventions (n=47). From the identified intervention-level and service-level adaptations, those regarding communication and intervention content were most frequently reported, and adjustment to the environment were least included. Most papers focused on CBT-based mental health interventions for anxiety. Additionally, 37 papers described caregiver involvement in therapy, such as being offered separate/combined sessions. Tables 1 and S6 contain descriptions of the included strategies and caregiver involvement.

##### Service-level strategies and adapted interventions

Ten papers explored service-level strategies applied to improve mental health care for autistic people across a service. These papers explored initiatives to improve the detection of autism (Ford et al., 2019; Hollocks et al., 2019; Stadnick et al., 2015), strategies for improving clinicians’ skills and knowledge of autism (Cervantes et al., 2019; Dreiling et al., 2022; Helverschou et al., 2021; Kuriakose et al., 2018), and general adaptations to standard practice concerning the way mental health services are organised for autistic people (Jones et al., 2021; Petty et al., 2021; Spain et al., 2017).

Twenty-eight papers described studies of adapted mental health interventions to meet the needs of autistic people. These included adaptations of group or individual CBT for anxiety (Bemmer et al., 2021; Burke et al., 2017; Chalfant et al., 2007; Cook et al., 2017; Driscoll et al., 2020; Ehrenreich-May et al., 2014; Ekman et al., 2015; Fujii et al., 2013; Hepburn et al., 2016; E. Higgins et al., 2019; Kilburn et al., 2019, 2020; Oerbeck et al., 2021; Ollendick et al., 2021; J. Reaven, Blakeley-Smith, Leuthe, et al., 2012; A. J. Russell et al., 2013; Storch et al., 2013, 2015, 2020; Sung et al., 2011; Wise et al., 2019; Wood et al., 2015), group CBT targeting emotion regulation (Factor et al., 2019; Scarpa & Reyes, 2011; Sofronoff et al., 2017; Swain et al., 2019), individual CBT for various mental health needs (Cooper et al., 2018), and Eye Movement Desensitisation and Reprocessing (EMDR) (Fisher et al., 2023). Studies with a comparison group most often compared the adapted interventions to non-active controls, and none compared it to a non-adapted version of the same intervention.

Seven top-level adaptation categories were identified from these papers exploring service- and intervention-level adaptations:

- Increasing knowledge and detection of autism (n=10, e.g., use of screening tools, clinician training)
- Adjustments to the physical environment (n=6, e.g., minimising sensory distractions, providing ear defenders, weighted blankets, fidget toys, movement breaks)
- Communication accommodations (n=20, e.g., being directive, adjusting the communication pace, using preferred language, using written information on whiteboard, activity books, agendas and visual aids like drawings, videos, using social stories, using a computer to reduce face-to-face contact)
- Accommodating individual differences (n=16, e.g., evaluating preferences and needs, encouraging special interests and hobbies, and tailoring treatment to these by being flexible with the treatment manual)
- Structural or procedural adaptations (n=15, e.g., changing the format, duration or number of sessions, having predictable session routines and structured approach to treatment with details communicated in advance)
- Intervention content adaptations (n=24, e.g., removing or simplifying psychoeducation and cognitive elements of the intervention, incorporating arts-based activities, using role-play, rewards, taking a progressive approach to treatment with opportunity for repetition and practice)
- Involving the wider support network (n=18, e.g., involving parents and child’s school to support active transfer of skills/therapy goals from clinic to home and school)

Several adaptations were reported within the same paper, meaning papers crossed several categories. Most papers provided a general rationale for these adaptations as addressing barriers to mental health care. There were limited descriptions of specific adaptations and their rationale. Table 2 shows a breakdown of sub-categories which map to these top-level categories. Table S9 includes details of the individual adaptations used by each paper.

**Table 2.** All service-level and intervention-level adaptations (simplified version) (*N* =38)

| Top-level categories | Sub-categories | Summary | *N* articles* |
| --- | --- | --- | --- |
| **Increase knowledge and detection of autism (n=10)** | Clinician training and skills | Training to provide clinicians with an overview of ASD and to tailor treatment to individual needs and increase self-efficacy, knowledge of autism and skills. Use of skills such as normalising experiences and prioritising the therapeutic relationship | 6 |
|  | Introduction of screening tools for the detection of autism | Use of assessments such as the ADOS, SCQ and DAWBA to improve the assessment and detection of autism | 4 |
| **Adjustments to the physical environment (n=6)** | Provide environmental and practical adjustments | Provide adjustments to minimise sensory distractions such as a low stimulus area, adjustments to noise, decor, scents, and lighting | 5 |
|  | Encourage the use of sensory resources and stimming | Provide sensory resources such as tactile objects, include movement breaks and exercises and use multi-sensory activities, encourage use of stimming behaviour | 5 |
| **Communication accommodations (n=20)** | Use of clear, simple and preferred language | Provide clear instructions and guidance, be more directive, monitor, adapt and slow the pace of communication, use preferred language where possible | 7 |
|  | Use of simple, written material and visual aids | Use of written information and external cues such as use of a whiteboard, activity books, worksheets, timers, agendas and calendars. Use of visual aids such as drawings, pictures, videos and leaflets | 16 |
|  | Provide communication support | Use of communication passports and social stories to support communication. | 1 |
|  | Use of technology | Incorporating technology into the intervention to aid communication | 2 |
| **Accommodate individual differences (n=16)** | Evaluate individual needs and preferences | Evaluate preferences, sensitivities, sensory needs, likes and dislikes and coping strategies. | 3 |
|  | Encourage individual's hobbies and interests | Include and ask about the individual's special interests and hobbies in therapy. | 4 |
|  | Tailor practice to individual needs and preferences | Tailor care plans and practice to individual differences such as incorporating approaches targeted at neurodevelopmental comorbidities, being flexible with the treatment manual and the session timings and ensuring that resources are appropriate for the person's gender | 13 |
| **Structural or procedural adaptations (n=15)** | Format of intervention | Reduce or increase the number and duration of sessions and exercises, conduct exposure in more varied settings | 8 |
|  | A structured and predictable approach | Having predictable session routines and a structured approach to treatment, with details communicated in advance | 8 |
| **Intervention content adaptations (n=24)** | Simplified content | Remove or simplify psychoeducation and cognitive elements of the intervention | 5 |
|  | Creative outlets and activities | Incorporating creativity and arts-based activities into the intervention | 3 |
|  | Use of role play or modelling | Using role play as a technique to reinforce learning during sessions, or modelling activities e.g., by video, or using a puppet or character | 6 |
|  | Use of a rewards system | Using reward systems to help reinforce learning | 4 |
|  | Taking it slow | Taking a slow/progressive approach to treatment, with opportunity for repetition and practice | 6 |
|  | Consider the role of autism | Consider the role of autism, develop an understanding of autism such as its characteristics and impact on daily life | 2 |
|  | Integration of emotion-focused strategies | Provide psychoeducation on emotions, arousal and feeling physiologically overwhelmed and exercises to access emotions | 7 |
|  | Integration of cognitive-behavioural approaches | Provide cognitive and behavioural strategies including building a positive self-image, coping strategies, and making links between behaviour, thoughts and feelings | 6 |
|  | Integration of social skills training | Integration of social skills training such as entering and maintaining conversations and managing disagreements | 6 |
| **Involving wider support network (n=18)** | Parental involvement | Involving parents in the intervention | 17 |
|  | Involving school | Involving or incorporating the child's school into the intervention | 3 |

Note. ADOS = Autism Diagnostic Observation Schedule, SCQ = Social Communication Questionnaire, DAWBA = Development and Well-Being Assessment
\* Several adaptations were often reported by the same article, meaning papers crossed several categories so the number of papers in this column does not add up to the total 38 contributing papers

##### Bespoke interventions

Nineteen papers described bespoke mental health interventions originally developed for autistic CYP, often in part by the authors themselves, and tested in their unmodified version. These included a novel combined group and individual intervention for anxiety (Murphy et al., 2017; White et al., 2013, 2015), individual interventions for anxiety in a virtual reality environment (Maskey, McConachie, et al., 2019; Maskey, Rodgers, et al., 2019), group interventions for anxiety (Keefer et al., 2017; Langdon et al., 2016; McConachie et al., 2014; Pickard et al., 2020; J. A. Reaven et al., 2009; J. Reaven et al., 2015, 2018; J. Reaven, Blakeley-Smith, Culhane-Shelburne, et al., 2012; ; Solish et al., 2020; Walsh et al., 2018), group interventions for anxiety, stress, and depression (McGillivray & Evert, 2014) and for depression only (Santomauro et al., 2016), all utilising CBT techniques. Additionally, they included a new group intervention for emotion regulation designed for autistic CYP based on mindfulness principles (Drüsedau et al., 2022).

#### Acceptability, feasibility, and effectiveness of strategies used to improve mental health care for autistic CYP

##### Evaluation of service-level strategies

Ten papers evaluated service-level strategies, grouped into three categories depending on their focus. The main findings of service-level strategies are presented in Table 3, with detailed results of individual studies in Table S12. Tables S10 shows the GRADE assessment for effectiveness outcomes.

**Table 3.**
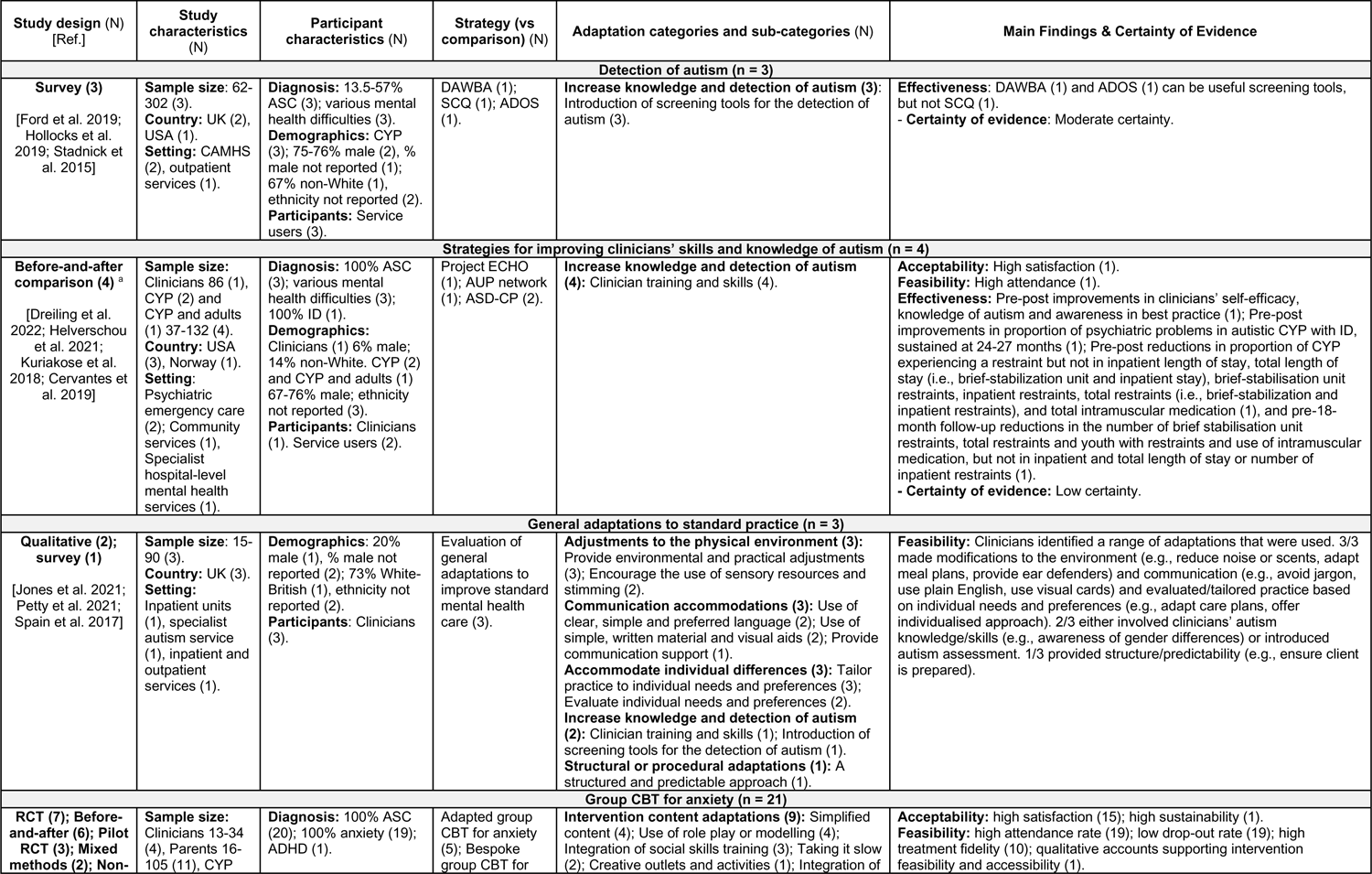
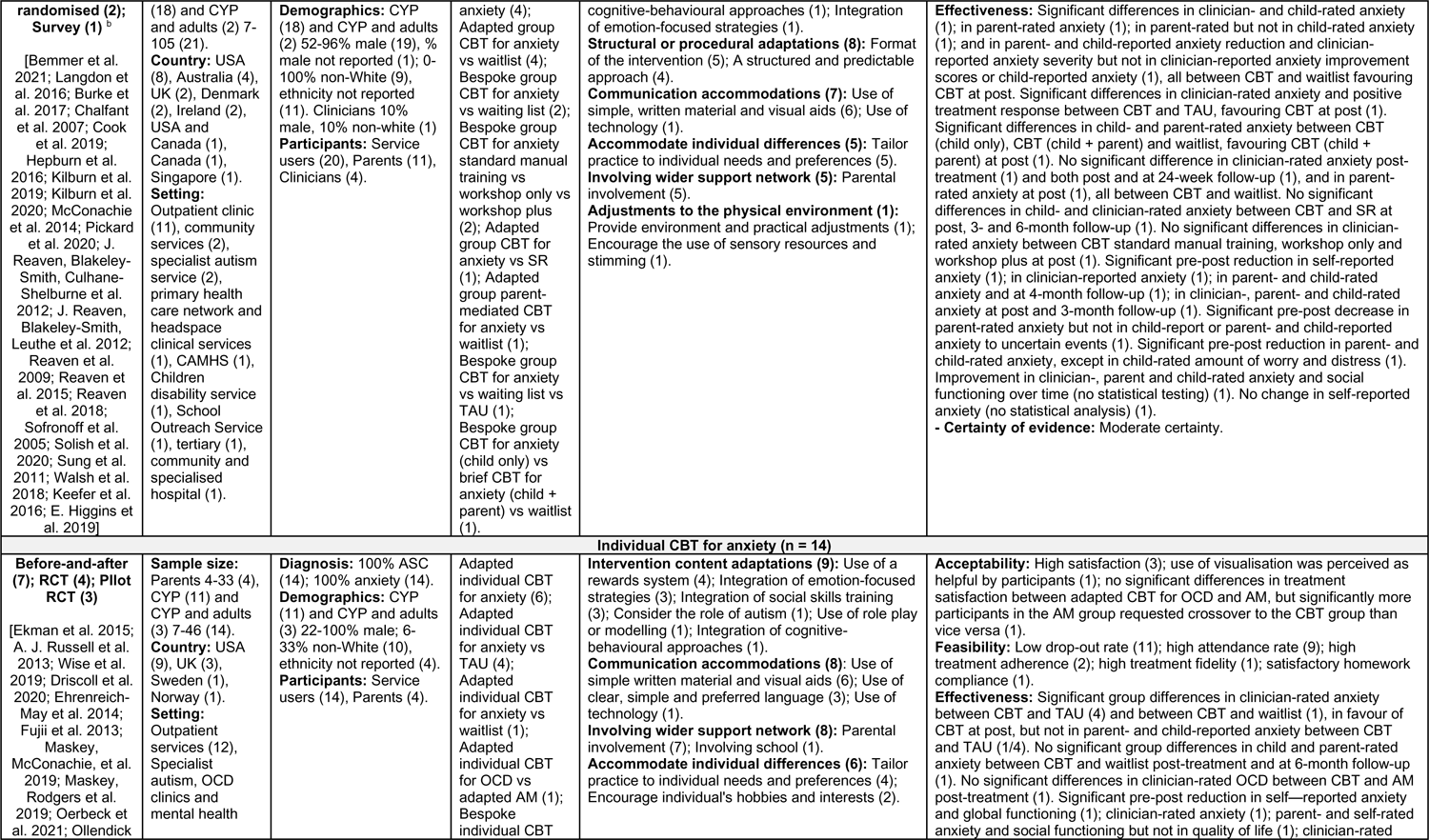
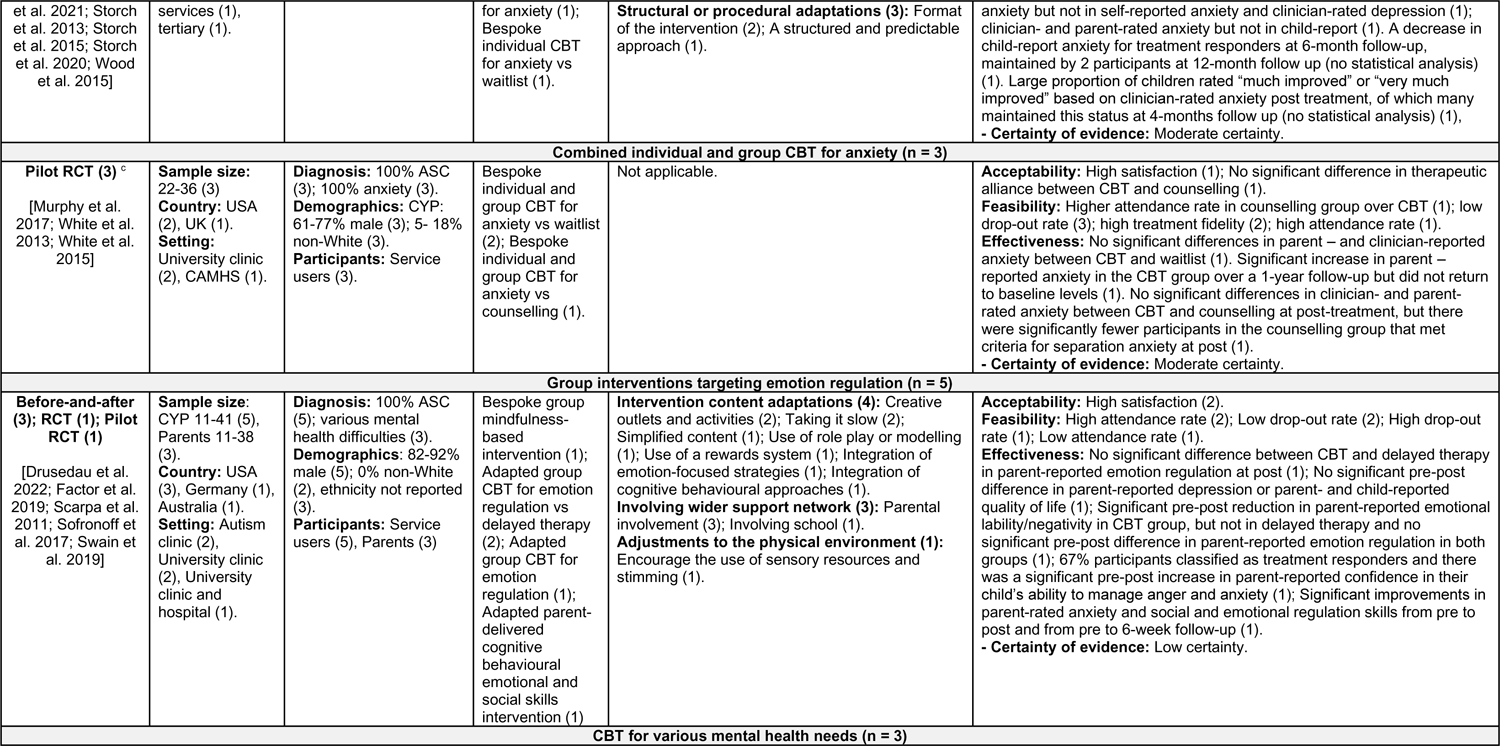
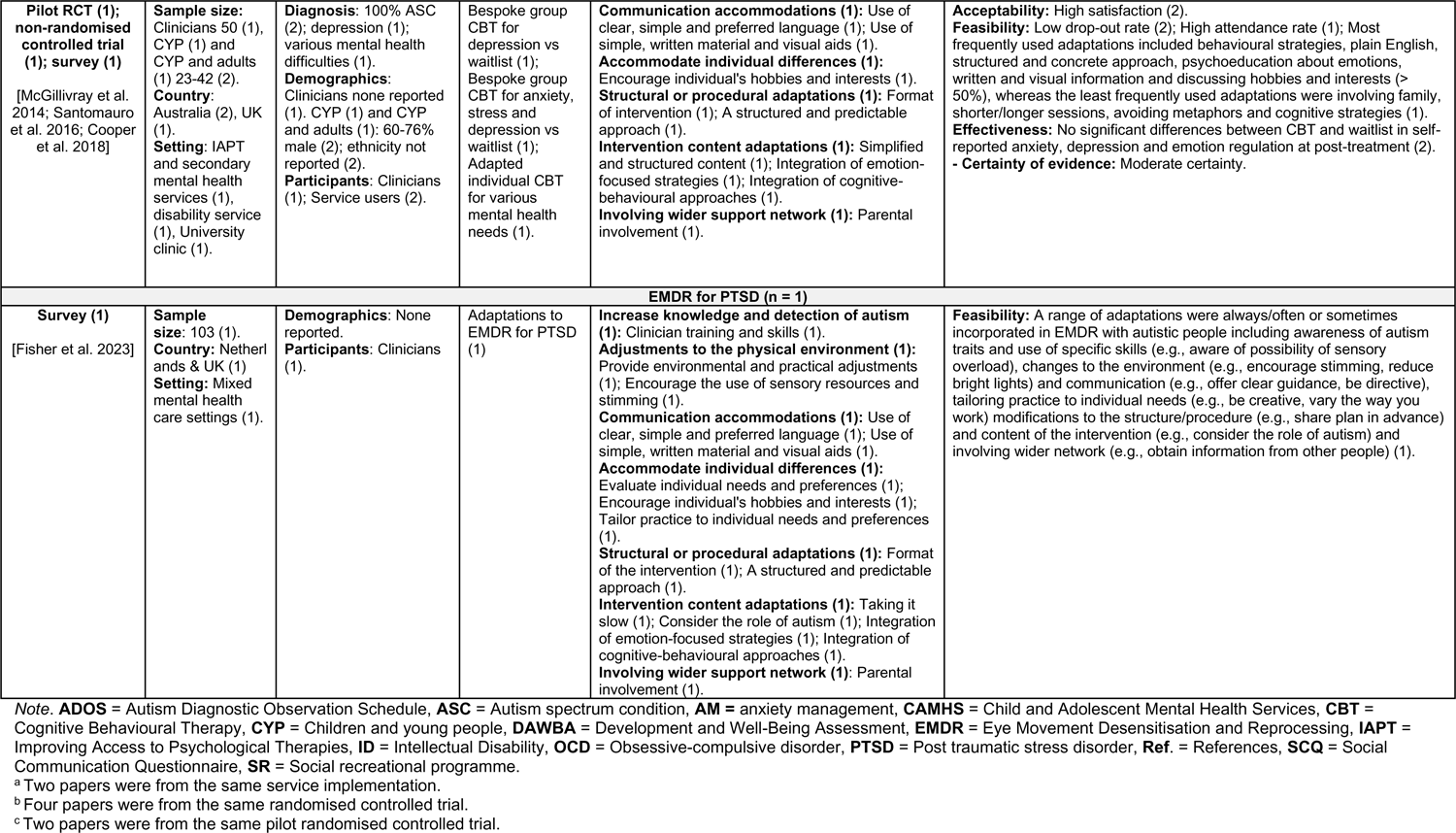
Main findings of individual/group and adapted/bespoke mental health interventions/strategies and service adaptations.

###### 1. Detection of autism (n=3)

Overall, moderate certainty evidence suggested that some screening tools may be helpful in detection of autism in mental health services (Table S10). The Development and Well-being Assessment (DAWBA) was found to have moderate agreement with practitioner diagnosis of autism in child and adolescent mental health services (CAMHS), suggesting it may be useful to aid the diagnostic process (Ford et al., 2019). Conversely, the Social Communication Questionnaire (SCQ) was found to not be an effective autism screening tool in CAMHS (Hollocks et al., 2019). The Autism Diagnostic Observation Schedule (ADOS) administered in community mental health services was found to identify autistic CYP referred for an autism assessment (Stadnick et al., 2015).

###### 2. Strategies for improving clinicians’ skills and autism knowledge (n=4)

Strategies, involving training and guiding clinicians to provide better care across the lifespan to autistic people with co-occurring mental health needs, included the Extension for Community Healthcare Outcomes autism model in community services (Dreiling et al., 2022), the Autism Intellectual Disability and Psychiatric Disorder network in specialist mental health services (Helverschou et al., 2021) and the Autism Spectrum Disorder Care Pathway in psychiatric emergency care (Cervantes et al., 2019; Kuriakose et al., 2018).

The Extension for Community Healthcare Outcomes autism model was found feasible and acceptable to clinicians (Dreiling et al., 2022). All strategies were associated with significant improvements over time; however, causality cannot be concluded since there were no comparison groups. Overall, low-certainty evidence suggested that some strategies for improving clinicians’ skills and knowledge of autism (Cervantes et al., 2019; Dreiling et al., 2022; Helverschou et al., 2021; Kuriakose et al., 2018) may be helpful in improving mental health of autistic individuals (Table S10).

###### 3. General adaptations to services (n=3)

Clinicians reported using a range of adaptations in inpatient units (Jones et al., 2021), a specialist autism service (Petty et al., 2021), and inpatient and outpatient services (Spain et al., 2017). All papers described clinicians modifying the environment and communication and reported clinicians evaluating and adapting practice based on individual needs. Only one reported on clinicians providing structure to reduce uncertainty (Petty et al., 2021). None of the papers evaluated the impact of these general adaptations.

##### Evaluation of interventions

Forty-seven of the included papers, broadly grouped based on similarities in type and focus in four intervention categories, evaluated the effectiveness of interventions in improving autistic individuals’ mental health and/or their acceptability/feasibility. The main findings of evaluated interventions are presented in Table 3, with detailed results of individual studies in Table S12. Tables S10 and S11 show the GRADE assessment for effectiveness outcomes.

###### 1. CBT for anxiety (n=38)

Thirty-five out of the 38 papers that tested adapted or bespoke individual, group, or combined individual and group CBT for anxiety, reported feasibility outcomes. All 35 papers reported the interventions were feasible largely based on low drop-out rates, high attendance rates and treatment fidelity (Bemmer et al., 2021; Chalfant et al., 2007; Driscoll et al., 2020; Ehrenreich-May et al., 2014; Fujii et al., 2013; Hepburn et al., 2016; E. Higgins et al., 2019; Keefer et al., 2017; Kilburn et al., 2019, 2020; Langdon et al., 2016; Maskey, McConachie, et al., 2019; Maskey, Rodgers, et al., 2019; McConachie et al., 2014; Murphy et al., 2017; Oerbeck et al., 2021; Ollendick et al., 2021; Pickard et al., 2020; J. A. Reaven et al., 2009; J. Reaven et al., 2015, 2018; J. Reaven, Blakeley-Smith, Culhane-Shelburne, et al., 2012; J. Reaven, Blakeley-Smith, Leuthe, et al., 2012; A. J. Russell et al., 2013; Sofronoff et al., 2005; Solish et al., 2020; Storch et al., 2013, 2015, 2020; Sung et al., 2011; Walsh et al., 2018; White et al., 2013, 2015; Wise et al., 2019; Wood et al., 2015). Twenty-three out of 38 papers reported acceptability outcomes of adapted/bespoke individual, group, or combined CBT for anxiety for either CYP, parents or clinicians. All 23 papers reported the interventions were acceptable based on participant-reported positive experiences and intervention satisfaction (Bemmer et al., 2021; Burke et al., 2017; Cook et al., 2017; Ekman et al., 2015; Hepburn et al., 2016; E. Higgins et al., 2019; Kilburn et al., 2019, 2020; Langdon et al., 2016; McConachie et al., 2014; Murphy et al., 2017; Oerbeck et al., 2021; Ollendick et al., 2021; Pickard et al., 2020; J. Reaven et al., 2015; J. Reaven, Blakeley-Smith, Culhane-Shelburne, et al., 2012; J. Reaven, Blakeley-Smith, Leuthe, et al., 2012; A. J. Russell et al., 2013; Sofronoff et al., 2005; Solish et al., 2020; Walsh et al., 2018; White et al., 2013; Wood et al., 2015).

Facilitators to acceptability reported by participants included perceived positive intervention impact (E. Higgins et al., 2019; Kilburn et al., 2020; Langdon et al., 2016; McConachie et al., 2014; Oerbeck et al., 2021; Ollendick et al., 2021; Sofronoff et al., 2005; Solish et al., 2020) and perceived usefulness of intervention’s information/activities/techniques (Bemmer et al., 2021; E. Higgins et al., 2019; Langdon et al., 2016; McConachie et al., 2014; Oerbeck et al., 2021; J. Reaven et al., 2015; J. Reaven, Blakeley-Smith, Culhane-Shelburne, et al., 2012; J. Reaven, Blakeley-Smith, Leuthe, et al., 2012; Solish et al., 2020; Walsh et al., 2018). Feeling accepted/supported by the group (Bemmer et al., 2021; E. Higgins et al., 2019; Langdon et al., 2016; McConachie et al., 2014), interaction with others (Bemmer et al., 2021; E. Higgins et al., 2019; Langdon et al., 2016; McConachie et al., 2014; Sofronoff et al., 2005), and individual preparatory sessions prior to group sessions (Langdon et al., 2016) also appeared important. Additionally, receiving preparatory handout for upcoming sessions (E. Higgins et al., 2019), perceived parental confidence with the intervention content and thus being able to support their child (Sofronoff et al., 2005; Solish et al., 2020), understanding assignments (Oerbeck et al., 2021), and getting rewards (Oerbeck et al., 2021) were seen as facilitators. Using visualisation was viewed as helpful (Ekman et al., 2015). Clinicians’ participation in a short training workshop appeared to facilitate higher acceptability, as opposed to receiving additional ongoing feedback or only a manual (Walsh et al., 2018).

Participants also reported barriers to acceptability, including perceiving the sessions to be too long/short (Bemmer et al., 2021; Langdon et al., 2016), difficulties with group dynamics (Bemmer et al., 2021; Langdon et al., 2016), feeling anxiety limited their learning (Bemmer et al., 2021), feeling the individual sessions involved too much talking (Oerbeck et al., 2021), perceived lack of learning (McConachie et al., 2014), dissatisfaction with visuals (Oerbeck et al., 2021), children‘s reluctance to talk to parents about content beyond the sessions (E. Higgins et al., 2019), difficulties with homework assignments (Oerbeck et al., 2021), and difficulties with making phone calls (Bemmer et al., 2021). Practical issues related to transport, parking, heating in session rooms and timings also appeared to hinder acceptability (E. Higgins et al., 2019; Langdon et al., 2016). The addition of bi-weekly feedback and consultation next to training workshops might have put clinicians under pressure (Walsh et al., 2018).

Clinicians who continued to implement group version of CBT for anxiety for at least four years following training reported tailoring, lengthening, removing, shortening, and supplementing the intervention’s components to enhance and adapt it to the learning needs of CYP and carers (Pickard et al., 2020). Positive clinicians’ views of the intervention’s effectiveness, ease of use, and fit with existing service were perceived as facilitators for sustained use of this intervention. Reported barriers included the intervention no longer being relevant to the service, services being unable to support delivery, clinicians no longer working clinically, inability to obtain funding for intervention, and difficulties with group format of the intervention due to insufficient staffing and challenges with recruiting a group of CYP of the same age and level of support needs (Pickard et al., 2020).

##### Effectiveness of CBT for anxiety

Thirty-six out of 38 papers evaluating CBT for anxiety reported effectiveness outcomes. Sixteen RCTs/pilot RCTs testing effectiveness of adapted/bespoke individual, group, and combined CBT for anxiety compared to any control group were included in three meta-analyses depending on the rater of the autistic person’s anxiety measure.

###### Child/self-rater meta-analysis

CBT was not significantly different from control, including treatment as usual (TAU), waitlist, adapted anxiety management (AM) and social recreation (SR), in reducing child/self-rated anxiety symptoms at EOT (*k* = 9, *g* = 0.34 [95% CI –0.15, 0.84], *p* = .173) (Figure 2) (moderate-certainty evidence, Table S11). There was significant heterogeneity among studies, *Q*(8)= 43.85, *p* < .001, *I^2^* = 81.75%. On removal of outliers (Chalfant et al., 2007; Langdon et al., 2016), there were no significant differences between groups at EOT (*k* = 7, *g* = 0.17 [95% CI –0.07, 0.40], *p* = .169), but heterogeneity reduced, *Q*(6) = 3.21, *p* = .782, *I^2^* = 0%.

**Figure 2.**
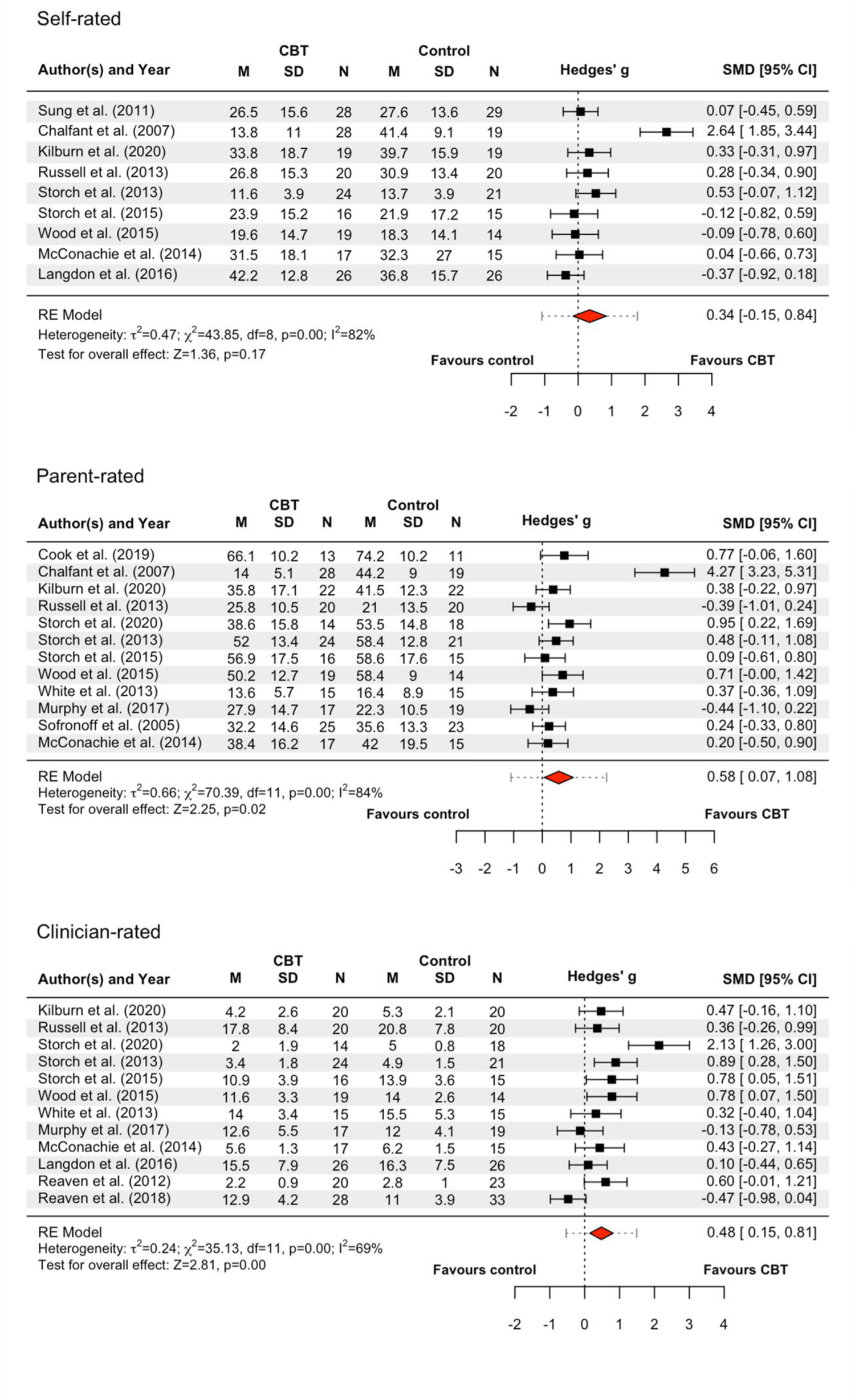
Forest plots of meta-analyses comparing cognitive behavioural therapy (CBT) for anxiety with any control group in reducing anxiety symptom severity in autistic individuals. *Note*. Continuous rather than dichotomous data were used, as this was the most frequent type of data across studies. Intention-to-treat was favoured over completer analysis. In cases of trials with more than two arms (Sofronoff et al., 2005; Reaven et al., 2018), we compared the most intensive arm (treatment) to the least intensive (control). The following clinician-rated outcome measures were acceptable and included in the meta-analysis: the Anxiety Diagnostic Interview Schedule (ADIS, Silverman & Albano 1996), the Pediatric Anxiety Rating Scale (PARS, RUPP, 2002), the Hamilton Rating Scale for Anxiety (HAM-A, Hamilton, 1959) and the Yale-Brown Obsessive Compulsive Scale (YBOCS, Goodman et al., 1989). Four studies (Storch et al., 2020; Storch et al., 2013; Storch et al., 2015; Wood et al., 2015) used multiple clinician-rated outcomes. Given this, we favoured primary outcome measures first (if reported in article or in protocol), followed by the most frequently used measure across studies (i.e., ADIS) to ensure consistency. Reaven et al. (2018) and Murphy et al. (2017) reported on individual symptoms on the ADIS, rather than the total, hence scores were combined. Acceptable parent/carer-rated outcome measures were the Spence Children’s Anxiety Scale (SCAS, Spence, 1998), the Multidimensional Anxiety Scale for Children (MASC, March, 1998), the Child and Adolescent Symptom Inventory-4 ASD Anxiety Scale (CASI-anx, Sukhodolsky et al., 2008), the Child Behaviour Checklist (CBCL, Achenbach & Rescorla, 2001) and the Children’s Obsessive Compulsive Inventory (CHOCI, Shafran et al., 2003). Child/self-rated outcome measures included the Spence Children’s Anxiety Scale (SCAS, Spence, 1998), the Revised Children’s Manifest Anxiety Scale (RCMAS, reference), the Revised Children’s Anxiety and Depression Scale (RCADS, Chorpita et al., 2005), the Obsessive Compulsive Inventory - Revised (OCI-R, Foa et al., 2002) and the Liebowitz Social Anxiety Scale (LSAS, Heimberg et al., 1999). One trial (Chalfant et al., 2007), used both the RCMAS and the SCAS, but the latter was favoured, as it was the most commonly used outcome measure. Storch et al. (2013) reported only subscales of the RCMAS, so the total mean was calculated.

###### Parent/carer-rater meta-analysis

There was a significant medium effect of CBT compared to control, including TAU, waitlist, counselling, adapted AM, and bespoke CBT (manual training only), in reducing parent/carer-ratings for anxiety symptoms at EOT (*k* = 12, *g* = 0.58 [95% CI 0.07, 1.08], *p* = .0246) (moderate-certainty evidence, Table S11), and significant heterogeneity, *Q*(11) = 70.39, *p* < .001, *I^2^* = 84.37%. After removal of outliers (Chalfant et al., 2007; Murphy et al., 2017; A. J. Russell et al., 2013), the effect size was still significant (*k* = 9, *g* = 0.44 [95% CI 0.21, 0.66], *p <* .001). Heterogeneity reduced, *Q*(8) = 4.99, *p* = .76, *I^2^* = 0%.

###### Clinician-rater meta-analysis

CBT had a significant small-to-medium effect on reducing clinician ratings for anxiety symptoms compared to control, including TAU, waitlist, counselling, and adapted AM (*k = 12, g* = 0.48 [95% CI 0.14, 0.81], *p* = .005) (moderate-certainty evidence, Table S11). There was significant heterogeneity, *Q*(11) = 35.13, p < .001, *I^2^* = 68.69%. On removal of outliers (J. Reaven et al., 2018; Storch et al., 2020), the effect size remained significant (*k* = 10, *g* = 0.44 [95% CI 0.24, 0.65], *p* < .001) and heterogeneity reduced, *Q*(9) = 8.58, *p* = .477, *I^2^* = 0%.

###### Meta-regression analyses

Bespoke CBT showed significantly worse clinician-rated anxiety at EOT compared to adapted CBT (*b* = −0.72 [95% CI –1.27, −0.18], *p* = .009), based on six bespoke against six adapted trials. There were no other significant moderators (Table S13).

Seven of the RCTs/pilot RCTs included in the meta-analyses evaluating CBT for anxiety reported non-anxiety outcomes (Kilburn et al., 2020; Langdon et al., 2016; A. J. Russell et al., 2013; Storch et al., 2013, 2015, 2020; White et al., 2013). Four indicated significant group differences in social functioning at EOT in favour of adapted/bespoke individual or combined individual and group CBT compared to non-active control (Storch et al., 2013, 2015, 2020; White et al., 2013). The remaining three, evaluating bespoke and adapted group CBT compared to non-active controls (Kilburn et al., 2020; Langdon et al., 2016), and adapted individual CBT for OCD compared to adapted AM (A. J. Russell et al., 2013) at EOT and follow-up, found no such effect. Two trials also showed no effect on depression (Langdon et al., 2016; A. J. Russell et al., 2013).

###### Studies not included in meta-analyses

Three pilot RCTs/RCTs that reported effectiveness outcomes for CBT for anxiety were not included in the meta-analysis due to having <10 participants per group (Fujii et al., 2013) or no EOT data (only follow-up) (Maskey, Rodgers, et al., 2019; White et al., 2015). They reported significant group differences in anxiety at EOT between adapted individual CBT and non-active control (Fujii et al., 2013), but no significant group differences in anxiety and social functioning at 6-months post-treatment between bespoke individual CBT and non-active control (Maskey, Rodgers, et al., 2019). While anxiety worsened over 1-year follow-up after treatment with bespoke individual and group CBT ended, it did not revert to pre-treatment severity 1-year post-treatment (White et al., 2015).

Two of the 36 papers reporting effectiveness outcomes were non-randomised controlled trials and reported an adapted/bespoke group CBT was effective for parent-reported CYP anxiety at EOT compared to non-active conditions (Hepburn et al., 2016; J. A. Reaven et al., 2009), but not for CYP-rated anxiety (J. A. Reaven et al., 2009).

Fifteen before-and-after comparisons examined the effectiveness of adapted/bespoke individual/group CBT for anxiety (Bemmer et al., 2021; Burke et al., 2017; Driscoll et al., 2020; Ehrenreich-May et al., 2014; Ekman et al., 2015; E. Higgins et al., 2019; Keefer et al., 2017; Kilburn et al., 2019; Maskey, McConachie, et al., 2019; Oerbeck et al., 2021; Ollendick et al., 2021; J. Reaven et al., 2015; J. Reaven, Blakeley-Smith, Leuthe, et al., 2012; Solish et al., 2020; Wise et al., 2019). Statistically significant improvements in outcomes over time were reported in 14 of these 15 studies (Bemmer et al., 2021; Driscoll et al., 2020; Ehrenreich-May et al., 2014; Ekman et al., 2015; E. Higgins et al., 2019; Keefer et al., 2017; Kilburn et al., 2019; Maskey, McConachie, et al., 2019; Oerbeck et al., 2021; Ollendick et al., 2021; J. Reaven et al., 2015; J. Reaven, Blakeley-Smith, Leuthe, et al., 2012; Solish et al., 2020; Wise et al., 2019). However, causality cannot be inferred since there were no comparison groups.

Considering all 36 papers that reported effectiveness outcomes of individual (Driscoll et al., 2020; Ehrenreich-May et al., 2014; Ekman et al., 2015; Fujii et al., 2013; Maskey, McConachie, et al., 2019; Maskey, Rodgers, et al., 2019; Oerbeck et al., 2021; Ollendick et al., 2021; A. J. Russell et al., 2013; Storch et al., 2013, 2015, 2020; Wise et al., 2019; Wood et al., 2015), group (Bemmer et al., 2021; Burke et al., 2017; Chalfant et al., 2007; Cook et al., 2017; Hepburn et al., 2016; E. Higgins et al., 2019; Keefer et al., 2017; Kilburn et al., 2019, 2020; Langdon et al., 2016; McConachie et al., 2014; J. A. Reaven et al., 2009; J. Reaven et al., 2015, 2018; J. Reaven, Blakeley-Smith, Culhane-Shelburne, et al., 2012; J. Reaven, Blakeley-Smith, Leuthe, et al., 2012; Sofronoff et al., 2005; Solish et al., 2020; Sung et al., 2011) and combined (Murphy et al., 2017; White et al., 2013, 2015) CBT for anxiety, which were synthesized narratively, moderate-certainty evidence suggested mixed results that these interventions may be helpful in reducing anxiety among autistic individuals (Table S10).

###### 2. Interventions targeting emotion regulation (n=5)

Three out of five papers evaluating adapted/bespoke group interventions targeting emotion regulation reported feasibility outcomes. Two papers separately reported that an adapted group CBT for autistic children aged 4-7 years with mental health difficulties (Factor et al., 2019) and a bespoke group mindfulness-based intervention for autistic children aged 7-12 years with mental health difficulties (Drüsedau et al., 2022) were feasible, based on low drop-out rates and high attendance. However, one paper found limited feasibility for a parent-delivered cognitive behavioural emotional and social skills intervention for autistic children as some parents reported difficulties with engaging their child to complete the programme, time constraints and interference of life events (Sofronoff et al., 2017). Two out of five papers reported on treatment satisfaction and showed the interventions were acceptable (Drüsedau et al., 2022; Swain et al., 2019). Children and parents reported they enjoyed and benefited from the group mindfulness-based intervention, although homework and having sessions on a weekly basis contributed to some dissatisfaction (Drüsedau et al., 2022). Parents noted that psychoeducation, support, and skills training components were helpful and reported high satisfaction with the adapted group CBT-based intervention for autistic children aged 4-8 years, with some parents reporting wanting more time for discussion and others noting some difficulties with generalisation of skills provided by the intervention (Swain et al., 2019).

Two out of five papers were RCTs. One did not statistically compare the two groups (Factor et al., 2019). The other was a pilot RCT, which preliminarily showed an adapted group CBT for children aged 5-7 years was not effective for emotion regulation but was effective for frequency of anger/anxiety episodes, use of emotion regulation strategies, and parent-reported perceived confidence in their child’s ability to manage their own anxiety and anger, all post-treatment compared to waitlist control (Scarpa & Reyes, 2011). The remaining three papers were before-and-after comparisons reporting on intervention effects over time (Drüsedau et al., 2022; Sofronoff et al., 2017; Swain et al., 2019), however, causality cannot be inferred since there were no comparison groups. Overall, low certainty evidence suggested mixed results regarding the effectiveness of some group interventions targeting emotion regulation to improve mental health of autistic CYP ((Drüsedau et al., 2022; Factor et al., 2019; Scarpa & Reyes, 2011; Sofronoff et al., 2017; Swain et al., 2019); Table S10).

###### 3. CBT for various mental health needs (n=3)

One paper examined therapists’ experiences of using CBT with autistic people and adaptations incorporated into their routine practice (Cooper et al., 2018). A range of adaptations were endorsed, including accommodating individual differences, changing the structure and content of interventions, and establishing communication preferences. Adaptations reported as being used less consistently included shorter/longer sessions, avoidance of metaphors and use of cognitive strategies. Most participants reported using CBT and favoured this approach over others.

Two papers reported on pilots of bespoke CBT group interventions for stress, anxiety and depression in CYP and adult, evaluated through a non-randomised trial (McGillivray & Evert, 2014), and depression in CYP through an RCT (Santomauro et al., 2016). Both interventions appeared to be feasible based on low drop-out rates. One paper examined acceptability and found most participants reported they enjoyed the intervention, finding the group setting most helpful, but with variations in which intervention tools were most helpful to manage depression (Santomauro et al., 2016). There were no statistically significant differences in depression between the CBT interventions and waitlist controls post-intervention (McGillivray & Evert, 2014; Santomauro et al., 2016). However, participants who had scored above the depression, anxiety and stress threshold pre-intervention benefited the most from the intervention (McGillivray & Evert, 2014). At three and nine month follow up, gains were sustained in one study (McGillivray & Evert, 2014) but not the other (Santomauro et al., 2016). Overall, moderate certainty evidence suggested mixed findings regarding the effectiveness of some CBT for anxiety, stress, and depression to improve mental health of autistic CYP ((McGillivray & Evert, 2014; Santomauro et al., 2016); Table S10).

###### 4. EMDR for post-traumatic stress disorder (PTSD) (n=1)

One Delphi study gathered perspectives on EMDR for autistic people and the adaptations therapists incorporated to standard protocols (Fisher et al., 2023). Participants reported tailoring therapy from the assessment onwards, such as by adopting a flexible and creative approach, adjusting the environment to suit sensory preferences, communicating clearly, taking more time in initial phases and before active processing commenced, and acknowledging the contribution of autism to the formulation.

#### Predictors of outcome

Five studies explored relevant predictors of treatment outcome, such as demographic variables, autism symptomatology, verbal Intelligence Quotient. Only parental trait anxiety showed an effect on change in child anxiety in one study ((White et al., 2015); Table S14).

## Discussion

This systematic review and meta-analysis explored strategies implemented within mental health services to improve mental health care for autistic CYP. Overall, 57 papers were included. Most tested CBT-based interventions.

Few studies identified service-level strategies largely related to increasing detection and knowledge of autism, and skills in working with autistic people in mental health services through screening tools, specialised care pathways, professional networks, and service-wide general adaptations. Most of the interventions comprised CBT for anxiety, with a few targeting emotion regulation and depression. Reported adaptations involved environmental and communication accommodations, accommodating individual differences, structural/procedural adaptations, intervention content adaptations, and engaging wider support networks (e.g., parents). However, we identified a lack of thorough description of the adaptations made and the rationale for use. Additionally, some adaptations reported, such as those relating to accommodating individual differences, are part of general good clinical practice rather than only specific to autism.

This review, together with our review on autistic adults (Loizou et al., 2023), shows a similar pattern of most papers reporting communication and intervention content adaptations, but few reporting environmental adjustments. Parental and school involvement was an adaptation category specifically relevant to CYP, but evidence on CYP views of such involvement was lacking. While the studies reporting involvement with the child’s school did not evaluate this aspect, one study indicated that children were satisfied with the treatment overall (Wood et al., 2015). Parental involvement was evaluated only in one trial, which showed parent groups and training parents as co-therapists to be involved in all aspects of CBT for anxiety enhanced the usefulness of the intervention when compared to a child-only group with minimal parental involvement (Sofronoff et al., 2005). A previous study reported friends and family were frequently used as sources of support by young autistic people with co-occurring mental health difficulties. However, participants with more severe mental health difficulties reported being reluctant to talk about their needs to friends and family due to stigma (Crane et al., 2018).

The identified bespoke interventions originally developed for autistic CYP were mostly based on CBT principles, with one being mindfulness-based. Most targeted anxiety, and a few emotion regulation, depression, or combination of mental health difficulties. Notably, there were no eligible studies investigating pharmacological interventions, although these are increasingly used for autistic CYP (Bachmann, Manthey, Kamp-Becker, Glaeske, & Hoffmann, 2013; Coury et al., 2012; Deb et al., 2021; Jobski, Höfer, Hoffmann, & Bachmann, 2017; Murray et al., 2014) despite limited evidence of effectiveness (Jobski et al., 2017) and clinical guidelines recommending caution when prescribing them for autistic CYP (NICE, 2021). Additionally, only one study evaluated telemental health, despite it being increasingly used in and since the pandemic (Appleton et al., 2021). Notably, only two studies included participants with co-occurring ID.

Evidence on effectiveness was of higher quality than in the review relating to adults (Loizou et al., 2023), due to more RCTs having been conducted. Nonetheless, the certainty of evidence for effectiveness, based on the GRADE system rating (Guyatt et al., 2008), ranged from low to moderate (Table S10 and S11), meaning further research is likely to significantly impact our confidence in the findings. The effectiveness results for service-level strategies suggest some screening tools may be helpful in identifying autism and clinician training may improve mental health care. The exploratory meta-analyses examining whether CBT for anxiety was superior to any comparison group in reducing anxiety symptoms severity at EOT among autistic individuals, showed no significant group differences in improving child/self-rated anxiety, significant medium effect of CBT on reducing parent/carer-rated anxiety, and significant small-to-medium effect on decreasing clinician-rated anxiety (all moderate-certainty evidence). Importantly, the presence of publication bias was detected, which warrants caution. However, it should be noted that upon removal of outliers, effect sizes remained relatively stable, suggesting bespoke/adapted CBT for anxiety may be effective in reducing anxiety in autistic CYP when viewed from the perspective of parents/carers or clinicians, but not of children/care recipients, compared to the active and non-active controls.

Inconsistencies in effect sizes with previous meta-analyses may be attributed to methodological differences such as inclusion criteria (e.g., some included non-adapted generic CBT (Sharma et al., 2021) and non-randomised controlled trials (Perihan et al., 2020; Ung, Selles, Small, & Storch, 2015)) or favouring different measures (Kreslins, Robertson, & Melville, 2015; Sharma et al., 2021; Sukhodolsky et al., 2013) and chosing pooled over separate meta-analyses (Perihan et al., 2020; Ung et al., 2015; Wichers, Van Der Wouw, Brouwer, Lok, & Bockting, 2023). Variation of effect sizes across different raters has been reported elsewhere (Kreslins et al., 2015; Sharma et al., 2021; Sukhodolsky et al., 2013). CYP often differ in reporting symptom severity in contrast to parents and clinicians, with higher correspondence reported for assessments of observable mental health concerns, assessments made by informants observing the child in the same setting, and for assessments using dimensional measures comparative to categorical ones (De Los Reyes et al., 2015; Smith, 2007). Rater blinding and observer bias could be influential factors contributing to informant discrepancies (Hróbjartsson, Emanuelsson, Thomsen, Hilden, & Brorson, 2014). A variety of anxiety measures were used, however not all are supported for use with autistic individuals (Glod et al., 2017; Jitlina et al., 2017; Lecavalier et al., 2014; May, Cornish, & Rinehart, 2015), particularly without adjustments.

The only significant moderator for effectiveness was type of CBT, with adapted CBT being superior to bespoke CBT in reducing clinician-rated anxiety in autistic individuals based on equal number of contributing bespoke and adapted trials. This should be carefully interpreted as the study quality of the bespoke trials appears slightly lower than that of the adapted trials, and the distinction between adapted and bespoke CBT potentially lacks robustness. Comparing group, individual and combined CBT showed no significant difference in parent- and clinician-rated anxiety in autistic individuals counter to another meta-analysis (Sharma et al., 2021), where significant advantage was found for individual CBT for clinician ratings. However, this difference could be due to the uneven number of trials contributing to each intervention format (Sharma et al., 2021).

Evidence for feasibility and acceptability, although sometimes not involving rigorous formal measurement strategies, was largely positive, similarly to the review relating to adults (Loizou et al., 2023). Clinicians reported using a range of service-level adaptations related to the physical environment, communication, accommodating individual needs, and a more structured and predictable approach, suggesting these can be implemented in routine clinical services (Jones et al., 2021; Petty et al., 2021; Spain et al., 2017). Additionally, a tele-mentoring platform to support mental health clinicians was evaluated as acceptable and feasible (Dreiling et al., 2022). The identified mental health interventions were also evaluated as acceptable and feasible. Adaptations concerning communication, intervention structure and content, and accommodating individual preferences were reported as often incorporated by clinicians when delivering CBT and EMDR to autistic people, supporting feasibility (Cooper et al., 2018; Fisher et al., 2023). Limited feasibility based on high drop-out rate was noted only for a parent-delivered behavioural emotional and social skills intervention as some parents struggled to implement the intervention (Sofronoff et al., 2017). A trial found the type of clinician training for bespoke group CBT for anxiety can affect autistic CYP, parent, and clinician acceptability (Walsh et al., 2018). Clinicians’ view of the intervention’s effectiveness, fit with existing service, ease of use and making further adaptations were reported as important factors that facilitated the feasibility of sustained use of a group CBT with autistic adults (Pickard et al., 2020).

### Strengths and limitations

This systematic review provides a comprehensive overview of strategies/adaptations tested in mental health care settings for autistic CYP, which is potentially useful for adapting care to the specific needs of this population. A strength was the co-produced nature of this review, with lived experience researchers involved throughout the project. A novel feature of this research is the inclusion of AIRA assessing the primary research’s inclusive practices that we hope will encourage appropriate adaptations in future research involving this population.

Regarding limitations of our review, the meta-analyses lacked inclusion of follow-up outcomes to determine if treatment gains are sustained over time, although few trials measured these. Additionally, our classification of ‘bespoke’ versus ‘adapted’ interventions depended on authors’ descriptions, which may lack robustness if evaluated against independently-rated criteria.

There were several limitations of the included studies that limited the quality of the review. Most autistic participants were male and white, and all studies were conducted in high-income countries, limiting the generalisability of findings. Evidence on effective strategies for autistic individuals with ID was lacking, indicating a potential bias in the selection of study participants (G. Russell et al., 2019). There was a lack of different types of strategies to CBT, such as pharmacological interventions or other psychological approaches, and there were noticeable gaps of interventions targeting other mental health difficulties besides anxiety. The included RCTs had small sample sizes (the mean CBT participants was 21 and controls 20) and probably lacked statistical power to detect significant group differences. Thus, large, high-quality studies with the potential to shape practice were missing. Importantly, many of the included papers lacked a comparison group, preventing improvements being credited to the intervention alone. Where comparisons were included, none were with a non-adapted version of the same intervention, limiting inferences about effectiveness of adaptations. There was a lack of clarity in the intervention-level adaptations reported in some papers limiting replication in further research and implementation in practice. Assessing the included papers with the novel and lived experience researcher-led AIRA showed co-produced research with involvement from autistic individuals and carers was missing, data collection methods and outcome measures often lacked autism inclusivity, and some interventions, of which most were CBT, appeared to involve some focus on masking autistic traits rather than improving mental health.

### Clinical implications and future directions

Better mental health care is a top priority for autistic people (Cusack & Sterry, 2016) and recognised as such by the World Health Organisation (World Health Assembly, 2014), the National Health Service (NHS) Long-Term Plan (NHS, 2019), and the NHS Autism Research Strategy (NHS, 2022). Autistic people experience high rates of mental health difficulties but face many barriers to accessing and benefiting from mental health care. This systematic review provided a list of strategies and adaptations to services and interventions found to be acceptable and feasible to implement in mental health care. Many of the identified adaptations (Table 2 and S9) are simple, reasonable adjustments not necessarily requiring further evaluation to be implemented in practice, or specific iterations of general good clinical practice. Tailoring mental health care to individual differences may be especially helpful in achieving effective mental health care, as autistic individuals vary in their support needs and presentation of autistic characteristics (Robledo, Donnellan, & Strandt-Conroy, 2012; Uljarević et al., 2017). A neurodivergence-informed approach to therapy (Chapman & Botha, 2023) and primary co-produced research to strengthen the evidence-base are necessary to strike a balance between personalising care and following evidence-based practice.

More research is needed to improve autism assessment so autistic CYP can benefit from mental health care. For more robust intervention research in this field, it is important to develop consensus that includes involvement of autistic people, on appropriate mental health outcome measures, meaningful treatment gain, and likelihood of the intervention to encourage masking over genuine mental health improvements. Identified differences in the magnitude and significance of effect sizes by rater (self, parent, clinician) suggest that pooling data across raters should be avoided. This review also identified a need for studies comparing autism-friendly adapted interventions and generic non-adapted interventions received in mental health services.

This review together with our review relating to autistic adults (Loizou et al., 2023) can contribute to national guidelines for mental health provision for autistic people to be tested in a small number of well-funded research projects prioritising co-production and ensuring participation from under-represented groups.

### Lived Experience Commentary written by Robin Jackson and Eva Driskell

We have lived experience as an autistic young person, now a Child and Adolescent Mental Health Services (CAMHS) professional, and as a carer of a child with autism and have spent a decade as service users of CAMHS. The examples we use reflect both of our lived experiences.

The review found that better training is required for CAMHS professionals to see the signs of autism earlier. Our experience supports this along with the need to fast track children to a diagnosis. Children with autism are left in mental distress and miss out on crucial years of education and parents and carers find themselves blamed by the educational establishment (and often the medical professionals) for what is seen as bad behaviour rather than a distressed child in need of help.

The review focused on strategies delivered in mental health service settings, but outside of the home, school is where children spend the most time. Evidence of school or parental involvement was lacking, but strategies could be delivered in a school setting alongside training for education professionals to recognise autistic traits and understand sensory issues. We experienced a huge improvement in mental health issues when the right adjustments were made, which also raises the question of whether mental health issues were co-occurring or caused by a lack of understanding and an over-stimulating environment.

Training and understanding of autism for children and young people (CYP) and carers is invaluable and not widely available. Personalised training helped us - a short in-person group course was very useful in explaining the issues and allowed the sharing of experiences. However, consideration of accessibility is needed, including aspects such as language, socio-economic background, and culture.

Neurotypical bias was evident in the research when the views of CYP were dismissed if their self-reported outcome measures found less improvement than clinician or parent reports meaning that the therapies could inadvertently encourage masking, which occurs when autistic traits are hidden to fit in with other people. Using cognitive behavioural therapy could result in autistic CYP learning to mask their traits to better ‘fit in’. Although this is measured as success in the papers reviewed, in the long term it can cause further distress and mental health issues.

More research is needed into the causes of mental health issues in autistic CYP and into how diagnosis, training of CYP and carers and environmental adjustments affect mental health. The review found that only 7% of research was carried out with autistic researchers involved. Future research should include autistic researchers to investigate which therapies work best for autistic CYP and develop new therapeutic approaches to be more effective for this population. Unfortunately, current therapies can be focused on changing the autistic young person’s behaviour to fit in better with neurotypical society rather than improving the autistic young person’s mental health.

## Acknowledgements

Autistica.

## Data Availability

Data were collected from publicly available research papers which are referenced.

## Author contributions

The working group collaboratively conceived and formulated the review questions. TS conducted the searches. The selection strategy was piloted by TS, PB. AK, TS, KS, AG, TP, UF conducted the title and abstract screening. TP, SL independently reviewed a random 10% of records. Full texts were screened independently in duplicate (TP, AG, AK, TS, DS, KS, SL, RC, JG, HB, UF) and discrepancies were resolved by discussion with a third reviewer (SJ). TP, SL, AG, AK, DS, RA, MM, GB, HB, JG, RC, UF extracted data. SL, RC piloted the data extraction form. Supported by PB, SL and TP independently double-extracted raw outcome data for the meta-analysis. SL ran the meta-analyses, supported by PB. RRO developed the Autism-Inclusive Research Assessment (AIRA), AK, RA extracted all relevant data for the AIRA, and RJ was involved as second assessor for the final criterion. TP, SL, AG, AK, DS, MM, GB, HB, JG, RC, UF assessed study quality. TP, SL, RA contributed to the evaluation of the strength of evidence about intervention effectiveness using the Grading of Recommendations Assessment, Development and Evaluation (GRADE) system. TP, SL, RA, DS collaboratively drafted the manuscript. All authors reviewed and contributed to the editing of the manuscript. All authors have approved the final manuscript.

