## Supplementary Material for "Approaches to improve mental health care for autistic children and young people: a systematic review and meta-analysis"

**Table S1** PRISMA checklist

| **Section and Topic** | **Item #** | **Checklist item** | **Location where item is reported** |
| --- | --- | --- | --- |
| **TITLE** | | |  |
| Title | 1 | Identify the report as a systematic review. | Page 1 |
| **ABSTRACT** | | |  |
| Abstract | 2 | See the PRISMA 2020 for Abstracts checklist. | Page 2 |
| **INTRODUCTION** | | |  |
| Rationale | 3 | Describe the rationale for the review in the context of existing knowledge. | Pages 3-4 |
| Objectives | 4 | Provide an explicit statement of the objective(s) or question(s) the review addresses. | Pages 3-4 |
| **METHODS** | | |  |
| Eligibility criteria | 5 | Specify the inclusion and exclusion criteria for the review and how studies were grouped for the syntheses. | Pages 4-7 |
| Information sources | 6 | Specify all databases, registers, websites, organisations, reference lists and other sources searched or consulted to identify studies. Specify the date when each source was last searched or consulted. | Page 4 |
| Search strategy | 7 | Present the full search strategies for all databases, registers and websites, including any filters and limits used. | Tables S2-S4 in supplementary |
| Selection process | 8 | Specify the methods used to decide whether a study met the inclusion criteria of the review, including how many reviewers screened each record and each report retrieved, whether they worked independently, and if applicable, details of automation tools used in the process. | Pages 4-7 |
| Data collection process | 9 | Specify the methods used to collect data from reports, including how many reviewers collected data from each report, whether they worked independently, any processes for obtaining or confirming data from study investigators, and if applicable, details of automation tools used in the process. | Pages 5-7 |
| Data items | 10a | List and define all outcomes for which data were sought. Specify whether all results that were compatible with each outcome domain in each study were sought (e.g. for all measures, time points, analyses), and if not, the methods used to decide which results to collect. | Pages 4-7 |
|  | 10b | List and define all other variables for which data were sought (e.g. participant and intervention characteristics, funding sources). Describe any assumptions made about any missing or unclear information. | Pages 4-7 |
| Study risk of bias assessment | 11 | Specify the methods used to assess risk of bias in the included studies, including details of the tool(s) used, how many reviewers assessed each study and whether they worked independently, and if applicable, details of automation tools used in the process. | Page 6 |
| Effect measures | 12 | Specify for each outcome the effect measure(s) (e.g. risk ratio, mean difference) used in the synthesis or presentation of results. | Pages 6-7 |
| Synthesis methods | 13a | Describe the processes used to decide which studies were eligible for each synthesis (e.g. tabulating the study intervention characteristics and comparing against the planned groups for each synthesis (item #5)). | Pages 6-7 |
|  | 13b | Describe any methods required to prepare the data for presentation or synthesis, such as handling of missing summary statistics, or data conversions. | Pages 6-7 |
|  | 13c | Describe any methods used to tabulate or visually display results of individual studies and syntheses. | Pages 6-7 |
|  | 13d | Describe any methods used to synthesize results and provide a rationale for the choice(s). If meta-analysis was performed, describe the model(s), method(s) to identify the presence and extent of statistical heterogeneity, and software package(s) used. | Pages 6-7 |
|  | 13e | Describe any methods used to explore possible causes of heterogeneity among study results (e.g. subgroup analysis, meta-regression). | Pages 6-7 |
|  | 13f | Describe any sensitivity analyses conducted to assess robustness of the synthesized results. | Pages 6-7 |
| Reporting bias assessment | 14 | Describe any methods used to assess risk of bias due to missing results in a synthesis (arising from reporting biases). | Pages 6-7 |
| Certainty assessment | 15 | Describe any methods used to assess certainty (or confidence) in the body of evidence for an outcome. | Page 6, Tables S10-S11 in supplementary |
| **RESULTS** | | |  |
| Study selection | 16a | Describe the results of the search and selection process, from the number of records identified in the search to the number of studies included in the review, ideally using a flow diagram. | Figure 1 |
|  | 16b | Cite studies that might appear to meet the inclusion criteria, but which were excluded, and explain why they were excluded. | Table S5 |
| Study characteristics | 17 | Cite each included study and present its characteristics. | Page 7-9, Table 1, tables S6 and S12 |
| Risk of bias in studies | 18 | Present assessments of risk of bias for each included study. | Page 15, Table S7 |
| Results of individual studies | 19 | For all outcomes, present, for each study: (a) summary statistics for each group (where appropriate) and (b) an effect estimate and its precision (e.g. confidence/credible interval), ideally using structured tables or plots. | Figure 2, Table 3, Table S12 and S13 in supplementary |
| Results of syntheses | 20a | For each synthesis, briefly summarise the characteristics and risk of bias among contributing studies. | Pages 7-18, Table 3, Table S12 in supplementary |
|  | 20b | Present results of all statistical syntheses conducted. If meta-analysis was done, present for each the summary estimate and its precision (e.g. confidence/credible interval) and measures of statistical heterogeneity. If comparing groups, describe the direction of the effect. | Pages 25-28, Figure 2, Table S13 |
|  | 20c | Present results of all investigations of possible causes of heterogeneity among study results. | Pages 25-28 |
|  | 20d | Present results of all sensitivity analyses conducted to assess the robustness of the synthesized results. | Pages 25-28 |
| Reporting biases | 21 | Present assessments of risk of bias due to missing results (arising from reporting biases) for each synthesis assessed. | Table S7 |
| Certainty of evidence | 22 | Present assessments of certainty (or confidence) in the body of evidence for each outcome assessed. | Pages 18-30, Table 3, Tables S10-S11 in supplementary |
| **DISCUSSION** | | |  |
| Discussion | 23a | Provide a general interpretation of the results in the context of other evidence. | Page 30-34 |
|  | 23b | Discuss any limitations of the evidence included in the review. | Page 32-33 |
|  | 23c | Discuss any limitations of the review processes used. | Page 32-33 |
|  | 23d | Discuss implications of the results for practice, policy, and future research. | Pages 33-34 |
| **OTHER INFORMATION** | | |  |
| Registration and protocol | 24a | Provide registration information for the review, including register name and registration number, or state that the review was not registered. | Page 4 |
|  | 24b | Indicate where the review protocol can be accessed, or state that a protocol was not prepared. | Page 4 |
|  | 24c | Describe and explain any amendments to information provided at registration or in the protocol. | Not applicable |
| Support | 25 | Describe sources of financial or non-financial support for the review, and the role of the funders or sponsors in the review. | Page 35 |
| Competing interests | 26 | Declare any competing interests of review authors. | Page 35 |
| Availability of data, code and other materials | 27 | Report which of the following are publicly available and where they can be found: template data collection forms; data extracted from included studies; data used for all analyses; analytic code; any other materials used in the review. | Page 35 |

**Table S2.** Ovid MEDLINE(R) search

|  | Query | Hits on 20/07/22 |
| --- | --- | --- |
| 1 | (autis* or Asperger* or ASC or ASD or PDD or pervasive development*).ab,kw,ti. | 84191 |
| 2 (MeSH) | Autistic Disorder/ or Autism Spectrum Disorder/ or Child Development Disorders, Pervasive/ or Asperger Syndrome/ | 44574 |
| 3  (MeSH) | Rett Syndrome/ | 2860 |
| 4 | 1 OR 2 OR 3 | 90125 |
| 5 | (mental health service* or health service* or hospital or GP or ward or inpatient* or "community mental health" or home treatment or crisis resolution or "child and adolescent mental health service" or CAMHS or CBT or "cognitive behavio* therap*" or DBT or "dialectical behavio* therap*" or family therapy or interpersonal therap* or psychodynamic or treatment* or intervention* or occupational therapy or mindfulness or psychological intervention* or psychological therap* or behavio* therap* or psychotherap* or "acceptance and commitment therap*" or ACT).ab,kw,ti. | 7210419 |
| 6 | (mental health or mental illness* or mental disorder* or mental condition* or anxi* or affect or depress* or behavio* problems or eating disorder*).ab,kw,ti. | 919484 |
| 7  (MeSH) | mental disorders/ or anxiety disorders/ or "bipolar and related disorders"/ or "disruptive, impulse control, and conduct disorders"/ or dissociative disorders/ or elimination disorders/ or "feeding and eating disorders"/ or mood disorders/ or neurocognitive disorders/ or neurotic disorders/ or personality disorders/ or "schizophrenia spectrum and other psychotic disorders"/ or substance-related disorders/ or "trauma and stressor related disorders"/ | 373099 |
| 8 | 6 OR 7 | 1142253 |
| 9 | 4 and 5 and 8 | 5262 |
| 10 | limit 9 to yr="1994 -Current" | 5073 |

**Table S3.** APA PsycINFO search

|  | Query | Hits on 20/07/22 |
| --- | --- | --- |
| 1  (MeSH) | autism spectrum disorders/ or neurodevelopmental disorders/ or autistic traits/ or developmental disabilities/ or rett syndrome/ | 65622 |
| 2 | (autis* or Asperger* or ASC or ASD or PDD or pervasive development*).ab,hw,id,ti. | 65688 |
| 3 | 1 OR 2 | 79011 |
| 4 | ("mental health" or "mental illness*" or "mental disorder*" or "mental condition*" or anxi* or affective or depress* or "behavio* problems" or "eating disorder*" or psychosis or schizophrenia or "psychotic disorder*").ab,hw,id,ti. | 998005 |
| 5  (MeSH) | mental disorders/ or affective disorders/ or anxiety disorders/ or bipolar disorder/ or borderline states/ or chronic mental illness/ or dissociative disorders/ or eating disorders/ or mental disorders due to general medical conditions/ or neurocognitive disorders/ or neurosis/ or personality disorders/ or psychosis/ or serious mental illness/ or sleep wake disorders/ or somatoform disorders/ or "stress and trauma related disorders"/ or "substance related and addictive disorders"/ | 235303 |
| 6 | 4 OR 5 | 1030648 |
| 7 | ("mental health service*" or "health service*" or hospital or GP or ward or inpatient* or "community mental health" or "home treatment" or "crisis resolution" or "child and adolescent mental health service" or CAMHS or CBT or "cognitive behavio* therap*" or DBT or "dialectical behavio* therap*" or "family therap*" or "interpersonal therap*" or psychodynamic or treatment* or intervention* or "occupational therap*" or mindfulness or "psychological intervention*" or "psychological therap*" or "behavio* therap*" or psychotherap* or "acceptance and commitment" or ACT).ab,hw,id,ti. | 1399839 |
| 8 | 3 AND 6 AND 7 | 9268 |
| 9 | limit 8 to yr="1994 -Current" | 8349 |

**Table S4.** CINAHL Plus search

|  | Query | Hits on 20/07/22 |
| --- | --- | --- |
| 1  (MeSH) | (MH "Autistic Disorder") OR (MH "Rett Syndrome") OR (MH "Developmental Disabilities") OR (MH "Asperger Syndrome") | 25,117 |
| 2 | autis* or Asperger* or ASC or ASD or PDD or pervasive development* | 25,938 |
| 3 | 1 OR 2 | 30,774 |
| 4  (MeSH) | (MH "Mental Disorders") OR (MH "Neurotic Disorders") OR (MH "Affective Disorders") OR (MH "Anxiety Disorders") OR (MH "Dissociative Disorders") OR (MH "Factitious Disorders") OR (MH "Somatoform Disorders") OR (MH "Personality Disorders") OR (MH "Psychotic Disorders") OR (MH "Substance Use Disorders") OR (MH "Psychological Trauma") OR (MH "Adjustment Disorders") OR (MH "Behavioral Symptoms") OR (MH "Behavioral and Mental Disorders") | 149,139 |
| 5 | "mental health" or "mental illness*" or "mental disorder*" or "mental condition*" or anxi* or affect or depress* or "behavio* problems" or "eating disorder*" or psychosis or schizophrenia or "psychotic disorder*" | 264,382 |
| 6 | 4 OR 5 | 362,727 |
| 7 | "mental health service*" or "health service*" or hospital or GP or ward or inpatient* or "community mental health" or "home treatment" or "crisis resolution" or "child and adolescent mental health service" or CAMHS or CBT or "cognitive behavio* therap*" or DBT or "dialectical behavio* therap*" or "family therap*" or "interpersonal therap*" or psychodynamic or treatment* or intervention* or "occupational therap*" or mindfulness or "psychological intervention*" or "psychological therap*" or "behavio* therap*" or psychotherap* or "acceptance and commitment" or ACT | 1,105,132 |
| 8 | 3 AND 6 AND 7 (**Limiters:** 1994-2022; Exclude MEDLINE records) | 2,344 |

**Table S5.** List of studies excluded at full-text screening and reasons for exclusion (n=479)

| **Reference** | **Reason for exclusion** |
| --- | --- |
| Odiyoor M, Tromans SJ, Alexander RT, Akbari S, Bell G, Bering S, et al. The role of specialist inpatient rehabilitation services for people with intellectual disability, autism and mental health, behavioural or forensic needs. Adv Ment Health Intellect Disabil. 2019 Sep 4;13(5):204–15. | Wrong publication/study design |
| McConachie H, Hoole S, Le Couteur AS. Improving mental health transitions for young people with autism spectrum disorder. Child Care Health Dev [Internet]. 2011 Nov 1 [cited 2023 Oct 13];37(6):764–6. Available from: https://onlinelibrary.wiley.com/doi/full/10.1111/j.1365-2214.2011.01238.x | Wrong publication/study design |
| Laugeson EA, Park MN. Using a CBT Approach to Teach Social Skills to Adolescents with Autism Spectrum Disorder and Other Social Challenges: The PEERS® Method. Journal of Rational - Emotive and Cognitive - Behavior Therapy [Internet]. 2014 Feb 5 [cited 2023 Oct 13];32(1):84–97. Available from: https://link.springer.com/article/10.1007/s10942-014-0181-8 | Wrong publication/study design |
| Lalanne L, Weiner L, Bertschy G. Treatment of Addiction in Adults with Autism Spectrum Disorder. 2017;377–95. | Wrong publication/study design |
| Doyen C, Goupil V, Desailly E, Oreve MJ, Kaye K. Télémédecine et troubles du spectre de l’autisme de l’enfant et de l’adolescent : guide théorique et pratique. Annales Médico-psychologiques, revue psychiatrique. 2019 Sep;177(7):702–9. | Wrong publication/study design |
| Shaw SR, Bruce J, Ouimet T, Sharma A, Glaser S. Young children with developmental disabilities and atypical antipsychotic medications: Dual diagnosis, direction, and debate. J Early Child Infant Psychol [Internet]. 2009;5:37–55. Available from: http://ovidsp.ovid.com/ovidweb.cgi?T=JS&PAGE=reference&D=psyc6&NEWS=N&AN=2010-01989-003 | Wrong publication/study design |
| Turan B, Kocarslan N, Gulsen M, Dursun OB. Your country is your routine: the evacuation, quarantine, and management of behavioral problems of a child with autism during COVID-19 pandemic. Dusunen Adam: Journal of Psychiatry and Neurological Sciences [Internet]. 2020;33(3):310–2. Available from: http://ovidsp.ovid.com/ovidweb.cgi?T=JS&PAGE=reference&D=psyc18&NEWS=N&AN=2021-15713-009 | Wrong publication/study design |
| Attwood T. Working with individuals on the spectrum. A spectrum of solutions for clients with autism: Treatment for adolescents and adults [Internet]. 2020;3–13. Available from: http://ovidsp.ovid.com/ovidweb.cgi?T=JS&PAGE=reference&D=psyc18&NEWS=N&AN=2020-16852-001 | Wrong publication/study design |
| Loomes R, Bryant-Waugh R. Widening the reach of family-based interventions for Anorexia Nervosa: autism-adaptations for children and adolescents. J Eat Disord [Internet]. 2021;9(1):157. Available from: http://ovidsp.ovid.com/ovidweb.cgi?T=JS&PAGE=reference&D=pmnm5&NEWS=N&AN=34863292 | Wrong publication/study design |
| Withrow NA. Why so picky? Food selectivity and how it impacts eating. A spectrum of solutions for clients with autism: Treatment for adolescents and adults [Internet]. 2020;173–9. Available from: http://ovidsp.ovid.com/ovidweb.cgi?T=JS&PAGE=reference&D=psyc18&NEWS=N&AN=2020-16852-022 | Wrong publication/study design |
| Schoultz P. Who responds to cognitive-behavioral group treatment? Associations between anxiety symptom reduction and autism symptom domains. Dissertation Abstracts International: Section B: The Sciences and Engineering [Internet]. 2017;77(11):No-Specified. Available from: http://ovidsp.ovid.com/ovidweb.cgi?T=JS&PAGE=reference&D=psyc14&NEWS=N&AN=2016-58393-015 | Wrong publication/study design |
| Campillo C, Herrera G, de Ganuza CR, Cuesta JL, Abellan R, Campos A, et al. Using Tic-Tac software to reduce anxiety-related behaviour in adults with autism and learning difficulties during waiting periods: A pilot study. Autism [Internet]. 2014;18(3):264–71. Available from: http://ovidsp.ovid.com/ovidweb.cgi?T=JS&PAGE=reference&D=psyc11&NEWS=N&AN=2014-11084-007 | Wrong publication/study design |
| Kurtz T. Utilizing exercise to treat symptoms of anxiety in autism spectrum disorder. Dissertation Abstracts International: Section B: The Sciences and Engineering [Internet]. 2018;79(12):No-Specified. Available from: http://ovidsp.ovid.com/ovidweb.cgi?T=JS&PAGE=reference&D=psyc15&NEWS=N&AN=2018-48576-058 | Wrong publication/study design |
| Koegel LK, Navab A, Ashbaugh K, Koegel RL. Using reframing to reduce negative statements in social conversation for adults with autism spectrum disorder. J Posit Behav Interv [Internet]. 2016;18(3):133–44. Available from: http://ovidsp.ovid.com/ovidweb.cgi?T=JS&PAGE=reference&D=psyc13&NEWS=N&AN=2016-29836-002 | Wrong publication/study design |
| Chapman R, Evans B. Using art-based Acceptance and Commitment Therapy (ACT) for an adolescent with anxiety and autism. Clin Case Stud [Internet]. 2020;19(6):438–55. Available from: http://ovidsp.ovid.com/ovidweb.cgi?T=JS&PAGE=reference&D=psyc17&NEWS=N&AN=2020-76780-003 | Wrong publication/study design |
| Gerhardt L, Smith J. The use of minecraft in the treatment of trauma for a child with autism spectrum disorder. J Fam Ther [Internet]. 2020;No-Specified. Available from: http://ovidsp.ovid.com/ovidweb.cgi?T=JS&PAGE=reference&D=psyc17&NEWS=N&AN=2020-36735-001 | Wrong publication/study design |
| Bridges S. Use of a learning disabilities and autism toolkit in mental health care. Nurs Times [Internet]. 2019;115(9):55–8. Available from: https://search.ebscohost.com/login.aspx?direct=true&AuthType=ip,shib&db=jlh&AN=138703495&site=ehost-live&scope=site&custid=s8454451 | Wrong publication/study design |
| Kuriakose S, Lahiri U. Understanding the Psycho-Physiological Implications of Interaction With a Virtual Reality-Based System in Adolescents With Autism: A Feasibility Study. IEEE Trans Neural Syst Rehabil Eng [Internet]. 2015;23(4):665–75. Available from: http://ovidsp.ovid.com/ovidweb.cgi?T=JS&PAGE=reference&D=med12&NEWS=N&AN=25643409 | Wrong publication/study design |
| Aman MG. Treatment planning for patients with autism spectrum disorders. J Clin Psychiatry [Internet]. 2005;66:38–45. Available from: http://ovidsp.ovid.com/ovidweb.cgi?T=JS&PAGE=reference&D=med6&NEWS=N&AN=16401149 | Wrong publication/study design |
| Watanabe T. Treatment of Major Depressive Disorder with Autism Spectrum Disorder by Acceptance and Commitment Therapy Matrix. Case Rep Psychiatry [Internet]. 2021;2021:5511232. Available from: http://ovidsp.ovid.com/ovidweb.cgi?T=JS&PAGE=reference&D=pmnm5&NEWS=N&AN=33880197 | Wrong publication/study design |
| Lin CE, Wood JJ, Storch EA, Sze KM. Treatment of childhood anxiety in autism spectrum disorders. Handbook of treating variants and complications in anxiety disorders [Internet]. 2013;83–95. Available from: http://ovidsp.ovid.com/ovidweb.cgi?T=JS&PAGE=reference&D=psyc10&NEWS=N&AN=2013-41454-005 | Wrong publication/study design |
| Reaven J. The treatment of anxiety symptoms in youth with high-functioning autism spectrum disorders: Developmental considerations for parents. Brain Res [Internet]. 2011;1380:255–63. Available from: http://ovidsp.ovid.com/ovidweb.cgi?T=JS&PAGE=reference&D=psyc8&NEWS=N&AN=2010-21992-001 | Wrong publication/study design |
| Ehrenreich-May J, Simpson G, Stewart LM, Kennedy SM, Rowley AN, Beaumont A, et al. Treatment of anxiety in older adolescents and young adults with autism spectrum disorders: A pilot study. Bull Menninger Clin [Internet]. 2020;84(2):105–36. Available from: http://ovidsp.ovid.com/ovidweb.cgi?T=JS&PAGE=reference&D=psyc17&NEWS=N&AN=2020-57770-001 | Wrong publication/study design |
| Kerns CM, Wood JJ, Kendall PC, Renno P, Crawford EA, Mercado RJ, et al. The Treatment of Anxiety in Autism Spectrum Disorder (TAASD) study: Rationale, design and methods. J Child Fam Stud [Internet]. 2016;25(6):1889–902. Available from: http://ovidsp.ovid.com/ovidweb.cgi?T=JS&PAGE=reference&D=psyc13&NEWS=N&AN=2016-06663-001 | Wrong publication/study design |
| Sarawgi S. Treating obsessive‐compulsive disorder in the presence of autism spectrum disorders. Brown University Child & Adolescent Behavior Letter [Internet]. 2021;37(2):1–4. Available from: https://search.ebscohost.com/login.aspx?direct=true&AuthType=ip,shib&db=jlh&AN=147991964&site=ehost-live&scope=site&custid=s8454451 | Wrong publication/study design |
| Whitehead JL. Treating AD-related anxiety as measured by the BASC in adolescents with Asperger’s disorder. Dissertation Abstracts International Section A: Humanities and Social Sciences [Internet]. 2005;66(6):2174. Available from: http://ovidsp.ovid.com/ovidweb.cgi?T=JS&PAGE=reference&D=psyc4&NEWS=N&AN=2005-99023-058 | Wrong publication/study design |
| Selles RR, Storch EA. Translation of anxiety treatment to youth with autism spectrum disorders. J Child Fam Stud [Internet]. 2013;22(3):405–13. Available from: http://ovidsp.ovid.com/ovidweb.cgi?T=JS&PAGE=reference&D=psyc10&NEWS=N&AN=2013-10414-010 | Wrong publication/study design |
| Maurice V, Russet F, Scocco P, McNicholas F, Santosh P, Singh SP, et al. Transition from child and adolescent mental health care to adult services for young people with Attention-Deficit/Hyperactivity Disorder (ADHD) or Autism Spectrum Disorder (ASD) in Europe: Barriers and recommendations. Encephale [Internet]. 2022; Available from: http://ovidsp.ovid.com/ovidweb.cgi?T=JS&PAGE=reference&D=medp&NEWS=N&AN=35725512 | Wrong publication/study design |
| Read M. Transforming Care: supporting people with learning disabilities, autism and mental health issues to move out of long-stay hospitals. Learning Disability Practice [Internet]. 2020;23(2):31–7. Available from: https://search.ebscohost.com/login.aspx?direct=true&AuthType=ip,shib&db=jlh&AN=148703706&site=ehost-live&scope=site&custid=s8454451 | Wrong publication/study design |
| Taylor JL. Transforming care for people with intellectual disabilities and autism in England. Lancet Psychiatry [Internet]. 2021;8(11):942–4. Available from: http://ovidsp.ovid.com/ovidweb.cgi?T=JS&PAGE=reference&D=psyc18&NEWS=N&AN=2022-00384-005 | Wrong publication/study design |
| Tchanturia K, Smith K, Glennon D, Burhouse A. Towards an Improved Understanding of the Anorexia Nervosa and Autism Spectrum Comorbidity: PEACE Pathway Implementation. Front Psychiatry [Internet]. 2020;11:640. Available from: http://ovidsp.ovid.com/ovidweb.cgi?T=JS&PAGE=reference&D=pmnm5&NEWS=N&AN=32733294 | Wrong publication/study design |
| Grob CS. 6.3 CURRENT RESEARCH ON PSILOCYBIN TREATMENT ON END-OF-LIFE ANXIETY AND MDMA TREATMENT OF SOCIAL ANXIETY IN AUTISM SPECTRUM DISORDER. J Am Acad Child Adolesc Psychiatry [Internet]. 2021;60(10):S9–S9. Available from: https://search.ebscohost.com/login.aspx?direct=true&AuthType=ip,shib&db=jlh&AN=152922493&site=ehost-live&scope=site&custid=s8454451 | Wrong publication/study design |
| Steinsiek J, Stone JW, Bedford J. 6.3 Current Treatment Practices for Depression in Children and Adolescents With Autism Spectrum Disorder and Intellectual and Developmental Disabilities. J Am Acad Child Adolesc Psychiatry [Internet]. 2021;60(10):S158–9. Available from: https://search.ebscohost.com/login.aspx?direct=true&AuthType=ip,shib&db=jlh&AN=152923326&site=ehost-live&scope=site&custid=s8454451 | Wrong publication/study design |
| Nowinski L. 15.3 CBT FOR ANXIETY IN AUTISM SPECTRUM DISORDER. J Am Acad Child Adolesc Psychiatry [Internet]. 2021;60(10):S281–S281. Available from: https://search.ebscohost.com/login.aspx?direct=true&AuthType=ip,shib&db=jlh&AN=152923076&site=ehost-live&scope=site&custid=s8454451 | Wrong publication/study design |
| Maddox BB. 37.3 Cognitive-Behavioral Treatment for Anxiety Disorders in Children and Adolescents With Autism Spectrum Disorder. J Am Acad Child Adolesc Psychiatry [Internet]. 2017;56:S55–S55. Available from: https://search.ebscohost.com/login.aspx?direct=true&AuthType=ip,shib&db=jlh&AN=125868472&site=ehost-live&scope=site&custid=s8454451 | Wrong publication/study design |
| Siegel M. 37.4 Pharmacologic Treatment of Anxiety Disorders and Obsessive-Compulsive Disorder in Children and Adolescents With Autism Spectrum Disorder. J Am Acad Child Adolesc Psychiatry [Internet]. 2017;56:S55–S55. Available from: https://search.ebscohost.com/login.aspx?direct=true&AuthType=ip,shib&db=jlh&AN=125867383&site=ehost-live&scope=site&custid=s8454451 | Wrong publication/study design |
| Sukhodolsky DG. 42.3 COGNITIVE AND BEHAVIORAL INTERVENTIONS FOR ANXIETY IN CHILDREN WITH AUTISM. J Am Acad Child Adolesc Psychiatry [Internet]. 2016;55:S325–S325. Available from: https://search.ebscohost.com/login.aspx?direct=true&AuthType=ip,shib&db=jlh&AN=118925827&site=ehost-live&scope=site&custid=s8454451 | Wrong publication/study design |
| Hardan A. 42.4 PSYCHOPHARMACOLOGICAL TREATMENT OF ANXIETY SYMPTOMS IN AUTISM SPECTRUM DISORDER. J Am Acad Child Adolesc Psychiatry [Internet]. 2016;55:S325–6. Available from: https://search.ebscohost.com/login.aspx?direct=true&AuthType=ip,shib&db=jlh&AN=118925603&site=ehost-live&scope=site&custid=s8454451 | Wrong publication/study design |
| Perkins M. 47.3 Fractured Treatment of Children and Adolescents With Mental Health Disorders and Co-Existing Autism Spectrum Disorder Developmental Disabilities and an Effort to Develop a Specialized Inpatient Psychiatric Unit. J Am Acad Child Adolesc Psychiatry [Internet]. 2017;56:S69–70. Available from: https://search.ebscohost.com/login.aspx?direct=true&AuthType=ip,shib&db=jlh&AN=125867742&site=ehost-live&scope=site&custid=s8454451 | Wrong publication/study design |
| £62 million investment aims to accelerate inpatient discharge. Learning Disability Practice [Internet]. 2020;23(4):6. Available from: https://search.ebscohost.com/login.aspx?direct=true&AuthType=ip,shib&db=jlh&AN=148704396&site=ehost-live&scope=site&custid=s8454451 | Wrong publication/study design |
| van Pelt BJ, Idris S, Jagersma G, Duvekot J, Maras A, van der Ende J, et al. The ACCEPT-study: Design of an RCT with an active treatment control condition to study the effectiveness of the Dutch version of PEERS for adolescents with autism spectrum disorder. BMC Psychiatry [Internet]. 2020;20. Available from: http://ovidsp.ovid.com/ovidweb.cgi?T=JS&PAGE=reference&D=psyc17&NEWS=N&AN=2020-39886-001 | Wrong publication/study design |
| Canon Garzon LF, a. Acceptance and Commitment Therapy (ACT) as an alternative for reducing psychological distress experienced by Latino parents of children with autism: A pilot study. Dissertation Abstracts International: Section B: The Sciences and Engineering [Internet]. 2013;74(6):No-Specified. Available from: http://ovidsp.ovid.com/ovidweb.cgi?T=JS&PAGE=reference&D=psyc10&NEWS=N&AN=2013-99241-092 | Wrong publication/study design |
| Boon ML. An adaptation of the mindful schools curriculum for adolescents with autism spectrum disorder: Implementation feasibility and impact on anxiety, rigidity, and mindfulness. Dissertation Abstracts International Section A: Humanities and Social Sciences [Internet]. 2018;79(7):No-Specified. Available from: http://ovidsp.ovid.com/ovidweb.cgi?T=JS&PAGE=reference&D=psyc15&NEWS=N&AN=2018-21178-044 | Wrong publication/study design |
| Kerns CM, Roux AM, Connell JE, Shattuck PT. Adapting cognitive behavioral techniques to address anxiety and depression in cognitively able emerging adults on the autism spectrum. Cogn Behav Pract [Internet]. 2016;23(3):329–40. Available from: http://ovidsp.ovid.com/ovidweb.cgi?T=JS&PAGE=reference&D=psyc13&NEWS=N&AN=2016-34950-002 | Wrong publication/study design |
| Gaus VL. Adult Asperger syndrome and the utility of cognitive-behavioral therapy. Journal of Contemporary Psychotherapy: On the Cutting Edge of Modern Developments in Psychotherapy [Internet]. 2011;41(1):47–56. Available from: http://ovidsp.ovid.com/ovidweb.cgi?T=JS&PAGE=reference&D=psyc8&NEWS=N&AN=2010-26914-006 | Wrong publication/study design |
| Jain N. Affective computing in the area of Autism. Dissertation Abstracts International: Section B: The Sciences and Engineering [Internet]. 2016;76(9):No-Specified. Available from: http://ovidsp.ovid.com/ovidweb.cgi?T=JS&PAGE=reference&D=psyc13&NEWS=N&AN=2016-26518-128 | Wrong publication/study design |
| Li Y, Sokhadze EM, Luo H, El-Baz AS, Elmaghraby AS. Affective virtual reality gaming for autism. Modern approaches to augmentation of brain function [Internet]. 2021;575–606. Available from: http://ovidsp.ovid.com/ovidweb.cgi?T=JS&PAGE=reference&D=psyc18&NEWS=N&AN=2022-10730-026 | Wrong publication/study design |
| Hardan A, Fung LK, King BH. Anxiety in Individuals With Autism Spectrum Disorder: Clinical Assessment, Biology, and Treatments. J Am Acad Child Adolesc Psychiatry [Internet]. 2016;55:S324–S324. Available from: https://search.ebscohost.com/login.aspx?direct=true&AuthType=ip,shib&db=jlh&AN=118925689&site=ehost-live&scope=site&custid=s8454451 | Wrong publication/study design |
| Pae CU, Forbes A, Patkar AA. Aripiprazole as adjunctive therapy for patients with major depressive disorder: Overview and implications of clinical trial data. CNS Drugs [Internet]. 2011;25(2):109–27. Available from: http://ovidsp.ovid.com/ovidweb.cgi?T=JS&PAGE=reference&D=psyc8&NEWS=N&AN=2011-03798-002 | Wrong publication/study design |
| Moskowitz LJ. Assessment and treatment of anxiety and problem behavior in children with autism spectrum disorders and intellectual disability. Dissertation Abstracts International: Section B: The Sciences and Engineering [Internet]. 2016;76(8):No-Specified. Available from: http://ovidsp.ovid.com/ovidweb.cgi?T=JS&PAGE=reference&D=psyc13&NEWS=N&AN=2016-17339-179 | Wrong publication/study design |
| Lai AG, Chang WH, Skuse D. Autism and mental illness in children and young people require standardised approaches for assessment and treatment. The Lancet regional health Europe [Internet]. 2022;16:100360. Available from: http://ovidsp.ovid.com/ovidweb.cgi?T=JS&PAGE=reference&D=pmnm&NEWS=N&AN=35368370 | Wrong publication/study design |
| Hodges C, Leach JO. Balancing our focus: The development of a collaborative approach between child development and children’s mental health services. International Journal of Integrated Care (IJIC) [Internet]. 2018;18:1–2. Available from: https://search.ebscohost.com/login.aspx?direct=true&AuthType=ip,shib&db=jlh&AN=138115024&site=ehost-live&scope=site&custid=s8454451 | Wrong publication/study design |
| Henton P, Deitrick L, Godfroy R, Horodeczny C, Madaris T, Mericle S, et al. Basic Activities and Anxiety Reduction in the Care of Canines (BAARCC) Protocol for Adults With Autism Spectrum Disorder. SIS Quarterly Practice Connections [Internet]. 2020;5(2):2–4. Available from: https://search.ebscohost.com/login.aspx?direct=true&AuthType=ip,shib&db=jlh&AN=143049465&site=ehost-live&scope=site&custid=s8454451 | Wrong publication/study design |
| Phillips Z. Behavioral change in a summer treatment program for children with ASD and ADHD. Dissertation Abstracts International: Section B: The Sciences and Engineering [Internet]. 2022;83(5):No-Specified. Available from: http://ovidsp.ovid.com/ovidweb.cgi?T=JS&PAGE=reference&D=psyc18&NEWS=N&AN=2022-34074-191 | Wrong publication/study design |
| Elliott SJ, Marshall D, Morley K, Uphoff E, Kumar M, Meader N. Behavioural and cognitive behavioural therapy for obsessive compulsive disorder (OCD) in individuals with autism spectrum disorder (ASD). Cochrane Database Syst Rev [Internet]. 2021;9:CD013173. Available from: http://ovidsp.ovid.com/ovidweb.cgi?T=JS&PAGE=reference&D=med20&NEWS=N&AN=34693989 | Wrong publication/study design |
| Dingfelder HE, M, ell DS. Bridging the research-to-practice gap in autism intervention: An application of diffusion of innovation theory. J Autism Dev Disord [Internet]. 2011;41(5):597–609. Available from: http://ovidsp.ovid.com/ovidweb.cgi?T=JS&PAGE=reference&D=psyc8&NEWS=N&AN=2011-07919-007 | Wrong publication/study design |
| Rowe G, Nevin H. Bringing “patient voice” into psychological formulations of in-patients with intellectual disabilities, autism spectrum disorder and severe challenging behaviours: Report of a service improvement pilot. Br J Learn Disabil [Internet]. 2014;42(3):177–84. Available from: http://ovidsp.ovid.com/ovidweb.cgi?T=JS&PAGE=reference&D=psyc11&NEWS=N&AN=2014-32986-002 | Wrong publication/study design |
| Bjastad JF. Can children and adolescents with Asperger’s syndrome/high-functioning autism benefit from cognitive behavioural therapy? Kan bam og ungdom med Aspergersyndrom og hoytfungerende autisme ha nytte av kognitiv atferdsterapi? [Internet]. 2011;48(1):69–74. Available from: http://ovidsp.ovid.com/ovidweb.cgi?T=JS&PAGE=reference&D=psyc8&NEWS=N&AN=2011-01719-012 | Wrong publication/study design |
| Scarpinato N, Bradley J, Kurbjun K, Bateman X, Holtzer B, Ely B. Caring for the child with an autism spectrum disorder in the acute care setting. Journal for Specialists in Pediatric Nursing [Internet]. 2010;15(3):244–54. Available from: http://ovidsp.ovid.com/ovidweb.cgi?T=JS&PAGE=reference&D=psyc7&NEWS=N&AN=2010-13433-009 | Wrong publication/study design |
| Burton C, Allan E, Eckhardt S, Le Grange D, Ehrenreich-May J, Singh M, et al. Case presentations combining family-based treatment with the unified protocols for transdiagnostic treatment of emotional disorders in children and adolescents for comorbid Avoidant Restrictive Food Intake Disorder and Autism Spectrum Disorder. J Can Acad Child Adolesc Psychiatry [Internet]. 2021;30(4):280–91. Available from: http://ovidsp.ovid.com/ovidweb.cgi?T=JS&PAGE=reference&D=psyc18&NEWS=N&AN=2022-04141-007 | Wrong publication/study design |
| Chandrasekhar T, Sikich L. Challenges in the diagnosis and treatment of depression in autism spectrum disorders across the lifespan. Dialogues Clin Neurosci [Internet]. 2015;17(2):219–27. Available from: http://ovidsp.ovid.com/ovidweb.cgi?T=JS&PAGE=reference&D=med12&NEWS=N&AN=26246795 | Wrong publication/study design |
| Beauvois L, Kverno K. Challenges in treating children with autism spectrum disorder: Implications for psychiatric-mental health nurse practitioners. J Psychosoc Nurs Ment Health Serv [Internet]. 2020;58(12):7–12. Available from: http://ovidsp.ovid.com/ovidweb.cgi?T=JS&PAGE=reference&D=psyc18&NEWS=N&AN=2021-00740-001 | Wrong publication/study design |
| Kuroda M, Kawakubo Y, Kuwabara H, Kano Y, Kamio Y. [Characteristics and adaptive problems of adults with subthreshold ASD in cognitive-behavioral intervention research for emotion regulation]. Seishin Shinkeigaku Zasshi [Internet]. 2013;115(6):623–9. Available from: http://ovidsp.ovid.com/ovidweb.cgi?T=JS&PAGE=reference&D=med10&NEWS=N&AN=23944120 | Wrong publication/study design |
| Attwood T. Cognitive Behaviour Therapy for Children and Adults with Asperger’s Syndrome. Behaviour Change [Internet]. 2004;21(3):147–61. Available from: http://ovidsp.ovid.com/ovidweb.cgi?T=JS&PAGE=reference&D=psyc4&NEWS=N&AN=2005-00662-001 | Wrong publication/study design |
| Gaus VL. Cognitive behavioural therapy for adults with autism spectrum disorder. Adv Ment Health Intellect Disabil [Internet]. 2011;5(5):15–25. Available from: http://ovidsp.ovid.com/ovidweb.cgi?T=JS&PAGE=reference&D=psyc8&NEWS=N&AN=2011-24380-003 | Wrong publication/study design |
| Nadeau JM, Arnold EB, Selles RR, Storch EA, Lewin AB. A cognitive-behavioral approach for anxiety in a preschool-aged child with autism spectrum disorder. Clin Case Stud [Internet]. 2015;14(1):47–60. Available from: http://ovidsp.ovid.com/ovidweb.cgi?T=JS&PAGE=reference&D=psyc12&NEWS=N&AN=2014-57674-004 | Wrong publication/study design |
| Oshima F, William M, y, Takahashi N, Tsuchiyagaito A, Kuwabara H, et al. Cognitive-behavioral family therapy as psychoeducation for adolescents with high-functioning autism spectrum disorders: Aware and Care for my Autistic Traits (ACAT) program study protocol for a pragmatic multisite randomized controlled trial. Trials [Internet]. 2020;21(1):814. Available from: http://ovidsp.ovid.com/ovidweb.cgi?T=JS&PAGE=reference&D=med18&NEWS=N&AN=32993775 | Wrong publication/study design |
| Kuroda M, Kawakubo Y, Kuwabara H, Yokoyama K, Kano Y, Kamio Y. A cognitive-behavioral intervention for emotion regulation in adults with high-functioning autism spectrum disorders: study protocol for a randomized controlled trial. Trials [Internet]. 2013;14:231. Available from: http://ovidsp.ovid.com/ovidweb.cgi?T=JS&PAGE=reference&D=med10&NEWS=N&AN=23880333 | Wrong publication/study design |
| Kirkl, Michelle M. Cognitive-behavioral therapy for anxiety in youth with autism: Paving the way to evidence-based practice. Dissertation Abstracts International: Section B: The Sciences and Engineering [Internet]. 2022;83(5):No-Specified. Available from: http://ovidsp.ovid.com/ovidweb.cgi?T=JS&PAGE=reference&D=psyc18&NEWS=N&AN=2022-34073-269 | Wrong publication/study design |
| Patterson T, Higgins M, Dyck DG. A collaborative approach to reduce hospitalization of developmentally disabled clients with mental illness. Psychiatric Services [Internet]. 1995;46(3):243–7. Available from: http://ovidsp.ovid.com/ovidweb.cgi?T=JS&PAGE=reference&D=psyc3&NEWS=N&AN=1995-34435-001 | Wrong publication/study design |
| van Schalkwyk GI, Beyer C, Martin A, Volkmar FR. College students with autism spectrum disorders: A growing role for adult psychiatrists. Journal of American College Health [Internet]. 2016;64(7):575–9. Available from: http://ovidsp.ovid.com/ovidweb.cgi?T=JS&PAGE=reference&D=psyc13&NEWS=N&AN=2016-41668-008 | Wrong publication/study design |
| Norris AL. Community capacity to provide mental/behavioral health services to people with developmental disabilities. Dissertation Abstracts International: Section B: The Sciences and Engineering [Internet]. 2015;76(6):No-Specified. Available from: http://ovidsp.ovid.com/ovidweb.cgi?T=JS&PAGE=reference&D=psyc12&NEWS=N&AN=2015-99240-405 | Wrong publication/study design |
| Torrey WC. Community care of adults with developmental disabilities and mental illness. Practicing psychiatry in the community: A manual [Internet]. 1996;153–72. Available from: http://ovidsp.ovid.com/ovidweb.cgi?T=JS&PAGE=reference&D=psyc3&NEWS=N&AN=1996-97758-009 | Wrong publication/study design |
| Maddox BB, Gaus VL. Community Mental Health Services for Autistic Adults: Good News and Bad News. Autism Adulthood [Internet]. 2019;1(1):15–9. Available from: http://ovidsp.ovid.com/ovidweb.cgi?T=JS&PAGE=reference&D=pmnm4&NEWS=N&AN=31667466 | Wrong publication/study design |
| Chu HC. A comparison of verbal-behavior and social-skills approaches for development of social interaction skills and concurrent reduction of aberrant behaviors of children with developmental disabilities in the context of matching theory. Dissertation Abstracts International Section A: Humanities and Social Sciences [Internet]. 1998;59(6):1974. Available from: http://ovidsp.ovid.com/ovidweb.cgi?T=JS&PAGE=reference&D=psyc3&NEWS=N&AN=1998-95023-185 | Wrong publication/study design |
| Wynn T, Brunetti S. Compounding naltrexone for the treatment of autism. Int J Pharm Compd [Internet]. 2009;13(4):296–9. Available from: http://ovidsp.ovid.com/ovidweb.cgi?T=JS&PAGE=reference&D=pmnm2&NEWS=N&AN=23966519 | Wrong publication/study design |
| Holttum JR, Lubetsky MJ, Eastman LE. Comprehensive management of trichotillomania in a young autistic girl. J Am Acad Child Adolesc Psychiatry [Internet]. 1994;33(4):577–81. Available from: http://ovidsp.ovid.com/ovidweb.cgi?T=JS&PAGE=reference&D=psyc3&NEWS=N&AN=1994-42056-001 | Wrong publication/study design |
| A comprehensive program for treating profoundly autistic children. Psychiatric Services [Internet]. 2000;51(11):1439–40. Available from: http://ovidsp.ovid.com/ovidweb.cgi?T=JS&PAGE=reference&D=psyc3&NEWS=N&AN=2000-12273-017 | Wrong publication/study design |
| Shochet IM, Saggers BR, Carrington SB, Orr JA, Wurfl AM, Duncan BM, et al. The Cooperative Research Centre for Living with Autism (Autism CRC) conceptual model to promote mental health for adolescents with ASD. Clin Child Fam Psychol Rev [Internet]. 2016;19(2):94–116. Available from: http://ovidsp.ovid.com/ovidweb.cgi?T=JS&PAGE=reference&D=psyc13&NEWS=N&AN=2016-21652-001 | Wrong publication/study design |
| Van Adel JM, Geier JD, Perry A, Reitzel JAM. Credible knowledge: a pilot evaluation of a modified GRADE method using parent-implemented interventions for children with autism. BMC Health Serv Res [Internet]. 2011;11:60. Available from: http://ovidsp.ovid.com/ovidweb.cgi?T=JS&PAGE=reference&D=med8&NEWS=N&AN=21426564 | Wrong publication/study design |
| Jacques C, Cloutier V, Bouchard S. The “Decoding of Social Interactions in Virtual Reality” tasks for autism spectrum people: Development of an intervention protocol and pilot testing. Annual Review of CyberTherapy and Telemedicine [Internet]. 2018;16:148–52. Available from: http://ovidsp.ovid.com/ovidweb.cgi?T=JS&PAGE=reference&D=psyc15&NEWS=N&AN=2019-27383-021 | Wrong publication/study design |
| Murphy D, Glaser K, Hayward H, Eklund H, Cadman T, Findon J, et al. Crossing the divide: a longitudinal study of effective treatments for people with autism and attention deficit hyperactivity disorder across the lifespan. 2018; Available from: http://ovidsp.ovid.com/ovidweb.cgi?T=JS&PAGE=reference&D=medp&NEWS=N&AN=30088881 | Wrong publication/study design |
| Ahc M. Digital Applications Used in Mental Health Treatment: Two Randomized Studies. Integrative Medicine Alert [Internet]. 2019;22(8):N.PAG-N.PAG. Available from: https://search.ebscohost.com/login.aspx?direct=true&AuthType=ip,shib&db=jlh&AN=138277020&site=ehost-live&scope=site&custid=s8454451 | Wrong publication/study design |
| White SW, Albano AM, Johnson CR, Kasari C, Ollendick T, Klin A, et al. Development of a cognitive-behavioral intervention program to treat anxiety and social deficits in teens with high-functioning autism. Clin Child Fam Psychol Rev [Internet]. 2010;13(1):77–90. Available from: http://ovidsp.ovid.com/ovidweb.cgi?T=JS&PAGE=reference&D=psyc7&NEWS=N&AN=2010-03378-003 | Wrong publication/study design |
| Banerjee D, Vajawat B, Varshney P. Digital gaming interventions: A novel paradigm in mental health? Perspectives from India. International Review of Psychiatry [Internet]. 2021;33(4):435–41. Available from: http://ovidsp.ovid.com/ovidweb.cgi?T=JS&PAGE=reference&D=psyc17&NEWS=N&AN=2020-88361-001 | Wrong publication/study design |
| Burbach FR, Stiles KM. Digital Mental Health and Neurodevelopmental Services: Case-Based Realist Evaluation. JMIR Form Res [Internet]. 2021;5(9):e29845. Available from: http://ovidsp.ovid.com/ovidweb.cgi?T=JS&PAGE=reference&D=pmnm5&NEWS=N&AN=34369382 | Wrong publication/study design |
| Shawler P. Does early intervention reduce the risk of future emotional and behavioral problems in children with autism spectrum disorder. Dissertation Abstracts International: Section B: The Sciences and Engineering [Internet]. 2017;78(4):No-Specified. Available from: http://ovidsp.ovid.com/ovidweb.cgi?T=JS&PAGE=reference&D=psyc14&NEWS=N&AN=2017-05719-178 | Wrong publication/study design |
| Keefer A, Vasa RA. DINOSAUR: An integrated cognitive-behavioral treatment for anxiety in young children with ASD. J Neurodev Disord [Internet]. 2021;13. Available from: http://ovidsp.ovid.com/ovidweb.cgi?T=JS&PAGE=reference&D=psyc18&NEWS=N&AN=2021-94054-001 | Wrong publication/study design |
| Drumming for Autism. Natural Solutions [Internet]. 2013;(151):12. Available from: https://search.ebscohost.com/login.aspx?direct=true&AuthType=ip,shib&db=jlh&AN=100038540&site=ehost-live&scope=site&custid=s8454451 | Wrong publication/study design |
| Barnett DW, Bell SH, Bauer A, Lentz Francis E. Jr, Petrelli S, Air A, et al. The Early Childhood Intervention Project: Building capacity for service delivery. Special Issue: Changing Teacher and Staff Behavior to Benefit Children [Internet]. 1997;12(4):293–315. Available from: http://ovidsp.ovid.com/ovidweb.cgi?T=JS&PAGE=reference&D=psyc3&NEWS=N&AN=1997-38839-002 | Wrong publication/study design |
| Pejic I. Early intervention for children with autistic spectrum disorders through a system of mobile services in the local community. Autism 360degree [Internet]. 2020;269–76. Available from: http://ovidsp.ovid.com/ovidweb.cgi?T=JS&PAGE=reference&D=psyc17&NEWS=N&AN=2019-72324-015 | Wrong publication/study design |
| Huntjens A, van den Bosch LMCW, Sizoo B, Kerkhof A, Huibers MJH, van der Gaag M. The effect of dialectical behaviour therapy in autism spectrum patients with suicidality and/ or self-destructive behaviour (DIASS): Study protocol for a multicentre randomised controlled trial. BMC Psychiatry [Internet]. 2020;20. Available from: http://ovidsp.ovid.com/ovidweb.cgi?T=JS&PAGE=reference&D=psyc17&NEWS=N&AN=2020-20540-001 | Wrong publication/study design |
| Wijnhoven LAMW, Creemers DHM, Engels RCME, Granic I. The effect of the video game Mindlight on anxiety symptoms in children with an Autism Spectrum Disorder. BMC Psychiatry [Internet]. 2015;15. Available from: http://ovidsp.ovid.com/ovidweb.cgi?T=JS&PAGE=reference&D=psyc12&NEWS=N&AN=2015-29816-001 | Wrong publication/study design |
| Ashworth S, Bamford J, Tully R. The effectiveness of a cbt-based intervention for depression symptoms with a female forensic inpatient with cognitive disability and autism. Journal of Forensic Psychiatry & Psychology [Internet]. 2020;No-Specified. Available from: http://ovidsp.ovid.com/ovidweb.cgi?T=JS&PAGE=reference&D=psyc17&NEWS=N&AN=2020-28696-001 | Wrong publication/study design |
| Myszak JP. Effectiveness of a computer program in increasing social skills in children with autism spectrum disorder. Dissertation Abstracts International Section A: Humanities and Social Sciences [Internet]. 2011;71(11):3899. Available from: http://ovidsp.ovid.com/ovidweb.cgi?T=JS&PAGE=reference&D=psyc8&NEWS=N&AN=2011-99090-294 | Wrong publication/study design |
| Kalek D. The effectiveness of a family-centered early intervention program for parents of children with developmental delays ages 0 through 3. Dissertation Abstracts International Section A: Humanities and Social Sciences [Internet]. 2009;69(7):2594. Available from: http://ovidsp.ovid.com/ovidweb.cgi?T=JS&PAGE=reference&D=psyc6&NEWS=N&AN=2009-99010-167 | Wrong publication/study design |
| McNally Keehn R. Effectiveness of cognitive-behavioral therapy for children with high-functioning autism and anxiety. Dissertation Abstracts International: Section B: The Sciences and Engineering [Internet]. 2011;71(11):7097. Available from: http://ovidsp.ovid.com/ovidweb.cgi?T=JS&PAGE=reference&D=psyc8&NEWS=N&AN=2011-99100-241 | Wrong publication/study design |
| Cullain RE. The effects of social stories on anxiety levels and excessive behavioral expressions of elementary school-aged children with autism. Dissertation Abstracts International Section A: Humanities and Social Sciences [Internet]. 2002;62(7):2383. Available from: http://ovidsp.ovid.com/ovidweb.cgi?T=JS&PAGE=reference&D=psyc4&NEWS=N&AN=2002-95001-073 | Wrong publication/study design |
| McDaid PK. The effects of stress inoculation training on the anxiety-related challenging behavior of middle school students with Asperger’s syndrome. Dissertation Abstracts International Section A: Humanities and Social Sciences [Internet]. 2007;68(4):1409. Available from: http://ovidsp.ovid.com/ovidweb.cgi?T=JS&PAGE=reference&D=psyc5&NEWS=N&AN=2007-99190-282 | Wrong publication/study design |
| Cunningham A, ra. Effects of the relationship enhancementrtm program on social skills, empathy and social support for adults with autism spectrum disorders. Dissertation Abstracts International: Section B: The Sciences and Engineering [Internet]. 2015;75(9):No-Specified. Available from: http://ovidsp.ovid.com/ovidweb.cgi?T=JS&PAGE=reference&D=psyc12&NEWS=N&AN=2015-99060-195 | Wrong publication/study design |
| Pryor FC. Efficacy of a computer-assisted cognitive-behavioral therapy program in treating youth with anxiety and co-occurring autism spectrum disorder: Camp Cope A-Lot. Dissertation Abstracts International: Section B: The Sciences and Engineering [Internet]. 2018;78(7):No-Specified. Available from: http://ovidsp.ovid.com/ovidweb.cgi?T=JS&PAGE=reference&D=psyc15&NEWS=N&AN=2017-19722-029 | Wrong publication/study design |
| Carlier S, der Paelt S, Ongenae F, De Backere F, De Turck F. Empowering Children with ASD and Their Parents: Design of a Serious Game for Anxiety and Stress Reduction. Sensors (Basel) [Internet]. 2020;20(4). Available from: http://ovidsp.ovid.com/ovidweb.cgi?T=JS&PAGE=reference&D=med17&NEWS=N&AN=32054025 | Wrong publication/study design |
| Sze KM, Wood JJ. Enhancing CBT for the treatment of autism spectrum disorders and concurrent anxiety. Behavioural and Cognitive Psychotherapy [Internet]. 2008;36(4):403–9. Available from: http://ovidsp.ovid.com/ovidweb.cgi?T=JS&PAGE=reference&D=psyc6&NEWS=N&AN=2008-15717-003 | Wrong publication/study design |
| Patriquin MA. Evidence-based treatment and conceptualization of autism spectrum disorder: Emotion regulation, social impairment, and anxiety as targets. Special Issue: Evidence-Based Treatment and Conceptualization of Autism Spectrum Disorder [Internet]. 2019;83(3):199–204. Available from: http://ovidsp.ovid.com/ovidweb.cgi?T=JS&PAGE=reference&D=psyc16&NEWS=N&AN=2019-57224-001 | Wrong publication/study design |
| Pickard K, Reyes N, Reaven J. Examining the inclusion of diverse participants in cognitive behavior therapy research for youth with autism spectrum disorder and anxiety. Autism [Internet]. 2019;23(4):1057–64. Available from: http://ovidsp.ovid.com/ovidweb.cgi?T=JS&PAGE=reference&D=psyc16&NEWS=N&AN=2019-29502-024 | Wrong publication/study design |
| Maroney MR. Exploring the efficacy of an online intervention in processing experiences of heterosexism among autistic-LGBTQ individuals. Dissertation Abstracts International: Section B: The Sciences and Engineering [Internet]. 2021;82(3):No-Specified. Available from: http://ovidsp.ovid.com/ovidweb.cgi?T=JS&PAGE=reference&D=psyc18&NEWS=N&AN=2020-79974-122 | Wrong publication/study design |
| Bruce S. Exploring the impact of the PEERS intervention on mental health and emotion regulation in adolescents with autism spectrum disorder. Dissertation Abstracts International: Section B: The Sciences and Engineering [Internet]. 2022;83(5):No-Specified. Available from: http://ovidsp.ovid.com/ovidweb.cgi?T=JS&PAGE=reference&D=psyc18&NEWS=N&AN=2022-34073-150 | Wrong publication/study design |
| Attwood T. Expressing and enjoying love and affection: A cognitive-behavioral program for children and adolescents with high-functioning ASD. CBT for children and adolescents with high-functioning autism spectrum disorders [Internet]. 2013;259–77. Available from: http://ovidsp.ovid.com/ovidweb.cgi?T=JS&PAGE=reference&D=psyc10&NEWS=N&AN=2013-32785-011 | Wrong publication/study design |
| Beck KB. Feasibility of mindfulness-based stress reduction for adults With Autism Spectrum Disorder. Dissertation Abstracts International: Section B: The Sciences and Engineering [Internet]. 2018;79(4):No-Specified. Available from: http://ovidsp.ovid.com/ovidweb.cgi?T=JS&PAGE=reference&D=psyc15&NEWS=N&AN=2018-11224-176 | Wrong publication/study design |
| Wilson CS. Feasibility of psychosis risk assessment for individuals diagnosed with an autism spectrum disorder: A cognitive exploration of Item Interpretation of the Structured Interview for psychosis-risk syndromes. Dissertation Abstracts International: Section B: The Sciences and Engineering [Internet]. 2018;79(3):No-Specified. Available from: http://ovidsp.ovid.com/ovidweb.cgi?T=JS&PAGE=reference&D=psyc15&NEWS=N&AN=2018-09132-116 | Wrong publication/study design |
| For Many Kids With Autism, Health Care Needs Go Unaddressed. Palaestra [Internet]. 2019;33(3):61. Available from: https://search.ebscohost.com/login.aspx?direct=true&AuthType=ip,shib&db=jlh&AN=139041523&site=ehost-live&scope=site&custid=s8454451 | Wrong publication/study design |
| Coffey A, a L., Shawler LA, Jessel J, Bain T, Nye M, et al. Generality of the practical functional assessment and skill-based treatment among individuals with autism and mental health disorders. Behavioral Interventions [Internet]. 2021;36(1):298–314. Available from: http://ovidsp.ovid.com/ovidweb.cgi?T=JS&PAGE=reference&D=psyc18&NEWS=N&AN=2020-85626-001 | Wrong publication/study design |
| Weiss JA, Lunsky Y. Group cognitive behaviour therapy for adults with Asperger syndrome and anxiety or mood disorder: A case series. Clin Psychol Psychother [Internet]. 2010;17(5):438–46. Available from: http://ovidsp.ovid.com/ovidweb.cgi?T=JS&PAGE=reference&D=psyc7&NEWS=N&AN=2010-19860-009 | Wrong publication/study design |
| Glynn K, Lyne J. Hidden addictions, improving eating disorder services and overcoming challenges in autism spectrum disorders. Ir J Psychol Med [Internet]. 2018;35(4):267. Available from: https://search.ebscohost.com/login.aspx?direct=true&AuthType=ip,shib&db=jlh&AN=133320955&site=ehost-live&scope=site&custid=s8454451 | Wrong publication/study design |
| Russell A, Gaunt D, Cooper K, Horwood J, Barton S, Ensum I, et al. Guided self-help for depression in autistic adults: the ADEPT feasibility RCT. Health Technol Assess [Internet]. 2019;23(68):1–94. Available from: http://ovidsp.ovid.com/ovidweb.cgi?T=JS&PAGE=reference&D=med16&NEWS=N&AN=31856942 | Wrong publication/study design |
| Roy A. How can psychiatrists make mental health services more accessible for people with autism? BJPsych Open [Internet]. 2021;7(6):e197. Available from: http://ovidsp.ovid.com/ovidweb.cgi?T=JS&PAGE=reference&D=pmnm5&NEWS=N&AN=34709152 | Wrong publication/study design |
| Schultz HM. Identification of mental health needs among adolescents with autism spectrum disorder: An investigation of a multi-informant approach. Dissertation Abstracts International: Section B: The Sciences and Engineering [Internet]. 2022;83(3):No-Specified. Available from: http://ovidsp.ovid.com/ovidweb.cgi?T=JS&PAGE=reference&D=psyc18&NEWS=N&AN=2021-94598-032 | Wrong publication/study design |
| Wendler D. Improv theater as a social cognition intervention for autism. Dissertation Abstracts International: Section B: The Sciences and Engineering [Internet]. 2019;80(8):No-Specified. Available from: http://ovidsp.ovid.com/ovidweb.cgi?T=JS&PAGE=reference&D=psyc16&NEWS=N&AN=2019-41141-135 | Wrong publication/study design |
| Selvap, iyan J. Improvising Pragmatic Cognitive-Behavioral Therapy for Depressed Adolescents With Asperger Syndrome. Prim Care Companion CNS Disord [Internet]. 2019;21(3). Available from: http://ovidsp.ovid.com/ovidweb.cgi?T=JS&PAGE=reference&D=med16&NEWS=N&AN=31265767 | Wrong publication/study design |
| Drahota AM. Intervening with independent daily living skills for high-functioning children with autism and concurrent anxiety disorders. Dissertation Abstracts International Section A: Humanities and Social Sciences [Internet]. 2009;69(7):2601. Available from: http://ovidsp.ovid.com/ovidweb.cgi?T=JS&PAGE=reference&D=psyc6&NEWS=N&AN=2009-99010-440 | Wrong publication/study design |
| Moskowitz LJ, Walsh CE, Mulder E, McLaughlin DM, Hajcak G, Carr EG, et al. Intervention for anxiety and problem behavior in children with autism spectrum disorder and intellectual disability. J Autism Dev Disord [Internet]. 2017;47(12):3930–48. Available from: http://ovidsp.ovid.com/ovidweb.cgi?T=JS&PAGE=reference&D=psyc14&NEWS=N&AN=2017-11487-001 | Wrong publication/study design |
| Danforth AL, Struble CM, Yazar-Klosinski B, Grob CS. MDMA-assisted therapy: A new treatment model for social anxiety in autistic adults. Prog Neuropsychopharmacol Biol Psychiatry [Internet]. 2016;64:237–49. Available from: http://ovidsp.ovid.com/ovidweb.cgi?T=JS&PAGE=reference&D=psyc13&NEWS=N&AN=2015-45278-023 | Wrong publication/study design |
| Di Marco M, Iacono T. Mental Health Assessment and Intervention for People With Complex Communication Needs Associated With Developmental Disabilities. J Policy Pract Intellect Disabil [Internet]. 2007;4(1):40–59. Available from: http://ovidsp.ovid.com/ovidweb.cgi?T=JS&PAGE=reference&D=psyc5&NEWS=N&AN=2007-03196-005 | Wrong publication/study design |
| Bal VH, Wilkinson E, Glascock V, Hastings RP, Jahoda A. Mechanisms of change in behavioral activation: Adapting depression treatment for autistic people. Cogn Behav Pract [Internet]. 2022;No-Specified. Available from: http://ovidsp.ovid.com/ovidweb.cgi?T=JS&PAGE=reference&D=psyc18&NEWS=N&AN=2022-57042-001 | Wrong publication/study design |
| Ward A, Russell A. Mental health services for adults with autism spectrum disorders. Advances in Mental Health and Learning Disabilities [Internet]. 2007;1(4):23–6. Available from: http://ovidsp.ovid.com/ovidweb.cgi?T=JS&PAGE=reference&D=psyc5&NEWS=N&AN=2010-24328-005 | Wrong publication/study design |
| Lake JK, Perry A, Lunsky Y. Mental health services for individuals with high functioning autism spectrum disorder. Autism Res Treat [Internet]. 2014;2014:502420. Available from: http://ovidsp.ovid.com/ovidweb.cgi?T=JS&PAGE=reference&D=pmnm3&NEWS=N&AN=25276425 | Wrong publication/study design |
| Giaquinto MB. Mindfulness, social problem solving, social anxiety and quality of life in college students with autism spectrum disorders. Dissertation Abstracts International Section A: Humanities and Social Sciences [Internet]. 2015;75(10):No-Specified. Available from: http://ovidsp.ovid.com/ovidweb.cgi?T=JS&PAGE=reference&D=psyc12&NEWS=N&AN=2015-99071-189 | Wrong publication/study design |
| Mirtazapine for anxiety in autism. Brown University Psychopharmacology Update [Internet]. 2022;33(7):7. Available from: https://search.ebscohost.com/login.aspx?direct=true&AuthType=ip,shib&db=jlh&AN=157235985&site=ehost-live&scope=site&custid=s8454451 | Wrong publication/study design |
| Stadnick NA, Brookman-Frazee L, M, ell DS, Kuelbs CL, Coleman KJ, et al. A mixed methods study to adapt and implement integrated mental healthcare for children with autism spectrum disorder. Pilot Feasibility Stud [Internet]. 2019;5:51. Available from: http://ovidsp.ovid.com/ovidweb.cgi?T=JS&PAGE=reference&D=pmnm&NEWS=N&AN=30976456 | Wrong publication/study design |
| Harrings R, ie K. Modified cognitive behavioral therapy (CBT) program for high functioning autism. Dissertation Abstracts International: Section B: The Sciences and Engineering [Internet]. 2018;79(10):No-Specified. Available from: http://ovidsp.ovid.com/ovidweb.cgi?T=JS&PAGE=reference&D=psyc15&NEWS=N&AN=2018-34221-134 | Wrong publication/study design |
| Williams DT, Hirsch S, Coffey B. Mood and anxiety symptoms in an adolescent with pervasive developmental disorder not otherwise specified and moderate mental retardation. J Child Adolesc Psychopharmacol [Internet]. 2007;17(5):721–6. Available from: http://ovidsp.ovid.com/ovidweb.cgi?T=JS&PAGE=reference&D=psyc5&NEWS=N&AN=2007-17730-017 | Wrong publication/study design |
| Moskowitz LJ, Mulder E, Walsh CE, McLaughlin DM, Zarcone JR, Proudfit GH, et al. A multimethod assessment of anxiety and problem behavior in children with autism spectrum disorders and intellectual disability. Am J Intellect Dev Disabil [Internet]. 2013;118(6):419–34. Available from: http://ovidsp.ovid.com/ovidweb.cgi?T=JS&PAGE=reference&D=psyc10&NEWS=N&AN=2014-03585-003 | Wrong publication/study design |
| Cashin A. Narrative therapy: A psychotherapeutic approach in the treatment of adolescents with Asperger’s disorder. Journal of Child and Adolescent Psychiatric Nursing [Internet]. 2008;21(1):48–56. Available from: http://ovidsp.ovid.com/ovidweb.cgi?T=JS&PAGE=reference&D=psyc6&NEWS=N&AN=2008-01715-006 | Wrong publication/study design |
| Fida A, Naveed S, Waqas A. Non-Specialist Mediated Interventions for Autism Spectrum Disorder: An Overview. J Psychosoc Nurs Ment Health Serv [Internet]. 2019;57(5):15–9. Available from: http://ovidsp.ovid.com/ovidweb.cgi?T=JS&PAGE=reference&D=med16&NEWS=N&AN=31042296 | Wrong publication/study design |
| Bloomfield BS, Fischer AJ, Dove M, Clark RR, Fife M. Parent teleconsultation to increase bites consumed: A demonstration across foods for a child with ARFID and ASD. Behav Anal Pract [Internet]. 2021;14(4):913–26. Available from: http://ovidsp.ovid.com/ovidweb.cgi?T=JS&PAGE=reference&D=psyc18&NEWS=N&AN=2021-65903-001 | Wrong publication/study design |
| Cook JM, Donovan CL, Garnett MS. A parent-mediated, cognitive behavioral therapy group treatment for young children with high-functioning autism spectrum disorder and comorbid anxiety: Development and case illustration of the Fun with Feelings program. J Cogn Psychother [Internet]. 2017;31(3):204–24. Available from: http://ovidsp.ovid.com/ovidweb.cgi?T=JS&PAGE=reference&D=psyc14&NEWS=N&AN=2017-54975-005 | Wrong publication/study design |
| Ridderinkhof A, Elmose M, de Bruin EI, Blom R, Salem-Guirgis S, ra, et al. A person-centered approach in investigating a mindfulness-based program for adolescents with autism spectrum disorder. Mindfulness (N Y) [Internet]. 2021;12(10):2394–414. Available from: http://ovidsp.ovid.com/ovidweb.cgi?T=JS&PAGE=reference&D=psyc18&NEWS=N&AN=2021-79648-001 | Wrong publication/study design |
| White SW, Ollendick T, Scahill L, Oswald D, Albano AM. Preliminary efficacy of a cognitive-behavioral treatment program for anxious youth with autism spectrum disorders. J Autism Dev Disord [Internet]. 2009;39(12):1652–62. Available from: http://ovidsp.ovid.com/ovidweb.cgi?T=JS&PAGE=reference&D=psyc6&NEWS=N&AN=2009-21951-003 | Wrong publication/study design |
| Lee D. Preliminary efficacy of a gamified, behavior change techniques-based mobile application puzzlewalk on increasing physical activity and reducing anxiety in adults with autism spectrum disorder. Dissertation Abstracts International: Section B: The Sciences and Engineering [Internet]. 2021;82(2):No-Specified. Available from: http://ovidsp.ovid.com/ovidweb.cgi?T=JS&PAGE=reference&D=psyc18&NEWS=N&AN=2020-67313-230 | Wrong publication/study design |
| Frazier J. Professionals judgements of social competencies for the treatment of autism: Treatment as usual vs. social skills training. Dissertation Abstracts International: Section B: The Sciences and Engineering [Internet]. 2021;82(10):No-Specified. Available from: http://ovidsp.ovid.com/ovidweb.cgi?T=JS&PAGE=reference&D=psyc18&NEWS=N&AN=2021-50284-014 | Wrong publication/study design |
| Russell A, Cooper K, Barton S, Ensum I, Gaunt D, Horwood J, et al. Protocol for a feasibility study and randomised pilot trial of a low-intensity psychological intervention for depression in adults with autism: the Autism Depression Trial (ADEPT). BMJ Open [Internet]. 2017;7(12):e019545. Available from: http://ovidsp.ovid.com/ovidweb.cgi?T=JS&PAGE=reference&D=med14&NEWS=N&AN=29203509 | Wrong publication/study design |
| Connan A. Psychoeducational program for High-functioning autism or Asperger syndrome adults. Un programme de psychoeducation pour les adultes avec autisme sans deficience intellectuelle [Internet]. 2019;29(3):140–8. Available from: http://ovidsp.ovid.com/ovidweb.cgi?T=JS&PAGE=reference&D=psyc16&NEWS=N&AN=2019-51896-006 | Wrong publication/study design |
| Maskey M, Lowry J, Rodgers J, McConachie H, Parr JR. Reducing specific phobia/fear in young people with autism spectrum disorders (ASDs) through a virtual reality environment intervention. PLoS One [Internet]. 2014;9(7):e100374. Available from: http://ovidsp.ovid.com/ovidweb.cgi?T=JS&PAGE=reference&D=med11&NEWS=N&AN=24987957 | Wrong publication/study design |
| Results for mirtazapine for anxiety in children with autism warrant further study. Brown University Psychopharmacology Update [Internet]. 2022;33(7):3–4. Available from: https://search.ebscohost.com/login.aspx?direct=true&AuthType=ip,shib&db=jlh&AN=157235983&site=ehost-live&scope=site&custid=s8454451 | Wrong publication/study design |
| Holdman R, Vigil D, Robinson K, Shah P, Contreras A, ra Elyse. Safety and Efficacy of Medical Cannabis in Autism Spectrum Disorder Compared with Commonly Used Medications. Cannabis Cannabinoid Res [Internet]. 2021; Available from: http://ovidsp.ovid.com/ovidweb.cgi?T=JS&PAGE=reference&D=medp&NEWS=N&AN=34432543 | Wrong publication/study design |
| Berney T. Services for adults with autism spectrum disorders. Advances in Mental Health & Learning Disabilities [Internet]. 2007;1(2):45–7. Available from: https://search.ebscohost.com/login.aspx?direct=true&AuthType=ip,shib&db=jlh&AN=104583835&site=ehost-live&scope=site&custid=s8454451 | Wrong publication/study design |
| Shulman C. Services for persons with autism in Israel. Special Issue: International priorities for developing autism services via the TEACCH Model-1 [Internet]. 2000;29(1):88–97. Available from: http://ovidsp.ovid.com/ovidweb.cgi?T=JS&PAGE=reference&D=psyc3&NEWS=N&AN=2000-12075-006 | Wrong publication/study design |
| Cholewicki JM. Services received and parental perception of quality of life for children with autism spectrum disorder. Dissertation Abstracts International Section A: Humanities and Social Sciences [Internet]. 2017;77(7):No-Specified. Available from: http://ovidsp.ovid.com/ovidweb.cgi?T=JS&PAGE=reference&D=psyc14&NEWS=N&AN=2016-47708-074 | Wrong publication/study design |
| Eisenbaum E, Womack R, Dahl K, Sheppard-Jones K, Singleton P, Moseley E, et al. Serving clients with developmental disabilities in clinical practice: utilizing a universal design framework. Soc Work Ment Health [Internet]. 2022;20(4):418–31. Available from: https://search.ebscohost.com/login.aspx?direct=true&AuthType=ip,shib&db=jlh&AN=156868190&site=ehost-live&scope=site&custid=s8454451 | Wrong publication/study design |
| Johnson-Patagoc K. Silver Award: CARITAS Peace Center’s Innovations and the Neurobehavioral Center–Special Programs for Children With Psychiatric Disorders and Developmental Disabilities. Psychiatric Services [Internet]. 2003;54(11):1532–3. Available from: http://ovidsp.ovid.com/ovidweb.cgi?T=JS&PAGE=reference&D=psyc4&NEWS=N&AN=2003-09657-020 | Wrong publication/study design |
| Omodak Y, Toya K. Social adaptation support in a psychiatric day hospital for adults with autism spectrum disorder (ASD) and mental illness. Japanese Journal of Special Education [Internet]. 2017;54(5):317–26. Available from: http://ovidsp.ovid.com/ovidweb.cgi?T=JS&PAGE=reference&D=psyc14&NEWS=N&AN=2017-29831-005 | Wrong publication/study design |
| Wallace NM. Staff-child interaction therapy versus treatment as usual in community-based wraparound Services: An evaluation of feasibility and child behavior problems. Dissertation Abstracts International: Section B: The Sciences and Engineering [Internet]. 2017;78(2):No-Specified. Available from: http://ovidsp.ovid.com/ovidweb.cgi?T=JS&PAGE=reference&D=psyc14&NEWS=N&AN=2017-01060-240 | Wrong publication/study design |
| Ryan R, Neligh GL, Aderman S. A state-university collaboration to serve persons with developmental disabilities and mental health needs. Community Ment Health J [Internet]. 1997;33(5):445–54. Available from: http://ovidsp.ovid.com/ovidweb.cgi?T=JS&PAGE=reference&D=psyc3&NEWS=N&AN=1997-42580-006 | Wrong publication/study design |
| Romney JS, Garcia M, a. TF-CBT Informed Teletherapy for Children with Autism and their Families. J Child Adolesc Trauma [Internet]. 2021;14(3):415–24. Available from: https://search.ebscohost.com/login.aspx?direct=true&AuthType=ip,shib&db=jlh&AN=151860663&site=ehost-live&scope=site&custid=s8454451 | Wrong publication/study design |
| Thapar A, Livingston LA, Eyre O, Riglin L. Practitioner Review: Attention-deficit hyperactivity disorder and autism spectrum disorder – the importance of depression. Journal of Child Psychology and Psychiatry [Internet]. 2023 Jan 1 [cited 2023 Oct 13];64(1):4–15. Available from: https://onlinelibrary.wiley.com/doi/full/10.1111/jcpp.13678 | Wrong publication/study design |
| Belcher HL. On Being Autistic and in Mental Health Crisis Care. Autism Adulthood [Internet]. 2022 Sep 1 [cited 2023 Oct 13];4(3):179. Available from: /pmc/articles/PMC9908276/ | Wrong publication/study design |
| Chapman R, Botha M. Neurodivergence-informed therapy. Dev Med Child Neurol [Internet]. 2023 Mar 1 [cited 2023 Oct 13];65(3):310–7. Available from: https://pubmed.ncbi.nlm.nih.gov/36082483/ | Wrong publication/study design |
| Puleo CM, Kendall PC. Anxiety disorders in typically developing youth: Autism spectrum symptoms as a predictor of cognitive-behavioral treatment. J Autism Dev Disord [Internet]. 2011;41(3):275–86. Available from: http://ovidsp.ovid.com/ovidweb.cgi?T=JS&PAGE=reference&D=psyc8&NEWS=N&AN=2011-03834-003 | Wrong population |
| Fell L, Goshe B, Traeger L, Perez G, Iannuzzi D, Park E, et al. Acceptability of a virtual mind-body group intervention for teen siblings of children with autism spectrum disorder. J Autism Dev Disord [Internet]. 2022;No-Specified. Available from: http://ovidsp.ovid.com/ovidweb.cgi?T=JS&PAGE=reference&D=psyc18&NEWS=N&AN=2022-69978-001 | Wrong population |
| Jones J, Blinkhorn A, McQueen M, Hewett L, Mills‐Rogers M, Hall L, et al. The adaptation and feasibility of dialectical behaviour therapy for adults with intellectual developmental disabilities and transdiagnoses: A pilot community‐based randomized controlled trial. Journal of Applied Research in Intellectual Disabilities [Internet]. 2021;34(3):805–17. Available from: https://search.ebscohost.com/login.aspx?direct=true&AuthType=ip,shib&db=jlh&AN=149879693&site=ehost-live&scope=site&custid=s8454451 | Wrong population |
| dosReis S, N’Dri L, Ross M, Camelo Castillo W, Reeves G, Butler B. Care Management for Youth With Comorbid Developmental and Mental Health Conditions: A Discrete Choice Experiment Pilot Study. Acad Pediatr [Internet]. 2020;20(2):241–9. Available from: http://ovidsp.ovid.com/ovidweb.cgi?T=JS&PAGE=reference&D=med17&NEWS=N&AN=31128382 | Wrong population |
| Settipani CA, Puleo CM, Conner BT, Kendall PC. Characteristics and anxiety symptom presentation associated with autism spectrum traits in youth with anxiety disorders. J Anxiety Disord [Internet]. 2012;26(3):459–67. Available from: http://ovidsp.ovid.com/ovidweb.cgi?T=JS&PAGE=reference&D=psyc9&NEWS=N&AN=2012-06706-011 | Wrong population |
| Devapriam J, Fosker H, Chester V, Gangadharan S, Hiremath A, Alex, et al. Characteristics and outcomes of patients with intellectual disability admitted to a specialist inpatient rehabilitation service. Journal of Intellectual Disabilities [Internet]. 2020;24(1):21–34. Available from: http://ovidsp.ovid.com/ovidweb.cgi?T=JS&PAGE=reference&D=psyc17&NEWS=N&AN=2020-13514-003 | Wrong population |
| Saunder L, Knight RA. CitySCaPE: Moving beyond indifference in education for pre-registration nurses about learning disability. Nurse Educ Pract [Internet]. 2017;26:82–8. Available from: http://ovidsp.ovid.com/ovidweb.cgi?T=JS&PAGE=reference&D=med14&NEWS=N&AN=28772114 | Wrong population |
| Lobregt-van Buuren E, Sizoo B, Mevissen L, de Jongh A. Eye Movement Desensitization and Reprocessing (EMDR) Therapy as a Feasible and Potential Effective Treatment for Adults with Autism Spectrum Disorder (ASD) and a History of Adverse Events. J Autism Dev Disord [Internet]. 2019 Jan 15 [cited 2023 Oct 13];49(1):151–64. Available from: https://pubmed.ncbi.nlm.nih.gov/30047096/ | Wrong population |
| Tchanturia K, Dandil Y, Li Z, Smith K, Leslie M, Byford S. A novel approach for autism spectrum condition patients with eating disorders: Analysis of treatment cost-savings. Eur Eat Disord Rev [Internet]. 2021 May 1 [cited 2023 Oct 13];29(3):514–8. Available from: https://pubmed.ncbi.nlm.nih.gov/32648631/ | Wrong population |
| Maskey M, Rodgers J, Ingham B, Freeston M, Evans G, Labus M, et al. Using Virtual Reality Environments to Augment Cognitive Behavioral Therapy for Fears and Phobias in Autistic Adults. Autism Adulthood [Internet]. 2019;1(2):134–45. Available from: http://ovidsp.ovid.com/ovidweb.cgi?T=JS&PAGE=reference&D=pmnm&NEWS=N&AN=31032480 | Wrong population |
| Brugha T, Tyrer F, Leaver A, Lewis S, Seaton S, Morgan Z, et al. Testing adults by questionnaire for social and communication disorders, including autism spectrum disorders, in an adult mental health service population. Int J Methods Psychiatr Res [Internet]. 2020;29(1). Available from: http://ovidsp.ovid.com/ovidweb.cgi?T=JS&PAGE=reference&D=psyc17&NEWS=N&AN=2020-02747-001 | Wrong population |
| Oshima F, Murata T, Ohtani T, Seto M, Shimizu E. A preliminary study of schema therapy for young adults with high-functioning autism spectrum disorder: a single-arm, uncontrolled trial. BMC Res Notes [Internet]. 2021;14(1):158. Available from: http://ovidsp.ovid.com/ovidweb.cgi?T=JS&PAGE=reference&D=med19&NEWS=N&AN=33926531 | Wrong population |
| Kiep M, Spek AA, Hoeben L. Mindfulness-based therapy in adults with an autism spectrum disorder: Do treatment effects last? Mindfulness (N Y) [Internet]. 2015;6(3):637–44. Available from: http://ovidsp.ovid.com/ovidweb.cgi?T=JS&PAGE=reference&D=psyc12&NEWS=N&AN=2014-11532-001 | Wrong population |
| Spek AA, van Ham NC, Nyklicek I. Mindfulness-based therapy in adults with an autism spectrum disorder: A randomized controlled trial. Res Dev Disabil [Internet]. 2013;34(1):246–53. Available from: http://ovidsp.ovid.com/ovidweb.cgi?T=JS&PAGE=reference&D=psyc10&NEWS=N&AN=2012-32676-027 | Wrong population |
| Spain D, Blainey SH, Vaillancourt K. Group cognitive behaviour therapy (CBT) for social interaction anxiety in adults with autism spectrum disorders (ASD). Res Autism Spectr Disord [Internet]. 2017;41:20–30. Available from: http://ovidsp.ovid.com/ovidweb.cgi?T=JS&PAGE=reference&D=psyc14&NEWS=N&AN=2017-40905-004 | Wrong population |
| Russell A, Gaunt DM, Cooper K, Barton S, Horwood J, Kessler D, et al. “The feasibility of low-intensity psychological therapy for depression co-occurring with autism in adults: The Autism Depression Trial (ADEPT)-A pilot randomised controlled trial”: Erratum. Autism [Internet]. 2020;24(6):1583. Available from: http://ovidsp.ovid.com/ovidweb.cgi?T=JS&PAGE=reference&D=psyc18&NEWS=N&AN=2020-54243-024 | Wrong population |
| Blainey SH, Rumball F, Mercer L, Evans LJ, Beck A. An evaluation of the effectiveness of psychological therapy in reducing general psychological distress for adults with autism spectrum conditions and comorbid mental health problems. Clin Psychol Psychother [Internet]. 2017;24(6):O1474–84. Available from: http://ovidsp.ovid.com/ovidweb.cgi?T=JS&PAGE=reference&D=psyc14&NEWS=N&AN=2017-32676-001 | Wrong population |
| Wijker C, Leontjevas R, Spek A, Enders-Slegers MJ. Effects of dog assisted therapy for adults with autism spectrum disorder: An exploratory randomized controlled trial. J Autism Dev Disord [Internet]. 2020;50(6):2153–63. Available from: http://ovidsp.ovid.com/ovidweb.cgi?T=JS&PAGE=reference&D=psyc17&NEWS=N&AN=2019-16405-001 | Wrong population |
| Sizoo BB, Kuiper E. Cognitive behavioural therapy and mindfulness based stress reduction may be equally effective in reducing anxiety and depression in adults with autism spectrum disorders. Res Dev Disabil [Internet]. 2017;64:47–55. Available from: http://ovidsp.ovid.com/ovidweb.cgi?T=JS&PAGE=reference&D=psyc14&NEWS=N&AN=2017-28543-006 | Wrong population |
| Harrison KB, McCredie MN, Reddy MK, Krishnan A, Engstrom A, Posey YS, et al. Assessing Autism Spectrum Disorder in Intellectually Able Adults with the Personality Assessment Inventory: Normative Data and a Novel Supplemental Indicator. J Autism Dev Disord [Internet]. 2020;50(11):3935–43. Available from: http://ovidsp.ovid.com/ovidweb.cgi?T=JS&PAGE=reference&D=med17&NEWS=N&AN=32170539 | Wrong population |
| Hare DJ, Gracey C, Wood C. Anxiety in high-functioning autism: A pilot study of experience sampling using a mobile platform. Autism [Internet]. 2016;20(6):730–43. Available from: http://ovidsp.ovid.com/ovidweb.cgi?T=JS&PAGE=reference&D=psyc13&NEWS=N&AN=2016-33459-009 | Wrong population |
| Flygare O, Andersson E, Ringberg H, Hellstadius AC, Edbacken J, En, et al. Adapted cognitive behavior therapy for obsessive-compulsive disorder with co-occurring autism spectrum disorder: A clinical effectiveness study. Autism [Internet]. 2020;24(1):190–9. Available from: http://ovidsp.ovid.com/ovidweb.cgi?T=JS&PAGE=reference&D=psyc17&NEWS=N&AN=2019-80393-015 | Wrong population |
| Pahnke J, Hirvikoski T, Bjureberg J, Bolte S, Jokinen J, Bohman B, et al. Acceptance and Commitment Therapy for autistic adults: An open pilot study in a psychiatric outpatient context. J Contextual Behav Sci [Internet]. 2019;13:34–41. Available from: http://ovidsp.ovid.com/ovidweb.cgi?T=JS&PAGE=reference&D=psyc16&NEWS=N&AN=2019-51847-005 | Wrong population |
| Russell J, Maguire S, Hunt GE, Kesby A, Suraev A, Stuart J, et al. Intranasal oxytocin in the treatment of anorexia nervosa: Randomized controlled trial during re-feeding. Psychoneuroendocrinology [Internet]. 2018 Jan 1 [cited 2023 Oct 13];87:83–92. Available from: https://pubmed.ncbi.nlm.nih.gov/29049935/ | Wrong population |
| Lodder A, Papadopoulos C, Randhawa G. SOLACE: A Psychosocial Stigma Protection Intervention to Improve the Mental Health of Parents of Autistic Children—A Feasibility Randomised Controlled Trial. J Autism Dev Disord [Internet]. 2020 Dec 1 [cited 2023 Oct 13];50(12):4477–91. Available from: https://link.springer.com/article/10.1007/s10803-020-04498-0 | Wrong population |
| Helverschou SB, Brunvold AR, Arnevik EA. Treating Patients With Co-occurring Autism Spectrum Disorder and Substance Use Disorder: A Clinical Explorative Study. Subst Abuse [Internet]. 2019 Apr 1 [cited 2023 Oct 13];13. Available from: /pmc/articles/PMC6472168/ | Wrong population |
| Vaz I. Visual symbols in healthcare settings for children with learning disabilities and autism spectrum disorder. Br J Nurs [Internet]. 2013;22(3):156–9. Available from: http://ovidsp.ovid.com/ovidweb.cgi?T=JS&PAGE=reference&D=med10&NEWS=N&AN=23411823 | Wrong population |
| Cannella-Malone HI, Tullis CA, Kazee AR. Using Antecedent Exercise to Decrease Challenging Behavior in Boys With Developmental Disabilities and an Emotional Disorder. J Posit Behav Interv [Internet]. 2011;13(4):230–9. Available from: https://search.ebscohost.com/login.aspx?direct=true&AuthType=ip,shib&db=jlh&AN=104706805&site=ehost-live&scope=site&custid=s8454451 | Wrong population |
| Perera B, Shaikh A, Singh N. The use of Community Treatment Orders in an intellectual disability service. Adv Ment Health Intellect Disabil [Internet]. 2013;7(3):129–34. Available from: http://ovidsp.ovid.com/ovidweb.cgi?T=JS&PAGE=reference&D=psyc10&NEWS=N&AN=2013-19232-001 | Wrong population |
| Ames JL, Massolo ML, Davignon MN, Qian Y, Cerros HJ, Croen LA. Transitioning youth with autism spectrum disorders and other special health care needs into adult primary care: A provider survey. Autism [Internet]. 2021;25(3):731–43. Available from: https://search.ebscohost.com/login.aspx?direct=true&AuthType=ip,shib&db=jlh&AN=150211309&site=ehost-live&scope=site&custid=s8454451 | Wrong population |
| Gentile JP, Gillig PM, Stinson K, Jensen J. Toward impacting medical and psychiatric comorbidities in persons with intellectual/developmental disabilities: An initial prospective analysis. Innov Clin Neurosci [Internet]. 2014;11(11):22–6. Available from: http://ovidsp.ovid.com/ovidweb.cgi?T=JS&PAGE=reference&D=psyc11&NEWS=N&AN=2015-01254-004 | Wrong population |
| Fang H, Li Y, Wu M, Gan F. Study on the validity of the theoretical paradigm of art therapy for vulnerable children. Disabil Rehabil Assist Technol [Internet]. 2022;1–12. Available from: http://ovidsp.ovid.com/ovidweb.cgi?T=JS&PAGE=reference&D=medp&NEWS=N&AN=35584289 | Wrong population |
| Temkin AB, Beaumont R, Wkya K, Hariton JR, Flye BL, Sheridan E, et al. Secret agent society: A randomized controlled trial of a transdiagnostic youth social skills group treatment. Res Child Adolesc Psychopathol [Internet]. 2022;No-Specified. Available from: http://ovidsp.ovid.com/ovidweb.cgi?T=JS&PAGE=reference&D=psyc18&NEWS=N&AN=2022-56586-001 | Wrong population |
| Jones EA, Fiani T, Stewart JL, Neil N, McHugh S, Fienup DM. Randomized controlled trial of a sibling support group: Mental health outcomes for siblings of children with autism. Autism [Internet]. 2020;24(6):1468–81. Available from: https://search.ebscohost.com/login.aspx?direct=true&AuthType=ip,shib&db=jlh&AN=144708745&site=ehost-live&scope=site&custid=s8454451 | Wrong population |
| Koch AD, Dobrindt J, Schutzwohl M. Prevalence of psychotropic medication and factors associated with antipsychotic treatment in adults with intellectual disabilities: A cross-sectional, epidemiological study in germany. Journal of Intellectual Disability Research [Internet]. 2021;No-Specified. Available from: http://ovidsp.ovid.com/ovidweb.cgi?T=JS&PAGE=reference&D=psyc18&NEWS=N&AN=2021-05738-001 | Wrong population |
| Sajith SG, Chiu J, Wong W, Chiam PC. Perceptions of psychiatry residents and non-residents on training needs and care related to patients with intellectual disability and mental health problems: A study from Singapore. J Intellect Dev Disabil [Internet]. 2019;44(1):73–80. Available from: http://ovidsp.ovid.com/ovidweb.cgi?T=JS&PAGE=reference&D=psyc16&NEWS=N&AN=2019-01097-008 | Wrong population |
| Sturmey P, Newton J, Cowley A, Bouras N, Holt G. The PAS-ADD Checklist: Independent replication of its psychometric properties in a community sample. The British Journal of Psychiatry [Internet]. 2005;186(4):319–23. Available from: http://ovidsp.ovid.com/ovidweb.cgi?T=JS&PAGE=reference&D=psyc4&NEWS=N&AN=2005-04200-010 | Wrong population |
| Allott K, van-der-El K, Bryce S, Hamilton M, Adams S, Burgat L, et al. Need for clinical neuropsychological assessment in headspace youth mental health services: A national survey of providers. Aust J Psychol [Internet]. 2019;71(2):108–16. Available from: http://ovidsp.ovid.com/ovidweb.cgi?T=JS&PAGE=reference&D=psyc16&NEWS=N&AN=2018-45459-001 | Wrong population |
| Kalb LG, Beasley J, Caoili A, Klein A. Improvement in mental health outcomes and caregiver service experiences associated with the START program. Am J Intellect Dev Disabil [Internet]. 2019;124(1):25–34. Available from: http://ovidsp.ovid.com/ovidweb.cgi?T=JS&PAGE=reference&D=psyc16&NEWS=N&AN=2019-19662-003 | Wrong population |
| Tchanturia K, Larsson E, Adamson J. How anorexia nervosa patients with high and low autistic traits respond to group Cognitive Remediation Therapy. BMC Psychiatry [Internet]. 2016;16. Available from: http://ovidsp.ovid.com/ovidweb.cgi?T=JS&PAGE=reference&D=psyc13&NEWS=N&AN=2016-47273-001 | Wrong population |
| Yaseen W, Steckle V, Sgro M, Barozzino T, Suleman S. Exploring stakeholder service navigation needs for children with developmental and mental health diagnoses. Journal of Developmental and Behavioral Pediatrics [Internet]. 2021;42(7):553–60. Available from: http://ovidsp.ovid.com/ovidweb.cgi?T=JS&PAGE=reference&D=psyc18&NEWS=N&AN=2022-31971-005 | Wrong population |
| Peens A, Pienaar AE, Nienaber AW. The effect of different intervention programmes on the self-concept and motor proficiency of 7- to 9-year-old children with DCD. Child Care Health Dev [Internet]. 2008;34(3):316–28. Available from: http://ovidsp.ovid.com/ovidweb.cgi?T=JS&PAGE=reference&D=psyc6&NEWS=N&AN=2008-04666-005 | Wrong population |
| Wentz E, Nyden A, Krevers B. Development of an internet-based support and coaching model for adolescents and young adults with ADHD and autism spectrum disorders: A pilot study. Eur Child Adolesc Psychiatry [Internet]. 2012;21(11):611–22. Available from: http://ovidsp.ovid.com/ovidweb.cgi?T=JS&PAGE=reference&D=psyc9&NEWS=N&AN=2012-30484-003 | Wrong population |
| Horwood J, Cooper K, Harvey H, Davies L, Russell A. The experience of autistic adults accessing adapted cognitive behaviour therapy: ADEPT (Autism Depression Trial) qualitative evaluation. Res Autism Spectr Disord. 2021 Aug 1;86:101802. | Wrong population |
| Conaughton RJ, Donovan CL, March S. Efficacy of an internet-based CBT program for children with comorbid High Functioning Autism Spectrum Disorder and anxiety: A randomised controlled trial. J Affect Disord [Internet]. 2017 Aug 15 [cited 2023 Oct 13];218:260–8. Available from: https://pubmed.ncbi.nlm.nih.gov/28477505/ | Wrong intervention |
| Zhang R, Jia MX, Zhang JS, Xu XJ, Shou XJ, Zhang XT, et al. Transcutaneous electrical acupoint stimulation in children with autism and its impact on plasma levels of arginine-vasopressin and oxytocin: a prospective single-blinded controlled study. Res Dev Disabil [Internet]. 2012 Jul [cited 2023 Oct 13];33(4):1136–46. Available from: https://pubmed.ncbi.nlm.nih.gov/22502839/ | Wrong intervention |
| Yamada T, Miura Y, Oi M, Akatsuka N, Tanaka K, Tsukidate N, et al. Examining the Treatment Efficacy of PEERS in Japan: Improving Social Skills Among Adolescents with Autism Spectrum Disorder. J Autism Dev Disord [Internet]. 2020 Mar 1 [cited 2023 Oct 13];50(3):976–97. Available from: https://pubmed.ncbi.nlm.nih.gov/31823217/ | Wrong intervention |
| Wijnhoven LAMW, Creemers DHM, Vermulst AA, Lindauer RJL, Otten R, Engels RCME, et al. Effects of the video game “Mindlight” on anxiety of children with an autism spectrum disorder: A randomized controlled trial. J Behav Ther Exp Psychiatry [Internet]. 2020 Sep 1 [cited 2023 Oct 13];68. Available from: https://pubmed.ncbi.nlm.nih.gov/32155470/ | Wrong intervention |
| Takei R, Matsuo J, Takahashi H, Uchiyama T, Kunugi H, Kamio Y. Verification of the utility of the social responsiveness scale for adults in non-clinical and clinical adult populations in Japan. BMC Psychiatry [Internet]. 2014 Nov 18 [cited 2023 Oct 13];14(1). Available from: https://pubmed.ncbi.nlm.nih.gov/25403232/ | Wrong intervention |
| Simonoff E, Mowlem F, Pearson O, Anagnostou E, Donnelly C, Hollander E, et al. Citalopram Did Not Significantly Improve Anxiety in Children with Autism Spectrum Disorder Undergoing Treatment for Core Symptoms: Secondary Analysis of a Trial to Reduce Repetitive Behaviors. J Child Adolesc Psychopharmacol [Internet]. 2022 May 1 [cited 2023 Oct 13];32(4):233–41. Available from: https://pubmed.ncbi.nlm.nih.gov/35501967/ | Wrong intervention |
| Pellicano E, Brett S, den Houting J, Heyworth M, Magiati I, Steward R, et al. COVID-19, social isolation and the mental health of autistic people and their families: A qualitative study. Autism [Internet]. 2022 May 1 [cited 2023 Oct 13];26(4):914–27. Available from: https://journals.sagepub.com/doi/full/10.1177/13623613211035936 | Wrong intervention |
| Ozsivadjian A, Hibberd C, Hollocks MJ. Brief report: The use of self-report measures in young people with autism spectrum disorder to access symptoms of anxiety, depression and negative thoughts. J Autism Dev Disord [Internet]. 2014 Sep 8 [cited 2023 Oct 13];44(4):969–74. Available from: https://link.springer.com/article/10.1007/s10803-013-1937-1 | Wrong intervention |
| McLay LL, France KG, Blampied NM, Hunter JE, van Deurs JR, Woodford EC, et al. Collateral Child and Parent Effects of Function‑Based Behavioral Interventions for Sleep Problems in Children and Adolescents with Autism. J Autism Dev Disord [Internet]. 2022 May 1 [cited 2023 Oct 13];52(5):2258–73. Available from: https://pubmed.ncbi.nlm.nih.gov/34110556/ | Wrong intervention |
| Mazzucchelli TG, Jenkins M, Sofronoff K. Building Bridges Triple P: Pilot study of a behavioural family intervention for adolescents with autism spectrum disorder. Res Dev Disabil [Internet]. 2018 May 1 [cited 2023 Oct 13];76:46–55. Available from: https://pubmed.ncbi.nlm.nih.gov/29550648/ | Wrong intervention |
| Maddox BB, Kang-Yi CD, Brodkin ES, Mandell DS. Treatment utilization by adults with autism and co-occurring anxiety or depression. Res Autism Spectr Disord [Internet]. 2018 Jul 1 [cited 2023 Oct 13];51:32–7. Available from: /pmc/articles/PMC6181232/ | Wrong intervention |
| Loring WA, L. Johnston R, Shui AM, Malow BA. Impact of a Brief Behavioral Intervention for Insomnia on Daytime Behaviors in Adolescents with Autism Spectrum Disorders. J Contemp Psychother [Internet]. 2018 Sep 1 [cited 2023 Oct 13];48(3):165–77. Available from: https://link.springer.com/article/10.1007/s10879-018-9381-3 | Wrong intervention |
| Lei J, Sukhodolsky DG, Abdullahi SM, Braconnier ML, Ventola P. Reduced anxiety following pivotal response treatment in young children with autism spectrum disorder. Res Autism Spectr Disord [Internet]. 2017 Nov 1 [cited 2023 Oct 13];43–44:1–7. Available from: /pmc/articles/PMC5761743/ | Wrong intervention |
| Gwynette MF, Lowe DW, Henneberry EA, Sahlem GL, Wiley MG, Alsarraf H, et al. Treatment of Adults with Autism and Major Depressive Disorder Using Transcranial Magnetic Stimulation: An Open Label Pilot Study. Autism Res [Internet]. 2020 Mar 1 [cited 2023 Oct 13];13(3):346–51. Available from: https://pubmed.ncbi.nlm.nih.gov/31944611/ | Wrong intervention |
| Grant PJ, Joseph LA, Farmer CA, Luckenbaugh DA, Lougee LC, Zarate CA, et al. 12-Week, Placebo-Controlled Trial of Add-on Riluzole in the Treatment of Childhood-Onset Obsessive–Compulsive Disorder. Neuropsychopharmacology [Internet]. 2014 [cited 2023 Oct 13];39(6):1453. Available from: /pmc/articles/PMC3988548/ | Wrong intervention |
| Danforth AL, Grob CS, Struble C, Feduccia AA, Walker N, Jerome L, et al. Reduction in social anxiety after MDMA-assisted psychotherapy with autistic adults: a randomized, double-blind, placebo-controlled pilot study. Psychopharmacology (Berl) [Internet]. 2018 Nov 1 [cited 2023 Oct 13];235(11):3137–48. Available from: https://pubmed.ncbi.nlm.nih.gov/30196397/ | Wrong intervention |
| Dandil Y, Smith K, Adamson J, Tchanturia K. Individual cognitive remediation therapy benefits for patients with anorexia nervosa and high autistic features. European Eating Disorders Review [Internet]. 2020 Jan 1 [cited 2023 Oct 13];28(1):87–91. Available from: https://onlinelibrary.wiley.com/doi/full/10.1002/erv.2707 | Wrong intervention |
| Ceranoglu TA, Wozniak J, Fried R, Galdo M, Hoskova B, DeLeon Fong M, et al. A Retrospective Chart Review of Buspirone for the Treatment of Anxiety in Psychiatrically Referred Youth with High-Functioning Autism Spectrum Disorder. J Child Adolesc Psychopharmacol [Internet]. 2019 Feb 2 [cited 2023 Oct 13];29(1):28. Available from: /pmc/articles/PMC6354602/ | Wrong intervention |
| Bejarano-Martín Á, Canal-Bedia R, Magán-Maganto M, Hernández Fabián A, Calvarro Castañeda AL, Manso de Dios S, et al. Effect of a Focused Social and Communication Intervention on Preterm Children with ASD: A Pilot Study. J Autism Dev Disord [Internet]. 2022 Apr 1 [cited 2023 Oct 13];52(4):1725–40. Available from: https://pubmed.ncbi.nlm.nih.gov/33991290/ | Wrong intervention |
| Adamson J, Leppanen J, Murin M, Tchanturia K. Effectiveness of emotional skills training for patients with anorexia nervosa with autistic symptoms in group and individual format. Eur Eat Disord Rev [Internet]. 2018 Jul 1 [cited 2023 Oct 13];26(4):367–75. Available from: https://pubmed.ncbi.nlm.nih.gov/29607561/ | Wrong intervention |
| Rossi LP, Lovisi GM, Abelha L, Gomide M. [Virtual Paths and Autism: access to health services from the perspective of Social Network Analysis]. Caminhos Virtuais e Autismo: acesso aos servicos de saude na perspectiva da Analise de Redes Sociais [Internet]. 2018;23(10):3319–26. Available from: http://ovidsp.ovid.com/ovidweb.cgi?T=JS&PAGE=reference&D=med15&NEWS=N&AN=30365851 | Wrong intervention |
| Holl, er E, Kaplan A, Cartwright C, Reichman D. Venlafaxine in children, adolescents, and young adults with autism spectrum disorders: An open retrospective clinical report. J Child Neurol [Internet]. 2000;15(2):132–5. Available from: http://ovidsp.ovid.com/ovidweb.cgi?T=JS&PAGE=reference&D=psyc3&NEWS=N&AN=2000-13928-004 | Wrong intervention |
| Gerrard D, Rhodes J, Lee R, Ling J. Using positive behavioural support (PBS) for STOMP medication challenge. Adv Ment Health Intellect Disabil [Internet]. 2019;13(3):102–12. Available from: http://ovidsp.ovid.com/ovidweb.cgi?T=JS&PAGE=reference&D=psyc16&NEWS=N&AN=2019-42666-002 | Wrong intervention |
| Smith MJ, Pinto RM, Dawalt L, Smith JD, Sherwood K, Miles R, et al. Using community-engaged methods to adapt virtual reality job-interview training for transition-age youth on the autism spectrum. Res Autism Spectr Disord [Internet]. 2020;71. Available from: http://ovidsp.ovid.com/ovidweb.cgi?T=JS&PAGE=reference&D=pmnm&NEWS=N&AN=34667480 | Wrong intervention |
| Ming X, Gordon E, Kang N, Wagner GC. Use of clonidine in children with autism spectrum disorders. Brain Dev [Internet]. 2008;30(7):454–60. Available from: http://ovidsp.ovid.com/ovidweb.cgi?T=JS&PAGE=reference&D=psyc6&NEWS=N&AN=2008-09076-003 | Wrong intervention |
| Chiri G, Warfield ME. Unmet need and problems accessing core health care services for children with autism spectrum disorder. Matern Child Health J [Internet]. 2012;16(5):1081–91. Available from: http://ovidsp.ovid.com/ovidweb.cgi?T=JS&PAGE=reference&D=psyc9&NEWS=N&AN=2012-17469-019 | Wrong intervention |
| Taleb MO. The treatment of children and adolescents autism: Establishing a program in Algers (2005/2008). Traitement de l’autisme de l’enfant et de l’adolescent: Mise en place d’un programme a Alger (2005/2008) [Internet]. 2010;58(3):159–67. Available from: http://ovidsp.ovid.com/ovidweb.cgi?T=JS&PAGE=reference&D=psyc7&NEWS=N&AN=2010-07259-010 | Wrong intervention |
| Brookman-Frazee L, Chlebowski C, Villodas M, Roesch S, Martinez K, ra. Training community therapists to deliver an individualized mental health intervention for autism spectrum disorder: Changes in caregiver outcomes and mediating role on child outcomes. J Am Acad Child Adolesc Psychiatry [Internet]. 2021;60(3):355–66. Available from: http://ovidsp.ovid.com/ovidweb.cgi?T=JS&PAGE=reference&D=psyc18&NEWS=N&AN=2020-83045-001 | Wrong intervention |
| Brookman-Frazee LI, Drahota A, Stadnick N. Training community mental health therapists to deliver a package of evidence-based practice strategies for school-age children with autism spectrum disorders: A pilot study. J Autism Dev Disord [Internet]. 2012;42(8):1651–61. Available from: http://ovidsp.ovid.com/ovidweb.cgi?T=JS&PAGE=reference&D=psyc9&NEWS=N&AN=2012-19817-013 | Wrong intervention |
| Dyson MW, Chlebowski C, Brookman-Frazee L. Therapists’ adaptations to an intervention to reduce challenging behaviors in children with autism spectrum disorder in publicly funded mental health services. J Autism Dev Disord [Internet]. 2019;49(3):924–34. Available from: http://ovidsp.ovid.com/ovidweb.cgi?T=JS&PAGE=reference&D=psyc16&NEWS=N&AN=2018-54684-001 | Wrong intervention |
| Drahota A, Stadnick N, Brookman-Frazee L. Therapist perspectives on training in a package of evidence-based practice strategies for children with autism spectrum disorders served in community mental health clinics. Administration and Policy in Mental Health and Mental Health Services Research [Internet]. 2014;41(1):114–25. Available from: http://ovidsp.ovid.com/ovidweb.cgi?T=JS&PAGE=reference&D=psyc11&NEWS=N&AN=2014-00152-007 | Wrong intervention |
| Anderberg E, Cox JC, Tass ESN, Erekson DM, Gabrielsen TP, Warren JS, et al. Sticking with it: Psychotherapy outcomes for adults with autism spectrum disorder in a university counseling center setting. Autism Research [Internet]. 2017;10(12):2048–55. Available from: http://ovidsp.ovid.com/ovidweb.cgi?T=JS&PAGE=reference&D=psyc14&NEWS=N&AN=2017-35687-001 | Wrong intervention |
| Siegel M, Milligan B, Chemelski B, Payne D, Ellsworth B, Harmon J, et al. Specialized inpatient psychiatry for serious behavioral disturbance in autism and intellectual disability. J Autism Dev Disord [Internet]. 2014;44(12):3026–32. Available from: http://ovidsp.ovid.com/ovidweb.cgi?T=JS&PAGE=reference&D=med11&NEWS=N&AN=24925543 | Wrong intervention |
| van den Heuvel RM, Wensing M, Geurts HM, Teunisse JP. The social support network of adults with an autism spectrum condition: An exploration using the network in action-questionnaire. J Autism Dev Disord [Internet]. 2022;No-Specified. Available from: http://ovidsp.ovid.com/ovidweb.cgi?T=JS&PAGE=reference&D=psyc18&NEWS=N&AN=2022-35600-001 | Wrong intervention |
| Hillier AJ, Fish T, Siegel JH, Beversdorf DQ. Social and vocational skills training reduces self-reported anxiety and depression among young adults on the autism spectrum. J Dev Phys Disabil [Internet]. 2011;23(3):267–76. Available from: http://ovidsp.ovid.com/ovidweb.cgi?T=JS&PAGE=reference&D=psyc8&NEWS=N&AN=2011-08445-008 | Wrong intervention |
| Malihi M, Nguyen J, Cardy RE, Eldon S, Petta C, Kushki A. Short report: Evaluating the safety and usability of head-mounted virtual reality compared to monitor-displayed video for children with autism spectrum disorder. Autism [Internet]. 2020;24(7):1924–9. Available from: http://ovidsp.ovid.com/ovidweb.cgi?T=JS&PAGE=reference&D=psyc18&NEWS=N&AN=2020-74826-029 | Wrong intervention |
| Gaigg SB, Flaxman PE, McLaven G, Shah R, Bowler DM, Meyer B, et al. Self-guided mindfulness and cognitive behavioural practices reduce anxiety in autistic adults: A pilot 8-month waitlist-controlled trial of widely available online tools. Autism [Internet]. 2020;24(4):867–83. Available from: http://ovidsp.ovid.com/ovidweb.cgi?T=JS&PAGE=reference&D=psyc18&NEWS=N&AN=2020-36416-006 | Wrong intervention |
| Thorkelson G, Laughlin SF, Turner KS, Ober N, H, en BL. Selective serotonin reuptake inhibitor monotherapy for anxiety disorders in children and adolescents with autism spectrum disorder: A chart review. J Child Adolesc Psychopharmacol [Internet]. 2019;29(9):705–11. Available from: http://ovidsp.ovid.com/ovidweb.cgi?T=JS&PAGE=reference&D=psyc16&NEWS=N&AN=2019-40359-001 | Wrong intervention |
| Courturier JL, Nicolson R. A retrospective assessment of citalopram in children and adolescents with pervasive developmental disorders. J Child Adolesc Psychopharmacol [Internet]. 2002;12(3):243–8. Available from: http://ovidsp.ovid.com/ovidweb.cgi?T=JS&PAGE=reference&D=psyc4&NEWS=N&AN=2002-04850-008 | Wrong intervention |
| Schohl KA, Van Hecke A V, Carson AM, Dolan B, Karst J, Stevens S. A replication and extension of the PEERS intervention: Examining effects on social skills and social anxiety in adolescents with autism spectrum disorders. J Autism Dev Disord [Internet]. 2014;44(3):532–45. Available from: http://ovidsp.ovid.com/ovidweb.cgi?T=JS&PAGE=reference&D=psyc11&NEWS=N&AN=2013-27307-001 | Wrong intervention |
| McVey AJ, Dolan BK, Willar KS, Pleiss S, Karst JS, Casnar CL, et al. A replication and extension of the PEERS for young adults social skills intervention: Examining effects on social skills and social anxiety in young adults with autism spectrum disorder. J Autism Dev Disord [Internet]. 2016;46(12):3739–54. Available from: http://ovidsp.ovid.com/ovidweb.cgi?T=JS&PAGE=reference&D=psyc13&NEWS=N&AN=2016-44909-001 | Wrong intervention |
| Golubchik P, Sever J, Weizman A. Reboxetine treatment for autistic spectrum disorder of pediatric patients with depressive and inattentive/hyperactive symptoms: An open-label trial. Clin Neuropharmacol [Internet]. 2013;36(2):37–41. Available from: http://ovidsp.ovid.com/ovidweb.cgi?T=JS&PAGE=reference&D=psyc10&NEWS=N&AN=2013-09660-001 | Wrong intervention |
| Singh NN, Lancioni GE, Medvedev ON, Hwang YS, Myers RE. Real-Time Telehealth Treatment Team Consultation for Self-Injury by Individuals with Autism Spectrum Disorder. Adv Neurodev Disord [Internet]. 2021;5(2):170–82. Available from: http://ovidsp.ovid.com/ovidweb.cgi?T=JS&PAGE=reference&D=pmnm&NEWS=N&AN=33490369 | Wrong intervention |
| Parker KJ, Oztan O, Libove RA, Mohsin N, Karhson DS, Sumiyoshi RD, et al. A randomized placebo-controlled pilot trial shows that intranasal vasopressin improves social deficits in children with autism. Sci Transl Med [Internet]. 2019;11(491). Available from: http://ovidsp.ovid.com/ovidweb.cgi?T=JS&PAGE=reference&D=med16&NEWS=N&AN=31043522 | Wrong intervention |
| McDougle CJ, Thom RP, Ravich, ran CT, Palumbo ML, Politte LC, et al. A randomized double-blind, placebo-controlled pilot trial of mirtazapine for anxiety in children and adolescents with autism spectrum disorder. Neuropsychopharmacology [Internet]. 2022;47(6):1263–70. Available from: http://ovidsp.ovid.com/ovidweb.cgi?T=JS&PAGE=reference&D=psyc18&NEWS=N&AN=2022-42059-001 | Wrong intervention |
| Marino F, Chila P, Failla C, Minutoli R, Vetrano N, Luraschi C, et al. Psychological Interventions for Children with Autism during the COVID-19 Pandemic through a Remote Behavioral Skills Training Program. J Clin Med [Internet]. 2022;11(5). Available from: http://ovidsp.ovid.com/ovidweb.cgi?T=JS&PAGE=reference&D=pmnm&NEWS=N&AN=35268284 | Wrong intervention |
| Caplan B, Chlebowski C, May G, Baker-Ericzen MJ, Connor W, Brookman-Frazee L. Psychotropic Medication Use by Children with Autism Served in Publicly Funded Mental Health Settings. J Dev Behav Pediatr [Internet]. 2022;43(5):252–61. Available from: http://ovidsp.ovid.com/ovidweb.cgi?T=JS&PAGE=reference&D=mesx&NEWS=N&AN=34855690 | Wrong intervention |
| Arnold LE, Luna RA, Williams K, Chan J, Parker RA, Wu Q, et al. Probiotics for gastrointestinal symptoms and quality of life in autism: A placebo-controlled pilot trial. J Child Adolesc Psychopharmacol [Internet]. 2019;29(9):659–69. Available from: http://ovidsp.ovid.com/ovidweb.cgi?T=JS&PAGE=reference&D=psyc16&NEWS=N&AN=2019-52988-001 | Wrong intervention |
| Siegel M, Beresford CA, Bunker M, Verdi M, Vishnevetsky D, Karlsson C, et al. Preliminary investigation of lithium for mood disorder symptoms in children and adolescents with autism spectrum disorder. J Child Adolesc Psychopharmacol [Internet]. 2014;24(7):399–402. Available from: http://ovidsp.ovid.com/ovidweb.cgi?T=JS&PAGE=reference&D=psyc11&NEWS=N&AN=2014-38849-006 | Wrong intervention |
| van Steensel FJA, Zegers VM, Bogels SM. Predictors of treatment effectiveness for youth with ASD and comorbid anxiety disorders: It all depends on the family? J Autism Dev Disord [Internet]. 2017;47(3):636–45. Available from: http://ovidsp.ovid.com/ovidweb.cgi?T=JS&PAGE=reference&D=psyc14&NEWS=N&AN=2016-61739-001 | Wrong intervention |
| Muskett A, Radtke SR, Ollendick T. A pilot study of one-session treatment for specific phobias in children with ASD traits. J Child Fam Stud [Internet]. 2020;29(4):1021–8. Available from: http://ovidsp.ovid.com/ovidweb.cgi?T=JS&PAGE=reference&D=psyc16&NEWS=N&AN=2019-61857-001 | Wrong intervention |
| Ritschel LA, Guy L, Maddox BB. A pilot study of dialectical behaviour therapy skills training for autistic adults. Behavioural and Cognitive Psychotherapy [Internet]. 2022;50(2):187–202. Available from: http://ovidsp.ovid.com/ovidweb.cgi?T=JS&PAGE=reference&D=psyc18&NEWS=N&AN=2021-88960-001 | Wrong intervention |
| Hill TL, Gray SAO, Baker CN, Boggs K, Carey E, Johnson C, et al. A pilot study examining the effectiveness of the PEERS program on social skills and anxiety in adolescents with autism spectrum disorder. J Dev Phys Disabil [Internet]. 2017;29(5):797–808. Available from: http://ovidsp.ovid.com/ovidweb.cgi?T=JS&PAGE=reference&D=psyc14&NEWS=N&AN=2017-26604-001 | Wrong intervention |
| Russo-Ponsaran NM, Evans-Smith B, Johnson JK, McKown C. A pilot study assessing the feasibility of a facial emotion training paradigm for school-age children with autism spectrum disorders. J Ment Health Res Intellect Disabil [Internet]. 2014;7(2):169–90. Available from: http://ovidsp.ovid.com/ovidweb.cgi?T=JS&PAGE=reference&D=psyc11&NEWS=N&AN=2014-15675-005 | Wrong intervention |
| Chen RY, Feltes JR, Tzeng WS, Lu ZY, Pan M, Zhao N, et al. Phone-Based Interventions in Adolescent Psychiatry: A Perspective and Proof of Concept Pilot Study With a Focus on Depression and Autism. JMIR Res Protoc [Internet]. 2017;6(6):e114. Available from: http://ovidsp.ovid.com/ovidweb.cgi?T=JS&PAGE=reference&D=pmnm4&NEWS=N&AN=28623183 | Wrong intervention |
| Wright H, Hall S, Hames A, Hardiman J, Mills R, Mills D. Pet dogs improve family functioning and reduce anxiety in children with Autism Spectrum Disorder. Anthrozoos [Internet]. 2015;28(4):611–24. Available from: http://ovidsp.ovid.com/ovidweb.cgi?T=JS&PAGE=reference&D=psyc12&NEWS=N&AN=2016-06600-006 | Wrong intervention |
| Zheng S, Taylor JL, Adams R, Pezzimenti F, Bishop SL. Perceived helpfulness of depression treatments among young adults with autism. Autism Research [Internet]. 2021;14(7):1522–8. Available from: http://ovidsp.ovid.com/ovidweb.cgi?T=JS&PAGE=reference&D=psyc18&NEWS=N&AN=2021-37277-001 | Wrong intervention |
| Stadnick NA, Drahota A, Brookman-Frazee L. Parent perspectives of an evidence-based intervention for children with autism served in community mental health clinics. J Child Fam Stud [Internet]. 2013;22(3):414–22. Available from: http://ovidsp.ovid.com/ovidweb.cgi?T=JS&PAGE=reference&D=psyc10&NEWS=N&AN=2013-10414-011 | Wrong intervention |
| Adams D, Young K, Simpson K, Keen D. Parent descriptions of the presentation and management of anxiousness in children on the autism spectrum. Autism [Internet]. 2019;23(4):980–92. Available from: http://ovidsp.ovid.com/ovidweb.cgi?T=JS&PAGE=reference&D=psyc16&NEWS=N&AN=2019-29502-016 | Wrong intervention |
| Lipinski S, Blanke ES, Suenkel U, Dziobek I. Outpatient psychotherapy for adults with high-functioning Autism Spectrum Condition: Utilization, treatment satisfaction, and preferred modifications. J Autism Dev Disord [Internet]. 2019;49(3):1154–68. Available from: http://ovidsp.ovid.com/ovidweb.cgi?T=JS&PAGE=reference&D=psyc16&NEWS=N&AN=2018-57493-001 | Wrong intervention |
| Malone RP, Cater J, Sheikh RM, Choudhury MS, Delaney MA. Olanzapine versus haloperidol in children with autistic disorder: An open pilot study. J Am Acad Child Adolesc Psychiatry [Internet]. 2001;40(8):887–93. Available from: http://ovidsp.ovid.com/ovidweb.cgi?T=JS&PAGE=reference&D=psyc3&NEWS=N&AN=2001-01750-003 | Wrong intervention |
| Charman T, Palmer M, Stringer D, Hallett V, Mueller J, Romeo R, et al. A novel group parenting intervention for emotional and behavioral difficulties in young autistic children: Autism Spectrum Treatment and Resilience (ASTAR): A randomized controlled trial. J Am Acad Child Adolesc Psychiatry [Internet]. 2021;60(11):1404–18. Available from: http://ovidsp.ovid.com/ovidweb.cgi?T=JS&PAGE=reference&D=psyc18&NEWS=N&AN=2021-61774-001 | Wrong intervention |
| Posey DJ, Guenin KD, Kohn AE, Swiezy NB, McDougle CJ. A naturalistic open-label study of mirtazapine in autistic and other pervasive developmental disorders. J Child Adolesc Psychopharmacol [Internet]. 2001;11(3):267–77. Available from: http://ovidsp.ovid.com/ovidweb.cgi?T=JS&PAGE=reference&D=psyc3&NEWS=N&AN=2001-05060-005 | Wrong intervention |
| de Bruin EI, Blom R, Smit FMA, van Steensel FJA, Bogels SM. MYmind: Mindfulness training for Youngsters with autism spectrum disorders and their parents. Autism [Internet]. 2015;19(8):906–14. Available from: http://ovidsp.ovid.com/ovidweb.cgi?T=JS&PAGE=reference&D=psyc12&NEWS=N&AN=2015-47118-003 | Wrong intervention |
| Salem-Guirgis S, ra, Albaum C, Tablon P, Riosa PB, Nicholas DB, et al. MYmind: A concurrent group-based mindfulness intervention for youth with autism and their parents. Mindfulness (N Y) [Internet]. 2019;10(9):1730–43. Available from: http://ovidsp.ovid.com/ovidweb.cgi?T=JS&PAGE=reference&D=psyc16&NEWS=N&AN=2019-10706-001 | Wrong intervention |
| Porter S, McConnell T, McLaughlin K, Lynn F, Cardwell C, Braiden HJ, et al. Music therapy for children and adolescents with behavioural and emotional problems: a randomised controlled trial. J Child Psychol Psychiatry [Internet]. 2017;58(5):586–94. Available from: http://ovidsp.ovid.com/ovidweb.cgi?T=JS&PAGE=reference&D=med14&NEWS=N&AN=27786359 | Wrong intervention |
| Charlot L, Deutsch CK, Albert A, Hunt A, Connor DF, McIlvane William J. Jr. Mood and anxiety symptoms in psychiatric inpatients with autism spectrum disorder and depression. J Ment Health Res Intellect Disabil [Internet]. 2008;1(4):238–53. Available from: http://ovidsp.ovid.com/ovidweb.cgi?T=JS&PAGE=reference&D=psyc6&NEWS=N&AN=2009-06298-002 | Wrong intervention |
| Crowell C, Sayis B, Benitez JP, Pares N. Mixed reality, full-body interactive experience to encourage social initiation for autism: Comparison with a control nondigital intervention. Cyberpsychol Behav Soc Netw [Internet]. 2020;23(1):5–9. Available from: http://ovidsp.ovid.com/ovidweb.cgi?T=JS&PAGE=reference&D=psyc17&NEWS=N&AN=2019-64597-001 | Wrong intervention |
| Ridderinkhof A, de Bruin EI, Blom R, Singh NN, Bogels SM. Mindfulness-based program for autism spectrum disorder: A qualitative study of the experiences of children and parents. Mindfulness (N Y) [Internet]. 2019;10(9):1936–51. Available from: http://ovidsp.ovid.com/ovidweb.cgi?T=JS&PAGE=reference&D=psyc16&NEWS=N&AN=2019-41630-001 | Wrong intervention |
| Kramer K, Vetter A, Schultz-Venrath U, Vogeley K, Reul S. Mentalization-Based Treatment in Groups for Adults With Autism Spectrum Disorder. Front Psychol [Internet]. 2021;12:708557. Available from: http://ovidsp.ovid.com/ovidweb.cgi?T=JS&PAGE=reference&D=pmnm5&NEWS=N&AN=34456821 | Wrong intervention |
| Koudys J, Perry A, Ho HSW, Charles M. Mental health status of youth diagnosed with ASD who received early intensive behavioral intervention as young children. Child Fam Behav Ther [Internet]. 2021;43(2):103–13. Available from: http://ovidsp.ovid.com/ovidweb.cgi?T=JS&PAGE=reference&D=psyc18&NEWS=N&AN=2021-31486-001 | Wrong intervention |
| Ryan S, Lai J, Weiss JA. Mental health service use among youth with autism spectrum disorder: A comparison of two age groups. Journal of Developmental and Behavioral Pediatrics [Internet]. 2018;39(1):20–7. Available from: http://ovidsp.ovid.com/ovidweb.cgi?T=JS&PAGE=reference&D=psyc15&NEWS=N&AN=2018-50130-003 | Wrong intervention |
| Song W, Salzer MS, Nonnemacher SL, Shea L. Lifespan service receipt and unmet needs among individuals on the autism spectrum. Administration and Policy in Mental Health and Mental Health Services Research [Internet]. 2022;No-Specified. Available from: http://ovidsp.ovid.com/ovidweb.cgi?T=JS&PAGE=reference&D=psyc18&NEWS=N&AN=2022-48585-001 | Wrong intervention |
| Afsharnejad B, Falkmer M, Black MH, Alach T, Lenhard F, Fridell A, et al. KONTAKT R social skills group training for Australian adolescents with autism spectrum disorder: a randomized controlled trial. Eur Child Adolesc Psychiatry [Internet]. 2021; Available from: http://ovidsp.ovid.com/ovidweb.cgi?T=JS&PAGE=reference&D=medp&NEWS=N&AN=34052908 | Wrong intervention |
| Parker KJ, Oztan O, Libove RA, Sumiyoshi RD, Jackson LP, Karhson DS, et al. Intranasal oxytocin treatment for social deficits and biomarkers of response in children with autism. PNAS Proceedings of the National Academy of Sciences of the United States of America [Internet]. 2017;114(30):8119–24. Available from: http://ovidsp.ovid.com/ovidweb.cgi?T=JS&PAGE=reference&D=psyc14&NEWS=N&AN=2017-32743-022 | Wrong intervention |
| Taylor BJ, S, ers KB, Kyle M, Pedersen KA, Veenstra-V, et al. Inpatient psychiatric treatment of serious behavioral problems in children with Autism Spectrum Disorder (ASD): Specialized versus general inpatient units. J Autism Dev Disord [Internet]. 2019;49(3):1242–9. Available from: http://ovidsp.ovid.com/ovidweb.cgi?T=JS&PAGE=reference&D=psyc16&NEWS=N&AN=2018-60386-001 | Wrong intervention |
| Durnik M, Dougherty JM, Andersson T, Persson B, Bjorevall GB, Emilsson B. Influence of the TEACCH program in Sweden. Special Issue: International priorities for developing autism services via the TEACCH Model-1 [Internet]. 2000;29(1):72–87. Available from: http://ovidsp.ovid.com/ovidweb.cgi?T=JS&PAGE=reference&D=psyc3&NEWS=N&AN=2000-12075-005 | Wrong intervention |
| Bryson SA, Ostmeyer KF. Increasing the effectiveness of community mental health center social skills groups for children with autism spectrum disorder: A training and consultation example. Administration and Policy in Mental Health and Mental Health Services Research [Internet]. 2014;41(6):808–21. Available from: http://ovidsp.ovid.com/ovidweb.cgi?T=JS&PAGE=reference&D=psyc11&NEWS=N&AN=2014-00253-001 | Wrong intervention |
| Guan K, Lau AS, Zhang A, Chlebowski C, Haine-Schlagel R, Brookman-Frazee L. In-Session Caregiver Behaviors during Evidence-Based Intervention Delivery for Children with ASD in Community Mental Health Services. Evid Based Pract Child Adolesc Ment Health [Internet]. 2019;4(1):55–71. Available from: http://ovidsp.ovid.com/ovidweb.cgi?T=JS&PAGE=reference&D=pmnm4&NEWS=N&AN=31263772 | Wrong intervention |
| Coleman-Fountain E, Buckley C, Beresford B. Improving mental health in autistic young adults: a qualitative study exploring help-seeking barriers in UK primary care. Br J Gen Pract [Internet]. 2020;70(694):e356–63. Available from: http://ovidsp.ovid.com/ovidweb.cgi?T=JS&PAGE=reference&D=med17&NEWS=N&AN=32312761 | Wrong intervention |
| Roughan LA, Parker JR, Mercer L. Improving interventions for parents of children and young people with autism spectrum disorder (ASD) in CAMHS. BMJ Open Qual [Internet]. 2019;8(2):e000261. Available from: http://ovidsp.ovid.com/ovidweb.cgi?T=JS&PAGE=reference&D=med16&NEWS=N&AN=31206044 | Wrong intervention |
| Uljarevic M, Richdale A, a L., McConachie H, Hedley D, Cai RY, et al. The hospital anxiety and depression scale: Factor structure and psychometric properties in older adolescents and young adults with autism spectrum disorder. Autism Research [Internet]. 2018;11(2):258–69. Available from: http://ovidsp.ovid.com/ovidweb.cgi?T=JS&PAGE=reference&D=psyc15&NEWS=N&AN=2017-41965-001 | Wrong intervention |
| Cidav Z, Marcus SC, M, ell DS. Home- and community-based waivers for children with autism: Effects on service use and costs. Intellect Dev Disabil [Internet]. 2014;52(4):239–48. Available from: http://ovidsp.ovid.com/ovidweb.cgi?T=JS&PAGE=reference&D=psyc11&NEWS=N&AN=2014-33058-001 | Wrong intervention |
| Ahmedani BK, Hock RM. Health care access and treatment for children with co-morbid autism and psychiatric conditions. Social Psychiatry and Psychiatric Epidemiology: The International Journal for Research in Social and Genetic Epidemiology and Mental Health Services [Internet]. 2012;47(11):1807–14. Available from: http://ovidsp.ovid.com/ovidweb.cgi?T=JS&PAGE=reference&D=psyc9&NEWS=N&AN=2012-28433-011 | Wrong intervention |
| Okuda T, Asano K, Numata N, Hirano Y, Yamamoto T, Tanaka M, et al. Feasibility of cognitive remediation therapy for adults with autism spectrum disorders: A single-group pilot study. Neuropsychiatr Dis Treat [Internet]. 2017;13. Available from: http://ovidsp.ovid.com/ovidweb.cgi?T=JS&PAGE=reference&D=psyc14&NEWS=N&AN=2017-35970-001 | Wrong intervention |
| Carlisle GK, Johnson RA, Wang Z, Bibbo J, Cheak-Zamora N, Lyons LA. Exploratory Study of Cat Adoption in Families of Children with Autism: Impact on Children’s Social Skills and Anxiety. J Pediatr Nurs [Internet]. 2021;58:28–35. Available from: http://ovidsp.ovid.com/ovidweb.cgi?T=JS&PAGE=reference&D=med18&NEWS=N&AN=33290937 | Wrong intervention |
| Catchpole R, Young A, Baer S, Salih T. Examining a novel, parent child interaction therapy-informed, behavioral treatment of selective mutism. J Anxiety Disord [Internet]. 2019;66. Available from: http://ovidsp.ovid.com/ovidweb.cgi?T=JS&PAGE=reference&D=psyc16&NEWS=N&AN=2019-48478-001 | Wrong intervention |
| Carminati GG, Gerber F, Kempf-Constantin N, Baud O. Evolution of adults with autism and profound intellectual disabilities living within a structured residential programme: A 21-month longitudinal study. Schweizer Archiv fur Neurologie und Psychiatrie [Internet]. 2007;158(5):233–41. Available from: http://ovidsp.ovid.com/ovidweb.cgi?T=JS&PAGE=reference&D=psyc5&NEWS=N&AN=2007-12671-004 | Wrong intervention |
| Silva EA da J, Medeiros W, ersonia Moreira Brito, Santos JPM Dos, Sousa JMM de, Costa FB da, et al. Evaluation of the efficacy and safety of cannabidiol-rich cannabis extract in children with autism spectrum disorder: randomized, double-blind and controlled placebo clinical trial. Trends Psychiatry Psychother [Internet]. 2022;44. Available from: http://ovidsp.ovid.com/ovidweb.cgi?T=JS&PAGE=reference&D=medp&NEWS=N&AN=35617670 | Wrong intervention |
| Cornwall PL, Simpson S, Gibbs C, Morfee V. Evaluation of radically open dialectical behaviour therapy in an adult community mental health team: Effectiveness in people with autism spectrum disorders. BJPsych Bull [Internet]. 2021;45(3):146–53. Available from: http://ovidsp.ovid.com/ovidweb.cgi?T=JS&PAGE=reference&D=psyc18&NEWS=N&AN=2021-51048-005 | Wrong intervention |
| Frolli A, ro, Ricci MC, Di Carmine F, Orefice A, Saviano E, et al. Emotional Rational Education Training Associated with Mindfulness for Managing Anxiety within Adolescents Affected by High-Functioning Autism: A Descriptive Study. Behavioral sciences (Basel, Switzerland) [Internet]. 2021;11(11). Available from: http://ovidsp.ovid.com/ovidweb.cgi?T=JS&PAGE=reference&D=pmnm5&NEWS=N&AN=34821617 | Wrong intervention |
| Roberts-Collins C, Mahoney-Davies G, Russell A, Booth A, Loades M. Emotion awareness and cognitive behavioural therapy in young people with autism spectrum disorder. Autism [Internet]. 2018;22(7):837–44. Available from: http://ovidsp.ovid.com/ovidweb.cgi?T=JS&PAGE=reference&D=psyc15&NEWS=N&AN=2018-50457-008 | Wrong intervention |
| Lind T, Lau AS, Gomez C, Rodriguez A, Guan K, Chlebowski C, et al. Emergent life events in the delivery of a caregiver-mediated evidence-based intervention for children with autism spectrum disorder in publicly funded mental health services. Autism [Internet]. 2020;24(5):1286–99. Available from: http://ovidsp.ovid.com/ovidweb.cgi?T=JS&PAGE=reference&D=psyc18&NEWS=N&AN=2020-45699-020 | Wrong intervention |
| Zamzow RM, Ferguson BJ, Stichter JP, Porges EC, Ragsdale A, ra S., et al. Effects of propranolol on conversational reciprocity in autism spectrum disorder: A pilot, double-blind, single-dose psychopharmacological challenge study. Psychopharmacology (Berl) [Internet]. 2016;233(7):1171–8. Available from: http://ovidsp.ovid.com/ovidweb.cgi?T=JS&PAGE=reference&D=psyc13&NEWS=N&AN=2016-02456-001 | Wrong intervention |
| Guo J, Li D. Effects of Image-Sandplay Therapy on the Mental Health and Subjective Well-Being of Children with Autism. Iran J Public Health [Internet]. 2021;50(10):2046–54. Available from: http://ovidsp.ovid.com/ovidweb.cgi?T=JS&PAGE=reference&D=pmnm&NEWS=N&AN=35223572 | Wrong intervention |
| Kaur M, Eigsti IM, Bhat A. Effects of a creative yoga intervention on the joint attention and social communication skills, as well as affective states of children with Autism Spectrum Disorder. Res Autism Spectr Disord [Internet]. 2021;88. Available from: http://ovidsp.ovid.com/ovidweb.cgi?T=JS&PAGE=reference&D=pmnm&NEWS=N&AN=34552663 | Wrong intervention |
| Chien YL, Tsai WC, Chen WH, Yang CL, Gau SSF, Soong WT, et al. Effectiveness, durability, and clinical correlates of the peers social skills intervention in young adults with autism spectrum disorder: The first evidence outside north america. Psychol Med [Internet]. 2021;No-Specified. Available from: http://ovidsp.ovid.com/ovidweb.cgi?T=JS&PAGE=reference&D=psyc18&NEWS=N&AN=2021-64660-001 | Wrong intervention |
| Brookman-Frazee L, Roesch S, Chlebowski C, Baker-Ericzen M, Ganger W. Effectiveness of training therapists to deliver an individualized mental health intervention for children with ASD in publicly funded mental health services: A cluster randomized clinical trial. JAMA Psychiatry [Internet]. 2019;76(6):574–83. Available from: http://ovidsp.ovid.com/ovidweb.cgi?T=JS&PAGE=reference&D=psyc16&NEWS=N&AN=2019-75734-003 | Wrong intervention |
| Brookman-Frazee L, Chlebowski C, Villodas M, Garl, Ann, McPherson J, et al. The effectiveness of training community mental health therapists in an evidence-based intervention for ASD: Findings from a hybrid effectiveness-implementation trial in outpatient and school-based mental health services. Autism [Internet]. 2022;26(3):678–89. Available from: http://ovidsp.ovid.com/ovidweb.cgi?T=JS&PAGE=reference&D=med20&NEWS=N&AN=34983251 | Wrong intervention |
| Cashin A, Browne G, Bradbury J, Mulder A. The effectiveness of narrative therapy with young people with autism. Journal of Child and Adolescent Psychiatric Nursing [Internet]. 2013;26(1):32–41. Available from: http://ovidsp.ovid.com/ovidweb.cgi?T=JS&PAGE=reference&D=psyc10&NEWS=N&AN=2013-02822-006 | Wrong intervention |
| Buchsbaum MS, Holl, er E, Haznedar MM, Tang C, Spiegel-Cohen J, et al. Effect of fluoxetine on regional cerebral metabolism in autistic spectrum disorders: A pilot study. International Journal of Neuropsychopharmacology [Internet]. 2001;4(2):119–25. Available from: http://ovidsp.ovid.com/ovidweb.cgi?T=JS&PAGE=reference&D=psyc3&NEWS=N&AN=2001-01759-003 | Wrong intervention |
| Golubchik P, Rapaport M, Weizman A. The effect of methylphenidate on anxiety and depression symptoms in patients with Asperger syndrome and comorbid attention deficit/hyperactivity disorder. Int Clin Psychopharmacol [Internet]. 2017;32(5):289–93. Available from: http://ovidsp.ovid.com/ovidweb.cgi?T=JS&PAGE=reference&D=psyc14&NEWS=N&AN=2017-34210-007 | Wrong intervention |
| Cohen SA, Fitzgerald BJ, Khan SRF, Khan A. The Effect of a Switch to Ziprasidone in an Adult Population With Autistic Disorder: Chart Review of Naturalistic, Open-Label Treatment. J Clin Psychiatry [Internet]. 2004;65(1):110–3. Available from: http://ovidsp.ovid.com/ovidweb.cgi?T=JS&PAGE=reference&D=psyc4&NEWS=N&AN=2004-10836-018 | Wrong intervention |
| Mills W, Kondakis N, Orr R, Warburton M, Milne N. Does Hydrotherapy Impact Behaviours Related to Mental Health and Well-Being for Children with Autism Spectrum Disorder? A Randomised Crossover-Controlled Pilot Trial. Int J Environ Res Public Health [Internet]. 2020;17(2). Available from: http://ovidsp.ovid.com/ovidweb.cgi?T=JS&PAGE=reference&D=med17&NEWS=N&AN=31952323 | Wrong intervention |
| Afsharnejad B, Falkmer M, Black MH, Alach T, Lenhard F, Fridell A, et al. Cross-cultural adaptation to Australia of the KONTAKT© social skills group training program for youth with autism spectrum disorder: A feasibility study. J Autism Dev Disord [Internet]. 2020;50(12):4297–316. Available from: http://ovidsp.ovid.com/ovidweb.cgi?T=JS&PAGE=reference&D=psyc17&NEWS=N&AN=2020-25995-001 | Wrong intervention |
| Brezis RS, Levin A, Oded Y, Zahavi O, Gampel G, Levit-Binnun N. A contemplative biofeedback intervention for adults with autism spectrum disorder: Feasibility of a community-based treatment. Appl Psychophysiol Biofeedback [Internet]. 2021;46(2):141–9. Available from: http://ovidsp.ovid.com/ovidweb.cgi?T=JS&PAGE=reference&D=psyc18&NEWS=N&AN=2021-13125-001 | Wrong intervention |
| Conner CM, Golt J, Righi G, Shaffer R, Siegel M, Mazefsky CA. A comparative study of suicidality and its association with emotion regulation impairment in large ASD and US census-matched samples. J Autism Dev Disord [Internet]. 2020;50(10):3545–60. Available from: http://ovidsp.ovid.com/ovidweb.cgi?T=JS&PAGE=reference&D=psyc17&NEWS=N&AN=2020-03418-001 | Wrong intervention |
| Abbott M, y, Bernard P, Forge J. Communicating a diagnosis of Autism Spectrum Disorder-A qualitative study of parents’ experiences. Clin Child Psychol Psychiatry [Internet]. 2013;18(3):370–82. Available from: http://ovidsp.ovid.com/ovidweb.cgi?T=JS&PAGE=reference&D=psyc10&NEWS=N&AN=2013-23097-005 | Wrong intervention |
| Green J, Leadbitter K, Ellis C, Taylor L, Moore HL, Carruthers S, et al. “Combined social communication therapy at home and in education for young autistic children in England (PACT-G): A parallel, single-blind, randomised controlled trial”: Correction. Lancet Psychiatry [Internet]. 2022;9(5):e21. Available from: http://ovidsp.ovid.com/ovidweb.cgi?T=JS&PAGE=reference&D=psyc18&NEWS=N&AN=2022-55778-021 | Wrong intervention |
| Hurwich-Reiss E, Chlebowski C, Lind T, Martinez K, ra, Best KM, et al. Characterizing therapist delivery of evidence-based intervention strategies in publicly funded mental health services for children with autism spectrum disorder: Differentiating practice patterns in usual care and AIM HI delivery. Autism [Internet]. 2021;25(6):1709–20. Available from: http://ovidsp.ovid.com/ovidweb.cgi?T=JS&PAGE=reference&D=med19&NEWS=N&AN=33779320 | Wrong intervention |
| Schiltz HK, McVey AJ, Dolan BK, Willar KS, Pleiss S, Karst JS, et al. Changes in depressive symptoms among adolescents with ASD completing the PEERS social skills intervention. J Autism Dev Disord [Internet]. 2018;48(3):834–43. Available from: http://ovidsp.ovid.com/ovidweb.cgi?T=JS&PAGE=reference&D=psyc15&NEWS=N&AN=2017-52908-001 | Wrong intervention |
| Corbett BA, Blain SD, Ioannou S, Balser M. Changes in anxiety following a randomized control trial of a theatre-based intervention for youth with autism spectrum disorder. Autism [Internet]. 2017;21(3):333–43. Available from: http://ovidsp.ovid.com/ovidweb.cgi?T=JS&PAGE=reference&D=psyc14&NEWS=N&AN=2017-10652-008 | Wrong intervention |
| van Steensel FJA, Bogels SM. Cbt for anxiety disorders in children with and without autism spectrum disorders. J Consult Clin Psychol [Internet]. 2015;83(3):512–23. Available from: http://ovidsp.ovid.com/ovidweb.cgi?T=JS&PAGE=reference&D=psyc12&NEWS=N&AN=2015-17270-001 | Wrong intervention |
| Bentz M, Pedersen SH, Moslet U. Case series of family-based treatment for restrictive-type eating disorders and comorbid autism: What can we learn? A brief report. Eur Eat Disord Rev [Internet]. 2022; Available from: http://ovidsp.ovid.com/ovidweb.cgi?T=JS&PAGE=reference&D=medp&NEWS=N&AN=35808867 | Wrong intervention |
| Howells K, Sivaratnam C, Lindor E, Hyde C, McGillivray J, Whitehouse A, et al. Can participation in a community organized football program improve social, behavioural functioning and communication in children with autism spectrum disorder? A pilot study. J Autism Dev Disord [Internet]. 2020;50(10):3714–27. Available from: http://ovidsp.ovid.com/ovidweb.cgi?T=JS&PAGE=reference&D=psyc17&NEWS=N&AN=2020-15115-001 | Wrong intervention |
| Kuschner ES, Morton HE, Maddox BB, de Marchena A, Anthony LG, Reaven J. The BUFFET Program: Development of a Cognitive Behavioral Treatment for Selective Eating in Youth with Autism Spectrum Disorder. Clin Child Fam Psychol Rev [Internet]. 2017;20(4):403–21. Available from: http://ovidsp.ovid.com/ovidweb.cgi?T=JS&PAGE=reference&D=med14&NEWS=N&AN=28534237 | Wrong intervention |
| Hampton LH, Roberts MY, Anderson E, Hobson A, a N., Kaat AJ, et al. Brief report: What diagnostic observation can teach us about disruptive behavior in young children with autism. Journal of Developmental and Behavioral Pediatrics [Internet]. 2021;42(1):55–60. Available from: http://ovidsp.ovid.com/ovidweb.cgi?T=JS&PAGE=reference&D=psyc18&NEWS=N&AN=2021-52385-008 | Wrong intervention |
| Stark KH, Barnes JC, Young ND, Gabriels RL. Brief report: Understanding crisis behaviors in hospitalized psychiatric patients with autism spectrum disorder-Iceberg Assessment Interview. J Autism Dev Disord [Internet]. 2015;45(11):3468–74. Available from: http://ovidsp.ovid.com/ovidweb.cgi?T=JS&PAGE=reference&D=psyc12&NEWS=N&AN=2015-40540-001 | Wrong intervention |
| Lipinski S, Boegl K, Blanke ES, Suenkel U, Dziobek I. A blind spot in mental healthcare? Psychotherapists lack education and expertise for the support of adults on the autism spectrum. Autism [Internet]. 2021;13623613211057972. Available from: http://ovidsp.ovid.com/ovidweb.cgi?T=JS&PAGE=reference&D=medp&NEWS=N&AN=34825580 | Wrong intervention |
| Hawkins JR, Weatherby N, Wrye B, Ujcich Ward K. Bergamot Aromatherapy for Medical Office-Induced Anxiety Among Children With an Autism Spectrum Disorder: A Randomized, Controlled, Blinded Clinical Trial. Holist Nurs Pract [Internet]. 2019;33(5):285–94. Available from: https://search.ebscohost.com/login.aspx?direct=true&AuthType=ip,shib&db=jlh&AN=138742113&site=ehost-live&scope=site&custid=s8454451 | Wrong intervention |
| Wittling K, Dufur JP, McClain A, Gettis M. Behavioral Coping Plans: One Inter-Professional Team’s Approach to Patient-Centered Care. J Pediatr Nurs [Internet]. 2018;41:135–9. Available from: http://ovidsp.ovid.com/ovidweb.cgi?T=JS&PAGE=reference&D=pmnm4&NEWS=N&AN=29778301 | Wrong intervention |
| Downs JM, Lechler S, Dean H, Sears N, Patel R, Shetty H, et al. The Association Between Comorbid Autism Spectrum Disorders and Antipsychotic Treatment Failure in Early-Onset Psychosis: A Historical Cohort Study Using Electronic Health Records. J Clin Psychiatry [Internet]. 2017;78(9):e1233–41. Available from: http://ovidsp.ovid.com/ovidweb.cgi?T=JS&PAGE=reference&D=med14&NEWS=N&AN=29125721 | Wrong intervention |
| Capriola-Hall NN, Brewe AM, Golt J, White SW. Anxiety and depression reduction as distal outcomes of a college transition readiness program for adults with autism. J Autism Dev Disord [Internet]. 2021;51(1):298–306. Available from: http://ovidsp.ovid.com/ovidweb.cgi?T=JS&PAGE=reference&D=psyc18&NEWS=N&AN=2020-39039-001 | Wrong intervention |
| Byrne G, Ghráda ÁN, O’Mahony T. Parent-Led Cognitive Behavioural Therapy for Children with Autism Spectrum Conditions. A Pilot Study. J Autism Dev Disord [Internet]. 2023 Jan 1 [cited 2023 Oct 13];53(1):263–74. Available from: https://link.springer.com/article/10.1007/s10803-022-05424-2 | Wrong intervention |
| Becker JL, Rogers EC, Burrows B. Anthrozoös A multidisciplinary journal of the interactions of people and animals Animal-assisted Social Skills Training for Children with Autism Spectrum Disorders. 2017 [cited 2023 Oct 13]; Available from: https://www.tandfonline.com/action/journalInformation?journalCode=rfan20 | Wrong intervention |
| Doherty M, Neilson S, O’Sullivan J, Carravallah L, Johnson M, Cullen W, et al. Barriers to healthcare and self-reported adverse outcomes for autistic adults: a cross-sectional study. BMJ Open [Internet]. 2022 Feb 1 [cited 2023 Oct 13];12(2):e056904. Available from: https://bmjopen.bmj.com/content/12/2/e056904 | Wrong intervention |
| Conner CM, White SW. Brief Report: Feasibility and Preliminary Efficacy of Individual Mindfulness Therapy for Adults with Autism Spectrum Disorder. J Autism Dev Disord [Internet]. 2018 Jan 1 [cited 2023 Oct 13];48(1):290–300. Available from: https://link.springer.com/article/10.1007/s10803-017-3312-0 | Wrong setting |
| Pickard K, Meyer A, Reyes N, T, a T, Reaven J. Using evaluative frameworks to examine the implementation outcomes of a cognitive behavioral therapy program for autistic students with anxiety within public school settings. Autism [Internet]. 2022;26(3):640–53. Available from: http://ovidsp.ovid.com/ovidweb.cgi?T=JS&PAGE=reference&D=med20&NEWS=N&AN=34961348    %[20211228// | Wrong setting |
| Jemison Pollard D. The use of music to improve social skills development in children diagnosed with autism. Dissertation Abstracts International Section A: Humanities and Social Sciences [Internet]. 2014;74(7):No-Specified. Available from: http://ovidsp.ovid.com/ovidweb.cgi?T=JS&PAGE=reference&D=psyc11&NEWS=N&AN=2014-99010-363 | Wrong setting |
| Ireri NW, White SW, Mbwayo AW. Treating anxiety and social deficits in children with autism spectrum disorder in two schools in Nairobi, Kenya. J Autism Dev Disord [Internet]. 2019;49(8):3309–15. Available from: http://ovidsp.ovid.com/ovidweb.cgi?T=JS&PAGE=reference&D=psyc16&NEWS=N&AN=2019-27644-001 | Wrong setting |
| Adamson J, Brede J, Babb C, Serpell L, Jones CRG, Fox J, et al. Towards identifying a method of screening for autism amongst women with restrictive eating disorders. European Eating Disorders Review [Internet]. 2022;No-Specified. Available from: http://ovidsp.ovid.com/ovidweb.cgi?T=JS&PAGE=reference&D=psyc18&NEWS=N&AN=2022-80073-001 | Wrong setting |
| Hillier A, Goldstein J, Murphy D, Trietsch R, Keeves J, Mendes E, et al. Supporting university students with autism spectrum disorder. Autism [Internet]. 2018;22(1):20–8. Available from: http://ovidsp.ovid.com/ovidweb.cgi?T=JS&PAGE=reference&D=psyc15&NEWS=N&AN=2018-03902-003 | Wrong setting |
| Ratcliffe B, Wong M, Dossetor D, Hayes S. Teaching social-emotional skills to school-aged children with Autism Spectrum Disorder: A treatment versus control trial in 41 mainstream schools. Res Autism Spectr Disord [Internet]. 2014;8(12):1722–33. Available from: http://ovidsp.ovid.com/ovidweb.cgi?T=JS&PAGE=reference&D=psyc11&NEWS=N&AN=2014-45692-012 | Wrong setting |
| Shochet IM, Saggers BR, Carrington SB, Orr JA, Wurfl AM, Duncan BM. A strength-focused parenting intervention may be a valuable augmentation to a depression prevention focus for adolescents with autism. J Autism Dev Disord [Internet]. 2019;49(5):2080–100. Available from: http://ovidsp.ovid.com/ovidweb.cgi?T=JS&PAGE=reference&D=psyc16&NEWS=N&AN=2019-04623-001 | Wrong setting |
| Hatfield MK, Ashcroft E, Maguire S, Kershaw L, Ciccarelli M. “stop and just breathe for a minute”: Perspectives of children on the autism spectrum and their caregivers on a mindfulness group. J Autism Dev Disord [Internet]. 2022;No-Specified. Available from: http://ovidsp.ovid.com/ovidweb.cgi?T=JS&PAGE=reference&D=psyc18&NEWS=N&AN=2022-78812-001 | Wrong setting |
| Beck KB, Northrup JB, Breitenfeldt KE, Porton S, Day TN, MacKenzie KT, et al. Stakeholder informed development of the Emotion Awareness and Skills Enhancement team-based program (EASE-Teams). Autism [Internet]. 2022;26(3):586–600. Available from: http://ovidsp.ovid.com/ovidweb.cgi?T=JS&PAGE=reference&D=med20&NEWS=N&AN=34903083    %[20211214// | Wrong setting |
| Kerr-Gaffney J, Harrison A, Tchanturia K. The social responsiveness scale is an efficient screening tool for autism spectrum disorder traits in adults with anorexia nervosa. European Eating Disorders Review [Internet]. 2020;No-Specified. Available from: http://ovidsp.ovid.com/ovidweb.cgi?T=JS&PAGE=reference&D=psyc17&NEWS=N&AN=2020-24910-001 | Wrong setting |
| Powell L. A school-based intervention involving massage and yoga for disadvantaged male adolescents: An exploratory study on a self-discovery program. The human pursuit of well-being: A cultural approach [Internet]. 2011;257–67. Available from: http://ovidsp.ovid.com/ovidweb.cgi?T=JS&PAGE=reference&D=psyc8&NEWS=N&AN=2011-15071-023 | Wrong setting |
| Shochet IM, Saggers BR, Carrington SB, Orr JA, Wurfl AM, Kelly RL, et al. A school-based approach to building resilience and mental health among adolescents on the autism spectrum: A longitudinal mixed methods study. School Mental Health: A Multidisciplinary Research and Practice Journal [Internet]. 2022;No-Specified. Available from: http://ovidsp.ovid.com/ovidweb.cgi?T=JS&PAGE=reference&D=psyc18&NEWS=N&AN=2022-26228-001 | Wrong setting |
| Weiss JA, Albaum C, Chan V, Maughan A, Tablon P, Black K, et al. A randomized waitlist‐controlled trial of cognitive behavior therapy to improve emotion regulation in children with autism. Journal of Child Psychology & Psychiatry [Internet]. 2018;59(11):1180–91. Available from: https://search.ebscohost.com/login.aspx?direct=true&AuthType=ip,shib&db=jlh&AN=132481149&site=ehost-live&scope=site&custid=s8454451 | Wrong setting |
| Clarke C, Hill V, Charman T. School based cognitive behavioural therapy targeting anxiety in children with autistic spectrum disorder: A quasi-experimental randomised controlled trail incorporating a mixed methods approach. J Autism Dev Disord [Internet]. 2017;47(12):3883–95. Available from: http://ovidsp.ovid.com/ovidweb.cgi?T=JS&PAGE=reference&D=psyc14&NEWS=N&AN=2016-22243-001 | Wrong setting |
| Braden BB, Pagni BA, Monahan L, Walsh MJM, Dixon M V, Delaney S, et al. Quality of life in adults with autism spectrum disorder: Influence of age, sex, and a controlled, randomized mindfulness-based stress reduction pilot intervention. Qual Life Res [Internet]. 2021;No-Specified. Available from: http://ovidsp.ovid.com/ovidweb.cgi?T=JS&PAGE=reference&D=psyc18&NEWS=N&AN=2021-97874-001 | Wrong setting |
| Vause T, Neil N, Jaksic H, Jackiewicz G, Feldman M. Preliminary randomized trial of function-based cognitive-behavioral therapy to treat obsessive compulsive behavior in children with autism spectrum disorder. Focus Autism Other Dev Disabl [Internet]. 2017;32(3):218–28. Available from: http://ovidsp.ovid.com/ovidweb.cgi?T=JS&PAGE=reference&D=psyc14&NEWS=N&AN=2017-36893-006 | Wrong setting |
| Hillier A, Greher G, Poto N, Dougherty M. Positive outcomes following participation in a music intervention for adolescents and young adults on the autism spectrum. Psychol Music [Internet]. 2012;40(2):201–15. Available from: http://ovidsp.ovid.com/ovidweb.cgi?T=JS&PAGE=reference&D=psyc9&NEWS=N&AN=2012-06623-005 | Wrong setting |
| Mackay BA, Shochet IM, Orr JA. A pilot randomised controlled trial of a school-based resilience intervention to prevent depressive symptoms for young adolescents with autism spectrum disorder: A mixed methods analysis. J Autism Dev Disord [Internet]. 2017;47(11):3458–78. Available from: http://ovidsp.ovid.com/ovidweb.cgi?T=JS&PAGE=reference&D=psyc14&NEWS=N&AN=2017-33775-001 | Wrong setting |
| Yang YJ, Chung KM. Pilot randomized control trial of an app-based cbt program for reducing anxiety in individuals with asd without intellectual disability. J Autism Dev Disord [Internet]. 2022;No-Specified. Available from: http://ovidsp.ovid.com/ovidweb.cgi?T=JS&PAGE=reference&D=psyc18&NEWS=N&AN=2022-72378-001 | Wrong setting |
| Ioannou S, Key A, ra P, Muscatello RA, Klemencic M, Corbett BA. Peer actors and theater techniques play pivotal roles in improving social play and anxiety for children with autism. Front Psychol [Internet]. 2020;11. Available from: http://ovidsp.ovid.com/ovidweb.cgi?T=JS&PAGE=reference&D=psyc18&NEWS=N&AN=2020-42786-001 | Wrong setting |
| Pagni BA, Walsh MJM, Foldes E, Sebren A, Dixon M V, Guerithault N, et al. The neural correlates of mindfulness-induced depression reduction in adults with autism spectrum disorder: A pilot study. J Neurosci Res [Internet]. 2020;98(6):1150–61. Available from: http://ovidsp.ovid.com/ovidweb.cgi?T=JS&PAGE=reference&D=med17&NEWS=N&AN=32090389    %[20200223// | Wrong setting |
| Kolmen BK, Feldman HM, H, en BL, Janosky JE. Naltrexone in young autistic children: A double-blind, placebo-controlled crossover study. J Am Acad Child Adolesc Psychiatry [Internet]. 1995;34(2):223–31. Available from: http://ovidsp.ovid.com/ovidweb.cgi?T=JS&PAGE=reference&D=psyc3&NEWS=N&AN=1995-34200-001 | Wrong setting |
| Locke J, Beidas RS, Marcus S, Stahmer A, Aarons GA, Lyon AR, et al. A mixed methods study of individual and organizational factors that affect implementation of interventions for children with autism in public schools. Implement Sci [Internet]. 2016;11(1):135. Available from: http://ovidsp.ovid.com/ovidweb.cgi?T=JS&PAGE=reference&D=med13&NEWS=N&AN=27724933    %[20161010// | Wrong setting |
| Stockall N, Blackwell W. Mindfulness training: Reducing anxiety in students with autism spectrum disorder. Early Child Educ J [Internet]. 2022;50(1):1–9. Available from: http://ovidsp.ovid.com/ovidweb.cgi?T=JS&PAGE=reference&D=psyc17&NEWS=N&AN=2020-85987-001 | Wrong setting |
| Choudhary S, Begum R. Management of Anxiety Related Disorder in Children with High Functioning Autism Spectrum Disorder Using Cognitive Behavioural Therapy. Indian J Physiother Occup Ther [Internet]. 2018;12(4):61–6. Available from: https://search.ebscohost.com/login.aspx?direct=true&AuthType=ip,shib&db=jlh&AN=132964889&site=ehost-live&scope=site&custid=s8454451 | Wrong setting |
| Ratcliffe B, Wong M, Dossetor D, Hayes S. Improving emotional competence in children with autism spectrum disorder and mild intellectual disability in schools: A preliminary treatment versus waitlist study. Behaviour Change [Internet]. 2019;36(4):216–32. Available from: http://ovidsp.ovid.com/ovidweb.cgi?T=JS&PAGE=reference&D=psyc16&NEWS=N&AN=2019-74460-003 | Wrong setting |
| Conner CM, White SW, Beck KB, Golt J, Smith IC, Mazefsky CA. Improving emotion regulation ability in autism: The Emotional Awareness and Skills Enhancement (EASE) program. Autism [Internet]. 2019;23(5):1273–87. Available from: http://ovidsp.ovid.com/ovidweb.cgi?T=JS&PAGE=reference&D=psyc16&NEWS=N&AN=2019-35473-018 | Wrong setting |
| Nazni P, Wesely EG, Nishadevi V. Impact of casein and gluten free dietary intervention on selected autistic children. Iran J Pediatr [Internet]. 2008;18(3):244–50. Available from: https://search.ebscohost.com/login.aspx?direct=true&AuthType=ip,shib&db=jlh&AN=105712273&site=ehost-live&scope=site&custid=s8454451 | Wrong setting |
| Blakeley-Smith A, Meyer AT, Boles RE, Reaven J. Group cognitive behavioural treatment for anxiety in autistic adolescents with intellectual disability: A pilot and feasibility study. Journal of Applied Research in Intellectual Disabilities [Internet]. 2021;34(3):777–88. Available from: http://ovidsp.ovid.com/ovidweb.cgi?T=JS&PAGE=reference&D=psyc18&NEWS=N&AN=2021-07136-001 | Wrong setting |
| Drmic IE, Aljunied M, Reaven J. Feasibility, acceptability and preliminary treatment outcomes in a school-based CBT intervention program for adolescents with ASD and anxiety in Singapore. J Autism Dev Disord [Internet]. 2017;47(12):3909–29. Available from: http://ovidsp.ovid.com/ovidweb.cgi?T=JS&PAGE=reference&D=psyc14&NEWS=N&AN=2017-02853-001 | Wrong setting |
| Davidson D, DiClemente CM, Hilvert E. Experiences and insights of college students with autism spectrum disorder: An exploratory assessment to inform interventions. Journal of American College Health [Internet]. 2021;No-Specified. Available from: http://ovidsp.ovid.com/ovidweb.cgi?T=JS&PAGE=reference&D=psyc18&NEWS=N&AN=2021-31473-001 | Wrong setting |
| Nguyen J, Cardy RE, Anagnostou E, Brian J, Kushki A. Examining the effect of a wearable, anxiety detection technology on improving the awareness of anxiety signs in autism spectrum disorder: a pilot randomized controlled trial. Mol Autism [Internet]. 2021;12(1):72. Available from: http://ovidsp.ovid.com/ovidweb.cgi?T=JS&PAGE=reference&D=med20&NEWS=N&AN=34775994    %[20211114// | Wrong setting |
| Tanksale R, Sofronoff K, Sheffield J, Gilmour J. Evaluating the effects of a yoga-based program integrated with third-wave cognitive behavioral therapy components on self-regulation in children on the autism spectrum: A pilot randomized controlled trial. Autism [Internet]. 2021;25(4):995–1008. Available from: http://ovidsp.ovid.com/ovidweb.cgi?T=JS&PAGE=reference&D=med18&NEWS=N&AN=33238718    %[20201125// | Wrong setting |
| Luxford S, Hadwin JA, Kovshoff H. Evaluating the effectiveness of a school-based cognitive behavioural therapy intervention for anxiety in adolescents diagnosed with Autism Spectrum Disorder. J Autism Dev Disord [Internet]. 2017;47(12):3896–908. Available from: http://ovidsp.ovid.com/ovidweb.cgi?T=JS&PAGE=reference&D=psyc14&NEWS=N&AN=2016-36183-001 | Wrong setting |
| Chadwick Z, Edmondson A, a, McDonald S. Engaging with animal-assisted interventions (AAIs): Exploring the experiences of young people with ASD/ADHD diagnoses. Support for Learning [Internet]. 2022;37(1):44–61. Available from: http://ovidsp.ovid.com/ovidweb.cgi?T=JS&PAGE=reference&D=psyc18&NEWS=N&AN=2022-35292-004 | Wrong setting |
| Cho A, Wood JJ, Ferrer E, Rosenau K, Storch EA, Kendall PC. Empirically-identified subgroups of children with autism spectrum disorder and their response to two types of cognitive behavioral therapy. Dev Psychopathol [Internet]. 2021;No-Specified. Available from: http://ovidsp.ovid.com/ovidweb.cgi?T=JS&PAGE=reference&D=psyc18&NEWS=N&AN=2022-13020-001 | Wrong setting |
| Breeman LD, van Lier PAC, Wubbels T, Verhulst FC, van der Ende J, Maras A, et al. Effects of the good behavior game on the behavioral, emotional, and social problems of children with psychiatric disorders in special education settings. J Posit Behav Interv [Internet]. 2016;18(3):156–67. Available from: http://ovidsp.ovid.com/ovidweb.cgi?T=JS&PAGE=reference&D=psyc13&NEWS=N&AN=2016-29836-004 | Wrong setting |
| Ooi YP, Lam CM, Sung M, Tan WTS, Goh TJ, Fung DSS, et al. Effects of cognitive-behavioural therapy on anxiety for children with high-functioning autistic spectrum disorders. Singapore Med J [Internet]. 2008;49(3):215–20. Available from: http://ovidsp.ovid.com/ovidweb.cgi?T=JS&PAGE=reference&D=med7&NEWS=N&AN=18363003 | Wrong setting |
| Katz J, Knight V, Mercer SH, Skinner SY. Effects of a universal school-based mental health program on the self-concept, coping skills, and perceptions of social support of students with developmental disabilities. J Autism Dev Disord [Internet]. 2020;50(11):4069–84. Available from: http://ovidsp.ovid.com/ovidweb.cgi?T=JS&PAGE=reference&D=psyc17&NEWS=N&AN=2020-25254-001 | Wrong setting |
| Garcia-Gomez A, Risco ML, Rubi JC, Guerrero E, Garcia-Pena IM. Effects of a program of adapted therapeutic horse-riding in a group of autism spectrum disorder children. Electronic Journal of Research in Educational Psychology [Internet]. 2014;12(1):107–28. Available from: http://ovidsp.ovid.com/ovidweb.cgi?T=JS&PAGE=reference&D=psyc11&NEWS=N&AN=2014-19008-005 | Wrong setting |
| Carey M, Sheehan D, Healy S, Knott F, Kinsella S. The Effects of a 16-Week School-Based Exercise Program on Anxiety in Children with Autism Spectrum Disorder. Int J Environ Res Public Health [Internet]. 2022;19(9). Available from: http://ovidsp.ovid.com/ovidweb.cgi?T=JS&PAGE=reference&D=mesx&NEWS=N&AN=35564866    %[20220430// | Wrong setting |
| Ben-Itzchak E, Zachor DA. Dog training intervention improves adaptive social communication skills in young children with autism spectrum disorder: A controlled crossover study. Autism [Internet]. 2021;25(6):1682–93. Available from: http://ovidsp.ovid.com/ovidweb.cgi?T=JS&PAGE=reference&D=med19&NEWS=N&AN=33752452    %[20210322// | Wrong setting |
| Smith E, Constantin A, Johnson H, Brosnan M. Digitally-mediated social stories support children on the autism spectrum adapting to a change in a “real-world” context. J Autism Dev Disord [Internet]. 2021;51(2):514–26. Available from: http://ovidsp.ovid.com/ovidweb.cgi?T=JS&PAGE=reference&D=psyc18&NEWS=N&AN=2020-41795-001 | Wrong setting |
| Edgington L, Hill V, Pellicano E. The design and implementation of a CBT-based intervention for sensory processing difficulties in adolescents on the autism spectrum. Res Dev Disabil [Internet]. 2016;59:221–33. Available from: http://ovidsp.ovid.com/ovidweb.cgi?T=JS&PAGE=reference&D=psyc13&NEWS=N&AN=2016-54440-023 | Wrong setting |
| Malihi M, Nguyen J, Cardy RE, Eldon S, Petta C, Kushki A. Data-Driven Discovery of Predictors of Virtual Reality Safety and Sense of Presence for Children With Autism Spectrum Disorder: A Pilot Study. Front Psychiatry [Internet]. 2020;11:669. Available from: http://ovidsp.ovid.com/ovidweb.cgi?T=JS&PAGE=reference&D=pmnm5&NEWS=N&AN=32903670    %[20200804// | Wrong setting |
| McNally Keehn RH, Lincoln AJ, Brown MZ, Chavira DA. The Coping Cat program for children with anxiety and autism spectrum disorder: A pilot randomized controlled trial. J Autism Dev Disord [Internet]. 2013;43(1):57–67. Available from: http://ovidsp.ovid.com/ovidweb.cgi?T=JS&PAGE=reference&D=psyc10&NEWS=N&AN=2013-00365-006 | Wrong setting |
| Selles RR, Arnold EB, Phares V, Lewin AB, Murphy TK, Storch EA. Cognitive-behavioral therapy for anxiety in youth with an autism spectrum disorder: A follow-up study. Autism [Internet]. 2015;19(5):613–21. Available from: http://ovidsp.ovid.com/ovidweb.cgi?T=JS&PAGE=reference&D=psyc12&NEWS=N&AN=2015-26958-011 | Wrong setting |
| Maisel ME, Stephenson KG, Cox JC, South M. Cognitive defusion for reducing distressing thoughts in adults with autism. Res Autism Spectr Disord [Internet]. 2019;59:34–45. Available from: http://ovidsp.ovid.com/ovidweb.cgi?T=JS&PAGE=reference&D=psyc16&NEWS=N&AN=2019-02726-005 | Wrong setting |
| Wood JJ, Kendall PC, Wood KS, Kerns CM, Seltzer M, Small BJ, et al. Cognitive behavioral treatments for anxiety in children with autism spectrum disorder: A randomized clinical trial. JAMA Psychiatry [Internet]. 2020;77(5):474–83. Available from: http://ovidsp.ovid.com/ovidweb.cgi?T=JS&PAGE=reference&D=psyc17&NEWS=N&AN=2020-37326-005 | Wrong setting |
| Wood JJ, Drahota A, Sze K, Har K, Chiu A, Langer DA. Cognitive behavioral therapy for anxiety in children with autism spectrum disorders: A randomized, controlled trial. Journal of Child Psychology and Psychiatry [Internet]. 2009;50(3):224–34. Available from: http://ovidsp.ovid.com/ovidweb.cgi?T=JS&PAGE=reference&D=psyc6&NEWS=N&AN=2009-03619-004 | Wrong setting |
| Oka T, Ishikawa SI, Saito A, Maruo K, Stickley A, Watanabe N, et al. Changes in self-efficacy in Japanese school-age children with and without high autistic traits after the Universal Unified Prevention Program: a single-group pilot study. Child Adolesc Psychiatry Ment Health [Internet]. 2021;15(1):42. Available from: http://ovidsp.ovid.com/ovidweb.cgi?T=JS&PAGE=reference&D=pmnm5&NEWS=N&AN=34446074    %[20210826// | Wrong setting |
| Gobrial E, Raghavan R. Calm child programme: Parental programme for anxiety in children and young people with autism spectrum disorder and intellectual disabilities. Journal of Intellectual Disabilities [Internet]. 2018;22(4):315–27. Available from: http://ovidsp.ovid.com/ovidweb.cgi?T=JS&PAGE=reference&D=psyc15&NEWS=N&AN=2018-61721-002 | Wrong setting |
| Wood JJ, Drahota A, Sze K, Van Dyke M, Decker K, Fujii C, et al. Brief report: Effects of cognitive behavioral therapy on parent-reported autism symptoms in school-age children with high-functioning autism. J Autism Dev Disord [Internet]. 2009;39(11):1608–12. Available from: http://ovidsp.ovid.com/ovidweb.cgi?T=JS&PAGE=reference&D=psyc6&NEWS=N&AN=2009-18914-014 | Wrong setting |
| Tse ACY. Brief report: Impact of a physical exercise intervention on emotion regulation and behavioral functioning in children with autism spectrum disorder. J Autism Dev Disord [Internet]. 2020;50(11):4191–8. Available from: http://ovidsp.ovid.com/ovidweb.cgi?T=JS&PAGE=reference&D=psyc17&NEWS=N&AN=2020-16911-001 | Wrong setting |
| Godfrey E, Haythorne D. Benefits of dramatherapy for Autism Spectrum Disorder: A qualitative analysis of feedback from parents and teachers of clients attending Roundabout dramatherapy sessions in schools. Dramatherapy [Internet]. 2013;35(1):20–8. Available from: http://ovidsp.ovid.com/ovidweb.cgi?T=JS&PAGE=reference&D=psyc10&NEWS=N&AN=2014-27627-003 | Wrong setting |
| Doernberg EA, Russ S, ra W, Dimitropoulos A. Believing in make-believe: Efficacy of a pretend play intervention for school-aged children with high-functioning autism spectrum disorder. J Autism Dev Disord [Internet]. 2021;51(2):576–88. Available from: http://ovidsp.ovid.com/ovidweb.cgi?T=JS&PAGE=reference&D=psyc18&NEWS=N&AN=2020-44224-001 | Wrong setting |
| Reaven J, Reyes N, Pickard K, T, a T, Morris MA. Addressing the needs of diverse youth with ASD and anxiety in public schools: Stakeholder input on adaptations of clinic-based facing your fears. School Mental Health: A Multidisciplinary Research and Practice Journal [Internet]. 2020;12(2):308–22. Available from: http://ovidsp.ovid.com/ovidweb.cgi?T=JS&PAGE=reference&D=psyc16&NEWS=N&AN=2019-73237-001 | Wrong setting |
| Bischof NL, Rapee RM, Hudry K, Bayer JK. Acceptability and caregiver-reported outcomes for young children with autism spectrum disorder whose parents attended a preventative population-based intervention for anxiety: A pilot study. Autism Research [Internet]. 2018;11(8):1166–74. Available from: http://ovidsp.ovid.com/ovidweb.cgi?T=JS&PAGE=reference&D=psyc15&NEWS=N&AN=2018-23315-001 | Wrong setting |
| McBride NM, Weinzimmer SA, La Buissonnière-Ariza V, Schneider SC, Ehrenreich May J, Lewin AB, et al. The Impact of Comorbidity on Cognitive-Behavioral Therapy Response in Youth with Anxiety and Autism Spectrum Disorder. Child Psychiatry Hum Dev [Internet]. 2020 Aug 1 [cited 2023 Oct 13];51(4):625–35. Available from: https://link.springer.com/article/10.1007/s10578-020-00961-2 | Wrong outcome |
| Kinnaird E, Norton C, Stewart C, Tchanturia K. Same behaviours, different reasons: what do patients with co-occurring anorexia and autism want from treatment? International Review of Psychiatry [Internet]. 2019 May 19 [cited 2023 Oct 13];31(4):308–17. Available from: https://www.tandfonline.com/doi/abs/10.1080/09540261.2018.1531831 | Wrong outcome |
| Kamio Y, Haraguchi H, Miyake A, Hiraiwa M. Brief report: Large individual variation in outcomes of autistic children receiving low-intensity behavioral interventions in community settings. Child Adolesc Psychiatry Ment Health. 2015 Mar 25;9(1). | Wrong outcome |
| Jager-Hyman S, Maddox BB, Crabbe SR, Mandell DS. Mental Health Clinicians’ Screening and Intervention Practices to Reduce Suicide Risk in Autistic Adolescents and Adults. J Autism Dev Disord [Internet]. 2020 Oct 1 [cited 2023 Oct 13];50(10):3450–61. Available from: https://pubmed.ncbi.nlm.nih.gov/32240486/ | Wrong outcome |
| Graham YNH, Gerrard D, Laight S, Brown R, Keith S, Hayes C. More than medication: Evaluating the role of the pharmacist independent prescriber in a community team for learning disabilities and behaviours deemed to be challenging. Br J Learn Disabil [Internet]. 2020 Sep 1 [cited 2023 Oct 13];48(3):232–41. Available from: https://onlinelibrary.wiley.com/doi/full/10.1111/bld.12323 | Wrong outcome |
| Franck N, Bon L, Dekerle M, Plasse J, Massoubre C, Pommier R, et al. Satisfaction and Needs in Serious Mental Illness and Autism Spectrum Disorder: The REHABase Psychosocial Rehabilitation Project. Psychiatr Serv [Internet]. 2019 Apr 1 [cited 2023 Oct 13];70(4):316–23. Available from: https://pubmed.ncbi.nlm.nih.gov/30691384/ | Wrong outcome |
| Au-Yeung SK, Bradley L, Robertson AE, Shaw R, Baron-Cohen S, Cassidy S. Experience of mental health diagnosis and perceived misdiagnosis in autistic, possibly autistic and non-autistic adults. Autism [Internet]. 2019 Aug 1 [cited 2023 Oct 13];23(6):1508–18. Available from: https://pubmed.ncbi.nlm.nih.gov/30547677/ | Wrong outcome |
| Selman LE, Fox F, Aabe N, Turner K, Rai D, Redwood S. ’You are labelled by your children’s disability’-A community-based, participatory study of stigma among Somali parents of children with autism living in the United Kingdom. Ethn Health [Internet]. 2018;23(7):781–96. Available from: http://ovidsp.ovid.com/ovidweb.cgi?T=JS&PAGE=reference&D=psyc15&NEWS=N&AN=2018-37361-006 | Wrong outcome |
| Parry H, Mee S. Working with female adults living with autism in secure inpatient services. Nurs Times [Internet]. 2022;118(1):34–7. Available from: https://search.ebscohost.com/login.aspx?direct=true&AuthType=ip,shib&db=jlh&AN=154701676&site=ehost-live&scope=site&custid=s8454451 | Wrong outcome |
| Sharpley C, Bitsika V, Sarmukadam K. Variation in Social Anxiety and Separation Anxiety over Age in ASD and Non-ASD Young Males: Issues for Clinical Assessment and Treatment-Planning. Intl J Disabil Dev Educ [Internet]. 2021;68(5):678–89. Available from: https://search.ebscohost.com/login.aspx?direct=true&AuthType=ip,shib&db=jlh&AN=151876726&site=ehost-live&scope=site&custid=s8454451 | Wrong outcome |
| Dodds L, Spencer A, Shea S, Fell D, Armson BA, Allen AC, et al. Validity of autism diagnoses using administrative health data. Chronic Dis Can [Internet]. 2009;29(3):102–7. Available from: http://ovidsp.ovid.com/ovidweb.cgi?T=JS&PAGE=reference&D=med7&NEWS=N&AN=19527568 | Wrong outcome |
| Chlebowski C, Hurwich-Reiss E, Wright B, Brookman-Frazee L. Using stakeholder perspectives to guide systematic adaptation of an autism mental health intervention for latinx families: A qualitative study. J Community Psychol [Internet]. 2020;48(4):1194–214. Available from: http://ovidsp.ovid.com/ovidweb.cgi?T=JS&PAGE=reference&D=psyc17&NEWS=N&AN=2019-77345-001 | Wrong outcome |
| McCabe H. Two decades of serving children with autism in the People’s Republic of China: Achievements and challenges of a state-run mental health center. Disabil Soc [Internet]. 2008;23(3):271–82. Available from: http://ovidsp.ovid.com/ovidweb.cgi?T=JS&PAGE=reference&D=psyc6&NEWS=N&AN=2008-05593-007 | Wrong outcome |
| DaWalt LS, Greenberg JS, Mailick MR. Transitioning together: A multi-family group psychoeducation program for adolescents with ASD and their parents. J Autism Dev Disord [Internet]. 2018;48(1):251–63. Available from: http://ovidsp.ovid.com/ovidweb.cgi?T=JS&PAGE=reference&D=psyc15&NEWS=N&AN=2017-46976-001 | Wrong outcome |
| Reis J, Marchini S, Delvenne V. The Transition from Child and Adolescent Mental Health Services (CAMHS) To Adult Mental Health Services (AMHS): What about Autism Spectrum Disorder? Psychiatr Danub [Internet]. 2021;33:52–7. Available from: http://ovidsp.ovid.com/ovidweb.cgi?T=JS&PAGE=reference&D=med20&NEWS=N&AN=34862893 | Wrong outcome |
| Brookman-Frazee L, Drahota A, Stadnick N, Palinkas LA. Therapist perspectives on community mental health services for children with autism spectrum disorders. Administration and Policy in Mental Health and Mental Health Services Research [Internet]. 2012;39(5):365–73. Available from: http://ovidsp.ovid.com/ovidweb.cgi?T=JS&PAGE=reference&D=psyc9&NEWS=N&AN=2012-21287-004 | Wrong outcome |
| Kerns CM, Collier A, a, Lewin AB, Storch EA. Therapeutic alliance in youth with autism spectrum disorder receiving cognitive-behavioral treatment for anxiety. Autism [Internet]. 2018;22(5):636–40. Available from: http://ovidsp.ovid.com/ovidweb.cgi?T=JS&PAGE=reference&D=psyc15&NEWS=N&AN=2018-34089-012 | Wrong outcome |
| Klebanoff SM, Rosenau KA, Wood JJ. The therapeutic alliance in cognitive-behavioral therapy for school-aged children with autism and clinical anxiety. Autism [Internet]. 2019;23(8):2031–42. Available from: http://ovidsp.ovid.com/ovidweb.cgi?T=JS&PAGE=reference&D=psyc16&NEWS=N&AN=2019-60457-014 | Wrong outcome |
| Brewe AM, Mazefsky CA, White SW. Therapeutic alliance formation for adolescents and young adults with autism: Relation to treatment outcomes and client characteristics. J Autism Dev Disord [Internet]. 2021;51(5):1446–57. Available from: http://ovidsp.ovid.com/ovidweb.cgi?T=JS&PAGE=reference&D=psyc18&NEWS=N&AN=2020-57367-001 | Wrong outcome |
| Schwartz AE, Kramer JM, Rogers ES, McDonald KE, Cohn ES. Stakeholder-driven approach to developing a peer-mentoring intervention for young adults with intellectual/developmental disabilities and co-occurring mental health conditions. Journal of Applied Research in Intellectual Disabilities [Internet]. 2020;33(5):992–1004. Available from: http://ovidsp.ovid.com/ovidweb.cgi?T=JS&PAGE=reference&D=psyc17&NEWS=N&AN=2020-15698-001 | Wrong outcome |
| Alnemary FM, Aldhalaan HM, Simon-Cereijido G, Alnemary FM. Services for children with autism in the Kingdom of Saudi Arabia. Autism [Internet]. 2017;21(5):592–602. Available from: https://search.ebscohost.com/login.aspx?direct=true&AuthType=ip,shib&db=jlh&AN=123635326&site=ehost-live&scope=site&custid=s8454451 | Wrong outcome |
| Kerns CM, Berkowitz SJ, Moskowitz LJ, Drahota A, Lerner MD, Newschaffer CJ. Screening and treatment of trauma-related symptoms in youth with autism spectrum disorder among community providers in the United States. Autism [Internet]. 2020;24(2):515–25. Available from: https://search.ebscohost.com/login.aspx?direct=true&AuthType=ip,shib&db=jlh&AN=141396077&site=ehost-live&scope=site&custid=s8454451 | Wrong outcome |
| McKenzie R, Dallos R, Stedmon J, Hancocks H, Vickery PJ, Barton A, et al. SAFE, a new therapeutic intervention for families of children with autism: a randomised controlled feasibility trial. BMJ Open [Internet]. 2020;10(12):e038411. Available from: http://ovidsp.ovid.com/ovidweb.cgi?T=JS&PAGE=reference&D=med18&NEWS=N&AN=33384385 | Wrong outcome |
| Lee C eun, Burke MM, DaWalt LS, Li C, Taylor JL. The role of parental advocacy in addressing service disparities for transition-aged youth on the autism spectrum. Autism [Internet]. 2022;26(4):1001–6. Available from: https://search.ebscohost.com/login.aspx?direct=true&AuthType=ip,shib&db=jlh&AN=156316997&site=ehost-live&scope=site&custid=s8454451 | Wrong outcome |
| Hollocks MJ, Wood JJ, Storch EA, Cho AC, Kerns CM, Kendall PC. Reward sensitivity predicts the response to cognitive behavioral therapy for children with autism and anxiety. Journal of Clinical Child and Adolescent Psychology [Internet]. 2022;No-Specified. Available from: http://ovidsp.ovid.com/ovidweb.cgi?T=JS&PAGE=reference&D=psyc18&NEWS=N&AN=2022-27522-001 | Wrong outcome |
| Bradley E, Lunsky Y, Palucka A, Homitidis S. Recognition of intellectual disabilities and autism in psychiatric inpatients diagnosed with schizophrenia and other psychotic disorders. Adv Ment Health Intellect Disabil [Internet]. 2011;5(6):4–18. Available from: http://ovidsp.ovid.com/ovidweb.cgi?T=JS&PAGE=reference&D=psyc8&NEWS=N&AN=2011-28305-002 | Wrong outcome |
| Mouridsen SE, Rich B, Isager T. Psychiatric morbidity in disintegrative psychosis and infantile autism: A long-term follow-up study. Psychopathology [Internet]. 1999;32(4):177–83. Available from: http://ovidsp.ovid.com/ovidweb.cgi?T=JS&PAGE=reference&D=psyc3&NEWS=N&AN=1999-03135-003 | Wrong outcome |
| Webb EL, Greenwood A, Hamer A, Sibley V. Profiling referrals to a trauma support service: needs and outcomes for staff working in a secure adult developmental disorder pathway. Adv Ment Health Intellect Disabil [Internet]. 2021;15(5):209–21. Available from: https://search.ebscohost.com/login.aspx?direct=true&AuthType=ip,shib&db=jlh&AN=152512873&site=ehost-live&scope=site&custid=s8454451 | Wrong outcome |
| Williams ME, Haranin EC. Preparation of mental health clinicians to work with children with co-occurring autism spectrum disorders and mental health needs. J Ment Health Res Intellect Disabil [Internet]. 2016;9(1):83–100. Available from: http://ovidsp.ovid.com/ovidweb.cgi?T=JS&PAGE=reference&D=psyc13&NEWS=N&AN=2016-28325-006 | Wrong outcome |
| Penner M, Anagnostou E, Ungar WJ. Practice patterns and determinants of wait time for autism spectrum disorder diagnosis in Canada. Mol Autism [Internet]. 2018;9:16. Available from: http://ovidsp.ovid.com/ovidweb.cgi?T=JS&PAGE=reference&D=med15&NEWS=N&AN=29541438 | Wrong outcome |
| Garg P, Lillystone D, Dossetor D, Eastwood J, Liaw ST. A Pilot Study for Understanding the Perceptions of Australian General Practitioners Regarding Psychopharmacology for Children With Autism Spectrum Disorders. J Prim Care Community Health [Internet]. 2016;7(4):258–64. Available from: http://ovidsp.ovid.com/ovidweb.cgi?T=JS&PAGE=reference&D=med13&NEWS=N&AN=27103117 | Wrong outcome |
| Arm, -Branger S, Poisson N, Gaudoneix-Taieb M, Ramos O. Pharmaco-therapeutic evaluation of medical treatments of adult autistics and multi-handicapped patients in a public mental health unit. Les traitements psychotropes prescrits dans un etablissement public de sante mentale accueillant des personnes atteintes d’autisme et des polyhandicapes [Internet]. 2009;35(4):370–6. Available from: http://ovidsp.ovid.com/ovidweb.cgi?T=JS&PAGE=reference&D=psyc6&NEWS=N&AN=2009-16663-010 | Wrong outcome |
| Camm-Crosbie L, Bradley L, Shaw R, Baron-Cohen S, Cassidy S. “People like me don’t get support”: Autistic adults’ experiences of support and treatment for mental health difficulties, self-injury and suicidality. Autism [Internet]. 2019;23(6):1431–41. Available from: http://ovidsp.ovid.com/ovidweb.cgi?T=JS&PAGE=reference&D=psyc16&NEWS=N&AN=2019-40103-009 | Wrong outcome |
| Walhout SJ, van Zanten J, DeFuentes-Merillas L, Sonneborn CK, Bosma M. Patients With Autism Spectrum Disorder and Co-occurring Substance Use Disorder: A Clinical Intervention Study. Subst Abuse [Internet]. 2022;16:11782218221085600. Available from: http://ovidsp.ovid.com/ovidweb.cgi?T=JS&PAGE=reference&D=pmnm&NEWS=N&AN=35465619 | Wrong outcome |
| Brookman-Frazee L, Baker-Ericzen M, Stadnick N, Taylor R. Parent perspectives on community mental health services for children with autism spectrum disorders. J Child Fam Stud [Internet]. 2012;21(4):533–44. Available from: http://ovidsp.ovid.com/ovidweb.cgi?T=JS&PAGE=reference&D=psyc9&NEWS=N&AN=2012-19355-001 | Wrong outcome |
| Clark M, Adams D. Parent perspectives of what helps and hinders their child on the autism spectrum to manage their anxiety. Clinical Psychologist [Internet]. 2020;No-Specified. Available from: http://ovidsp.ovid.com/ovidweb.cgi?T=JS&PAGE=reference&D=psyc17&NEWS=N&AN=2020-34866-001 | Wrong outcome |
| Fraser R, Cotton S, Gentle E, Angus B, Allott K, Thompson A. Non-expert clinicians’ detection of autistic traits among attenders of a youth mental health service. Early Interv Psychiatry [Internet]. 2012;6(1):83–6. Available from: http://ovidsp.ovid.com/ovidweb.cgi?T=JS&PAGE=reference&D=psyc9&NEWS=N&AN=2012-02426-012 | Wrong outcome |
| Shivers CM, Sonnier‐Netto L, Lee GK. Needs and Experiences of Family Caregivers of Individuals With Autism Spectrum Disorders Across the Lifespan. J Policy Pract Intellect Disabil [Internet]. 2019;16(1):21–9. Available from: https://search.ebscohost.com/login.aspx?direct=true&AuthType=ip,shib&db=jlh&AN=135291566&site=ehost-live&scope=site&custid=s8454451 | Wrong outcome |
| Borgi M, Ambrosio V, Cordella D, Chiarotti F, Venerosi A. Nationwide survey of healthcare services for autism spectrum disorders (ASD) in Italy. Adv Neurodev Disord [Internet]. 2019;3(3):306–18. Available from: http://ovidsp.ovid.com/ovidweb.cgi?T=JS&PAGE=reference&D=psyc16&NEWS=N&AN=2019-55780-008 | Wrong outcome |
| Battaglia M, Detrick S, Fern, ez A. Multidisciplinary treatment for adults with autism spectrum disorder and co-occurring mental health disorders: Adapting clinical research tools to everyday clinical practice. J Ment Health Res Intellect Disabil [Internet]. 2016;9(4):232–49. Available from: http://ovidsp.ovid.com/ovidweb.cgi?T=JS&PAGE=reference&D=psyc13&NEWS=N&AN=2016-58842-003 | Wrong outcome |
| Storch EA, Wood JJ, Guzick AG, Small BJ, Kerns CM, Ordaz DL, et al. Moderators of response to personalized and standard care cognitive-behavioral therapy for youth with autism spectrum disorder and comorbid anxiety. J Autism Dev Disord [Internet]. 2022;52(2):950–8. Available from: http://ovidsp.ovid.com/ovidweb.cgi?T=JS&PAGE=reference&D=psyc18&NEWS=N&AN=2021-35353-001 | Wrong outcome |
| G, ia-Abellan H, Nieto C, Garcia-Rubio C. Mindfulness for adults with autism spectrum disorder and intellectual disability: A pilot study. J Intellect Disabil [Internet]. 2022;17446295221107284. Available from: http://ovidsp.ovid.com/ovidweb.cgi?T=JS&PAGE=reference&D=medp&NEWS=N&AN=35673766 | Wrong outcome |
| Heifetz M, Dyson A, a. Mindfulness-based group for teens with developmental disabilities and their parents: A pilot study. Mindfulness (N Y) [Internet]. 2017;8(2):444–53. Available from: http://ovidsp.ovid.com/ovidweb.cgi?T=JS&PAGE=reference&D=psyc14&NEWS=N&AN=2016-46901-001 | Wrong outcome |
| Williams ZJ, Cascio CJ, Woynaroski TG. Measuring subjective quality of life in autistic adults with the PROMIS global-10: Psychometric study and development of an autism-specific scoring method. Autism [Internet]. 2022;13623613221085364. Available from: http://ovidsp.ovid.com/ovidweb.cgi?T=JS&PAGE=reference&D=medp&NEWS=N&AN=35403453 | Wrong outcome |
| Jones HF, Ho ACC, Sharma S, Mohammad SS, Kothur K, Patel S, et al. Maternal thyroid autoimmunity associated with acute-onset neuropsychiatric disorders and global regression in offspring. Dev Med Child Neurol [Internet]. 2019;61(8):984–8. Available from: http://ovidsp.ovid.com/ovidweb.cgi?T=JS&PAGE=reference&D=psyc16&NEWS=N&AN=2019-06787-001 | Wrong outcome |
| Kennedy CH. Manipulating antecedent conditions to alter the stimulus control of problem behavior. J Appl Behav Anal [Internet]. 1994;27(1):161–70. Available from: http://ovidsp.ovid.com/ovidweb.cgi?T=JS&PAGE=reference&D=psyc3&NEWS=N&AN=1994-34400-001 | Wrong outcome |
| Borgi M, Chiarotti F, Aresu G, Gitti F, Fazzi E, Pierini A, et al. Management of Autism Spectrum Disorder in Italian Units of Child and Adolescent Mental Health: Diagnostic and Referral Pathways. Brain Sci [Internet]. 2022;12(2). Available from: http://ovidsp.ovid.com/ovidweb.cgi?T=JS&PAGE=reference&D=pmnm&NEWS=N&AN=35204027 | Wrong outcome |
| Maddox BB, Miyazaki Y, White SW. Long-term effects of CBT on social impairment in adolescents with ASD. J Autism Dev Disord [Internet]. 2017;47(12):3872–82. Available from: http://ovidsp.ovid.com/ovidweb.cgi?T=JS&PAGE=reference&D=psyc14&NEWS=N&AN=2016-16993-001 | Wrong outcome |
| Benevides TW, Shore SM, Palmer K, Duncan P, Plank A, Andresen ML, et al. Listening to the autistic voice: Mental health priorities to guide research and practice in autism from a stakeholder-driven project. Autism [Internet]. 2020;24(4):822–33. Available from: http://ovidsp.ovid.com/ovidweb.cgi?T=JS&PAGE=reference&D=psyc18&NEWS=N&AN=2020-36416-003 | Wrong outcome |
| Babb C, Brede J, Jones CRG, Elliott M, Zanker C, Tchanturia K, et al. “It’s not that they don’t want to access the support . . . it’s the impact of the autism”: The experience of eating disorder services from the perspective of autistic women, parents and healthcare professionals. Autism [Internet]. 2021;25(5):1409–21. Available from: http://ovidsp.ovid.com/ovidweb.cgi?T=JS&PAGE=reference&D=med19&NEWS=N&AN=33588579 | Wrong outcome |
| Spain D, Milner V, Mason D, Iannelli H, Attoe C, Ampegama R, et al. Improving cognitive behaviour therapy for autistic individuals: A delphi survey with practitioners. Journal of Rational-Emotive & Cognitive-Behavior Therapy [Internet]. 2022;No-Specified. Available from: http://ovidsp.ovid.com/ovidweb.cgi?T=JS&PAGE=reference&D=psyc18&NEWS=N&AN=2022-53635-001 | Wrong outcome |
| Brice S, Rodgers J, Ingham B, Mason D, Wilson C, Freeston M, et al. The importance and availability of adjustments to improve access for autistic adults who need mental and physical healthcare: findings from UK surveys. BMJ Open [Internet]. 2021;11(3):e043336. Available from: http://ovidsp.ovid.com/ovidweb.cgi?T=JS&PAGE=reference&D=med19&NEWS=N&AN=33737429 | Wrong outcome |
| Maddox BB, Crabbe S, Beidas RS, Brookman-Frazee L, Cannuscio CC, Miller JS, et al. “I wouldn’t know where to start”: Perspectives from clinicians, agency leaders, and autistic adults on improving community mental health services for autistic adults. Autism [Internet]. 2020;24(4):919–30. Available from: http://ovidsp.ovid.com/ovidweb.cgi?T=JS&PAGE=reference&D=psyc18&NEWS=N&AN=2020-36416-009 | Wrong outcome |
| Cai RY, Richdale A, a L., Dissanayake C, Uljarevic M. How does emotion regulation strategy use and psychological wellbeing predict mood in adults with and without autism spectrum disorder? A naturalistic assessment. J Autism Dev Disord [Internet]. 2020;50(5):1786–99. Available from: http://ovidsp.ovid.com/ovidweb.cgi?T=JS&PAGE=reference&D=psyc17&NEWS=N&AN=2019-10469-001 | Wrong outcome |
| Cummings JR, Lynch FL, Rust KC, Coleman KJ, Madden JM, Owen-Smith AA, et al. Health services utilization among children with and without autism spectrum disorders. J Autism Dev Disord [Internet]. 2016;46(3):910–20. Available from: http://ovidsp.ovid.com/ovidweb.cgi?T=JS&PAGE=reference&D=psyc13&NEWS=N&AN=2015-51198-001 | Wrong outcome |
| Hofer J, Hoffmann F, Dorks M, Kamp-Becker I, Kupper C, Poustka L, et al. Health services use and costs in individuals with autism spectrum disorder in Germany: Results from a survey in ASD outpatient clinics. J Autism Dev Disord [Internet]. 2022;52(2):540–52. Available from: http://ovidsp.ovid.com/ovidweb.cgi?T=JS&PAGE=reference&D=psyc18&NEWS=N&AN=2021-28305-001 | Wrong outcome |
| Maddox BB, Crabbe SR, Fishman JM, Beidas RS, Brookman-Frazee L, Miller JS, et al. Factors influencing the use of cognitive-behavioral therapy with autistic adults: A survey of community mental health clinicians. J Autism Dev Disord [Internet]. 2019;49(11):4421–8. Available from: http://ovidsp.ovid.com/ovidweb.cgi?T=JS&PAGE=reference&D=psyc16&NEWS=N&AN=2019-46445-001 | Wrong outcome |
| Hinde K, Mason J. Health practitioner knowledge and confidence in diagnosis and treatment of mental health issues in people with intellectual disabilities. J Intellect Dev Disabil [Internet]. 2020;45(3):269–78. Available from: https://search.ebscohost.com/login.aspx?direct=true&AuthType=ip,shib&db=jlh&AN=144525237&site=ehost-live&scope=site&custid=s8454451 | Wrong outcome |
| Adams L, Adamo N, Hollocks MJ, Valmaggia L, Brewster A, Watson J, et al. Examining clinicians’ concerns delivering telemental health interventions directly to autistic individuals during COVID-19. Res Autism Spectr Disord [Internet]. 2022;94:101956. Available from: http://ovidsp.ovid.com/ovidweb.cgi?T=JS&PAGE=reference&D=pmnm&NEWS=N&AN=35369648 | Wrong outcome |
| Piravej K, Tangtrongchitr P, Ch, arasiri P, Paothong L, Sukprasong S. Effects of Thai traditional massage on autistic children’s behavior. The Journal of Alternative and Complementary Medicine [Internet]. 2009;15(12):1355–61. Available from: http://ovidsp.ovid.com/ovidweb.cgi?T=JS&PAGE=reference&D=psyc6&NEWS=N&AN=2010-00064-004 | Wrong outcome |
| Drahota A, Wood JJ, Sze KM, Van Dyke M. Effects of cognitive behavioral therapy on daily living skills in children with high-functioning autism and concurrent anxiety disorders. J Autism Dev Disord [Internet]. 2011;41(3):257–65. Available from: http://ovidsp.ovid.com/ovidweb.cgi?T=JS&PAGE=reference&D=psyc8&NEWS=N&AN=2011-03834-001 | Wrong outcome |
| Zhang B, Liang S, Chen J, Chen L, Chen W, Tu S, et al. Effectiveness of peer-mediated intervention on social skills for children with autism spectrum disorder: a randomized controlled trial. Transl Pediatr [Internet]. 2022;11(5):663–75. Available from: http://ovidsp.ovid.com/ovidweb.cgi?T=JS&PAGE=reference&D=pmnm&NEWS=N&AN=35685075 | Wrong outcome |
| Bejarano-Martin A, Canal-Bedia R, Magan-Maganto M, Fern, ez-Alvarez C, Martin-Cilleros MV, et al. “Early detection, diagnosis and intervention services for young children with autism spectrum disorder in the European Union (ASDEU): Family and professional perspectives”: Correction. J Autism Dev Disord [Internet]. 2020;50(9):3395–6. Available from: http://ovidsp.ovid.com/ovidweb.cgi?T=JS&PAGE=reference&D=psyc17&NEWS=N&AN=2019-74370-001 | Wrong outcome |
| Zhou Y, Yin H, Wang M, Wang J. The effect of family-focused psychoeducational therapy for autism spectrum disorder children’s parents on parenting self-efficacy and emotion. Arch Psychiatr Nurs [Internet]. 2019;33(1):17–22. Available from: http://ovidsp.ovid.com/ovidweb.cgi?T=JS&PAGE=reference&D=psyc16&NEWS=N&AN=2018-39673-001 | Wrong outcome |
| Bates K, Morgan H, Crosby E, Nurse K, Flynn A, Stern D, et al. Developing digital approaches for adolescents and young adults with autism and learning disabilities: Tools to facilitate access and shared decision-making. Educational and Child Psychology [Internet]. 2021;38(3):124–39. Available from: http://ovidsp.ovid.com/ovidweb.cgi?T=JS&PAGE=reference&D=psyc18&NEWS=N&AN=2021-91171-008 | Wrong outcome |
| Green K, Williams C, Partridge I, Wright B. Developing a child and adolescent mental health service for children with learning disabilities. Psychiatric Bulletin [Internet]. 2001;25(7):264–7. Available from: http://ovidsp.ovid.com/ovidweb.cgi?T=JS&PAGE=reference&D=psyc3&NEWS=N&AN=2001-07825-008 | Wrong outcome |
| Stagi P, Galeotti S, Mimmi S, Starace F, Castagnini AC. Continuity of care from child and adolescent to adult mental health services: Evidence from a regional survey in Northern Italy. Eur Child Adolesc Psychiatry [Internet]. 2015;24(12):1535–41. Available from: http://ovidsp.ovid.com/ovidweb.cgi?T=JS&PAGE=reference&D=psyc12&NEWS=N&AN=2015-30328-001 | Wrong outcome |
| Wigham S, Ingham B, Le Couteur A, Wilson C, Ensum I, Parr JR. Consensus statements on optimal adult post-autism diagnosis support and services: Delphi process following a UK survey of autistic adults, relatives and clinicians. Autism [Internet]. 2022;13623613221097502. Available from: http://ovidsp.ovid.com/ovidweb.cgi?T=JS&PAGE=reference&D=medp&NEWS=N&AN=35670069 | Wrong outcome |
| Hepburn SL, Stern JA, Blakeley-Smith A, Kimel LK, Reaven JA. Complex psychiatric comorbidity of treatment-seeking youth with autism spectrum disorder and anxiety symptoms. J Ment Health Res Intellect Disabil [Internet]. 2014;7(4):359–78. Available from: http://ovidsp.ovid.com/ovidweb.cgi?T=JS&PAGE=reference&D=psyc11&NEWS=N&AN=2014-42717-005 | Wrong outcome |
| Babb C, Brede J, Jones CRG, Serpell L, M, y W, et al. A comparison of the eating disorder service experiences of autistic and non-autistic women in the uk. European Eating Disorders Review [Internet]. 2022;No-Specified. Available from: http://ovidsp.ovid.com/ovidweb.cgi?T=JS&PAGE=reference&D=psyc18&NEWS=N&AN=2022-79007-001 | Wrong outcome |
| Fong VC, Lee BS, Iarocci G. A community-engaged approach to examining barriers and facilitators to accessing autism services in Korean immigrant families. Autism [Internet]. 2022;26(2):525–37. Available from: http://ovidsp.ovid.com/ovidweb.cgi?T=JS&PAGE=reference&D=psyc18&NEWS=N&AN=2022-30803-019 | Wrong outcome |
| Roudbarani F, Tablon Modica P, Maddox BB, Bohr Y, Weiss JA. Clinician factors related to the delivery of psychotherapy for autistic youth and youth with attention-deficit hyperactivity disorder. Autism [Internet]. 2022;13623613221106400. Available from: http://ovidsp.ovid.com/ovidweb.cgi?T=JS&PAGE=reference&D=medp&NEWS=N&AN=35786029 | Wrong outcome |
| Melin J, Jansson-Frojmark M, Olsson NC. Clinical practitioners’ experiences of psychological treatment for autistic children and adolescents with school attendance problems: A qualitative study. BMC Psychiatry [Internet]. 2022;22. Available from: http://ovidsp.ovid.com/ovidweb.cgi?T=JS&PAGE=reference&D=psyc18&NEWS=N&AN=2022-49443-001 | Wrong outcome |
| Southey SJ, Stoddart KP. Clinical intervention with autistic adolescents and adults during the first two months of the COVID-19 pandemic: Experiences of clinicians and their clients. Int Soc Work [Internet]. 2021;64(5):777–82. Available from: https://search.ebscohost.com/login.aspx?direct=true&AuthType=ip,shib&db=jlh&AN=152311569&site=ehost-live&scope=site&custid=s8454451 | Wrong outcome |
| Moseley RL, Gregory NJ, Smith P, Allison C, Baron-Cohen S. A “choice”, an “addiction”, a way “out of the lost”: exploring self-injury in autistic people without intellectual disability. Mol Autism [Internet]. 2019;10:18. Available from: http://ovidsp.ovid.com/ovidweb.cgi?T=JS&PAGE=reference&D=med16&NEWS=N&AN=31007885 | Wrong outcome |
| Brookman-Frazee L, Chlebowski C, Suhrheinrich J, Finn N, Dickson KS, Aarons GA, et al. Characterizing shared and unique implementation influences in two community services systems for autism: Applying the EPIS framework to two large-scale autism intervention community effectiveness trials. Administration and Policy in Mental Health and Mental Health Services Research [Internet]. 2020;47(2):176–87. Available from: http://ovidsp.ovid.com/ovidweb.cgi?T=JS&PAGE=reference&D=psyc17&NEWS=N&AN=2019-16578-001 | Wrong outcome |
| Brookman-Frazee LI, Taylor R, Garl, Ann F. Characterizing community-based mental health services for children with autism spectrum disorders and disruptive behavior problems. J Autism Dev Disord [Internet]. 2010;40(10):1188–201. Available from: http://ovidsp.ovid.com/ovidweb.cgi?T=JS&PAGE=reference&D=psyc7&NEWS=N&AN=2010-19875-003 | Wrong outcome |
| Kerns CM, Maddox BB, Kendall PC, Rump K, Berry L, ra, et al. Brief measures of anxiety in non-treatment-seeking youth with autism spectrum disorder. Autism [Internet]. 2015;19(8):969–79. Available from: http://ovidsp.ovid.com/ovidweb.cgi?T=JS&PAGE=reference&D=psyc12&NEWS=N&AN=2015-47118-009 | Wrong outcome |
| White SW, Schry AR, Maddox BB. Brief report: The assessment of anxiety in high-functioning adolescents with autism spectrum disorder. J Autism Dev Disord [Internet]. 2012;42(6):1138–45. Available from: http://ovidsp.ovid.com/ovidweb.cgi?T=JS&PAGE=reference&D=psyc9&NEWS=N&AN=2012-14193-027 | Wrong outcome |
| Pedersen KA, Santangelo SL, Gabriels RL, Righi G, Erard M, Siegel M. Behavioral Outcomes of Specialized Psychiatric Hospitalization in the Autism Inpatient Collection (AIC): A Multisite Comparison. J Autism Dev Disord [Internet]. 2018;48(11):3658–67. Available from: http://ovidsp.ovid.com/ovidweb.cgi?T=JS&PAGE=reference&D=med15&NEWS=N&AN=29170939 | Wrong outcome |
| Raymaker DM, McDonald KE, Ashkenazy E, Gerrity M, Baggs AM, Kripke C, et al. Barriers to healthcare: Instrument development and comparison between autistic adults and adults with and without other disabilities. Autism [Internet]. 2017;21(8):972–84. Available from: http://ovidsp.ovid.com/ovidweb.cgi?T=JS&PAGE=reference&D=psyc14&NEWS=N&AN=2017-44446-007 | Wrong outcome |
| Frank HE, Kagan ER, Storch EA, Wood JJ, Kerns CM, Lewin AB, et al. Accommodation of anxiety in youth with autism spectrum disorder: Results from the TAASD study. Journal of Clinical Child and Adolescent Psychology [Internet]. 2022;51(2):219–29. Available from: http://ovidsp.ovid.com/ovidweb.cgi?T=JS&PAGE=reference&D=psyc17&NEWS=N&AN=2020-47887-001 | Wrong outcome |
| Kerns CM, Renno P, Kendall PC, Wood JJ, Storch EA. Anxiety Disorders Interview Schedule-Autism Addendum: Reliability and validity in children with autism spectrum disorder. Journal of Clinical Child and Adolescent Psychology [Internet]. 2017;46(1):88–100. Available from: http://ovidsp.ovid.com/ovidweb.cgi?T=JS&PAGE=reference&D=psyc14&NEWS=N&AN=2017-06060-006 | Wrong outcome |
| Meyer AT, Moody EJ, Keefer A, O’Kelley S, Duncan A, Blakeley-Smith A, et al. Effect of Co-occurring Psychiatric Disorders on Treatment of Children with Autism Spectrum Disorder and Anxiety. J Autism Dev Disord [Internet]. 2023 Feb 1 [cited 2023 Oct 13];53(2):569–79. Available from: https://pubmed.ncbi.nlm.nih.gov/32462457/ | Wrong outcome |
| Henderson-Laidlaw M, Hall S. Using cognitive behavioural therapy in individuals with intellectual disability. Learning Disability Practice [Internet]. 2020;23(3):40–5. Available from: https://search.ebscohost.com/login.aspx?direct=true&AuthType=ip,shib&db=jlh&AN=148703719&site=ehost-live&scope=site&custid=s8454451 | Unable to retrieve |
| González GC, Lester MR. Treatment services for individuals diagnosed with autism spectrum disorders in Quito, Ecuador. Perspectives on Global Issues in Communication Sciences & Related Disorders [Internet]. 2012;2(1):19–26. Available from: https://search.ebscohost.com/login.aspx?direct=true&AuthType=ip,shib&db=jlh&AN=104488215&site=ehost-live&scope=site&custid=s8454451 | Unable to retrieve |
| Hewitt L. Social Anxiety and Pragmatics in Autism Spectrum Disorders: Cognitive Behavioral Therapy and Its Relation to Speech-Language Pathology. Perspect Lang Learn Educ [Internet]. 2014;21(1):5–12. Available from: https://search.ebscohost.com/login.aspx?direct=true&AuthType=ip,shib&db=jlh&AN=104046365&site=ehost-live&scope=site&custid=s8454451 | Unable to retrieve |
| de Mir, a J. San Diego’s Project Connect: services for people with developmental disabilities. Alcoholism & Drug Abuse Weekly [Internet]. 2008;20(7):5–6. Available from: https://search.ebscohost.com/login.aspx?direct=true&AuthType=ip,shib&db=jlh&AN=105878650&site=ehost-live&scope=site&custid=s8454451 | Unable to retrieve |
| Davidson DA. Meeting the Mental Health Needs of Individuals with High-Functioning Autism Spectrum Disorders. Communique (Wash DC) [Internet]. 2016;(4):5–7. Available from: https://search.ebscohost.com/login.aspx?direct=true&AuthType=ip,shib&db=jlh&AN=124268634&site=ehost-live&scope=site&custid=s8454451 | Unable to retrieve |
| Jia YN, Gu JH, Wei QL, Jing YZ, Gan XY, Du XZ. [Effect of scalp acupuncture stimulation on mood and sleep in children with autism spectrum disorder]. Zhen Ci Yan Jiu [Internet]. 2021;46(11):948–52. Available from: http://ovidsp.ovid.com/ovidweb.cgi?T=JS&PAGE=reference&D=med20&NEWS=N&AN=34865332 | Unable to retrieve |
| Damodharan SK, Joji R. Delays in the diagnosis of autistic spectrum disorder in a community child and adolescent mental health service(CAMHS). Autism [Internet]. 2006;10(6):649–50. Available from: http://ovidsp.ovid.com/ovidweb.cgi?T=JS&PAGE=reference&D=med6&NEWS=N&AN=17203569 | Unable to retrieve |
| Davidson PW, Morris D, Cain NN. Community services for people with developmental disabilities and psychiatric or severe behaviour disorders. Psychiatric and behavioural disorders in developmental disabilities and mental retardation [Internet]. 1999;359–72. Available from: http://ovidsp.ovid.com/ovidweb.cgi?T=JS&PAGE=reference&D=psyc3&NEWS=N&AN=1999-02953-021 | Unable to retrieve |
| Chun TH, Katz ER, Duffy SJ, Gerson RS. Challenges of managing pediatric mental health crises in the emergency department. Child Adolesc Psychiatr Clin N Am [Internet]. 2015;24(1):21–40. Available from: http://ovidsp.ovid.com/ovidweb.cgi?T=JS&PAGE=reference&D=psyc12&NEWS=N&AN=2014-49476-006 | Unable to retrieve |

**Table S6.** Study design and study population characteristics

|  | **Author (Year)** | **Country** | **Aim** | **Study design** | **Setting** | **Baseline N** | **Participants** | **Strategy** | **Caregiver involvement in therapy** | **Comparison** |
| --- | --- | --- | --- | --- | --- | --- | --- | --- | --- | --- |
| **Detection of autism in mental health care** | | | | | | | | | | |
| 1 | Ford et al. (2019) | UK | To explore the levels of agreement about the diagnoses of autistic spectrum conditions between the referrer, practitioner and a research diagnosis, as well as the stability of the practitioner’s diagnosis over time. | Survey (Prospective cohort study) | Child and Adolescent Mental Health Services | CYP: 302 | 13.5% (n = 41) ASC research diagnosis, 19.5% (n = 59) suspected autism, of these 49% (n = 29) were given an ASC research diagnosis, 32% (n = 19) were given a definite ASC practitioner diagnosis and 42% (n = 25) were given a possible ASC practitioner diagnosis; 100% accessing Child and Adolescent Mental Health Services.  CYP. Age range 5-11 years. | Detection of autism using the DAWBA. | Not applicable. | Not applicable |
| 2 | Hollocks et al. (2019) | UK | To evaluate the sensitivity and specificity of the SCQ in a non-selected clinical sample of young people referred for an autism assessment. | Survey (Service evaluation) | Child and Adolescent Mental Health Services | CYP: 77 | 100% suspected autism, of which 57% (n = 44) met criteria for autism diagnosis; 100% accessing Child and Adolescent Mental Health Services.  CYP. *M* age = 12.8 years (*SD* = 3.6), range 6-19; 75.3% male. | Detection of autism using the SCQ. | Not applicable. | Not applicable |
| 3 | Stadnick et al. (2015) | USA | To examine the use of the ADOS when administered by clinicians to children referred for an autism assessment. | Cross-sectional survey (service evaluation) | Outpatient community-based mental health clinics | CYP: 62 | 56.4% received final ASC diagnosis; 100% accessing community mental health clinics; 37% anxiety disorders, 25% mood disorders, 19% disruptive behaviour disorder, 22% other; 58% ADHD.  CYP. *M* age = 10.69 years (*SD* = 3.48), range 5-18; 76% male; 67% non-White. | Detection of autism using the ADOS | Not applicable. | Not applicable |
| **Strategies for improving clinicians’ skills and autism knowledge** | | | | | | | | | | |
| 4 | Cervantes et al. (2019) | USA | To compare the outcomes of patients receiving the ASD-CP during an 18-month follow-up period to the outcomes of patients from the 18-month pre-implementation and 18-month initial evaluation periods, to assess maintenance of positive effects. | Before-and-after comparison (service implementation) | Psychiatric emergency care | CYP: 52 | 100% ASC diagnosis; co-occurring mental health difficulties.  CYP. Pre-implementation: 35.3% age 4-12 years and 64.7% age 13-17 years, 76.5% male. Post-implementation: 60% age 4-12 years and 40% age 13-17 years, 95% male  Follow-up: 66.7% age 4-12 years, 33.3% age 13-17 years; 100% male. | ASD-CP consisting of a modular staff training, toolkit and prescribed practices to be utilised with the person.  Face-to-face; 45 minutes x 4 training modules. Delivered by hospital staff who have been trained. | Not applicable. | Comparing pre-implementation (n = 17), post-implementation (n = 20) and follow-up (n = 15) |
| 5 | Dreiling et al. (2022) | USA | To develop and implement Project ECHO for community mental health providers. | Non-randomised service evaluation | Community services | Staff: 86 | Mental health providers working with autistic people across the lifespan.  *M* age = 42.22 years (*SD* = 10.6), range 25-66; 6% male; 14% non-White. | Project ECHO, a tele-mentoring platform to support mental health professionals when working with autistic people with co-occurring mental health difficulties.  Virtual; 90 minutes x 10 bi-weekly sessions over a period of 6 months. Panel consisted of two senior psychologists and 1 clinician and 1 parent advocate/professional autism resource specialist. | Not applicable. | Not applicable |
| 6 | Helverschou et al. (2021) | Norway | To describe patterns of psychiatric and behaviour problems in autistic people with ID, treated by the AUP network. | Before-and-after comparison (service evaluation) | Specialist hospital-level mental health services | CYP and adults: 132 | 100% ASC diagnosis; 100% ID diagnosis; co-occurring: 32.6% psychosis, 50.8% depression, 44.7% anxiety, 15.9% OCD.    CYP and adults. *M* age = 28.6 years (*SD* = 10.6), range 16-66; 67% male. | AUP network consisting of meetings and seminars to guide mental health professionals in providing specialised mental health care to autistic people with ID.  Face-to-face across 8 clinics. Network meetings occur yearly over 6 days, and seminars occur every other year for 2 days. Delivered by professionals. | Not applicable. | Not applicable |
| 7 | Kuriakose et al. (2018) - linked to Cervantes et al. (2019) | USA | To evaluate the effectiveness of a specialised autism spectrum care pathway for adolescents in inpatients psychiatric units. | Before-and-after comparison (service implementation) | Psychiatric emergency care | CYP: 37 | 100% ASC diagnosis; co-occurring mental health difficulties.  CYP. Pre-implementation: 35.3% age 4-12 years and 64.7% age 13-17 years; 76.5% male. Post-implementation: 60% age 4-12 years and 40% age range 13-17 years; 95% male. | ASD-CP consisting of a modular staff training, toolkit and prescribed practices to be utilised with the person.  Face-to-face; 45 minutes x 4 training modules. Delivered by hospital staff who have been trained. | Not applicable. | Comparing pre-implementation (n = 17) and post-implementation (n = 20) |
| **General adaptations to standard practice** | | | | | | | | | | |
| 8 | Jones et al. (2021) | UK | To explore skills and adaptations to inpatient units. | Cross-sectional survey | Inpatient psychiatric services. | Staff: 90 | Clinicians, some of whom had experience working with autistic people across the lifespan. | Evaluation of strategies and adaptations to psychiatric inpatient care. | Not applicable. | Not applicable |
| 9 | Petty et al. (2021) | UK | To inform service development within mental health services for autistic clients. | Qualitative (ethnographic technique) | Specialist autism service | Staff: 15 | Staff working with autistic people across the lifespan.  Age range 25-44 years; 20% male, 73% White British. | Evaluation of adaptations to improve mental health care. | Not applicable. | Not applicable |
| 10 | Spain et al. (2017) | UK | To ascertain professional perspectives about social anxiety in autism and to establish how, if at all, clinicians and researchers adapt their practice when working with this population. | Qualitative (thematic analysis) | Inpatient and outpatient services | Staff: 21 | Multidisciplinary team professionals working with autistic people across the life span with co-occurring social anxiety.  All had several years’ experience of working with people with developmental disorders. | Evaluation of general adaptations to standard practice. | Not applicable. | Not applicable |
| **Group CBT for anxiety** | | | | | | | | | | |
| 11 | Bemmer et al. (2021) | Australia | To evaluate the benefit, tolerability and acceptability of adapted CBT for social anxiety. | Before-and-after comparison | Research clinic within primary health care network and headspace clinical services | CYP and adults: 84 | 100% ASC diagnosis.    CYP and adults. *M* age = 22.77 years (*SD* = 5.31), range 16-38; 60% male. | Adapted CBT for social anxiety.    Group; face-to-face; 2.5 hours x 8 weekly sessions. Facilitated by staff with prior experience working with autistic people. 60-minute debrief following each session. | Yes. Participants were required to nominate a social support person (e.g., parent, partner, housemate or friend) to help practice skills, increase compliance with homework tasks and to increase the ability to generalise the application of CBT strategies to different social contexts. | Not applicable |
| 12 | Burke et al. (2017) | Ireland | To investigate the impact of adapted “FRIENDS for Life” among autistic children aged between 10 and 11 years. | Mixed methods (quantitative: before-and-after comparison; qualitative: thematic analysis) | Specialist autism service | CYP: 7 | 100% ASC diagnosis; 100% referred for anxiety intervention.  CYP. Age range 10 – 11 years. | Adapted CBT for anxiety (‘FRIENDS for life’ programme).  Group; 2 hours x 10 weekly sessions. Delivered by facilitators. | No information about involvement in sessions, but two parent and one teacher information evenings were delivered to share pictures of the immediate FRIENDS environment, the facilitators and location of the service, with the aim of facilitating the assent process, increasing engagement and supporting the process. | Not applicable |
| 13 | Chalfant et al. (2007) | Australia | To investigate the effectiveness of CBT for autistic children who have a co-occurring anxiety. | RCT | School Outreach Service of the Autism Association in New South Wales | CYP: 47 | 100% ASC diagnosis; 100% anxiety disorder diagnosis.  CYP. *M* age = 10.8 years (*SD* = 1.35), range 8-13; 74.5% male. | Adapted CBT for anxiety (manualized ‘Cool Kids’ programme) (n = 28).  Group; face-to-face; 2 hours x 12 weekly sessions. Delivered by clinical psychologists. | Yes. Families planned out exposure tasks and were asked to make a daily diary entry to record the practice and outcomes from their exposure activities. Parent-based group CBT manual was also adapted to be used in concurrent family sessions. | Waitlist (n = 19) |
| 14 | Cook et al. (2017) | Australia | To investigate the efficacy of a parent-mediated CBT group anxiety intervention with autistic young children. | Pilot RCT | University Psychology Clinic | CYP: 31  Parents: 31 | 100% ASC diagnosis; 100% met cut-off anxiety levels.  CYP. *M* age = 5 years (*SD* = 0.83); 87.1% male. CBT: *M* age = 5.5 years (*SD* = 0.88); 85.7% male. Waitlist: *M* age 5.42 years (*SD* = 0.81); 88.2% male. | Adapted parent-mediated CBT for anxiety (‘Fun with Feelings’ programme) for children aged 4-6 years (n = 14).  Group; face-to-face; 90 minutes x 9 weekly sessions + 3 monthly booster sessions. Delivered by psychologists. | Yes. Parents were instructed in CBT strategies and skills, that they in turn taught to their children. | Waitlist (n = 17) |
| 15 | Hepburn et al. (2016) | USA | To detail the pilot testing of a telehealth version of an empirically supported intervention targeting anxiety in autistic youth. | Non-randomised controlled trial | Specialist autism clinic | CYP: 33  Parents: 33 | 100% ASC diagnosis; 100% met cut-off anxiety levels.  CYP. 81.8% male; 12.1% non-White. CBT: *M* age = 11.5 years (*SD* = 2.67); 82.4% male; 12.5% non-White. Waitlist: *M* age = 12.1 years (*SD* = 1.96); 81.3% male; 11.8% non-White. | Adapted CBT for anxiety (telehealth manualized ‘FYF’ programme) (n = 17).  Group; virtual; 1.5 hours x 12 sessions over 3–4-months. Delivered by therapists over a video conferencing platform. | Yes. They had individual & combined sessions. Youth and parents attended the  sessions together, but the youth usually left the session approximately 20-30 minutes before the parents and therapists so that the therapists could provide additional support to parents in their roles as coaches. | Waitlist (n = 16) |
| 16 | E. Higgins et al. (2019) | Ireland | To investigate the impact of the ‘Special FRIENDS’ programme on anxiety levels. | Mixed methods (quantitative: before-and-after comparison; qualitative: thematic analysis) | Children disability service | CYP: 12 | 100% ASC diagnosis; 100% referred for specific anxiety management intervention.  *M* age = 10.25 years (*SD* = 1.26), range 9-12; 82% male. | Adapted CBT for anxiety (‘Special FRIENDS’ programme).  Group; 1 hour x 12 weekly sessions + 2 booster sessions at 1 and 3 months after programme completion. Delivered by psychologist, with assistance from trainee educational psychologists. | Yes. Parent component consisting of psycho-educational sessions. | Not applicable |
| 17 | Keefer et al. (2017) - linked to Reaven et al. (2018) | USA | To compare levels of intolerance of uncertainty, anxiety and worry in autistic youth to ascertain change before and after FYF and to examine whether high and low levels of pre-intervention intolerance uncertainty predicted response to FYF. | Before-and-after comparison | University outpatient clinic | CYP: 43  Parents: 43 | 100% ASC diagnosis; 100% clinically significant anxiety symptoms.  CYP. *M* age = 11.18 years (*SD* = 2.02); 81% male. | Bespoke CBT for anxiety (manualized ‘FYF’ programme).  Group; face-to-face; 90 minutes x 12 sessions. Delivered by clinical psychologist and co-therapists. | Yes. Separate parent meetings and parent-child dyads. | Not applicable |
| 18 | Kilburn et al. (2019) | Denmark | To investigate the feasibility of the manualised CBT group programme 'Cool Kids’ for anxiety adapted for autistic children in a general hospital setting. | Before-and-after comparison | Outpatient clinic at the Centre of Child and Adolescent Psychiatry | CYP: 9 | 100% ASC diagnosis; 100% referred to clinic with anxiety symptoms.  CYP. Age range 9-13 years; 77.7% male. | Adapted CBT for anxiety (manualized ‘Cool Kids’ programme).  Group; 2 hours x 12 sessions over 15-week period (first 9 weekly and remaining fortnightly). Delivered by psychologists and clinicians with experience of autism and CBT. | Yes. Parents had separate and conjoint sessions. Not necessary for the same parent to attend every session. | Not applicable |
| 19 | Kilburn et al. (2020) | Denmark | To investigate the efficacy of the manualised CBT ‘Cool Kids’ anxiety programme. | RCT | Outpatient clinic in a general child psychiatric hospital | CYP: 49  Parents: 49 | 100% ASC diagnosis; 100% clinically significant anxiety symptoms.  CYP. *M* age = 11.34 years (*SD* = 1.77); 57% male. CBT: *M* age = 11.99 years (*SD* = 1.70); 60% male. Waitlist: M age = 10.68 years (SD = 1.60); 54.2% male. | Adapted CBT for anxiety (manualized ‘Cool Kids’ programme) (n = 25).  Group; 2 hours x 10 weekly sessions with one-week break after session 3, 6, 8. Sessions consisted of time spent with child and parent separately and conjointly. Delivered by main therapist and co-therapist. | Yes. Parents had separate and conjoint sessions. | Waitlist (n = 24) |
| 20 | Langdon et al. (2016) | UK | To examine whether bespoke CBT for anxiety is feasible and likely to be efficacious. | Pilot single-blind RCT | Community-based settings | CYP and adults: 52 | 100% ASC diagnosis; 100% met cut-off anxiety levels.  CYP and adults. *M* age = 35.9 years (*SD* = 14.6), 17-65; 52% male; 0% non-White. CBT: *M* age 33.1 years (*SD* = 14.6), range 20-64; 46% male; 96% White British. Waitlist: *M* age 38.7 years (*SD* = 14.3), range 17-65; 58% male; 100% White British. | Bespoke CBT for anxiety + TAU (n = 26).  Group; face-to-face; 1 hour x 24 weekly manualized sessions (participants received 3 initial sessions of 1:1 CBT, followed by 21 group CBT sessions). Delivered by clinical psychologist or CBT therapist. | No information. | Waitlist (n = 26) |
| 21 | McConachie et al. (2014) | UK | To investigate the acceptability and feasibility of adapted group therapy for anxiety in autistic children. | Pilot RCT | Child and Adolescent Mental Health Services | CYP: 32  Parents: 32 | 100% ASC diagnosis; 100% anxiety disorder diagnosis.  CYP. CBT: *M* age = 11.7 years (*SD* = 1.4); boys:girls 15:2. Delayed therapy: *M* age = 11.8 years (*SD* = 1.3); boys:girls 13:2. | Bespoke CBT for anxiety (manualized ‘Exploring Feelings’ programme) (n = 17).  Group; 2 hours x 7 sessions. Delivered by two group leaders. | Yes. Parents had their own group run in parallel and worked through the same exercises and materials. | Delayed therapy (n = 15) |
| 22 | Pickard et al. (2020) - linked to Reaven et al. (2018) | USA | To examine the implementation and treatment outcomes in response to a group CBT programme for autistic youth with co-occurring anxiety. | Mixed methods survey | University-based clinics | Staff: 34 | Clinicians working with autistic CYP and trained to deliver FYF.  10% male; 10% non-White. | Bespoke CBT for anxiety for youth 8-14 years (manualized ‘FYF’ programme)  Group; face-to-face, 90 minutes x 14 weekly sessions. Delivered by clinical psychologist and co-therapists. | Yes. Separate parent meetings and parent-child dyads. | Not applicable |
| 23 | Reaven et al. (2009) | USA | To assess the effectiveness of an original, manualised, cognitive-behavioural group treatment with parental involvement, in the reduction of anxiety symptoms in autistic children. | Non-randomised controlled trial | Community-based services | CYP: 33  Parents: 33 | 100% ASC diagnosis; 100% clinically significant anxiety symptoms.  CYP. 78.8% male; 18.2% non-White. *M* age = 132 months (*SD* = 22.80), range 97-177. CBT: 70% male; 20% non-White. Waitlist: 82.6% male; 17.4% non-White. | Bespoke CBT for anxiety (manualized ‘FYF’ programme) (n = 10).  Group; 90 minutes x 12 weekly sessions. Delivered by clinical psychologist, or graduate student or postdoctoral fellow under psychologist’s supervision. | Yes. Separate parent meetings and parent-child dyads. | Waitlist (n = 23) |
| 24 | J. Reaven, Blakeley-Smith, Culhane-Shelburne et al. (2012) | USA | To examine the efficacy of a family-focused group CBT program developed specifically for autistic children with anxiety. | RCT | University outpatient clinic | CYP: 50  Parents: 50 | 100% ASC diagnosis; 100% clinically significant anxiety symptoms.  CYP. 96% male; 16% non-White. CBT: *M* age = 125.75 months (*SD* = 21.47); 100% male; 8.3% non-White. TAU: *M* age = 125 months (*SD* = 20.45); 92.3% male; 23.1% non-White. | Bespoke CBT for anxiety (manualized ‘FYF’ programme) (n = 24).  Group; face-to-face; 90 minutes x 12 sessions. Delivered by clinical psychologist and co-therapists. | Yes. Separate parent meetings and parent-child dyads. | TAU (n = 26) |
| 25 | J. Reaven, Blakeley-Smith, Leuthe et al. (2012) | USA | To develop an intervention for treating anxiety in autistic adolescents based on a CBT programme designed for school-aged children. | Before-and-after comparison study | Research clinic | CYP: 24 | 100% ASC diagnosis; 100% clinically significant anxiety symptoms.  CYP. *M* age = 15.5 years (*SD* = 13.4), range 13.4-18; 62.5% male; 33.3% non-White. | Adapted CBT for anxiety (manualized ‘FYF’ programme - adolescent version)  Group; face-to-face, 90 minutes x 14 sessions + 1 booster session. Delivered by two clinical psychologists and two co-therapists. | Yes. Separate parent meetings and parent-child dyads. | Not applicable |
| 26 | Reaven et al. (2015) | USA and Canada | To train clinicians to deliver FYF to fidelity and examine feasibility of the programme for novel settings. | Before and after comparison study (feasibility study) | Tertiary paediatric health centre | CYP: 16  Parents: 16  Clinicians: 13 | 100% ASC diagnosis; 100% clinically significant anxiety symptoms.  CYP*. M* age = 10.4 years (*SD* = 1.5), range 8-13; 85.7% male; 0% non-White. | Bespoke CBT for anxiety (manualized ‘FYF’ programme).  Group; face-to-face; 90 minutes x 12 sessions. Delivered by clinical psychologist and co-therapists. | Yes. Separate parent meetings and parent-child dyads. | Not applicable |
| 27 | Reaven et al. (2018) | USA | To expand on previous research and explore the implementation of group CBT for anxiety in autistic youth ages 8–14. | RCT (3-group parallel design) | University-affiliated outpatient clinics | CYP: 91  Parents: 91  Clinicians: 34 | 100% ASC diagnosis; 100% clinically significant anxiety symptoms.  CYP. 83.5% male; 33% non-White. Manual: *M* age = 132.9 months (*SD* = 20.4); 90.9% male; 9.1% non-White. Workshop only: *M* age = 130.8 months (*SD* = 27.6); 86.7% male; 16.7% non-White. Workshop plus: *M* age = 135.5 months (*SD* = 19.5); 71.4% male; 57% non-White. | Bespoke CBT for anxiety (manualized ‘FYF’ programme).  Group; face-to-face, 90 minutes x 14 weekly sessions. Delivered by clinical psychologist and co-therapists. | Yes. Separate parent and child meetings and parent-child dyads. | 3 groups receiving intervention, each group had therapists trained in a different way: standard manual (n = 33); workshop only (n = 30), workshop plus (n = 28) |
| 28 | Sofronoff et al. (2005) | Australia | To evaluate the effectiveness of a brief CBT intervention for anxiety with autistic children. | RCT | University psychology clinic | CYP: 71 | 100% ASC diagnosis; 100% parent-report anxiety; 42.2% ADHD.  CYP. 87.3% male. CBT child only: *M* age = 10.56 years (*SD* = 0.99), range 9-12; 86.9%% male. CBT child + parent: *M* age = 10.54 years (*SD* = 1.26), range 9-12; 88% male.  Waitlist: *M* age = 10.75 years (*SD* = 1.04) range 9-12; 86.9% male. | Bespoke manualised CBT for anxiety (Exploring Feelings; (child only n = 23; child + parent n = 25).  Group; 2 hours x 6 sessions. Delivered by postgraduate students in clinical psychology. | Yes. Child only - Parents received no training but after each session met with therapists for feedback. Child + parent - Parents formed parent groups and a therapist trained parents to work as co-therapists in all components of the intervention. | Waitlist (n = 23) |
| 29 | Solish et al. (2020) | Canada | To examine the effectiveness of a manualised group CBT programme. | Before-and-after comparison (service evaluation) | Community clinics and specialised hospital | CYP: 105  Parents: 105 (completers) | 100% ASC diagnosis; 100% anxiety symptoms; ID (n = 4).  CYP. Specialised hospital: *M* age 10.08 years (*SD* = 1.71); 74.1% male. Community: *M* age 10.87 (*SD* = 1.72); 70.6% male. | Bespoke CBT for anxiety (manualized ‘FYF’ programme)  Group; 1.5 hours x 14 weekly sessions. Delivered by at least one clinician. | Yes. Separate parent and child meetings and parent-child dyads. | Not applicable |
| 30 | Sung et al. (2011) | Singapore | To compare the effects of a 16-week CBT programme and adapted Social Recreational programme on anxiety in autistic children. | RCT | Outpatient mental health clinic for children and adolescents | CYP: 70 | 100% ASC diagnosis; 100% anxiety-related issues.  CYP. 9-16 years; 100% non-White. 94.3% male. CBT: *M* age = 11.33 years (*SD* = 2.03); 94.4% male; 100% non-White. Social recreational: *M* age = 11.09 years (*SD* = 1.53); 94.1% male; 100% non-White. | Adapted CBT for anxiety (n = 36)  Group; face-to-face; 90 minutes x 16 weekly sessions, manualized but retained flexibility. Delivered by therapists. | No. | Adapted Social Recreational programme (n = 34) |
| 31 | Walsh et al. (2018) - linked to Reaven et al. (2018) | USA | This study focuses on parent, youth, and clinician acceptability of a well-researched CBT programme, Facing Your Fears, for autistic youth with anxiety. | RCT (3 group parallel design) | University-affiliated outpatient clinics | CYP: 80  Parents: 80 (completers)  Clinicians: 34 | 100% diagnosed with ASC; 100% clinically significant symptoms of anxiety.  CYP. *M* age = 133.35 (*SD* = 23.59); 83.8% male; 32.6% non-White. | Bespoke CBT for anxiety (manualized ‘FYF’ programme)  Group; face-to-face, 90 minutes x 14 weekly sessions. Delivered by clinical psychologist and co-therapists. | Yes. Separate parent and child meetings and parent-child dyads. | 3 groups receiving intervention, each group had therapists trained in a different way: standard manual; workshop only, workshop plus |
| **Individual CBT for anxiety** | | | | | | | | | | |
| 32 | Driscoll et al. (2020) | USA | To pilot and explore the feasibility  and efficacy of a family-centred CBT protocol. | Before-and-after comparison (open pilot) | Outpatient clinics of two urban hospitals affiliated with a medical school | CYP: 16 | 100% ASC diagnosis; 100% anxiety disorder diagnosis; 12.5% OCD; 37.5% ADHD.  CYP. *M* age = 5.7 years (*SD* = 1.4), range 3-7; 81.25% male; 88% European-American, 6% Asian-American, 6% African-American. | Adapted CBT for anxiety (manualized ‘Being Brave’ programme).  Individual; face-to-face; 1 hour x 15-20 weekly sessions. Delivered by psychologist. | Yes. Parent meetings to provide psychoeducation (3 sessions), to teach them how to play with their children in a relaxed way (3 sessions) and to maintain gains (1 final session). Next 8-13 sessions involved therapy with child and 1 or both parents. | Not applicable |
| 33 | Ehrenreich-May et al. (2014) | USA | To determine whether BIACA, in its original form, would be feasible and evidence preliminary efficacy with younger adolescents. | Before-and-after comparison (open trial) | University treatment centres | CYP: 20 | 100% ASC diagnosis; 100% anxiety disorder diagnosis.  CYP. *M* age = 12.2 years (*SD* = 1.11), range 11-14; 90% male; 15% non-White | Adapted CBT for anxiety (modified ‘BIACA’ programme).  Individual; 60-90 minutes x 16 weekly sessions with modular, flexible format. 30 minutes of each session are spent with adolescent (14 modules), followed by 30 minutes with the parent (12 modules). Delivered by clinicians and therapists. | Yes. Parent modules were provided. | Not applicable |
| 34 | Ekman et al. (2015) | Sweden | To investigate benefits of adapted CBT for anxiety using visualisation and communication. | Before-and-after comparison (quasi-experimental open pilot) | Private clinic, child and adolescent psychiatric clinic, treatment centre for youth | CYP and adults: 18 | 100% ASC diagnosis; 100% anxiety disorder diagnosis and avoidance behaviour.  CYP and adults. *M* age for teens = 14.9 years (*SD* = 1.5), range 13-17; 22.2% male. *M* age for adults = 29.8 years (*SD* = 4.4), range 23-36, 38.9% male. | Adapted CBT (not manual-based) combined with visualised language for anxiety.  Individual; face-to-face; 45-60 minutes x 15 sessions fortnightly or at client's convenience. Delivered by CBT therapists with prior experience working with autistic people. | No information. | Not applicable |
| 35 | Fujii et al. (2013) | USA | To examine the efficacy of an intensive 32-week family-based CBT to treat co-occurring anxiety disorders in autistic youth ages 7–11 years. | Pilot RCT | University clinic and an associated autism community clinic | CYP: 12 | 100% ASC diagnosis; 100 % anxiety disorder diagnosis.  CYP. *M* age = 8.80 years (*SD* = 1.60), 7-11; 75% male, 25% non-White. CBT: *M* age = 8.7 years (*SD* = 1.8); 71% male; 14% non-White. TAU: *M* age = 9 years (*SD* = 1.6); 80% male; 40% non-White. | Adapted CBT for anxiety (modified ‘BIACA’ programme) (n = 7).  Individual; face-to-face; 90 minutes x 32 weekly sessions with modular format (30 minutes of each session spent with child and parent(s) separately and 30 minutes conjointly). Delivered by graduate or postgraduate psychology/psychiatry students. | Yes. Parents had separate and conjoint sessions. | TAU (n = 5) |
| 36 | Maskey, McConachie, et al. (2019) | UK | To explore a modified version of graded exposure treatment alongside CBT in autistic young people with specific fears and phobias. | Before-and-after comparison | NHS/university setting | CYP: 8 | 100% ASC diagnosis; 100% had specific phobia/fear.  CYP. Age range 8-12 years; 100% male. | Bespoke CBT for fears and phobias using flat screen computer delivery of images (‘Blue Room’).  Individual; 20 minutes x 4 sessions over 2 half days, with a week apart. Delivered by a psychologist. | Can’t tell. Parents observed sessions from an adjacent room via a one-way mirror. At the end of sessions, there was planning with family how to gradually increase exposure to the phobia outside of the clinic setting. | Not applicable |
| 37 | Maskey, Rodgers et al. (2019) | UK | To examine the feasibility and acceptability of using an immersive virtual reality environment alongside CBT for autistic young people experiencing specific phobia. | Feasibility RCT | NHS/university setting | CYP: 32  Parents: 32 | 100% ASC diagnosis; 100% had social/specific phobia.  CYP. 78.1% male; 6.25% non-White. CBT: *M* age = 130.13 months (*SD* = 28.38), range 89-174; 81.3% male; 0% non-White. Delayed therapy: *M* age = 129 months (*SD* = 21.51), range 90-157; 75% male; 12.5% non-White. | Bespoke CBT for fears and phobias with virtual reality (‘Blue Room’) (n = 16).  Individual; 20 minutes x 4 sessions over 2 half days, with a week apart. Delivered by a psychologist. | Can’t tell. Parent and child attended a 45-minute session with their allocated therapist prior to the commencement of the intervention. Parents watched treatment via a video link. | Delayed therapy (n = 16) |
| 38 | Oerbeck et al. (2021) | Norway | To test the feasibility of the CBT programme "Less stress" for co-occurring anxiety disorders in autistic children. | Before-and-after comparison study | Child and Adolescent Mental Health Clinics | CYP: 10 | 100% ASC diagnosis; 100% met cut-off anxiety levels.  CYP. *M* age = 9.5 years, range 8-12; 80% male. | Adapted manual-based CBT for anxiety (‘Less Stress’ programme).  Individual; face-to-face, 13 weekly sessions + 3 monthly follow up sessions. Delivered by therapists. | Yes. Parents joined all sessions. Two parent sessions were also offered, one before the commencement of programme and one in the middle of the programme. | Not applicable |
| 39 | Ollendick et al. (2021) | USA | To examine the initial feasibility and efficacy of a version of OST that was modified for autistic children. | Before-and-after comparison study | Clinic | CYP: 9 | 100% ASC diagnosis; 100% specific phobia diagnosis.  CYP. Age range 6-14 years; 100% male; 33% non-White. | Adapted CBT for specific phobia (‘OST’).  Individual; 3 hours x 1 session, followed by 4 1-hour weekly booster sessions. Delivered by clinicians. | Yes. Parents joined during 2^nd^ hour of the 1^st^ session. Families completed weekly practice logs. | Not applicable |
| 40 | A. J. Russell et al. (2013) | UK | To evaluate adapted CBT for OCD. | Single-blind RCT | Specialist autism, OCD clinics and mental health services | CYP and adults: 46 | 100% ASC diagnosis; 100% OCD diagnosis.  CYP and adults. M age = 26.9 years, 14-65; 76.1% male. CBT: *M* age = 28.6 years (*SD* = 11.3), range 14-49; 82.6% male. Anxiety management: *M* age = 25.2 (*SD* = 13.5), range 14-65; 69.6% male. | Adapted CBT for OCD (n = 23).  Individual; face-to-face; 60 minutes x up to 20 sessions. Delivered by psychologists with prior experience in treating OCD. | No information. | Adapted anxiety management (n = 23) |
| 41 | Storch et al. (2013) | USA | To examine the efficacy of a modular CBT protocol relative to treatment as TAU among autistic children with clinically significant anxiety. | RCT | University based mental health clinic | CYP: 45  Parents: 45 | 100% ASC diagnosis; 100% anxiety disorder diagnosis.  CYP. *M* age = 8.89 years (*SD* = 1.3), 7-11; 80% male; 15.5% non-White. CBT: *M* age = 8.83 years (*SD* = 1.31); 79.2% male; 8.3% non-White. TAU: *M* age = 8.95 years (*SD* = 1.40); 81% male; 23.8% non-White. | Adapted CBT for anxiety (‘BIACA’ programme) (n = 24)  Individual; 60-90 minutes x 16 weekly sessions with modular, flexible format. Child and parent components. Delivered by doctoral or post-doctoral psychology graduates. | Yes. Parent component. Parents also may have been present and/or involved in aspects of the child component. | TAU (n = 21) |
| 42 | Storch et al. (2015) | USA | To examine the efficacy of a personalised, modular CBT protocol among autistic early adolescents with co-occurring anxiety relative to TAU. | RCT | University based multidisciplinary behavioural health clinic specialising in the treatment of pediatric anxiety | CYP: 31  Parents: 31 | 100% ASC diagnosis; 100% anxiety disorder diagnosis.  CYP. *M* age = 12.74 years (*SD* = 1.34), 11–16; 80.6% male; 16.1% non-White. CBT: *M* age = 12.75 years (*SD* = 1.24); 75% male; 25% non-White. TAU: *M* age 12.73 years (*SD* = 1.49); 86.7% male; 6.7% non-White. | Adapted CBT for anxiety (developmentally modified ‘BIACA’ programme) (n = 16)  Individual; 60-90 minutes x 16 weekly sessions with modular, flexible format. Child and parent components. Delivered by doctoral or post-doctoral psychology graduates. | Yes. Parents were included in the majority of sessions including both child and parent components. | TAU (n = 15) |
| 43 | Storch et al. (2020) | USA | To examine the efficacy of family-based FET compared to TAU. | Pilot RCT | Tertiary care clinic specialising in pediatric obsessive compulsive and anxiety disorders | CYP: 32 | 100% ASC diagnosis; 100% anxiety disorder diagnosis.  CYP. *M* = 10.03 years (*SD* = 2.81), 6-17; 81.2% male; 25% non-White. FET: *M* age = 10.07 years (*SD* = 2.89); 71% male; 21.4%. TAU: *M* age = 10 years (*SD* = 2.83); 88.9% male; 27.7% non-White. | Adapted FET for anxiety (n = 14)  Individual; face-to-face; 45-55 minutes x 12 weekly sessions. Delivered by doctoral level clinical psychology students or fellows. | Yes. Parents joined in sessions. | TAU (n = 18) |
| 44 | Wise et al. (2019) | USA | To develop and examine the feasibility of adapted CBT for anxiety. | Before-and-after comparison (open trial) | University-based health clinic specialising in the treatment of anxiety | CYP and adults: 7 | 100% ASC diagnosis; 100% anxiety disorder diagnosis.    CYP and adults. *M* age = 17.14 years (*SD* = 1.68), range 16-20; 57.1% male; 14% non-White. | Adapted modularized CBT for anxiety (TALAA programme).    Individual; face-to-face; 60 minutes x 16 weekly sessions. Offered optional work-readiness program. Delivered by doctoral level students in clinical psychology. | Yes. Parents were encouraged to be involved in sessions, however not compulsory. | Not applicable |
| 45 | Wood et al. (2015) | USA | To evaluate efficacy for treating acute anxiety symptoms in autistic adolescents. | RCT | University clinic and associated autism community clinic | CYP: 33  Parents: 33 | 100% ASC diagnosis; 100% anxiety disorder diagnosis.  CYP. *M* = 12.3 years (*SD* = 1.14), 11-15; 69.7% male; 33.3% non-White. CBT: *M* age = 12.4 years (*SD* = 1.3); 68% male; 37% non-White. Waitlist: *M* age = 12.2 (*SD* = 0.98); 71% male; 28% non-White. | Adapted CBT for anxiety (developmentally modified ‘BIACA’ programme) (n = 19)  Individual; face-to-face; 90 minutes x 16 weekly sessions with modular, flexible format (30 minutes of each session spent with child and parent(s) separately and 30 minutes conjointly). Delivered by clinicians and therapists. | Yes. Parents had separate and conjoint sessions with the child. | Waitlist (n = 14) |
| **Individual and group CBT for anxiety** | | | | | | | | | | |
| 46 | Murphy et al. (2017) | UK | To compare the use of CBT against person-centred counselling for anxiety in autistic young people, ages 12–18. | Pilot RCT | Child and Adolescent Mental Health Services | CYP: 36 | 100% ASC diagnosis; 100% anxiety disorder diagnosis.  CYP. 61.1% male; 5.5% non-White. CBT: *M* age = 14.94, (*SD* = 1.63); 59% male; 6% non-White. Waitlist: *M* age = 15.56 (*SD* = 1.91); 63% male; 5% non-White | Bespoke CBT for anxiety (‘MASSI’ programme) (n = 17).  Individual and group; 12 individual sessions + booster session if needed and five group sessions. Delivered by clinical psychologist or counsellor. | Yes. Parent sessions were offered (no further information). | Person-centred, non-directive counselling (n = 19) |
| 47 | White et al. (2013) | USA | To examine the feasibility of the MASSI programme and preliminary results related to its efficacy. | Pilot RCT | University-affiliated clinic specialising in autism treatment | CYP: 30 | 100% ASC diagnosis; 100% anxiety disorder diagnosis.  CYP. *M* age = 175 (15 years); 77% male; 17.4% non-White. CBT: *M* age = 170 months (*SD* = 14); 73% male; 21% non-White. Waitlist: *M* age = 180 months (*SD* = 15); 80% male; 7% non-White. | Bespoke CBT for anxiety (‘MASSI’ programme) (n = 15)  Individual and group; 60-70 minutes x up to 13 individual sessions and 75 minutes x 7 group sessions. Delivered by therapists. | Yes. Parent education and coaching after each individual therapy session. Parents joined towards the end of individual sessions for approximately 15 minutes. | Waitlist (n = 15) |
| 48 | White et al. (2015) - linked to White et al. (2013) | USA | To examine the course of anxiety and long-term stability of reductions in anxiety, in autistic adolescents who received CBT for anxiety. | Pilot RCT | University-affiliated clinic specialising in autism treatment | CYP: 22 (completers) | 100% ASC; 100% anxiety disorder diagnosis.  CYP. *M* age = 174.05 months (*SD* = 18.66); 72.7% male; 18.1% non-White. | Bespoke CBT for anxiety (‘MASSI’ programme) (n = 11)  Individual and group; 60-70 minutes x up to 13 individual sessions and 75 minutes x 7 group sessions. Delivered by therapists. | Yes. Parent education and coaching after each individual therapy session. Parents joined towards the end of individual sessions for approximately 15 minutes. | Waitlist (n = 11) |
| **Interventions targeting emotional regulation** | | | | | | | | | | |
| 49 | Drusedau et al. (2022) | Germany | To investigate the feasibility, acceptance, and effectiveness of the TüTASS programme. | Before-and-after comparison (pilot study) | Outpatient unit at University Hospital of Psychiatry and Psychotherapy | CYP: 30 | 100% ASC diagnosis; 24% mixed disorders of conduct and emotions, 16% other behavioural and emotional disorders with onset in childhood/adolescence, 8% anxiety disorders, 4% emotional disorders with onset specific to childhood; 4% reaction to severe stress and adjustment disorders.  CYP. *M* age = 10.08 years (*SD* = 1.32), range 7-12; 92% male. | Bespoke mindfulness-based intervention (structured ‘TüTASS’ programme).  Group; face-to-face; 90 minutes x 12 weekly sessions. Delivered by 2 group leaders/therapists. | Yes. Parents were involved in sessions as much as possible. There were also two parent-therapist conferences and one family appointment. | Not applicable |
| 50 | Factor et al. (2019) | USA | To improve upon the statistical power of the initial pilot study to provide more convincing evidence of the efficacy of STAMP. | RCT | Autism clinic | CYP: 23  Parents: 23 | 100% ASC diagnosis; 100% difficulties with managing anger or anxiety.  CYP. *M* age = 5.46 (*SD* = 1.01), 4-7; 82.6% male; 0% non-White. CBT: *M* age = 5.54 years (*SD* = 0.94); 75% male; 0 non-White. Waitlist: *M* age = 5.36 years (*SD* = 1.12); 90.9% male; 0% non-White. | Adapted CBT for emotion regulation (‘STAMP’ programme) for younger children (4-7 years) (n = 12).  Group; 1 hour x 9 weekly sessions. | Yes. Parents attended group sessions. They also watched the child sessions on a monitor. | Delayed therapy (n = 11) |
| 51 | Scarpa et al. (2011) | USA | To test the efficacy of a developmentally modified CBT for autistic young children to teach emotion regulation strategies for reducing anger and anxiety. | Pilot RCT | Autism clinic | CYP: 11  Parents: 11 | 100% ASC diagnosis.  CYP. Age range 5-7 years; 81.8% male. | Adapted CBT for emotion regulation (n = 5)  Group; 1 hour x 9 weekly sessions. Delivered by clinical graduate students and trained staff. | Yes. 9 parent group sessions took place. Parents were also able to watch the children’s sessions on a monitor. | Delayed therapy (n = 6) |
| 52 | Sofronoff et al. (2017) | Australia | To investigate the usefulness of a self-directed multi-component social and emotional skills training programme for autistic children with weekly therapist support to parents provided via Skype. | Before-and-after comparison | University clinic | CYP: 41  Parents: 38^a^ | 100% ASC diagnosis; 9.5% anxiety, 2.4% depression; ADHD 17.5%, learning difficulties 4.9%.  *M* age = 9.56 years, range 7 years and 11 months-12; 87.8% male. | Adapted cognitive behavioural emotional and social skills intervention (‘SAS’ programme).  Group; 90 minutes x 10 weekly sessions. Delivered by parents with weekly therapist support provided via Skype (lasting 30-60 minutes). | Yes. Parents delivered the intervention. | Not applicable |
| 53 | Swain et al. (2019) | USA | To examine response to group CBT in terms of individual-level change in autistic young children. | Before-and-after comparison study | University associated community clinic and a hospital | CYP: 18 | 100% ASC diagnosis.  CYP. *M* age = 73.89 months (*SD* = 11.87), range 53-90; 88.8% male; 0% non-White. | Adapted CBT for young children for emotion regulation (aged 4-8) (‘STAMP’ programme)  Group; 60 minutes x 9 weekly sessions. Delivered by clinicians. | Yes. Concurrent parent groups. | Not applicable |
| **CBT for various mental health needs** | | | | | | | | | | |
| 54 | Cooper et al. (2018) | UK | To investigate therapists’ knowledge and experience of working within a CBT framework with autistic people. | Cross-sectional survey | IAPT and secondary mental health services | Staff: 50 | Therapists working with autistic people across the lifespan. | Adapted CBT.  Individual. Delivered by psychological therapists. | Yes. 48% of therapists involved family member in sessions. | Not applicable |
| 55 | McGillivray et al. (2014) | Australia | To test CBT whether reduced negative and anxious thinking patterns and symptoms of stress, anxiety and depression. | Non-randomised controlled trial | Disability service agency | CYP and adults: 42 | 100% ASC diagnosis; 100% symptoms of depressed mood, anxiety, stress and/or negative automatic thoughts.  CYP and adults. *M* age = 20.6 years (*SD* = 4.1), 15-25; 76.2% male. CBT: *M* age = 20.27 years (*SD* = 4.39); 73.1% male. Waitlist: *M* age = 20.50 (*SD* = 3.4); 81.3% male. | Bespoke CBT for anxiety, stress and depression (‘think well, feel well and be well’ programme) (n = 26).  Group; face-to-face; 2 hours x 9 weekly sessions. Delivered by practitioners who were experts in delivering CBT. | No information. | Waitlist (n = 16) |
| 56 | Santomauro et al. (2016) | Australia | To evaluate the feasibility, acceptability and preliminary efficacy of a cognitive behavioural intervention for depression in autistic adolescents. | Pilot RCT | University clinic | CYP: 20 (completers) | 100% ASC diagnosis; 100% depressive symptoms.  CYP. *M* age = 15.75 years (*SD* = 1.37); 60% male. CBT *M* age = 16 years (*SD* = 1.33). Waitlist: *M* age = 15.5 years (*SD* = 1.43). | Bespoke CBT for depression (‘Exploring depression’ programme) (n = 11)  Group; 1 hour x 10 weekly sessions + booster session 4 weeks later. Delivered by 2 clinical psychologists. | Can’t tell. If an adolescent missed a session, then they had the opportunity to attend a one-on-one catch-up session with the psychologists along with their parent. | Waitlist (n = 12) |
| **EMDR for PTSD** | | | | | | | | | | |
| 57 | Fisher et al. (2023) | Netherlands; UK | To develop therapist consensus about adaptations to EMDR that are important when working with autistic people. | Delphi survey (3 rounds) | Psychological therapies, community mental health, ID, forensic and tertiary services, independent practice, education, military, voluntary organisations | Staff: 103 | EMDR therapists working with autistic people across the lifespan. | Adapted EMDR.  Delivered by trained therapists. | Can’t tell. 86% of therapists obtained information from other people. | Not applicable |

*Note*. Where participant characteristics are not listed in the table this means that they were not reported in the paper. **ADHD** = Attention Deficit Hyperactivity Disorder, **ADOS** = Autism Diagnostic Observation Schedule, **ASC** = Autism Spectrum Conditions, **ASD-CP** = Autism Spectrum Disorder Care Pathway, **AUP** = Pathway Autism Intellectual Disability and Psychiatric Disorder, **BIACA** = Behavioral Interventions for Anxiety in Children with Autism, **CBT** = Cognitive Behavioural Therapy, **CYP** = Children and Young People, **DAWBA** = Development and Well-Being Assessment, **ECHO** = Extension for Community Healthcare Outcomes, **EMDR** = Eye Movement Desensitisation and Reprocessing, **FET** = Family-based exposure-focused treatment, **FYF** = Facing Your Fears, **IAPT** = Improving Access to Psychological Therapies, **ID** = Intellectual Disability, **MASSI** = Multimodal Anxiety and Social Skills Intervention, **OCD** = Obsessive Compulsive Disorder, **OST** = One-Session Treatment, **RCT** = Randomised Controlled Trial, **SAS** = Secret Agent Society, **SCQ** = Social Communication Questionnaire, **STAMP** = Stress and Anger Management Programme, **TALAA** = Treatment of Anxiety in Late Adolescents with Autism, **TAU** = Treatment as Usual, **TüTASS** = Tübinger Training for Autism Spectrum Disorders.

^a^ Three parents had two children in the study.

**Table S7.** Mixed Methods Appraisal Tool (MMAT) quality assessment

Sorted by category of study designs.

| **Authors** | **Category of study designs** | **Criterion 1** | **Criterion 2** | **Criterion 3** | **Criterion 4** | **Criterion 5** | **Total** |
| --- | --- | --- | --- | --- | --- | --- | --- |
| Petty et al. (2021) | Qualitative study | Yes | Yes | Yes | Yes | Yes | 5 |
| Spain et al. (2017) | Qualitative study | Yes | Yes | Yes | Yes | Yes | 5 |
| Chalfant et al. (2007) | Randomised controlled trials | Yes | Yes | Yes | No | Yes | 4 |
| Cook et al. (2017) | Randomised controlled trials | Yes | Yes | No | Can't tell | Can't tell | 2 |
| Factor et al. (2019) | Randomised controlled trials | Can't tell | Yes | Can't tell | No | Can't tell | 1 |
| Fujii et al. (2013) | Randomised controlled trials | Can't tell | Yes | Yes | Yes | Yes | 4 |
| Kilburn et al. (2020) | Randomised controlled trials | Yes | Yes | Yes | Yes | Yes | 5 |
| Maskey, Rodgers et al. (2019) | Randomised controlled trials | Yes | Yes | Yes | Yes | Yes | 5 |
| Murphy et al. (2017) | Randomised controlled trials | Can't tell | Yes | Can't tell | Yes | Yes | 3 |
| J. Reaven, Blakeley-Smith, Culhane-Shelburne et al. (2012) | Randomised controlled trials | Can't tell | Yes | Yes | Yes | Yes | 4 |
| Reaven et al. (2018) | Randomised controlled trials | Can't tell | Yes | Yes | No | Yes | 3 |
| A. J. Russell et al. (2013) | Randomised controlled trials | Yes | Yes | Yes | Yes | Yes | 5 |
| Santomauro et al. (2016) | Randomised controlled trials | Can't tell | Yes | Yes | No | No | 2 |
| Scarpa et al. (2011) | Randomised controlled trials | Can't tell | Yes | Yes | No | Yes | 3 |
| Sofronoff et al. (2005) | Randomised controlled trials | Can't tell | Yes | Yes | Can't tell | Yes | 3 |
| Storch et al. (2013) | Randomised controlled trials | Yes | Yes | Yes | Yes | Yes | 5 |
| Storch et al. (2015) | Randomised controlled trials | Yes | Yes | Yes | Yes | Yes | 5 |
| Storch et al. (2020) | Randomised controlled trials | Yes | Yes | No | Yes | Yes | 4 |
| Sung et al. (2011) | Randomised controlled trials | Yes | Yes | Yes | Yes | Yes | 5 |
| Walsh et al. (2018) | Randomised controlled trials | Can't tell | Yes | Yes | No | Yes | 3 |
| White et al. (2013) | Randomised controlled trials | Yes | Yes | Yes | Yes | No | 4 |
| White et al. (2015) | Randomised controlled trials | Yes | Yes | Yes | Yes | No | 4 |
| Wood et al. (2015) | Randomised controlled trials | Yes | Yes | Yes | Yes | Yes | 5 |
| Bemmer et al. (2021) | Non-randomised study | Can't tell | Yes | Yes | Yes | Yes | 4 |
| Cervantes et al. (2019) | Non-randomised study | Yes | Yes | Can't tell | Yes | Can't tell | 3 |
| Dreiling et al. (2022) | Non-randomised study | Can't tell | Can't tell | No | No | Yes | 1 |
| Driscoll et al. (2020) | Non-randomised study | Can't tell | Yes | Yes | No | Yes | 3 |
| Drusedau et al. (2022) | Non-randomised study | Can't tell | Yes | Yes | No | Yes | 3 |
| Ehrenreich-May et al. (2014) | Non-randomised study | Can't tell | Yes | Yes | No | Yes | 3 |
| Ekman et al. (2015) | Non-randomised study | Can't tell | No | Can't tell | No | Can't tell | 0 |
| Helverschou et al. (2021) | Non-randomised study | Can't tell | Yes | Yes | Can't tell | Yes | 3 |
| Hepburn et al. (2016) | Non-randomised study | Can't tell | Yes | Yes | Yes | Yes | 4 |
| Keefer et al. (2017) | Non-randomised study | Yes | Yes | No | No | Yes | 3 |
| Kilburn et al. (2019) | Non-randomised study | Can't tell | Yes | Yes | No | Yes | 3 |
| Kuriakose et al. (2018) | Non-randomised study | Yes | Yes | Can't tell | Yes | Can't tell | 3 |
| Maskey, McConachie, et al. (2019) | Non-randomised study | No | Yes | Yes | No | Yes | 3 |
| McGillivray et al. (2014) | Non-randomised study | Can't tell | Yes | Yes | Yes | Yes | 4 |
| Oerbeck et al. (2021) | Non-randomised study | Can't tell | Yes | Yes | No | Yes | 3 |
| Ollendick et al. (2021) | Non-randomised study | No | Yes | Yes | Can't tell | Yes | 3 |
| Reaven et al. (2009) | Non-randomised study | No | Yes | Yes | No | Yes | 3 |
| J. Reaven, Blakeley-Smith, Leuthe et al. (2012) | Non-randomised study | No | Yes | Yes | Can't tell | Yes | 3 |
| Reaven et al. (2015) | Non-randomised study | No | Yes | Yes | No | Yes | 3 |
| Sofronoff et al. (2017) | Non-randomised study | Yes | Yes | No | Yes | Yes | 4 |
| Solish et al. (2020) | Non-randomised study | No | Yes | Yes | Yes | Yes | 4 |
| Swain et al. (2019) | Non-randomised study | Yes | Yes | Yes | Can't tell | Yes | 4 |
| Wise et al. (2019) | Non-randomised study | No | Yes | Yes | No | Yes | 3 |
| Cooper et al. (2018) | Quantitative descriptive study | Yes | Yes | Can't tell | Yes | Yes | 4 |
| Ford et al. (2019) | Quantitative descriptive study | Yes | Can't tell | Yes | Can't tell | Yes | 3 |
| Hollocks et al. (2019) | Quantitative descriptive study | Yes | Yes | Yes | Can't tell | Yes | 4 |
| Jones et al. (2021) | Quantitative descriptive study | Yes | No | Can't tell | Can't tell | Yes | 2 |
| Stadnick et al. (2015) | Quantitative descriptive study | Yes | Yes | Yes | Can't tell | Yes | 4 |
| Burke et al. (2017) | Mixed methods study | No | No | No | Yes | No | 1 |
| Fisher et al. (2022) | Mixed methods study | Yes | Yes | Yes | Yes | Can't tell | 4 |
| E. Higgins et al. (2019) | Mixed methods study | Yes | Yes | Yes | Yes | Can't tell | 4 |
| Langdon et al. (2016) | Mixed methods study | Yes | Yes | Yes | Yes | Yes | 5 |
| McConachie et al. (2014) | Mixed methods study | Yes | Yes | Yes | Yes | Yes | 5 |
| Pickard et al. (2020) | Mixed methods study | Yes | Yes | Yes | Yes | Yes | 5 |

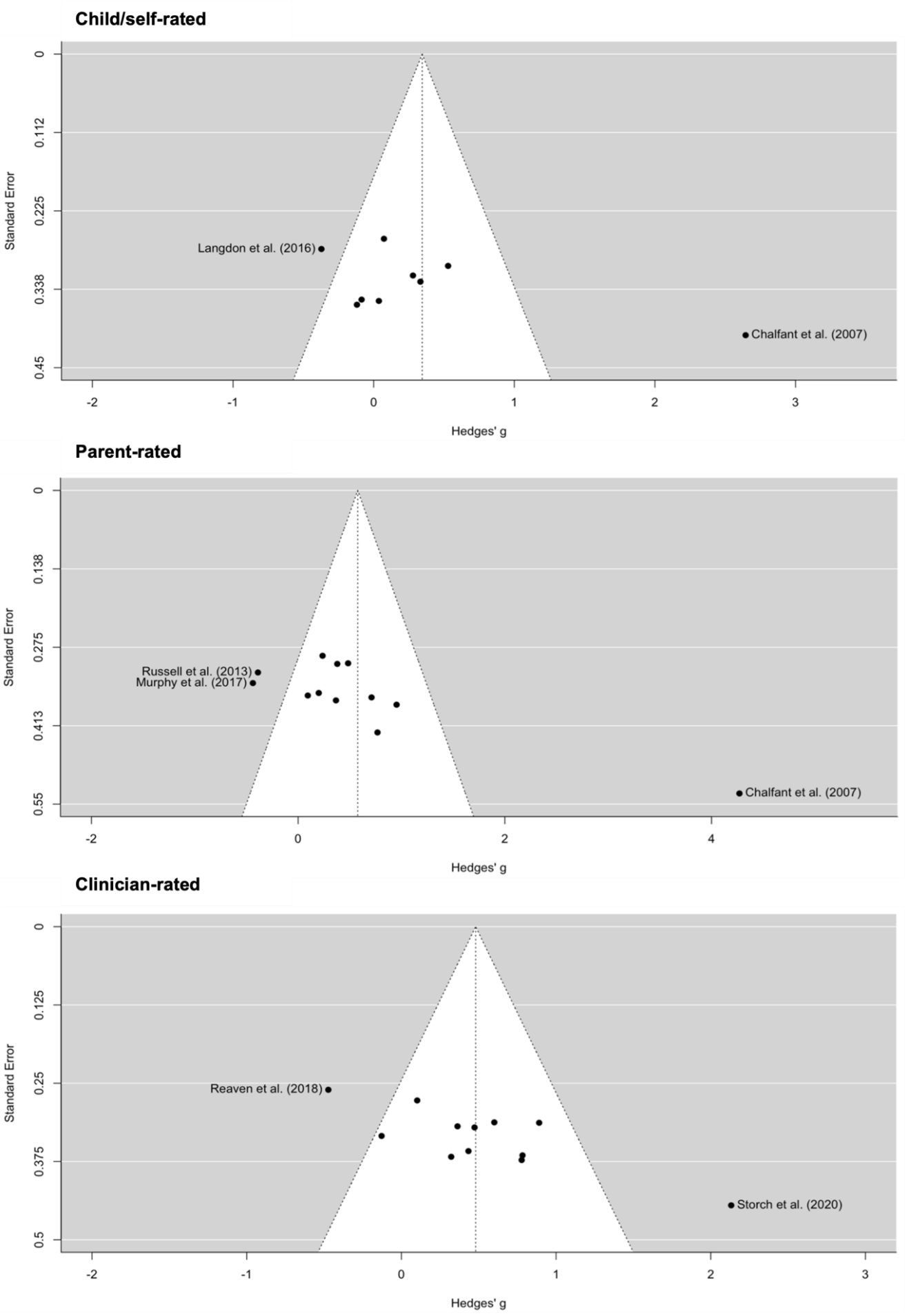
**Figure S1.** Funnel plots for child/self-rated anxiety, parent- and clinician-rated child and young person’s anxiety

**Table S8.** Autism-Inclusive Research Assessment (AIRA)

| **Author (Year)** | **Any reported involvement from people with lived experience in the design, conduct, or writing up of the articles?** | **For articles with qualitative elements, were adjustments made to the data collection process to facilitate wide participation e.g., allowing non-verbal/non-oral communication for interviews?** | **For articles with quantitative elements, were adjustments made to the data collection tools to facilitate wide participation e.g., adapting Likert scales for greater precision, straightforward language, defining key terms?** | **For articles with quantitative elements, were any of the relevant outcome measures to the review adapted (or reported to have been validated) for autistic people (e.g., measure of autistic quality of life)?** | **For articles with quantitative elements, did the intervention/strategy involve any focus (not just related to the relevant measures to the review) on getting people to mask/change autistic behaviours?** |
| --- | --- | --- | --- | --- | --- |
| Bemmer et al. (2021) | None reported | None reported. | None reported | Yes: some measures were reported as previously validated in research with autistic people: "DASS-21 is a self-report measure of depression, anxiety and stress, and assesses symptom severity over the past week, and has recently been validated for use in ASC populations". Other measures were reported as previously used with autistic people: "The LSAS is one of the most commonly used measures of social anxiety in adult ASC populations"; "K10 has been used in similar studies to measure overall symptoms of distress, rather than disorder-specific (anxiety/depression) symptoms in autistic adults"; "The SIAS and SPS are partner measures used to assess social anxiety and have previously been used to measure social anxiety levels in ASC populations". | Yes: The primary outcome measure was the Social Responsiveness Scale, which assessed social skill functioning and autistic symptoms in adults and measures a reduction in autistic behaviours as a positive outcome. Therefore, reducing autistic traits and potentially getting autistic people to mask was a focus of the study. |
| Burke et al. (2017) | None reported | Yes: As stated in paper, the childrens evaluation forms provided qualitative data and "Children’s evaluation forms were presented with an adapted Likert-based rating scale using visuals." | Yes: As stated in paper, regarding the Children Beck Youth Anxiety Self-report Inventory "To facilitate understanding and accuracy of rating, a child-friendly explanation of the concept of frequency terms: “never, sometimes, often or always” accompanied each scale, in addition to a structured description of how to complete each scale." | None reported | No: intervention aimed to build skills and understanding rather than encourage masking. As an aside, the use of functioning labels is really uncomfortable, especially given that the paper was published after the move away from functioning labels. |
| Cervantes et al. (2019) | None reported | Not applicable - no qualitative element | None reported | None reported | No: The intervention was around adaptations to the environment and to ways of working rather than on the young people having to mask or change. |
| Chalfant et al. (2007) | None reported | Not applicable - no qualitative element | None reported | None reported | No: The intervention is about managing anxiety and there is a heavy focus on relaxation strategies. No behaviours that could be related to masking were measured as positive outcomes. |
| Cook et al. (2017) | None reported | Not applicable - no qualitative element | None reported- only parents involved, no reporting of whether these parents were autistic. | None reported | No: There was potential for the intervention to encourage masking depending on what goals were considered on the 'hierarchy of fears'. However, the intervention encouraged parents to work with their child on this and there was a lot of education around autistic needs. It was internalising rather than externalising behaviours that were reduced, and that reduction reported as a positive outcome, which might even suggest less masking. |
| Cooper at al. (2018) | None reported | Not applicable. | None reported. Data were collected from providers, no reporting of whether these providers were autistic. | None reported. Data were collected from providers, no reporting of whether these providers were autistic. | Not applicable. |
| Dreiling et al. (2022) | Yes: The multidisciplinary expert panel (“hub team”) who facilitated the Project ECHO included one regional parent advocate to share a parent perspective with participating providers. In each cohort, the parent advocate was recruited from the cohort’s local region to provide information about local resources. Additionally, the specific didactic topics (in the Project ECHO curriculum) were chosen based on feedback from a series of focus groups with rural families who provided input on the mental health needs of their autistic child. | None reported. Data were collected from providers, no reporting of whether these providers were autistic. | None reported. Data were collected from providers, no reporting of whether these providers were autistic. | None reported. Data were collected from providers, no reporting of whether these providers were autistic. | Not applicable. |
| Driscoll et al. (2020) | None reported | Not applicable - no qualitative element | None reported | Yes - the study explicitly stated that PARS has demonstrated validity for autistic people | Yes: Exposure exercises for managing social anxiety included rewarding increased eye contact, which could encourage masking. 'Sensory fears' were also a target of treatment, which could have led to children learning to mask their discomfort when they are exposed to sensory environments they find overwhelming. |
| Drusedau et al. (2022) | None reported | Not applicable - no qualitative element | None reported | None reported | No: Although the intervention itself did not directly encourage masking, the goal of 'improving negative behaviours' means that masking is likely to have been picked up as a positive. |
| Ehrenreich-May et al. (2014) | None reported | Not applicable - no qualitative element | None reported | None reported | No: There was potential for the intervention to encourage masking depending on what goals were considered on the 'hiararchy of fears'. However, the intervention encouraged parents to work with their child on this and there was a lot of education around autistic needs. It was internalising rather than externalising behaviours that were reduced and that reduction reported as a positive outcome, which might even suggest less masking. |
| Ekman et al. (2015) | None reported | Not applicable. | None reported. | None reported | Yes: The study's results showed improvement in psychological, social and occupational functioning ability on the GAF. The GAF scale prompts include consideration of 'meaningful social relationships' which leaves it open to neurotypical bias and to an encouragement of masking or changing autistic behaviours because increased social interaction is then seen as a positive. In some cases, increased social interaction may not be what the autistic person wants and may not be meaningful to them but may be a result of masking behaviour. |
| Factor et al. (2019) | None reported | Not applicable - no qualitative element | None reported | None reported | No: intervention aimed to build skills and understanding rather than encourage masking. |
| Fisher et al. (2023) | Yes: study reported seeking informal feedback about the scope and aims of the first survey, categories of questions included and barriers to EMDR from one autistic adult. Additionally, the study reported informally discussing the study findings and interpretation with two autistic adults and a parent/carer of an autistic child. Participants also included autistic therapists. | None reported. Data were collected from providers, and participants included autistic therapists. | None reported. Data were collected from therapists, including autistic therapists | None reported. Data were collected from therapists, including autistic therapists. | Not applicable. |
| Ford et al. (2019) | None reported | Not applicable - no qualitative element | None reported | None reported | No |
| Fujii et al. (2013) | None reported | Not applicable - no qualitative element | None reported | None reported | Yes - Goals included being able to 'keep it in' in school (referring to emotional meltdowns) the reduction in meltdowns in class was measured as a positive outcome. That suggests a focus on masking autistic behaviours without consideration of the external sensory factors that may be influencing them. |
| Helverschou et al. (2021) | None reported | Not applicable. | None reported. Measures completed by caregivers. | Yes: the study reported the PAC to be a specific screening checklist for identification of individuals with autism and intellectual disability in need of psychiatric services, and a psychiatric instrument developed specifically for autistic individuals that has been found to discriminate reliably between psychiatric symptoms and the core autistic characteristics. The study reported that PAC has also been found to distinguish between autistic adults and those with intellectual disabilities with and without psychiatric conditions, and to a certain extent between people with different psychiatric conditions, especially psychosis and obsessive-compulsive disorder. | Yes: The use of the ABC focuses on behaviour that is challenging to people around the autistic person and measures a reduction in these behaviours as a positive outcome. Therefore, reducing autistic traits and potentially getting autistic people to mask was a focus of the study. Additionally, the significant correlation between PAC and ABC suggests that reducing 'challenging' behaviour may lead to change the response of others around the person, and may thereby reduce feelings linked to depression/anxiety, indicating potential masking. |
| Hepburn et al. (2016) | None reported | Not applicable - no qualitative element | None reported | Yes: This paper used SCARED and explicitly stated that The SCARED has been documented to be a “potentially appropriate” for in use in studies of psychosocial intervention with youth with ASD. | No: Focus was on anxiety symptoms. As with previous studies there is potential for masking to be encouraged depending on the 'fears' selected and whether these are chosen by the child or the parent and whether these relate to autism-related difficulties such as sensory needs. |
| Higging et al. (2019) | None reported | None reported - only parents interviewed, no indication if parents were autistic | None reported | None reported | No: Focus was on anxiety symptoms. As with previous studies there is potential for masking to be encouraged depending on the 'fears' selected and whether these are chosen by the child or the parent and whether these relate to autism-related difficulties such as sensory needs. |
| Hollocks et al. (2019) | None reported | Not applicable - no qualitative element | None reported | None reported | No: focus on detection of autism not intervention |
| Jones et al. (2021) | None reported | None reported. Data were collected from providers, no reporting of whether these providers were autistic. | None reported. | None reported. Data were collected from providers, no reporting of whether these providers were autistic. | Not applicable. |
| Keefer et al. (2017) | None reported | Not applicable - no qualitative element | Yes - "For the present study, the IUS-C was modified to support child comprehension and to accommodate common language difficulties in children with ASD. As such, specific modifications were made to reduce ambiguity as well as complexity of syntax and grammar. Some of the modifications included re-wording items from passive to active voice (e.g., changed ‘‘Surprise events upset me greatly.’’ to ‘‘I don’t like to be surprised by new plans or activities.’’), and consistently using first person pronouns to reduce abstraction and increase personalization (e.g., changed ‘‘One should always think ahead to avoid surprises.’’ to ‘‘I always try to think ahead in order to avoid surprises.’’). Additionally, the original 5-point Likert scale was reduced to 4 points to minimize complexity" | Yes - "Internal consistency for both parent and child report has been found to be adequate in ASD samples" | No |
| Kilburn et al. (2019) | None reported | Not applicable - no qualitative element | None reported | None reported | No |
| Kilburn et al. (2020) | None reported | Not applicable - no qualitative element | None reported | None reported | No: Focus was on anxiety symptoms. As with previous studies there is potential for masking to be encouraged depending on the 'fears' selected and whether these are chosen by the child or the parent and whether these relate to autism-related difficulties such as sensory needs. |
| Kuriakose et al. (2018) | None reported | Not applicable - no qualitative element | None reported | None reported | No: The intervention was around adaptations to the environment and to ways of working rather than on the young people having to mask or change. |
| Langdon et al. (2016) | None reported | None reported. | None reported. | None reported | No: There is no evidence to suggest a focus on masking or autism-related outcomes specifically. |
| Maskey, Rodgers et al. (2019) | None reported | Not applicable - no qualitative element | None reported | Yes: Paper used the SCQ and stated "It is used internationally, and has high sensitivity and specificity for an ASD diagnosis." Also, used Spence Children’s Anxiety Scale-parent version (SCAS-P) and child version (SCAS-C) and explicitly stated "The measure has been widely used in ASD studies. High internal consistency for the total scale score has been reported and both convergent and divergent validity." | No: phobias were the target of intervention. Reduction in autistic traits was not measured or seen as a positive outcome |
| Maskey, McConachie, et al. (2019) | None reported | Not applicable - no qualitative element | None reported | None reported | No: phobias were the target of intervention. Reduction in autistic traits was not measured or seen as a positive outcome |
| McConachie et al. (2014) | None reported | None reported | None reported | None reported | No: Focus was on anxiety symptoms. As with previous studies there is potential for masking to be encouraged depending on the 'fears' selected and whether these are chosen by the child or the parent and whether these relate to autism-related difficulties such as sensory needs. |
| McGillivray et al. (2014) | None reported | Not applicable. | None reported | None reported | No: the intervention did not focus on getting people to mask or change autistic behaviours and was focused on what worked for autistic people. The DASS-41 measure in particular was good because it measured physical responses to anxiety rather than behaviours, reducing the potential of measuring masking behaviours as a positive outcome. |
| Murphy et al. (2017) | None reported | Not applicable - no qualitative element | None reported | Yes: The paper explicity it states it uses the CASI-anx which "has been developed specifically to measure anxiety in children with ASD" | Yes - Improvements in the Social Responsiveness Scale Score were considered positive outcomes. It is unclear what the social skills training entailed, but it may have involved masking of autistic traits such as increasing eye contact and increasing social contact and learning 'socially acceptable' responses and body language. |
| Oerbeck et al. (2021) | None reported | Not applicable - no qualitative element | None reported | None reported | No: Focus was on anxiety symptoms. As with previous studies there is potential for masking to be encouraged depending on the 'fears' selected and whether these are chosen by the child or the parent and whether these relate to autism-related difficulties such as sensory needs. |
| Ollendick et al. (2021) | None reported | Not applicable - no qualitative element | None reported | None reported | No: Focus was on anxiety symptoms. As with previous studies there is potential for masking to be encouraged depending on the 'fears' selected and whether these are chosen by the child or the parent and whether these relate to autism-related difficulties such as sensory needs. |
| Petty et al. (2021) | Yes: The study reported that all study materials were revised by people with lived experience of autism, including the outcome measures, the design of the study and the interpretation of the findings. | None reported. Data were collected from providers, no reporting of whether these providers were autistic. | Not applicable. | Not applicable. | Not applicable. |
| Pickard et al. (2020) | None reported | Not applicable - no qualitative element | None reported - clinicians only, no reporting of whether these clinicians were autistic. | None reported - clinicians only, no reporting of whether these clinicians were autistic. | Not applicable - clinicians only |
| Reaven et al. (2009) | None reported | Not applicable - no qualitative element | None reported | None reported | No: Focus was on anxiety symptoms. As with previous studies there is potential for masking to be encouraged depending on the 'fears' selected and whether these are chosen by the child or the parent and whether these relate to autism-related difficulties such as sensory needs. |
| J. Reaven, Blakeley-Smith, Culhane-Shelburne et al. (2012) | None reported | Not applicable - no qualitative element | None reported | None reported | No: Focus was on anxiety symptoms. As with previous studies there is potential for masking to be encouraged depending on the 'fears' selected and whether these are chosen by the child or the parent and whether these relate to autism-related difficulties such as sensory needs. |
| J. Reaven, Blakeley-Smith, Leuthe et al. (2012) | None reported | Not applicable - no qualitative element | None reported | None reported | No: Focus was on anxiety symptoms. As with previous studies there is potential for masking to be encouraged depending on the 'fears' selected and whether these are chosen by the child or the parent and whether these relate to autism-related difficulties such as sensory needs. |
| Reaven et al. (2015) | None reported | Not applicable - no qualitative element | None reported | Yes - "Youth with ASD tend to under-report anxiety symptoms and, as such, clinicians tend to rely more upon parent report of symptoms on the ADIS than on child report. Thus, only the parent version was used in this study." | No: Focus was on anxiety symptoms. As with previous studies there is potential for masking to be encouraged depending on the 'fears' selected and whether these are chosen by the child or the parent and whether these relate to autism-related difficulties such as sensory needs. |
| Reaven et al. (2018) | None reported | Not applicable - no qualitative element | None reported | None reported | No: Focus was on anxiety symptoms. As with previous studies there is potential for masking to be encouraged depending on the 'fears' selected and whether these are chosen by the child or the parent and whether these relate to autism-related difficulties such as sensory needs. |
| A. J. Russell et al. (2013) | None reported | Not applicable. | Yes: as described - "at the start of each clinical interview, care was taken to ensure that the participant was cognisant of the phenomena to be rated, that the discomfort and anxiety basis for each potential [obsessive-compulsive] symptom was clearly established using visual tools if necessary. Eliciting of symptoms was achieved if needed by enquiring about daily routines in total before gathering further phenomenological information. Communication style and preferences of each individual were also taken into account when administering the [primary outcome measure]." | None reported | No: Communication style and preference were taken into account when administering the YBOCS symptom checklist for OCD symptoms, suggesting that the potential for masking behaviours to be seen as a positive outcome was minimised. |
| Santomauro et al. (2016) | None reported | Not applicable - no qualitative element | None reported | None reported | No - focus was on symptoms of depression |
| Scarpa et al. (2011) | None reported | Not applicable - no qualitative element | None reported | None reported | Unclear - It's unclear what is taught particularly in the social skills element. Measuring reduced 'outbursts' could potentially indicate children who have learned to mask their autistic behaviours and instead hold in their emotional responses. |
| Sofronoff et al. (2005) | None reported | Not applicable - no qualitative element | None reported | None reported | No |
| Sofronoff et al. (2017) | None reported | Not applicable - no qualitative element | None reported - only parents involved, no reporting of whether these parents were autistic. | None reported | No |
| Solish et al. (2020) | Yes: Questionnaire item development was informed by knowledge on parenting autistic children | Not applicable - no qualitative element | None reported | Yes - "Group Questionnaires, Parent and Child versions. Developed by authors to be used pre- and post-intervention. Includes quantitative ratings (e.g., levels of anxiety and its interference), and qualitative responses (e.g., strategies a parent is currently using). Item development was informed by knowledge of anxiety and ASD and on parenting children with ASD" | No: Focus was on anxiety symptoms. As with previous studies there is potential for masking to be encouraged depending on the 'fears' selected and whether these are chosen by the child or the parent and whether these relate to autism-related difficulties such as sensory needs. |
| Spain et al. (2017) | None reported | None reported. Data were collected from providers, no reporting of whether these providers were autistic. | Not applicable - no quantitative element | Not applicable - no quantitative element | Not applicable - no quantitative element |
| Stadnick et al. (2015) | None reported | Not applicable - no qualitative element | Yes - "Children were administered one of the modules based on their language and developmental level" | None reported. | No - relates to diagnosis and not intervention |
| Storch et al. (2013) | None reported | Not applicable - no qualitative element | None reported | None reported | Yes: the Social Responsiveness Scale was used as an outcome measure for reduction in autistic traits around social communication and repetitive behaviours. This means that masking of autistic behaviours would have been noted as a positive outcome. |
| Storch et al. (2015) | None reported | Not applicable - no qualitative element | None reported | Yes - "The PARS has excellent psychometric properties in youth with ASD and anxiety" | Yes: the Social Responsiveness Scale was used as an outcome measure for reduction in autistic traits around social communication and repetitive behaviours. This means that masking of autistic behaviours would have been noted as a positive outcome. |
| Storch et al. (2020) | None reported | Not applicable - no qualitative element | None reported | None reported | Yes: The Social Responsiveness Scale was used as an outcome measure with reduction of autistic traits being seen as a positive. That said, none of the modules of the intervention were aimed at targetting core autistic traits so the intervention itself did not focus on getting autistic people to mask. |
| Sung et al. (2011) | None reported | Not applicable - no qualitative element | None reported | None reported | No: focus was on reducing anxiety symptoms. Social skills work described did not aim to change autistic traits |
| Swain et al. (2019) | None reported | Not applicable - no qualitative element | None reported | None reported | Yes: The Social Responsiveness Scale was used as an outcome measure for reduction in autistic traits around social communication and repetitive behaviours. This means that masking of autistic behaviours would have been noted as a positive outcome. |
| Walsh et al. (2018) | None reported | Not applicable - no qualitative element | Yes - "Youth used a 5-point pictorial scale to rate the helpfulness of each activity. A rating of “1” was paired with a sad face and a rating of “5” was paired with a happy face." | Yes - "Results from a recent study confirm the 41-item measures five-factor structure and suggest good sensitivity (.71) and specificity (.67) among parents of youth with ASD (Stern et al. 2014)" | No: focus was on anxiety symptoms and acceptability of the approach. |
| White et al. (2013) | None reported | Not applicable - no qualitative element | None reported | Yes - "Although it has not been widely used as an outcome measure, there is emerging evidence that it is sensitive to change with treatment of people with ASD" and "Similar to modifications made by Leyfer et al. (2006), clinicians administering the ADIS-C/P were mindful of the symptom overlap between ASD and anxiety disorders. For instance, avoidance of social situations was coded as an anxiety symptom only if it was related to fear of evaluation rather than disinterest in social situations." | Yes: The social responsiveness scale was used as an outcome measure for reduction in autistic traits around social communication and repetitive behaviours. This means that masking of autistic behaviours would have been noted as a positive outcome. |
| White et al. (2015) | None reported | Not applicable - no qualitative element | None reported | None reported | No: Although the Social Responsiveness Scale was used, this was to measure autistic traits and degree of neurotypical percieved social difficulties to establish whether this is a factor in improvement in anxiety symptoms after CBT. |
| Wise et al. (2019) | None reported | Not applicable. | None reported. | None reported | Unclear: It is not clear exactly what the CBT protocol entailed specifically or how much of the exposure work was lead by the autistic persons wants. Therefore, it is possible that some of the exposure work required the autistic person to learn to mask and that some of the thought challenging was invalidating the autistic person's experiences. Interestingly, the clinician rated scales showed a significant reduction in anxiety while the self-rated scales did not. This was explained with 'lack of insight' on the part of autistic participants, rather than considering the effects of the autistic people masking traits. |
| Wood et al. (2015) | None reported | Not applicable - no qualitative element | None reported | Yes - "The MASC-P has previously been found to have robust psychometric properties, including in ASD samples" and "Psychometric properties of the PARS Total Score in ASD samples are acceptable; the adolescents in this study were part of a larger study of the PARS in youth with ASD in which interrater reliability and test-retest reliability for the PARS Total Score were good (ICC = 0.86 and 0.83, respectively)." | Yes: The Social Responsiveness Scale was used as an outcome measure for reduction in autistic traits around social communication and repetitive behaviours. This means that masking of autistic behaviours would have been noted as a positive outcome. |

*Note.* **ABC** = Aberrant Behaviour Checklist, **ADIS-C** = Adapted Anxiety Disorders Interview Schedule-Children, **ASD** = Autism Spectrum Disorder, **CASI-Anx**=Child and Adolescent symptom inventory-4 ASD anxiety scale, **CBT** = Cognitive Behavioural Therapy, **ECHO** = Extension for community healthcare outcomes, **EMDR** = Eye movement desensitisation and reprocessing, **DASS** = Depression Anxiety Stress Scale, **GAF** = Global Functioning Rating scale, IUS-C = Modified intolerance of uncertainty scale child-report, **K10** = Kessler psychological distress scale, **LSAS** = Liebowitz Social Anxiety Scale, **MASC-P** = Multidimensional anxiety scale for children-parent version, **OCD** = Obsessive Compulsive Disorder, **PAC** = Psychopathology in Autism Checklist, **PARS** = Paediatric Anxiety Rating Scale, **SCARED** = Screen for Anxiety and Related Emotional Disorders in Children, **SCAS – C/P**= Spence Children’s Anxiety Scale-Child/-Parent, **SCQ** = Social Communication Questionnaire, **SIAS** = Social Interaction Anxiety Scale, **SPS** = Social Phobia Scale, **YBOCS** = Yale-Brown Obsessive Compulsive Scale.

**Table S9.** All service-level and intervention-level adaptations (detailed version) (*N* = 38)

| **Top-level categories** | **Sub-categories** | **Summary** | ***N* studies** | **Adaptations** | **Type of intervention** | **Rationale** |
| --- | --- | --- | --- | --- | --- | --- |
| Increase knowledge and detection of autism | Clinician training and skills | Training to provide clinicians with an overview of ASD and to tailor treatment to individual needs and increase self-efficacy, knowledge of autism and skills. Use of skills such as normalising experiences and prioritising the therapeutic relationship. | 6 | Training staff. The ASD-CP consisted of a modular staff training, toolkit and prescribed practices to be utilized with the patient. The training was structured as four 45-min modules consisting of lecture, video examples, interactive exercises, and role-play.  Module content included: an overview of ASD and foundation principles for intervention (e.g. ensuring basic needs are met), predictability (e.g. importance of a schedule), communication, and information on agitation and strategies to reduce it (Kuriakose et al., 2018; Cervantes et al., 2019). | - | General rationale - to address or accommodate barriers |
|  |  |  |  | Training was supported by a toolkit (including a brief specialized assessment checklist and visual supports to increase functional communication between staff and patients) and specified strategies e.g. ideas for activities, a coping card which can be used to help CYP choose a coping strategy (Kuriakose et al., 2018; Cervantes et al., 2019). | - | General rationale - to address or accommodate barriers |
|  |  |  |  | Development of the Extension for Community Healthcare Outcomes (Project ECHO) Autism model (Dreiling et al. 2022) | - | To increase mental health provider self-efficacy, knowledge of autism, and problem-solving skills |
|  |  |  |  | Development of the Autism Intellectual Disability and Psychiatry Disorder (AUP) network (Helverschou et al., 2021) | - | To improve professional competence and the quality of specialized mental health services for individuals with autism, intellectual disability, and psychiatric disorders |
|  |  |  |  | Normalise experiences (Fisher et al., 2023) | EMDR | General rationale: to address or accommodate barriers |
|  |  |  |  | Prioritise the therapeutic relationship above everything else (Fisher et al., 2023) | EMDR | General rationale: to address or accommodate barriers |
|  |  |  |  | Assure you are well tuned in to the client before starting (Fisher et al., 2023) | EMDR | General rationale: to address or accommodate barriers |
|  |  |  |  | Be open to learning from the client and celebrate each person's uniqueness (Fisher et al., 2023) | EMDR | General rationale: to address or accommodate barriers |
|  |  |  |  | Be aware of the possibility of sensory overload (Fisher et al., 2023) | EMDR | General rationale: to address or accommodate barriers |
|  |  |  |  | Maintain the specialist skillset of staff (having skilled or trained staff including staff being skilled to communicate clearly and understand the needs of people with autism) (Petty et al., 2021) | - | Not reported |
|  |  |  |  | Maintain awareness of gender differences (being aware that autism can present in many ways; female presentations of autism and masking, with elaborations suggesting this is not explicitly tied to client sex) (Petty et al., 2021) | - | Not reported |
|  |  |  |  | Maintain awareness of gendered socialisation (being aware that people are socialised in different ways regarding gender; that gender can influence one’s life experiences) (Petty et al., 2021) | - | Not reported |
|  |  |  |  | Know how someone identifies (knowing, asking or checking the gender someone identifies with, including awareness of people identifying in many ways in terms of gender) (Petty et al., 2021) | - | Not reported |
|  |  |  |  | Do not make assumptions (not making assumptions, having no expectations or being open-minded around gender, gender identity, sexuality or ways of addressing clients) (Petty et al., 2021) | - | Not reported |
|  |  |  |  | Respond to the person in front of you. Some may take well to rating scales and questions about cognitions, others may not at all (Fisher et al., 2023) | EMDR | General rationale: to address or accommodate barriers |
|  | Introduction of screening tools for the detection of autism | Use of assessments such as the ADOS, SCQ and DAWBA to improve the assessment and detection of autism. | 4 | Assessment of autism (Jones et al., 2021) | - | Not reported |
|  |  |  |  | Use of DAWBA (Ford et al., 2019) | - | Not reported |
|  |  |  |  | Parents and teachers completing the SCQ (Hollocks et al., 2019) | - | Not reported |
|  |  |  |  | Use of ADOS and SCQ (Stadnick et al., 2015) | - | Not reported |
| Adjustments to the physical environment | Provide environmental and practical adjustments | Provide adjustments to minimise sensory distractions such as a low stimulus area, adjustments to noise, decor, scents, and lighting. | 5 | The FriEnds environment was ecologically adapted to harness engagement for children with autism spectrum disorder, with five environmental zones created each with specific functions (Burke et al., 2017) | CBT | To harness engagement in the intervention |
|  |  |  |  | Check about preferred lighting prior to starting any intervention, and reduce noise (Spain et al., 2017) | - | To ensure the clinical  environment is not overly stimulating |
|  |  |  |  | Change the environment (e.g., reduce bright lights or distracting noises, provide fiddle toys) (Fisher et al., 2023) | EMDR | To reduce sensory demands |
|  |  |  |  | Open-access low-stimulus area (Jones et al., 2021) | - | Not reported |
|  |  |  |  | Low-stimulus area on request (Jones et al., 2021) | - | Not reported |
|  |  |  |  | Scheduled access low-stimulus area (Jones et al., 2021) | - | Not reported |
|  |  |  |  | Lighting adaptations (Jones et al., 2021) | - | Not reported |
|  |  |  |  | Check the suitability of the sensory environment (considering environment suitability; included checking the sensory environment and removing irritable or overwhelming things where possible) (Petty et al., 2021) | - | Not reported |
|  |  |  |  | Check suitability of lighting (considering lighting suitability, brightness or harshness) (Petty et al., 2021) | - | Not reported |
|  |  |  |  | Provide a sensory friendly environment (reducing sensory input to reduce overwhelm) (Petty et al., 2021) | - | Not reported |
|  |  |  |  | Reduce noise (included choosing quieter rooms, avoiding flapping blinds or tapping a pen) (Petty et al., 2021) | - | Not reported |
|  |  |  |  | Provide adjustable lighting (included trying to offer natural lighting or a range of lamp/lighting options) (Petty et al., 2021) | - | Not reported |
|  |  |  |  | Reduce scents (cooking or food smells were considered; strong disinfectants or air fresheners were avoided) (Petty et al., 2021) | - | Not reported |
|  |  |  |  | Neutralise decor (a neutral colour scheme avoided bright colours for the walls, furniture, carpets, clothing or accessories) (Petty et al., 2021) | - | Not reported |
|  |  |  |  | Reduce the number of items in the environment (practitioners maintained a minimal amount of objects within each room) (Petty et al., 2021) | - | Not reported |
|  |  |  |  | Avoid patterns (patterns in the environment, including on walls, carpets or clothes, were avoided) (Petty et al., 2021) | - | Not reported |
|  |  |  |  | Control outside noise (lawn mowers, traffic, maintenance works, simultaneous appointments and open windows were each described as noises to manage) (Petty et al., 2021) | - | Not reported |
|  |  |  |  | Offer space (choice was given to clients where possible of the size and layout of the room, especially for psychological therapy sessions; choice was given on seating arrangement) (Petty et al., 2021) | - | Not reported |
|  |  |  |  | Neutralise all sensory demands (a plain and neutral sensory environment was described, including reflecting on possible sensory demands) (Petty et al., 2021) | - | Not reported |
|  |  |  |  | Keep to plain design (keeping things plain or neutral included minimal decoration and uncluttered rooms or walls) (Petty et al., 2021) | - | Not reported |
|  |  |  |  | Ensure suitable noise levels (controlling noise included minimising sounds outside the building, using quieter rooms, having quiet waiting rooms or minimising noise from phones, clocks or equipment inside the building) (Petty et al., 2021) | - | Not reported |
|  |  |  |  | Consider the room seating arrangement (considering room seating arrangements, including where to sit, letting the client choose their seat or checking they are comfortable with seating closeness) (Petty et al., 2021) | - | Not reported |
|  |  |  |  | Utilise a protected building or space (having a designated building or space for autism services ensured design and environment decisions could be maintained) (Petty et al., 2021) | - | Not reported |
|  |  |  |  | Use signs up to modify the environment (signs encouraged clients to adjusts their environment, for example to shut the blinds or turn music off) (Petty et al., 2021) | - | Not reported |
|  |  |  |  | Ability to adapt meal plans to sensory requirements (Jones et al., 2021) | - | Not reported |
|  |  |  |  | Always offer sessions at the same time and place (Fisher et al., 2023) | EMDR | General rationale: to address or accommodate barriers |
|  | Encourage the use of sensory resources and stimming | Provide sensory resources such as tactile objects, include movement breaks and exercises and use multi-sensory activities, encourage use of stimming behaviour. | 5 | Movement breaks and exercises recommended by the occupational therapy department (Burke et al., 2017) | CBT | To meet the sensory needs of individuals with autism. |
|  |  |  |  | Sensory tools were provided at each session, including various tactile objects such as therabands and theraputty (Burke et al., 2017) | CBT | To meet the sensory needs of individuals with autism. |
|  |  |  |  | The use of relaxation strategies (Sofronoff et al., 2017) | CBT | Not reported |
|  |  |  |  | Ear defenders (Jones et al., 2021) | - | Not reported |
|  |  |  |  | Weighted blankets (Jones et al., 2021) | - | Not reported |
|  |  |  |  | Stress ball (Jones et al., 2021) | - | Not reported |
|  |  |  |  | Relaxing music (Jones et al., 2021) | - | Not reported |
|  |  |  |  | Provide sensory resources (sensory or fidget toys were available) (Petty et al., 2021) | - | Not reported |
|  |  |  |  | Encourage them to use stimming behaviour as self-soothing if it works (Fisher et al., 2023) | EMDR | General rationale: to address or accommodate barriers |
| Communication accommodations | Use of clear, simple and preferred language | Provide clear instructions and guidance, be more directive, monitor, adapt and slow the pace of communication, use preferred language where possible. | 7 | Use more directive interweaves than usual (Fisher et al., 2023) | EMDR | General rationale: to address or accommodate barriers |
|  |  |  |  | Give explicit permission to ask question (Fisher et al., 2023) | EMDR | General rationale: to address or accommodate barriers |
|  |  |  |  | Be more directive in style (i.e., less Socratic, with fewer open-ended questions) (Fisher et al., 2023) | EMDR | General rationale: to address or accommodate barriers |
|  |  |  |  | Spell things out in black and white and be more directive than usual during history taking (Fisher et al., 2023) | EMDR | General rationale: to address or accommodate barriers |
|  |  |  |  | Use more prompts and suggestions to find positive cognition (Fisher et al., 2023) | EMDR | General rationale: to address or accommodate barriers |
|  |  |  |  | Be very clear with clients what the preparation phase is about and why it is necessary (Fisher et al., 2023) | EMDR | General rationale: to address or accommodate barriers |
|  |  |  |  | Offer clear guidance on what to do after the session and what they might experience after the session (Fisher et al., 2023) | EMDR | General rationale: to address or accommodate barriers |
|  |  |  |  | Adapt communication (being aware of communication with clients meant communicating clearly informed by an understanding of autism) (Petty et al., 2021) | - | Not reported |
|  |  |  |  | Monitor own communication (descriptions of being aware of one’s own communication, including communication styles and skills, pragmatic communication (Petty et al., 2021) | - | Not reported |
|  |  |  |  | Communicate clearly (using clear, direct, firm, concrete or verbally explicit communication, language, or requests, adapting communication to client’s language profile or avoiding jargon) (Petty et al., 2021) | - | Not reported |
|  |  |  |  | Slow the pace of communication (communicating more slowly, including thinking about talking pace or giving clients more time to process language) (Petty et al., 2021) | - | Not reported |
|  |  |  |  | Be prepared to adjust communication (included having information available in different formats or being able to explain things in a different way) (Petty et al., 2021) | - | Not reported |
|  |  |  |  | Check for understanding (checking that the client understands or has understood) (Petty et al., 2021) | - | Not reported |
|  |  |  |  | Repeat their feedback to them (Fisher et al., 2023) | EMDR | To aid processing |
|  |  |  |  | Minor, language-based modifications were made to the BIACA protocol (Ehrenreich-May et al., 2014) | CBT | These amendments were made in order to make sure the intervention was developmentally appropriate for anxious adolescents who have autism at this stage of evaluation. |
|  |  |  |  | Language specifically adapted for children and adolescents with ASD (Storch et al., 2020) | FET | General rationale - to make treatment suitable for CYP with ASD |
|  |  |  |  | Little reliance on metaphors or colloquialisms (Spain et al., 2017) |  | Not reported |
|  |  |  |  | Conversation includes didactic questions as well as a socratic style (Spain et al., 2017). |  | Not reported |
|  |  |  |  | Avoid metaphors in therapy (Cooper et al., 2018) | CBT | Not reported |
|  |  |  |  | Avoid metaphor (Fisher et al., 2023) | EMDR | General rationale: to address or accommodate barriers |
|  |  |  |  | Be clear in communication (communicating clearly; included descriptions of being black and white, using simple and concise language or giving salient summary points) (Petty et al., 2021) | - | Not reported |
|  |  |  |  | Avoid ambiguity (avoiding ambiguity or ambiguous expressions in communication, including non-literal or figurative language) (Petty et al., 2021) | - | Not reported |
|  |  |  |  | Avoid idioms (avoiding idioms in communication. This include giving examples of idioms with explanation of avoiding such language) (Petty et al., 2021) | - | Not reported |
|  |  |  |  | Use a literal description. Ask the person to explain what we would see if looking at a photo or a still of a movie (Fisher et al., 2023) | EMDR | General rationale: to address or accommodate barriers |
|  |  |  |  | Concrete/special interest-related analogies were used (A. J. Russell et al., 2013) | CBT | To convey psychological concepts |
|  |  |  |  | Use plain English more than with other clients (Cooper et al., 2018) | CBT | Not reported |
|  |  |  |  | Use very clear language. Do not assume that they have necessarily understood what you intended to say (Fisher et al., 2023) | EMDR | General rationale: to address or accommodate barriers |
|  |  |  |  | Take time to understand the language they use around thoughts and emotions, and mirror this (Fisher et al., 2023) | EMDR | General rationale: to address or accommodate barriers |
|  |  |  |  | Use their own language to describe emotions (Fisher et al., 2023) | EMDR | General rationale: to address or accommodate barriers |
|  |  |  |  | Be aware of how they communicate their level of arousal through behaviour and use this information to evaluate how they are coping during sessions (Fisher et al., 2023) | EMDR | General rationale: to address or accommodate barriers |
|  |  |  |  | Be particularly mindful of language (Fisher et al., 2023) | EMDR | People may be very sensitive to failure and ‘getting it wrong’ |
|  | Use of simple written material and visual aids | Use of written information and external cues such as use of a whiteboard, activity books, worksheets, timers, agendas and calendars. Use of visual aids such as drawings, pictures, videos and leaflets. | 16 | Adapted intervention-level adaptations: Using visual aides e.g. the daily agenda and rules were visually displayed (Burke et al., 2017) | CBT | To reinforce predictability, structure and routine. |
|  |  |  |  | More visual aids and structured worksheets were used than in the original intervention (Chalfant et al., 2007; Kilburn et al., 2019; Kilburn et al., 2020) | CBT | General rationale - to address or accommodate barriers |
|  |  |  |  | The use of activity books (Cook et al., 2017) | CBT | To ensure that the content was accessible and suitable for young children with HFASD |
|  |  |  |  | Use of visual supports for each concept (Cook et al., 2017) | CBT | To ensure that the content was accessible and suitable for young children with HFASD |
|  |  |  |  | Use of visual aids and visual charts to lay out coping plans and exposure hierarchies (Driscoll et al., 2020) | CBT | Not reported |
|  |  |  |  | Provision of visual supports (Hepburn et al., 2016) | CBT | To ensure the intervention was appropriate for YP with ASD |
|  |  |  |  | The program differs from traditional CBT in that it is more “visually focused” and less based on having the children write in a workbook (Oerbeck et al., 2021) | CBT | General rationale - to make intervention more suited to CYP with ASD |
|  |  |  |  | A visual schedule was used to support children in tracking their progress. The schedule consisted of an image of a road with a set number of stops; children progressed along the road by completing various exposure steps, earning a break at the end of each hour by reaching the end of the road (Ollendick et al., 2021) | CBT | General rationale - to make intervention more suited to CYP with ASD |
|  |  |  |  | More written and visual information (Cooper et al., 2018) | CBT | Not reported |
|  |  |  |  | Use visual aids (e.g., drawing, pictures, videos) (Fisher et al., 2023) | EMDR | General rationale: to address or accommodate barriers |
|  |  |  |  | Create a visual timeline (Fisher et al., 2023) | EMDR | General rationale: to address or accommodate barriers |
|  |  |  |  | Include props (e.g., charts about feelings and an emotion wheel) (Fisher et al., 2023) | EMDR | To help them identify emotions |
|  |  |  |  | Use visual or simplified version of ratings scales (Fisher et al., 2023) | EMDR | General rationale: to address or accommodate barriers |
|  |  |  |  | Use an image of a place rather than imaginal calm place (Fisher et al., 2023) | EMDR | General rationale: to address or accommodate barriers |
|  |  |  |  | Visual tools were used (A. J. Russell et al., 2013) | CBT | To convey psychological concepts |
|  |  |  |  | Visual signage or orientation tools (Jones et al., 2021) | - | Not reported |
|  |  |  |  | Visual help/cue cards (Jones et al., 2021) | - | Not reported |
|  |  |  |  | Increased use of visual aids (Wise et al., 2019) | CBT | Not reported |
|  |  |  |  | Adapt written correspondence (written correspondence included clear and specific written communication or references to letters with information, checked by experts by experience) (Petty et al., 2021) | - | Not reported |
|  |  |  |  | Use agendas (utilising or giving the option of an agenda, sometimes specified as a written or visual agenda, or using visual strategies to explain session structure) (Petty et al., 2021) | - | Not reported |
|  |  |  |  | Make information available for clients about the service (available information included leaflets, information packs or documents about the service and what to expect from a visit, photographs showing clients where they are going or who they will meet) (Petty et al., 2021) | - | Not reported |
|  |  |  |  | Mentalise and materialize the mental states and pictures on the whiteboard while speaking (Ekman et al., 2015) | CBT | To illustrate and systematize for the client |
|  | Provide communication support | Use of communication passports and social stories. | 1 | Communication passports (Jones et al., 2021) | - | Not reported |
|  |  |  |  | Social stories (Jones et al., 2021) | - | Not reported |
|  | Use of technology | Incorporating technology into the intervention to aid communication | 2 | The PowerPoint program aids in having participant and therapist focused on an external object (the computer), thus reducing the face-to-face contact often found difficult for children with ASD (Oerbeck et al., 2021). | CBT | General rationale - to make intervention more suited to CYP with ASD |
|  |  |  |  | Technology was incorporated in the form of a personalized digital assistant (J. Reaven, Blakeley-Smith, Leuthe et al., 2012) | CBT | General rationale - to accommodate the needs of teenagers with ASD |
| Accommodate individual differences | Evaluate individual needs and preferences | Evaluate preferences, sensitivities, sensory needs, likes and dislikes and coping strategies. | 3 | Assessment of likes and dislikes (Jones et al., 2021) | - | Not reported |
|  |  |  |  | Bespoke sensory assessment (Jones et al., 2021) | - | Not reported |
|  |  |  |  | Assess sensory preferences and sensitivities (Fisher et al., 2023) | EMDR | General rationale: to address or accommodate barriers |
|  |  |  |  | Assessment of coping strategies (Jones et al., 2021) | - | Not reported |
|  |  |  |  | Find out if the client has sensory needs (understanding if the client has sensory needs) (Petty et al., 2021) | - | Not reported |
|  |  |  |  | Agree etiquette for making eye contact (reducing, avoiding or not expecting eye contact; thinking or asking clients about their eye contact preferences) (Petty et al., 2021) | - | Not reported |
|  |  |  |  | Use a preference notifications system (using a computer system where gender and associated preferences can be added as visible notifications for all staff) (Petty et al., 2021) | - | Not reported |
|  |  |  |  | Know pronoun or name preferences (descriptions included knowing, asking, checking or using the preferred pronouns, names or terminology) (Petty et al., 2021) | - | Not reported |
|  |  |  |  | Check suitability of clinician gender (ensuring a client is comfortable with the clinician’s gender; offering a chaperone or a different therapist) (Petty et al., 2021) | - | Not reported |
|  |  |  |  | Presence of a standardised protocol for people with autism (specific protocol for admission, assessment and management of people with autism) (Jones et al., 2021) | - | Not reported |
|  | Encourage individual’s hobbies and interests | Include and ask about individual’s special interests and hobbies in therapy. | 4 | Clinicians asked children and their parents about any special interests their child might have and attempted to incorporate these interests into treatment (Ollendick et al., 2021). | CBT | General rationale - to make intervention more suited to CYP with ASD |
|  |  |  |  | Discussing individual hobbies and interests as part of therapy (Cooper et al., 2018) | CBT | Not reported |
|  |  |  |  | Ask about and include special interests throughout the therapy (Fisher et al., 2023) | EMDR | General rationale: to address or accommodate barriers |
|  |  |  |  | Use their special interests and how they feel when engaged in it as a resource (Fisher et al., 2023) | EMDR | General rationale: to address or accommodate barriers |
|  |  |  |  | Incorporation of specific interests into treatment (Wise et al., 2019) | CBT | General rationale: to address or accommodate barriers |
|  | Tailor practice to individual needs and preferences | Tailor care plans and practice to individual differences such as incorporating approaches targeted at neurodevelopmental comorbidities, being flexible with the treatment manual and the session timings and ensuring that resources are appropriate for the person’s gender. | 13 | Treatment manuals applied flexibly (Chalfant et al., 2007; Kilburn et al., 2019; Kilburn et al., 2020) | CBT | To take into account individual needs of children and parents |
|  |  |  |  | Using coping self-statements chosen by the child, and individually-tailored coping plans (Driscoll et al., 2020). | CBT | Not reported |
|  |  |  |  | Manualised treatment was personalised to the specific child, engaging them via their own interests and motivators and designing exposures addressing the child's specific fears and worries (Driscoll et al., 2020). | CBT | Not reported |
|  |  |  |  | If a child was accustomed to using social stories, these were incorporated into the intervention (Driscoll et al., 2020). | CBT | To remind the child of the coping plan and of how practicing facing a feared situation helps them become braver. |
|  |  |  |  | Flexibility in delivery, appropriate to the group’s interests, needs and learning styles (E. Higgins et al., 2019) | CBT | Not reported |
|  |  |  |  | Ensure that appointments are offered at a convenient time (Spain et al., 2017) | - | General rationale: to make practice more accessible |
|  |  |  |  | Encourage people to be ‘active participants’ whereby their views about the pace and content of clinical work are sought (Spain et al., 2017) | - | To provide more opportunities to develop assertiveness skills |
|  |  |  |  | Treatment followed a modular, flexible format in which therapy modules were selected on an ongoing basis by the therapist and supervisor (Storch et al., 2013) | CBT | General rationale - to make treatment suitable for CYP with ASD |
|  |  |  |  | Additional sessions and treatment modules were tailored to individual’s needs (Storch et al., 2013). | CBT | General rationale - to make treatment suitable for CYP with ASD |
|  |  |  |  | The instructional method retained flexibility and allows for differences in therapy facilitation between the younger group (9–12 years old) and the older group (13–16 years old) (Sung et al., 2011) | CBT | Therapists need to be sensitive to the developmental differences between children of different ages and intellectual maturity for therapy to be effective |
|  |  |  |  | The CBT program was also tailored to meet the cultural and developmental profiles of children with ASD in the Asian population (Sung et al., 2011) | CBT | To meet needs of the population in this country |
|  |  |  |  | Additional modules were implemented as needed to address social and adaptive skill deficits/problems, poor motivation, social/school issues, and comorbid conditions (Storch et al., 2015; Wood et al., 2015) | CBT | General rationale - to make treatment suitable for CYP with ASD |
|  |  |  |  | Care plans based on individual needs specific to people with autism (Jones et al., 2021) | - | Not reported |
|  |  |  |  | Try different types of bilateral stimulation (e.g., eye movements, tapping, auditory sounds) (Fisher et al., 2023) | EMDR | General rationale: to address or accommodate barriers |
|  |  |  |  | Vary the way you work (e.g., on the floor, walking, use play, engage with their hobbies and interests) (Fisher et al., 2023) | EMDR | General rationale: to address or accommodate barriers |
|  |  |  |  | Offer alternatives, prompts and suggestions for cognitions (Fisher et al., 2023) | EMDR | General rationale: to address or accommodate barriers |
|  |  |  |  | Ask for all the elements but if they cannot provide information, go with whatever is given (Fisher et al., 2023) | EMDR | General rationale: to address or accommodate barriers |
|  |  |  |  | Proceed without an image if they struggle with finding an image (Fisher et al., 2023) | EMDR | General rationale: to address or accommodate barriers |
|  |  |  |  | Skip negative cognition altogether if it causes problems (Fisher et al., 2023) | EMDR | General rationale: to address or accommodate barriers |
|  |  |  |  | Use any sensory modality as a target, not necessarily an image (Fisher et al., 2023) | EMDR | General rationale: to address or accommodate barriers |
|  |  |  |  | Allow the positive cognition to emerge during processing rather than identifying it beforehand (Fisher et al., 2023) | EMDR | General rationale: to address or accommodate barriers |
|  |  |  |  | Be flexible and creative (Fisher et al., 2023) | EMDR | General rationale: to address or accommodate barriers |
|  |  |  |  | Don't emphasise keeping logs between sessions if difficult for the client (Fisher et al., 2023) | EMDR | General rationale: to address or accommodate barriers |
|  |  |  |  | Do not expect generalisation (Fisher et al., 2023) | EMDR | General rationale: to address or accommodate barriers |
|  |  |  |  | Use more physical movement (Fisher et al., 2023) | EMDR | General rationale: to address or accommodate barriers |
|  |  |  |  | Use softeners for the positive cognition (e.g., instead of I am strong, use I am starting to believe that I am strong) (Fisher et al., 2023) | EMDR | General rationale: to address or accommodate barriers |
|  |  |  |  | Use fantasy figures as resources (e.g., superheroes) (Fisher et al., 2023) | EMDR | General rationale: to address or accommodate barriers |
|  |  |  |  | Consider small traumas as well as big traumas as possible targets (Fisher et al., 2023) | EMDR | General rationale: to address or accommodate barriers |
|  |  |  |  | Use flash (Fisher et al., 2023) | EMDR | General rationale: to address or accommodate barriers |
|  |  |  |  | Use a present-day target first (Fisher et al., 2023) | EMDR | General rationale: to address or accommodate barriers |
|  |  |  |  | Make time for the person to debrief about their week (Fisher et al., 2023) | EMDR | General rationale: to address or accommodate barriers |
|  |  |  |  | Let the client choose and control length of sets (Fisher et al., 2023) | EMDR | General rationale: to address or accommodate barriers |
|  |  |  |  | Think in terms of a 'positive engaging focus' rather than necessarily a 'calm place' (Fisher et al., 2023) | EMDR | General rationale: to address or accommodate barriers |
|  |  |  |  | Expect to add to history taking throughout the therapy as new information emerges (Fisher et al., 2023) | EMDR | General rationale: to address or accommodate barriers |
|  |  |  |  | Be ready to reformulate throughout the therapy and to shift the focus. Whilst you might start with symptoms, later work could focus on identity, the impact of neurodiversity and adapting to diagnosis (Fisher et al., 2023) | EMDR | General rationale: to address or accommodate barriers |
|  |  |  |  | Keep it simple, even with things that are very complex, and adapt to the person’s level of understanding (Fisher et al., 2023) | EMDR | General rationale: to address or accommodate barriers |
|  |  |  |  | Be creative with the calm place (e.g., use drawings, emojis, pictures, media clips, animals, fiddle toys, smells) (Fisher et al., 2023) | EMDR | General rationale: to address or accommodate barriers |
|  |  |  |  | Focus first on strengths and interests and then move onto problems and history (Fisher et al., 2023) | EMDR | General rationale: to address or accommodate barriers |
|  |  |  |  | End with a relaxing and positive activity (Fisher et al., 2023) | EMDR | General rationale: to address or accommodate barriers |
|  |  |  |  | Include your own thoughts as part of the debrief (Fisher et al., 2023) | EMDR | General rationale: to address or accommodate barriers |
|  |  |  |  | Use their background and history to identify resource possibilities (Fisher et al., 2023) | EMDR | General rationale: to address or accommodate barriers |
|  |  |  |  | Focus on quality of life and functioning (Fisher et al., 2023) | EMDR | General rationale: to address or accommodate barriers |
|  |  |  |  | Focus on quality of life and functioning to assess progress (Fisher et al., 2023) | EMDR | General rationale: to address or accommodate barriers |
|  |  |  |  | Don't insist on or encourage eye contact (Fisher et al., 2023) | EMDR | General rationale: to address or accommodate barriers |
|  |  |  |  | Offer your own observations of what has changed (Fisher et al., 2023) | EMDR | General rationale: to address or accommodate barriers |
|  |  |  |  | Offer a flexible and individualised approach (adapting to each individual, including using techniques known to work for them) (Petty et al., 2021) | - | Not reported |
|  |  |  |  | Adapt questioning for female representation (going beyond standard questions in an assessment) (Petty et al., 2021) | - | Not reported |
|  |  |  |  | Offer gender appropriate resources (ensuring that any resources used are appropriate for the gender a client identifies with) (Petty et al., 2021) | - | Not reported |
|  |  |  |  | Offer flexible session timings (being flexible with appointments included breaking appointments or having shorter sessions) (Petty et al., 2021) | - | Not reported |
| Structural or procedural adaptations | Format of the intervention | Reduce or increase the number and duration of sessions and exercises, conduct exposure in more varied settings. | 8 | Changes made to time devoted to certain aspects of programme: the largest components of the programme were devoted to relaxation (three treatment sessions and two booster sessions) and exposure (4½ treatment sessions and all booster sessions) (Chalfant et al., 2007; Kilburn et al., 2019; Kilburn et al., 2020) | CBT | These aspects involve more concrete exercises and place less emphasis on the children’s communication skills, which are impaired in people with high functioning autism. |
|  |  |  |  | Exposure was conducted in more varied settings than usual (Driscoll et al., 2020) | CBT | To promote generalisation |
|  |  |  |  | The number of in-session exposure practices was increased and occurred almost exclusively within the adolescent group and without direct parent involvement in-session (J. Reaven, Blakeley-Smith, Leuthe et al., 2012) | CBT | General rationale - to accommodate the needs of teenagers with ASD |
|  |  |  |  | The program was extended over a longer period of time (6 months) (Chalfant et al., 2007; Kilburn et al., 2019; Kilburn et al., 2020) | CBT | Not reported |
|  |  |  |  | Longer treatment course (Fujii et al., 2013) | CBT | To allow more time for adaptations to intervention |
|  |  |  |  | Adding two booster sessions and homework tasks to reinforce strategies explored (E. Higgins et al., 2019) | CBT | To ensure long-term maintenance of skills |
|  |  |  |  | Shorter or longer sessions (Cooper et al., 2018) | CBT | Not reported |
|  | A structured and predictable approach | Having predictable session routines and a structured approach to treatment, with details communicated in advance | 8 | Share a plan in advance for each session (Fisher et al., 2023) | EMDR | So that the client knows what to expect |
|  |  |  |  | The “transition activity” facilitated children’s gradual integration to the group, upon arrival. Each week a structured activity was put in place where children could engage in a structured task, for example, Lego, colouring, arts and crafts (Burke et al., 2017). | CBT | To reduce anxiety on arrival to the group |
|  |  |  |  | A highly structured predictable format in activities (Cook et al., 2017) | CBT | To ensure that the content was accessible and suitable for young children with HFASD |
|  |  |  |  | Predictable session routines (Hepburn et al., 2016) | CBT | To ensure the intervention was appropriate for YP with ASD |
|  |  |  |  | Predictable routines (E. Higgins et al., 2019) | CBT | Not reported |
|  |  |  |  | A more structured and concrete approach to therapeutic work (Cooper et al. 2018) | CBT | Not reported |
|  |  |  |  | A structured and therapist-directed approach to sessional and homework content was taken (A. J. Russell et al., 2013) | CBT | Not reported |
|  |  |  |  | Find out about the client in advance (finding out about the client in advance; included current wellbeing and priorities and checking notes about their gender preferences) (Petty et al., 2021) | - | Not reported |
|  |  |  |  | Find out about the client in advance from significant people (finding out about the client in advance from family, teachers, carers or other significant people) (Petty et al., 2021) | - | Not reported |
|  |  |  |  | Find out about the client in advance from case notes (finding out about the client in advance specifically by reading case notes or diagnostic reports) (Petty et al., 2021) | - | Not reported |
|  |  |  |  | Ensure the client is prepared for what will happen (ensuring the client is prepared for what is going to happen or what questions will be asked during the session) (Petty et al., 2021) | - | Not reported |
|  |  |  |  | Ensure the client is prepared about the purpose of the appointment (including descriptions about why they are there and ensuring consent) (Petty et al., 2021) | - | Not reported |
| Intervention content adaptations | Simplified content | Remove or simplify psychoeducation and cognitive elements of the intervention | 5 | The information in the cognitive activities and for the tasks involving generating helpful and unhelpful thoughts and thought challenging were simplified e.g., rather than have to come up with their own ideas of helpful and unhelpful thoughts, they could identify examples from a worksheet (Chalfant et al., 2007; Kilburn et al., 2019; Kilburn et al., 2020). | CBT | The changes to the cognitive component were made because of the children’s language and/or communication impairments |
|  |  |  |  | Adaptations to make intervention developmentally appropriate for younger children (Factor et al., 2019). | Stress and Anger Management Program (STAMP) | To adapt intervention for younger children |
|  |  |  |  | Cognitive work (e.g., identifying and challenging negative beliefs) was simplified and used to support the behavioural components (role plays, exposure tasks and out-of-session practice tasks) that formed the core interventions in the program (Bemmer et al., 2021). | CBT | General rationale: considered the needs of adults with SAD and co-occuring ASD who have difficulty implementing typical cognitive interventions due to limited introspection and a poorer understanding of social rules and norms |
|  | Creative outlets and activities | Incorporating creativity and arts-based activities into the intervention | 3 | An emphasis on drawing, collage, and other creative outlets designed for young children (Cook et al., 2017). | CBT | To ensure that the content was accessible and suitable for young children with HFASD |
|  |  |  |  | Using singing and stories (Swain et al., 2019) | CBT | To make the intervention engaging and developmentally appropriate |
|  |  |  |  | Greater use of songs, stories, and play activities (Scarpa et al., 2011) | CBT | To be developmentally appropriate for 5- to 7-year-old children with ASD. |
|  | Use of role play or modelling | Using role play as a technique to reinforce learning during sessions, or modelling activities e.g. by video, or using a puppet or character | 6 | FriEnds incorporated the implementation of regular puppet shows where both facilitators and children would interact in the creation of scripts and role plays centred on course material (Burke et al., 2017) | CBT | Not reported |
|  |  |  |  | Use of puppets (Cook et al., 2017) | CBT | To ensure that the content was accessible and suitable for young children with HFASD |
|  |  |  |  | Video modelling activities (Hepburn et al., 2016) | CBT | To ensure the intervention was appropriate for YP with ASD |
|  |  |  |  | Including role plays, games and activities (E. Higgins et al., 2019) | CBT | Not reported |
|  |  |  |  | Role-plays (Sofronoff et al., 2017) | CBT | Not reported |
|  |  |  |  | Use of characters as part of a computer game (Sofronoff et al., 2017) | CBT | Not reported |
|  |  |  |  | The use of a 'role model' character who investigates and works with anxiety by conducting experiments (exposures) and uses a richly equipped toolbox to fix anxiety arising in these experiments (Oerbeck et al., 2021) | CBT | General rationale - to make intervention more suited to CYP with ASD |
|  |  |  |  | The program is concrete (avoiding abstract terms) and activity based (with role-playing, games and to exposing themselves for the things they fear (exposure tasks) both within sessions and as home assignment tasks) (Oerbeck et al., 2021) | CBT | General rationale - to make intervention more suited to CYP with ASD |
|  | Use of a rewards system | Using reward systems to help reinforce learning | 4 | Emphasis on rewards in anxiety-focused modules (Storch et al., 2020; Wood et al., 2015) | FET | General rationale - to make treatment suitable for CYP with ASD |
|  |  |  |  | Children were rewarded for skill usage between sessions with points in a Rewards Diary, which were exchanged for home-based rewards (Sofronoff et al., 2017) | CBT | Not reported |
|  |  |  |  | Emphasis on a reward system (Wise et al., 2019) | CBT | To help motivate the adolescent to engage in exposure therapy |
|  | Taking it slow | Taking a slow/progressive approach to treatment, with opportunity for repetition and practice. | 6 | Shorter activity duration (Cook et al., 2017) | CBT | To ensure that the content was accessible and suitable for young children with HFASD |
|  |  |  |  | Shorter sessions (Scarpa et al., 2011) | CBT | To be developmentally appropriate for 5- to 7-year-old children with ASD. |
|  |  |  |  | Multiple opportunities for repetition and practice (Cook et al., 2017) | CBT | To ensure that the content was accessible and suitable for young children with HFASD |
|  |  |  |  | An emphasis on repeated practice of well-rehearsed coping plans in a stepwise series of situations (Driscoll et al., 2020) | CBT | This is well-suited to the behavioral styles of anxious children with ASD and is consistent with social skills approaches that emphasize rehearsal of scripts to cope with novel or challenging situations. |
|  |  |  |  | Repetition of key concepts (Hepburn et al., 2016) | CBT | To ensure the intervention was appropriate for YP with ASD |
|  |  |  |  | “Home Missions” were completed by children between sessions (Sofronoff et al., 2017) | CBT | To facilitate skill generalisation |
|  |  |  |  | Slow down every phase (Fisher et al., 2023) | EMDR | General rationale: to address or accommodate barriers |
|  |  |  |  | Take graduated/progressive approach towards full trauma processing (Fisher et al., 2023) | EMDR | General rationale: to address or accommodate barriers |
|  |  |  |  | Take longer to close down and leave extra time for a debrief (Fisher et al., 2023) | EMDR | General rationale: to address or accommodate barriers |
|  |  |  |  | Use shorter sets and a more frequent return to target (Fisher et al., 2023) | EMDR | General rationale: to address or accommodate barriers |
|  |  |  |  | Use a progressive approach to processing, starting with the 'tip of the finger' (Fisher et al., 2023) | EMDR | General rationale: to address or accommodate barriers |
|  |  |  |  | Start with short sets and build up tolerance from there (Fisher et al., 2023) | EMDR | General rationale: to address or accommodate barriers |
|  | Consider the role of autism | Consider the role of autism, develop an understanding of autism such as its characteristics and impact on daily life | 2 | Consider the role of autism within the conceptualisation (Fisher et al., 2023) | EMDR | General rationale: to address or accommodate barriers |
|  |  |  |  | The CBT intervention was adapted to consider barriers to working with children with ASD (e.g., low motivation, comorbidity) (Storch et al., 2013) | CBT | General rationale - to make treatment suitable for CYP with ASD |
|  | Integration of emotion-focused strategies | Provide psychoeducation on emotions, arousal and feeling physiologically overwhelmed and exercises to access emotions. | 7 | A larger affective education component (Cook et al., 2017) | CBT | To ensure that the content was accessible and suitable for young children with HFASD |
|  |  |  |  | A relative de-emphasis on identifying and modifying anxiety-provoking cognitions (Driscoll et al., 2020) | CBT | Not reported |
|  |  |  |  | Creating an ‘Emotional Toolbox’ of coping strategies (Swain et al., 2019) | CBT | Not reported |
|  |  |  |  | Include exercises to facilitate accessing emotions (Fisher et al., 2023) | EMDR | General rationale: to address or accommodate barriers |
|  |  |  |  | Psychoeducation about emotions (Cooper et al., 2018) | CBT | Not reported |
|  |  |  |  | If required, educational sessions about understanding and rating anxiety were provided (A. J. Russell et al., 2013) | CBT | Not reported |
|  |  |  |  | Provide extra psychoeducation around trauma, arousal and feeling physiologically overwhelmed (Fisher et al., 2023) | EMDR | General rationale: to address or accommodate barriers |
|  |  |  |  | Psychoeducation on anxiety and ASD for both the adolescent and parent when involved in therapy (Wise et al., 2019) | CBT | General rationale: to address or accommodate barriers |
|  | Integration of cognitive-behavioural approaches | Provide cognitive and behavioural strategies including building a positive self-image, coping strategies and making links between behaviour, thoughts and feelings. | 6 | A greater emphasis on behavioural versus cognitive strategies (Cook et al., 2017) | CBT | To ensure that the content was accessible and suitable for young children with HFASD |
|  |  |  |  | Cognitive components to teach children to recognize unhelp-ful thoughts while replacing these with helpful thoughts, and children are taught social problem solving skills (Sofronoff et al., 2017) | CBT | Not reported |
|  |  |  |  | Behavioural strategies to introduce change (Cooper et al., 2018) | CBT | Not reported |
|  |  |  |  | Behavioural interventions were integrated within treatment sessions, and as a focus of weekly homework (Bemmer et al., 2021) | CBT | To facilitate engagement and promote positive treatment outcomes |
|  |  |  |  | Cognitive strategies to introduce change (Cooper et al., 2018) | CBT | Not reported |
|  |  |  |  | Focus on building a positive self-image and coping strategies rather than pathologising and eliminating symptoms (Fisher et al., 2023) | EMDR | General rationale: to address or accommodate barriers |
|  |  |  |  | Install a positive self-view as a resource (Fisher et al., 2023) | EMDR | General rationale: to address or accommodate barriers |
|  |  |  |  | Do more cognitive work if necessary to identify a positive cognition (Fisher et al., 2023) | EMDR | General rationale: to address or accommodate barriers |
|  |  |  |  | Ensuring the building blocks for treatment (i.e., understanding and differentiating emotions, particularly anxiety, and making links between thoughts, feelings and behaviours) were in place (A. J. Russell et al., 2013) | CBT | Not reported |
|  | Integration of social skills training | Integration of social skills training such as entering and maintaining conversations and managing disagreements | 6 | Emphasis on theory of mind in all activities (Burke et al., 2017). | CBT | To help social interaction between children |
|  |  |  |  | Emphasis on feelings in others and recognising feelings in oneself, facilitators also verbalised their thoughts and feelings (Burke et al., 2017). | CBT | To help social interaction between children |
|  |  |  |  | Exposure exercises for social anxiety emphasise role plays and reinforce practice of social skills, such as greetings, eye contact, and initiating and maintaining conversations (Driscoll et al., 2020). | CBT | Not reported |
|  |  |  |  | Including other ASD specific content around friendship skills (Fujii et al., 2013) | CBT | Not reported |
|  |  |  |  | A specific social skills module was developed (J. Reaven, Blakeley-Smith, Leuthe et al., 2012) | CBT | To address deficits in social skills functioning |
|  |  |  |  | Inclusion of structured frameworks for teaching of social skills such as entering and maintaining conversations and managing disagreements (Bemmer et al., 2021) | CBT | To make the anxiety-based interventions more effective for adults with ASD |
|  |  |  |  | Treatment components (e.g., independence and social skills) (Wise et al., 2019) | CBT | To address ASD-specific deficits |
| Involving wider support network | Parental involvement | Involving parents in the intervention | 17 | A parent-based group CBT manual was also adapted from the Cool Kids program to use with the families in concurrent sessions to the child program. The parent program addressed anxiety education, relaxation strategies, cognitive restructuring exercises, graded exposure, parent management training and relapse prevention (Chalfant et al., 2007; Kilburn et al., 2019; Kilburn et al., 2020) | CBT | Not reported |
|  |  |  |  | Parent involvement (Driscoll et al., 2020) | CBT | Not reported |
|  |  |  |  | Parents took a larger role in planning exposure hierarchies (Driscoll et al., 2020) | CBT | If children were less verbal |
|  |  |  |  | Parents allowed in room during intervention (Ollendick et al., 2021) | CBT | General rationale - to make intervention more suited to CYP with ASD |
|  |  |  |  | Parent-teen dyadic work was included to identify primary anxiety diagnoses and related goals (J. Reaven, Blakeley-Smith, Leuthe et al., 2012) | CBT | General rationale - to accommodate the needs of teenagers with ASD |
|  |  |  |  | The parent curriculum was modified to focus on the unique developmental challenges of adolescence, in addition to the provision of psychoeducation of anxiety and overview of CBT techniques and strategies (J. Reaven, Blakeley-Smith, Leuthe et al., 2012) | CBT | General rationale - to accommodate the needs of teenagers with ASD |
|  |  |  |  | The incorporation of parent-training. In addition to the child group session, a simultaneous psycho-educational parent group was included in which parents met with another therapist and were able to watch their children’s sessions (Scarpa et al., 2011) | CBT | To be developmentally appropriate for 5- to 7-year-old children with ASD. |
|  |  |  |  | Concurrent parent and child therapy groups where parents receiving psychoeducation about autism and emotion regulation and observe the children’s session (Swain et al., 2019) | CBT | Not reported |
|  |  |  |  | Involvement of parents or parents present in the sessions - taking child's clinical and developmental needs into account (Storch et al., 2013) | CBT | General rationale - to make treatment suitable for CYP with ASD |
|  |  |  |  | Parents were included for the majority of sessions (Storch et al., 2015) | CBT | To improve treatment motivation, to facilitate treatment progress and generalization, promote autonomy and communication skills, and problem solve treatment barriers. |
|  |  |  |  | Parents involved in the entire session (Storch et al., 2020) | FET | Teaching parents to serve as social support/coaches for exposure, enhance motivation and generalization, and reduce accommodation of anxiety |
|  |  |  |  | Parent support of home-based exposures (Wood et al., 2015) | CBT | Parental involvement in treatment programme |
|  |  |  |  | Parent psycho-educational sessions in which parents are helped to understand anxiety, develop appropriate strategies to deal with their own anxiety (E. Higgins et al., 2019) | CBT | Not reported |
|  |  |  |  | Weekly group parental Skype sessions for parents (Sofronoff et al., 2017) | CBT | Not reported |
|  |  |  |  | Involving a family member in sessions (Cooper et al., 2018) | CBT | Not reported |
|  |  |  |  | Use storytelling, perhaps including information from others (Fisher et al., 2023) (storytelling in EMDR involves the use of caregivers) | EMDR | General rationale: to address or accommodate barriers |
|  |  |  |  | Parents were often encouraged to be a part of therapy when possible (Wise et al., 2019) | CBT | Parental involvement is commonly suggested as beneficial when working with youth with ASD |
|  |  |  |  | Obtain information from other people as well as the person themselves (Fisher et al., 2023) | EMDR | General rationale: to address or accommodate barriers |
|  | Involving school | Involving or incorporating the child’s school into the intervention | 3 | Expanded the treatment focus to include explicit home–school collaboration. This included an active transfer of skills from the clinic to the home and school setting (e.g., via school consultation), as well as the delivery of therapeutic interventions such as a peer-mediated social intervention directly in the school setting (Fujii et al., 2013) | CBT | To improve functioning at school |
|  |  |  |  | Teacher handouts review strategies the child has learnt and provide the teacher with tips on how to support the child at school (Sofronoff et al., 2017) | CBT | Not reported |
|  |  |  |  | One to two consultations are offered to teachers or other school personnel in support of the overall exposure-therapy goals that may take place on school grounds (e.g., participating in peer interactions during lunch) employing the same behavioural support strategies offered to parents (Wood et al., 2015) | CBT | General rationale - to optimize treatment for individuals with ASD |

*Note.* **ADOS** = Autism Diagnostic Observation Schedule, **ASD** = Autism Spectrum Disorder, **CYP** = Children and Young People, **CBT** = Cognitive Behavioural Therapy, **DAWBA** = Development and Wellbeing Assessment**, EMDR** = Eye Movement Desensitisation and Reprocessing, **FET** = Family-based Exposure-focused Treatment, **SCQ** = Social Communication Questionnaire.

**Table S10.** GRADE Assessment for effectiveness outcomes (narrative synthesis)

| **N studies** | **Study quality** | **Concerns about certainty** | **Inconsistency** | **Concerns about certainty** | **Indirectness** | **Concerns about certainty** | **Imprecision** | **Concerns about certainty** | **Publication bias** | **Concerns about certainty** | **Certainty** |
| --- | --- | --- | --- | --- | --- | --- | --- | --- | --- | --- | --- |
| **Individual CBT for anxiety** | | | | | | | | | | | |
| 14 | 7/14 studies were of high, 6/14 was of moderate, and 1/14 studies was of low methodological quality. | No concern | 4/14 reported significant group differences in clinician-rated anxiety between CBT and TAU and 1/14 between CBT and waitlist, in favour of CBT at post, but not in parent- and child-reported anxiety between CBT and TAU in 1/4. 1/14 reported no significant group differences in child and parent-rated anxiety between CBT and waitlist post-treatment and at 6-month follow-up. 1/14 reported no significant differences in clinician-rated OCD between CBT and AM post-treatment. 1/14 reported significant pre-post reduction in self—reported anxiety and global functioning; 1/14 clinician-rated anxiety; 1/14 parent- and self-rated anxiety and social functioning but not in quality of life; 1/14 clinician-rated anxiety but not in self-reported anxiety and clinician-rated depression; 1/14 clinician- and parent-rated anxiety but not in child-report. 1/14 reported clinician-rated anxiety improvement post-treatment, maintained at 4 moths follow up (no statistical analysis), and 1/14 reported a decrease in child-report anxiety for treatment responders at 6 months follow-up, maintained by 2 participants at 12 month follow up (no statistical analysis). Therefore, there is minor inconsistency across the results of these studies. | Borderline | 14/14 studies used established rating scales. 7/14 measured effect of time. | Borderline | 14/14 had a sample size under 100 individuals. There were 14 contributing studies. | Serious Concern | No publication bias is suspected, as both significant and non-significant findings were found. | No concern | moderate certainty ⊕⊕⊕O |
| **Group CBT for anxiety** | | | | | | | | | | | |
| 19 | 10/19 studies were of high, 7/19 was of moderate, and 2/19 studies was of low methodological quality. | No concern | 1/19 reported no significant differences in child- and clinician-rated anxiety between CBT and SR at post, 3- and 6-month follow-up. 1/19 reported no significant difference in clinician-rated anxiety post-treatment and 1/19 both post and at 24-week follow-up, and 1/19 in parent-rated anxiety at post, all between CBT and waitlist. 1/19 reported no significant differences in clinician-rated anxiety between CBT standard manual training, workshop only and workshop plus at post. Significant differences in clinician- and child-rated anxiety (1/19); in parent-rated anxiety (1/19); in parent-rated but not in child-rated anxiety (1/19); and in parent- and child-reported anxiety reduction and clinician-reported anxiety severity but not in clinician-reported anxiety improvement scores or child-reported anxiety (1/19), all between CBT and waitlist in favour of CBT at post. Significant differences in clinician-rated anxiety and positive treatment response between CBT and TAU, favouring CBT at post (1/19). Significant differences in child- and parent-rated anxiety between CBT (child only), CBT (child + parent) and waitlist, favouring CBT (child + parent) at post (1/19). Significant pre-post reduction in self-reported anxiety (1); in clinician-reported anxiety (1); in parent- and child-rated anxiety and at 4-month follow-up (1); in clinician-, parent- and child-rated anxiety at post and 3-month follow-up (1). Significant pre-post decrease in parent-rated anxiety but not in child-report or parent- and child-reported anxiety to uncertain events (1/19). Significant pre-post reduction in parent- and child-rated anxiety, except in child-rated amount of worry and distress (1/19). Improvement in clinician-, parent and child-rated anxiety and social functioning over time (no statistical testing) (1/19). No change in self-reported anxiety (no statistical analysis) (1/19). Therefore, there is minor inconsistency across the results of these studies. | Borderline | 19/19 studies used established rating scales. 8/19 measured effect of time. 2/19 analysed group by time interaction, no randomisation.9/19 analysed group by time interaction | Borderline | 18/19 had a sample size under 100 individuals. There were 19 contributing studies. | Serious Concern | No publication bias is suspected, as both significant and non-significant findings were found. | No concern | moderate certainty ⊕⊕⊕O |
| **Combined individual and group CBT for anxiety** | | | | | | | | | | | |
| 3 | 2/3 studies were of high, 1/3 was of moderate methodological quality. | No concern | No significant differences in parent – and clinician-reported anxiety between CBT and waitlist (1/3). Significant increase in parent –reported anxiety in the CBT group over a 1-year follow-up but did not return to baseline levels (1/3). No significant differences in anxiety between CBT and counselling at post-treatment, but there were significantly fewer participants in the counselling group that met criteria for separation anxiety at post (1/3). Therefore, there is minor inconsistency across the results of these studies. | Borderline | 3/3 studies used established rating scales. 1/3 measured effect of time. 2/3 analysed group by time interaction. | Borderline | 3/3 had a sample size under 100 individuals. There were 3 contributing studies. | Serious Concern | No publication bias is suspected, as both significant and non-significant findings were found. | No concern | moderate certainty ⊕⊕⊕O |
| **Group interventions targeting emotion regulation** | | | | | | | | | | | |
| 5 | 2/5 studies were of high, 2/5 was of moderate, and 1/5 studies was of low methodological quality. | No concern | No significant difference between CBT and delayed therapy in parent-reported emotion regulation at post (1/5); No significant pre-post difference in parent-reported depression or parent- and child-reported quality of life (1/5); Significant pre-post reduction in parent-reported emotional lability/negativity in CBT group, but not in delayed therapy and no significant pre-post difference in parent-reported emotion regulation in both groups (1/5); 67% participants classified as treatment responders and there was a significant pre-post increase in parent-reported confidence in their child’s ability to manage anger and anxiety (1/5); Significant improvements in parent-rated anxiety and social and emotional regulation skills from pre to post and from pre to 6-week follow-up (1/5). Therefore, there is minor inconsistency across the results of these studies. | Borderline | 5/5 studies used established rating scales. 4/5 measured effect of time. 1/5 analysed group by time interaction | Serious concerns | 5/5 had a sample size under 100 individuals. There were 5 contributing studies. | Serious Concern | No publication bias is suspected, as both significant and non-significant findings were found. | No concern | low certainty ⊕⊕OO |
| **CBT for various mental health needs** | | | | | | | | | | | |
| 2 | 1/2 studies were of high, 1/2 was of low methodological quality. | Borderline | 2/2 reported no significant differences between CBT and waitlist in self-reported anxiety, depression and emotion regulation at post-treatment, thus the results are sufficiently consistent. | No concern | 2/2 studies used established rating scales. 1/2 analysed group by time interaction, no randomisation. 1/2 analysed group by time interaction | Borderline | 2/2 had a sample size under 100 individuals. There were 2 contributing studies. | Serious Concern | No publication bias is suspected. Only 2 contributing studies | Borderline | moderate certainty ⊕⊕⊕O |
| **Detection of autism** | | | | | | | | | | | |
| 3 | 2/3 studies were of high, 1/3 was of moderate methodological quality. | No concern | 2/3 studies report on tools that can identify individuals for whom specialised autism assessment is needed. 1/3 reported on a tool lacking suitability for screening. Therefore, there is minor inconsistency across the results of these studies. | Borderline | 3/3 studies used established rating scales to assess detection of autism | No concern | 2/3 had a sample size below 100. There were 3 contributing studies. | Serious Concern | No publication bias is suspected, as both significant and non-significant findings were found. | No concern | moderate certainty ⊕⊕⊕O |
| **Strategies for improving clinicians’ skills and knowledge of autism** | | | | | | | | | | | |
| 4 | 3/4 studies were of moderate, 1/4 was of low methodological quality. | No concern | 1/4 reported significant improvement in clinician self-efficacy, autism knowledge and awareness in best-practice treatment considerations for autistic people from pre- to post-treatment. 1/4 reported significant improvements in proportion with psychiatric disorders from referral to after 12 months, sustained at 24-27 months. 1/4 reported pre- to post-care pathway implementation reductions in number of youth restraints and 1/4 pre-pathway implementation and 18-month follow-up reductions in the number of brief stabilisation unit restraints, total restraints and youth with restrains and use of intramuscular medication, but not length of stay or number of inpatient restraints. | Borderline | 1/4 used established rating scales. 2/4 used established data collection methods to assess service use, i.e., inpatient length of stay from clinical records. 1/4 used less direct proxies for effectiveness. Overall, studies were heterogeneous in the measures of outcomes used. 4/4 measured effect of time. | Serious concerns | 3/4 had a sample size below 100. There were 4 contributing studies | Serious Concern | No publication bias is suspected, as both significant and non-significant findings were found. | No concern | low certainty ⊕⊕OO |

*Note****.* CBT** = Cognitive Behavioural Therapy, **OCD** = Obsessive compulsive disorder

**Method:** The Grading of Recommendations Assessment, Development and Evaluation (GRADE) system (Guyatt et al., 2008), adapted for narrative synthesis according to (Murad et al., 2017) and according to methodological aspects of research contributing to the research question about effectiveness of strategies to improve mental health care for autistic people. The certainty of evidence for each outcome was independently assessed by two people, after which they met to address any inconsistencies. Each GRADE domain could obtain ‘no concerns’, ‘borderline’ or ‘serious concerns’ rating and the overall certainty for each outcome started as high and was lowered for any ‘serious concerns’. The GRADE domains were rated accordingly:

- **Study quality** – ‘no concerns’ were noted if ≤ 33% of the contributing studies were of low quality, ‘borderline’ if 34-67% were of low quality, and ‘serious concerns’ if > 67% were of low quality based on quality ratings (see Table S7).
- **Inconsistency** – consistency of the direction of change and the magnitude of effects across the research evidence was evaluated. ‘No concerns’ were noted when most studies reported associations/effects in the same direction, or where there was only one contributing study and therefore the inconsistency was impossible to tell. ‘Serious concerns’ were noted when there was evidence of opposite directions of change (e.g., significant improvement and significant worsening), and ‘borderline’ concerns were notes when there was evidence of significant improvement/worsening and no significant change. The outcome of interest was mental health outcomes, particularly the mental health outcome being targeted with the intervention, and time X group interaction analyses were prioritised.
- **Indirectness** – a judgement was made on the degree of similarity of the research evidence with the research question of interest, reflecting on how directly the available evidence answered the specific research questions set out in the review. For example, measures of effect of time instead of group by time interactions and lack of randomisation contributed to down-ratings for this domain.
- **Imprecision** – a judgement was made based on the total number of contributing studies and their sample size. The sample size threshold used for the relevant analysis was 100. ‘Serious concerns’ were noted when there was only one contributing study, even if its sample size was above 100 individuals.
- **Publication bias** – we considered if studies contributing to an outcome reported significant and non-significant results, or if publication bias was likely due to missing evidence. ‘Serious concerns’ were assigned if there were only one contributing study per outcome, as this may have indicated a shift in research publication priorities.

**Table S11.** GRADE Assessment for effectiveness outcomes (meta-analyses)

| **No. of studies** | **Study quality** | **Concerns about certainty** | **Inconsistency** | **Concerns about certainty** | **Indirectness** | **Concerns about certainty** | **Imprecision** | **Concerns about certainty** | **Publication bias** | **Concerns about certainty** | **Certainty** |
| --- | --- | --- | --- | --- | --- | --- | --- | --- | --- | --- | --- |
| **CBT for anxiety: Child/Self-rater meta-analysis** | | | | | | | | | | | |
| 9 | 9/9 studies were of high methodological quality | No concern | There was significant heterogeneity among studies before removing outliers, Q(8)= 43.85, p < .001, I2 = 81.75%. I2 is 81.75%, thus there is considerable proportion of variation in point estimates. However, on removal of outliers (Chalfant et al. 2007; Langdon et al., 2016) heterogeneity reduced, Q(6) = 3.21, p = .782, I2 = 0%. Only one study has non-overlapping CI. Point estimates ranged from -0.37 to 2.64. Based on CIs, there were no significant group differences in child/self-reported anxiety post-treatment in all but one study which found significant group differences in favour of CBT. | Borderline | All studies were RCTs testing effectiveness of CBT for anxiety vs active/non-active control among autistic people. 9/9 studies used established rating scales for anxiety. | No concern | 9/9 had a sample size under 100 individuals. All studies have very wide CI around the estimate of the effect. | Serious concern | Egger’s test was significant (child/self z = 2.13, p = .033), suggesting the presence of publication bias. There were 9 contributing RCTs. | Borderline | moderate certainty ⊕⊕⊕O |
| **CBT for anxiety: Parent/carer-rater meta-analysis** | | | | | | | | | | | |
| 12 | 9/12 studies were of high, 2/12 of moderate and 1/12 of low methodological quality | No concern | There was significant heterogeneity among studies before removing outliers, Q(11) = 70.39, p < .001, I2 = 84.37%. I2 is 84.37%, thus there is considerable proportion of variation in point estimates. However, on removal of outliers (Chalfant et al., 2007; Murphy et al., 2007; A. J. Russell et al., 2013), heterogeneity reduced, Q(8) = 4.99, p = .76, I2 = 0%. Only one study has non-overlapping CI. Point estimates ranged from -0.44 to 4.27. Based on CIs, there were no significant group differences in parent-reported anxiety post-treatment in all but two studies which found significant group differences in favour of CBT. | Borderline | All studies were RCTs testing effectiveness of CBT for anxiety vs active/non-active control among autistic people. 12/12 studies used established rating scales for anxiety. | No concern | 12/12 had a sample size under 100 individuals. All studies have very wide CI around the estimate of the effect. | Serious concern | Egger’s test was significant (parent z = 4.70, p < .001), suggesting the presence of publication bias. There were 12 contributing RCTs. | Borderline | moderate certainty ⊕⊕⊕O |
| **CBT for anxiety: Clinician-rater meta-analysis** | | | | | | | | | | | |
| 12 | 10/12 were of high and 2/12 were of moderate methodological quality | No concern | There was significant heterogeneity among studies before removing outliers, Q(11) = 35.13, p < .001, I2 = 68.69%. I2 is 84.37%, thus there is substantial proportion of variation in point estimates. However, on removal of outliers (Storch et al., 2020; Reaven et al., 2018) heterogeneity reduced, Q(9) = 8.58, p = .477, I2 = 0%. All studies had overlapping CI. Point estimates ranged from -0.47 to 2.13. There were no significant group differences in clinician-reported anxiety post-treatment in all but four studies which found significant group differences in favour of CBT. | Borderline | All studies were RCTs testing effectiveness of CBT for anxiety vs active/non-active control among autistic people. 12/12 studies used established rating scales for anxiety. | No concern | 12/12 had a sample size under 100 individuals. All studies have very wide CI around the estimate of the effect. | Serious concern | Egger’s test was significant (clinician z = 3.99, p < .001), suggesting the presence of publication bias. There were 12 contributing RCTs. | Borderline | moderate certainty ⊕⊕⊕O |

*Note****.* CBT** = Cognitive Behavioural Therapy, **CI** = Confidence intervals, **RCT =** Randomised controlled trial.

**Method:** The Grading of Recommendations Assessment, Development and Evaluation (GRADE) system (Guyatt et al., 2008) was used to evaluate the quality of effectiveness evidence contributing to each of the three meta-analyses depending on the rater (i.e., child/self, parent/carer and clinician). The certainty of evidence for each outcome was independently assessed by two people, after which they met to address any inconsistencies. Each GRADE domain could obtain ‘no concerns’, ‘borderline’ or ‘serious concerns’ rating and the overall certainty for each outcome started as high and was downgraded for any ‘serious concerns’. The GRADE domains were operationalized accordingly:

- **Study quality** – the proportion of trials that were of low methodological quality was considered (See Table S7).
- **Inconsistency** – heterogeneity or variance of point estimates across trials and overlap of confidence intervals (CI) was considered.
- **Indirectness** - a judgement was made on the degree of similarity of the research evidence with the research question of interest, reflecting on how directly the available evidence answered the specific research questions of each meta-analyses.
- **Imprecision** - a judgement was made based on the trials’ sample size (the sample size threshold used for the relevant analysis was 100) and trials’ width of CIs around the estimate of the effect.
- **Publication bias** – the number of contributing studies as well as visual inspection of the funnel plots and results of the statistical test for asymmetry were considered.

**Table S12.** Full results by study

*Adapted and bespoke mental health interventions*

| **Authors** | **Strategy vs comparison** | **Outcomes/measures** | **Adaptation categories and sub-categories** | **Acceptability/Feasibility findings** | **Effectiveness findings** |
| --- | --- | --- | --- | --- | --- |
| **CBT for anxiety** | | | | | |
| Sung et al. (2011) | Adapted group CBT for anxiety vs adapted Social recreational programme | Primary and secondary outcomes not specified.  SCAS-C - Child -self report measuring severity of anxiety symptoms in children.  CGI-S - Clinician rated measure of severity of the patient's illness. | Accommodate individual differences - Tailor practice to individual needs and preferences | Feasibility - Intervention attendance rate: 33/36 received allocated CBT intervention, and 31/34 received allocated comparison Social Recreational programme. - Intervention drop-out rate: 6/70 randomised in one of two groups did not complete the 16-week programme (3 in each of the groups) due to lack of interest, clashes with school schedules, preferred enrolment in other services, and changes in dosage of medication due to other co-occurring psychiatric conditions. A further 5 had dropped out at 3 and 6 months, 3 from the CBT and 2 from Social Recreational group. | Mental Health Outcomes using SCAS-C and CGI-S: - SCAS-C (completers analyses): No significant main effect for Group and interaction effects for Time x Group was found, indicating that both groups did not differ across time for child reports on anxiety. Analyses from the repeated measures showed that there were signiﬁcant main effects for Time for panic attack, F (3, 134) = 6.10, p = 0.001, n2= .11, generalized anxiety, F (3, 129) = 3.28, p = 0.03, n2= .06, and total anxiety score, F (3, 124) = 3.03, p = 0.04, n2 = .06. Post-hoc comparisons of the main effects showed that both groups reported signiﬁcantly fewer generalized anxiety and total anxiety symptoms at 6 months. Children in the Social Recreational group also reported signiﬁcantly fewer total anxiety symptoms at post-treatment and signiﬁcantly fewer panic attack symptoms at 6 months. - CGI-S (completers analyses): No significant differences between the two groups at different time points for severity of anxiety. In both groups, the percentage of participants in the ‘‘Normal’’ range increased from pre-treatment to 6-month follow-up, with a range of 6.06 to 37.04%. In addition, the percentage of participants in the ‘‘Moderately ill’’ range decreased from pre-treatment to 6-month follow-up, with a range of 3.70 to 29.03%. At post-treatment, 0% of participants in the Social Recreational group were markedly ill, indicating an improvement of 9.68%. In the CBT group there were no participants who were markedly ill.  - The sample size for intent-to-treat was 70. Participants’ last available scores were carried forward to post-treatment (T2) and 3-month follow-up (T3). All significant findings for treatment completers were maintained in intent-to-treat analysis. |
| E. Higgins et al. (2019) | Adapted group CBT for anxiety (“Special FRIENDS” programme) | Primary and secondary outcome measures not specified.  SCAS - child and parent-report measure of child's anxiety symptoms.  Feedback form - parents completed this form at post intervention, rating and commenting on observed changes in their child’s anxiety levels, their child’s enjoyment of the programme and ideas for programme improvement. Topic guide for semi-structured interviews with parents to gather qualitative feedback on the impact of the programme. | Accommodate individual differences - Tailor practice to individual needs and preferences Structural or procedural adaptations - Format of the intervention - A structured and predictable approach Intervention content adaptations - Use of role play or modelling Involving wider support network - Parental involvement | Feasibility - Intervention attendance rate: 9-12/12 participants attended each week, with attendance rates falling in Week 8 to 5/12 participants due to Easter Holidays. Weekly session summaries recorded high overall session attendance by children and parents.  - Drop-out rate: No one dropped out during treatment. All children and their parents completed assessments at pre- and post-intervention. 9/12 children and their parents completed assessment at 4-month follow-up. Acceptability - Experience of care: parents revealed their child enjoyed their time in attendance with six parents rating their child’s overall enjoyment of the programme as “excellent” and five rated their child’s enjoyment as “good” (N = 12). In relation to impact, data revealed three subthemes; 2 (a) decreased anxiety levels, 2 (b) development of CBT skills and 2 (c) opportunity for social interaction. The prepared handout was identified as helpful in the preparation for upcoming sessions with the sessions also allowing for the development of a parental network in some cases. Parents described how their child felt accepted in the group and developed awareness that others may be experiencing and feeling the same way. Barriers to success were also identified and these were the timing of the sessions which occurred during school time and the reluctance of children to talk to parents about content outside of the sessions. | Effect of time (no comparison group)  Mental health outcome using SCAS: - There was an overall improvement in children’s Total Anxiety scores from pre- (T1 M= 38.33, SD= 10.83) to 1-month post-intervention (T2 M= 29.67, SD= 6.56) and 4-month follow-up (T3 M= 26.67, SD= 7.66), F(2, 16) = 4.02, p= 0.04, g(T1 toT2) = 0.93, g(T1 to T3) = 1.2, g(T2 to T3) = 0.62.  - There was an overall improvement in parent’s-rated child's Total Anxiety scores from pre- (T1 M= 37.89, SD= 14.64) to 1-month post-intervention (T2 M= 34.44, SD= 15.57) and 4-month follow-up (T3 M= 28.67, SD= 13.86), F(2, 16) = 3.88,p= 0.04, g(T1 to T2) = .22, g(T1 to T3) = .62, g(T2 to T3) = .38. |
| Kilburn et al. (2019) | Adapted group CBT for anxiety ('Cool Kids' program) | Primary and secondary outcome measures not specified.  ADIS-C/P - clinician-rated measure assessing parent and child endorsement of disorder.  SCAS - child and parent-report measure of child's anxiety symptoms.  CATS - measure of automatic thoughts completed by children ESQ - measuring children and parents’ satisfaction with the treatment CALIS - measure of anxiety’s impact on life completed by child and parents | Communication Accommodations     - Use of simple, written material and visual aids   Accommodate individual differences - Tailor practice to individual needs and preferences Structural or procedural adaptations - Format of intervention Intervention content adaptations - Simplified content Involving wider support network - Parental involvement | Feasibility - Intervention attendance: 6/9 children attended all 12 sessions. 2/12 children attended 11 sessions. 1 child only attended 8 sessions. The 9 children participating completed a mean of 11.3 out of 12 sessions. One child only participated in 8/12 sessions due to difficulties being in a group setting. One child managed only to participate in 8 out of 12 sessions due to difficulties in being in a group session.  - Drop-out rate: No one dropped out before treatment. Only 1 person dropped out between post-intervention and 3-month follow-up.  - Intervention fidelity: All sessions assessed (a random selection of 3 out of the complete 12 sessions for each running programme) adhered to the manual. Acceptability - Satisfaction with care: 7 children and 8 parents completed the ESQ and showed overall satisfaction with the programme. Most parents (83%) and children (61%) gave maximum positive points to the different statements. | Effect of time (no comparison group)  Mental Health Outcomes using the ADIS-C/P, SCAS, CALIS, CATS: - No significance testing was employed due to the small number of participants. - Results from baseline (pre-) to post-treatment showed an improvement on the severity of the primary anxiety diagnosis with a large effect size (d = 1.25). The reduction in CSR from post-treatment to 3-month follow-up showed a medium effect size (d = 0.46). For all anxiety diagnoses, a large effect size in the overall mean CSR scores was evident from pre- to post-treatment (d = 1.70), whereas the effect size post-treatment to 3-month follow-up (d = 0.23) was small. The percentage of children recovered and no longer meeting the criteria for any anxiety diagnosis on the ADIS/CP after the treatment was 22.2%. This number rose to 55.5% at 3-month follow-up. Following treatment, 55.5% of the children had recovered from their primary anxiety diagnosis and at 3-month follow-up this was evident for seven out of nine children (77.7%). - The results from the self-reported questionnaire on anxiety symptoms (SCAS) showed an improvement from pre- to post-treatment in both the children’s responses (d = 0.71) and the parent’s responses (d = 0.67). There was no further improvement from post-treatment to follow-up according to both the child (d = 0.00) and the parent questionnaire (d = -0.00).  - Children’s negative self-statements measured with CATS showed a large positive effect from pre- to post-treatment (d = 0.95) but no effect from post-treatment to follow-up (d = -0.05). Social outcome using CALIS:  - For the children, results for the anxiety’s impact on life (CALIS) showed a very small positive effect (d = 0.04) from pre- to post-treatment. However, in the post-treatment to follow-up period actual worsening was detected (d = -0.29). No significance testing. |
| Kilburn et al. (2020) | Adapted group CBT for anxiety ('Cool Kids' program) vs Waitlist control | Primary outcome measures: ADIS-C/P - clinician-rated measure assessing parent and child endorsement of disorder. Secondary outcome measures: SCAS - child and parent-report measure of child's anxiety symptoms.  CATS - measure of automatic thoughts completed by children CALIS - measure of anxiety’s impact on life completed by child and parents ESQ - measuring children and parents’ satisfaction with the treatment | Communication Accommodations     - Use of simple, written material and visual aids   Accommodate individual differences - Tailor practice to individual needs and preferences Structural or procedural adaptations - Format of intervention Intervention content adaptations - Simplified content Involving wider support network - Parental involvement | Feasibility - Intervention attendance rate: 21/25 (84%) completed CBT. For CBT group the mean number of sessions attended was 8.28 (SD=2.72). In the waitlist group 20 children were offered CBT treatment after the waiting list period and 15 (75%) completed. Hence, of the 45 children offered CBT, 36 (73%) completed it. The mean number of sessions attended by the completers was 9.44 (SD=0.73).  - Drop-out: 4/25 (16%) children-parent dyads attended less than 7 CBT sessions (non-completers). Additionally, one family in the CBT group did not provide post measurement. During the waitlist period 4/24 (16.6%) families withdrew. Additionally, in the waitlist group, one child did not accept the CBT offer and 4 discontinued the program after attending only a few sessions due to not being able to attend a group setting (n=3) or wanting to seek other treatment (n=1).  - Intervention adherence: Of the 100 intervention sessions conducted for the CBT and the waitlist group after wait-list (10 groups, 10 sessions each), 20 sessions were coded for adherence to program: 90% met criteria of including 90% or more of the required session elements. Acceptability - Treatment satisfaction: of all parent-child dyads who completed the programme, 19 (53%) of the children and 20 (56%) of the parents completed the satisfaction questionnaire. 13 (68%) of the children and 14 (70%) of the parents felt that the treatment had helped the children. 10 (57%) of the children and 20 (100%) of the parents would recommend the treatment to others with the same problems. | Primary Outcome Mental health outcomes using ADIS-C/P: - ADIS-CSR primary diagnoses: Time by condition effect = -0.82, p = 0.202 (non-significant); CBT pre-to-post differences: p < 0.001, d = 1.05; WL pre-to-post differences: p = 0.012, d = 0.71. - ADIS number of anxiety diagnoses: Time by condition effect: -0.39, p = -0.279 (non-significant); CBT pre-to-post differences: p = 0.011, d = 0.59; WL pre-to-post differences: p = 0.325, d = 0.33. Secondary Outcomes Mental health outcomes using SCAS, CATS: - SCAS-children: Time by condition effect: -7.22, p = 0.044 (significant); CBT pre-to-post differences: p = 0.077, d = 0.33; WL pre-to-post differences: p = 0.281, d = -0.20. - SCAS-parents: Time by condition effect: -10.12, p = 0.005 (significant); CBT pre-to-post differences: p < 0.001, d = 0.56; WL pre-to-post differences: p = 0.506, d = -0.17. - CATS- social threat: Time by condition effect: -4.13, p = 0.069 (non-significant); CBT pre-to-post differences: p = 0.014, d = 0.40; WL pre-to-post differences: p = 0.911, d = 0.02. - CATS - physical threat: Time by condition effect: 2.85, p = 0.214 (non-significant); CBT pre-to-post differences: p = 0.129, d = 0.40, WL pre-to-post differences: p = 0.809, d = -0.04. Social outcome using CALIS:  - A clinically relevant, but not statistically significant effect of time by condition was found on the scale measuring the parent’s assessment of child's anxiety interference on the child’s life (p=0.078), together with a statistically significant moderate effect from pre- to post-treatment in the CBT condition (d=0.47, p=0.026). No effect of time by condition was found on the scale measuring children’s opinion of interference of anxiety in own life (p=0.701) but a small positive effect from pre- to post-treatment was found in the CBT group (d=0.11, p=0.685). |
| Chalfant et al. (2007) | Adapted group CBT for anxiety (‘Cool Kids’ programme) vs Waitlist control | Primary and secondary outcome measures not specified.  ADIS-C/P - clinician-rated measure assessing parent and child endorsement of disorder. RCMAS - child-reported measure of child’s chronic anxiety (trait). SCAS - child-report measure of child's anxiety symptoms. CATS - measure of automatic thoughts completed by children | Communication Accommodations     - Use of simple, written material and visual aids   Accommodate individual differences - Tailor practice to individual needs and preferences Structural or procedural adaptations - Format of intervention Intervention content adaptations - Simplified content Involving wider support network - Parental involvement | Feasibility - Intervention drop-out: 4 participants dropped out of the intervention. Participants were considered to have dropped out of the CBT program if they attended up to the first two sessions and then discontinued from the program. Two of the participants dropped out because their parents’ work schedules did not permit regular attendance, one of the participants dropped out because they moved out of area of the treatment centre and one of the participants dropped out because they felt the group was of no benefit to them.  - Intervention attendance: 28 CBT completers | Mental Health Outcomes pre-post using the ADIS-C/P, RCMAS, SCAS, CATS: - A significant Group x Time interaction F (1,45) = 63.792, p < 0.001, g2 = .586 was found indicating a significantly greater reduction between pre- and post-treatment in the number of anxiety disorder diagnoses for participants in the CBT condition than for those on the waitlist. A significant main effect was found for Time F (1,45) = 73.661, p < 0.001, g2 = .621 indicating an overall reduction in the number of anxiety disorder diagnoses across the CBT and waitlist conditions.  - A significant difference was found between the CBT and waitlist groups in their number of post-treatment anxiety diagnoses, t (1,45) = 5.074, p < 0.01. The CBT group had a significantly smaller number of anxiety diagnoses at post-treatment than the waitlist group. Follow-up paired t-tests indicated a significant difference between the number of pre- and post-treatment anxiety diagnoses for the CBT group but no significant differences between the number of pre- and post-treatment anxiety diagnoses for the WL, t (1,27) = 10.408, p < .01 and t (1,18) =1.000, p > 0.05, respectively.  - For the RCMAS, the ANOVA revealed a significant Group X Time interaction, F(1,45) = 71.145, p < 0.005, g2 = .613, showing a significantly greater reduction between pre- and post-treatment in self-reported anxiety for participants in the CBT condition than for those in the waitlist and a significant main effect for time, F(1,45) = 96.888, p < .005, g2 = .683, demonstrating an overall reduction of self-reported anxiety for both CBT and WL conditions.  - For the SCAS, the ANOVA revealed significant main and interaction effects, F (1,45) = 58.127, p< .005, g2= .564, and F (1,45) = 51.544, p< 0.005, g2= .534, respectively. The significant interaction demonstrated a significantly greater reduction between pre- and post-treatment in the anxiety symptoms for the CBT condition than for the WL. The main effect for time revealed an overall reduction of self-reported anxiety symptoms for all the conditions.  - On the CATS Internalising Thoughts Scales, there was a significant main effect for time, F (1,45) = 37.583, p< 0.005, g2= .455, showing an overall reduction in self-reported internalising thoughts about worries and self-esteem for both the CBT and waitlist condition. |
| J. Reaven, Blakeley-Smith, Leuthe et al. (2012) | Adapted group CBT for anxiety (Facing Your Fears-Adolescent Version - FYF-A) program) | Primary and secondary outcomes not specified.  ADIS-C/P - clinical interviews assessing the presence of anxiety disorders SCARED - Parent and child self-report. measure assessing clinical anxiety symptoms. CGI-S - to assess global severity for the four anxiety diagnoses. Assessed by clinical psychologists by reviewing parent report on the ADIS and SCARED.  CGI-I - to assess treatment responsiveness. Assessed by clinical psychologists by reviewing parent report on the ADIS and SCARED. | Communication accommodations - Use of technology Structural or procedural adaptations - Format of the intervention Intervention content adaptations - Integration of social skills training Involving wider support network - Parental involvement | Feasibility - Attendance: 7/24 families attended 100% of group treatment sessions, 13/24 attended 92.8%, 3/24 attended 85.7%, 1/24 attended 78.6% of sessions. - Drop out: Of the 26 families that started treatment, 2 dropped out before session two. Acceptability - Satisfaction: Of the 15 parent activities, 14 (93%) were rated as “helpful” or “very helpful.” Of the 14 teen activities, 13 (93%) were rated as “somewhat helpful” or higher. | Effect of time (no comparison group)  Mental Health Outcomes using CGI-S, CGI-I, SCARED and ADIS-C/P: - All paired t-tests revealed significant reductions in total anxiety symptoms as reported by parents from pre-treatment to post-treatment (t = 2.875, p = 0.009) and from pre-treatment to 3-months follow-up (t = 3.821, p = 0.001) using SCARED. - Significant reductions in anxiety symptoms were also reported by teens from pretreatment to posttreatment (t =3.896, p = .001) and from pretreatment to follow-up (t = 3.032, p = .008) using SCARED.  - There was a significant difference in clinician rating of anxiety severity (CGI-S), based upon parent report (Z =2.53, p = 0.011), meaning severity of anxiety symptoms significantly decreased post-intervention. - Nearly 46% of teen participants (11/24) obtained a CGI-I score reflecting a positive treatment response, 33% (8/24) obtained a CGIS-I score indicating “some” improvement, 21% (5/24) obtained a CGIS-I score indicating “no change” in symptoms, and no participants’ symptoms worsened postintervention (Cohen’s d = 0.90) |
| Burke et al. (2017) | Adapted group CBT for anxiety ('FRIENDS for life' programme) | Primary and secondary outcome measures not specified.  BAY-I - self-reported anxiety. Parental interviews post intervention. Participant evaluation questionnaire with Likert scale.  Narrative observation of interventions. | Adjustments to the physical environment - Provide environment and practical adjustments - Encourage the use of sensory resources and stimming Communication accommodations - Use of simple, written material and visual aids Structural or procedural adaptations - A structured and predictable approach Intervention content adaptations - Use of role play or modelling - Integration of social skills training | Acceptability - Satisfaction of care: Feedback from children post-intervention was favourable, where 85% reported that they would engage in the FRIENDS programme again. 71% of children identified that they “really enjoyed” the programme while one child (14%) noted that they “liked it”. One participant (14%) reported that he “did not enjoy it”. | Effect of time (no comparison group) Mental Health Outcomes pre-post using the BAY-I: - Of the seven children who participated, only one self-reported “mildly elevated anxiety symptoms” before engaging with the FRIENDS programme. Post-intervention, this child remained within the same range. All other children (n = 6) presented within the “average range” of anxiety pre- and post-intervention. |
| Cook et al. (2017) | Adapted group parent-mediated CBT for anxiety (‘Fun with Feelings’ programme) for children aged 4-6 years vs Waitlist control | Primary and secondary outcome measures not specified.  PAS - parent-report measure assessing the severity of child's anxiety symptoms. CBCL-Anx - parent report measuring frequency of child anxiety problems. Treatment satisfaction - parent-rated. | Communication adaptations: - Use of simple, written material and visual aids  Structural or procedural adaptations - A structured and predictable approach Intervention content adaptations - Creative outlets and activities - Use of play or role modelling - Taking it slow - Integration of emotion-focused strategies | Acceptability - Treatment satisfaction: Parents appeared generally satisfied with the programme, with means for all but one of the satisfaction items being 3 or above on a 5-point scale. | Mental Health Outcomes using the PAS & CBCL-Anx: - PAS: due to a computer malfunction, PAS was only used at pre-assessment for eligibility, even though it was planned as the primary outcome measure. - CBCL-Anx: there was no significant time or group × time effects for child anxiety, suggesting that neither group improved over time from pre- to post-treatment for the completer sample (n=24 parents): pre-treatment: CBT group M=67.71, SD=12.63; waitlist M= 75.58, SD=8.50; post-treatment: CBT group M= 66.07, SD =10.17; waitlist M= 74.17, SD =10.2, p = 0.938. With respect to intention to treat analysis (n=31) conducted from pre- to post-treatment, there were no significant group × time effects for child anxiety.  - 3-month follow-up (time effect of CBT group): reductions on the CBCL-Anx scale from pre-treatment to 3-month follow-up approached significance (p= 0.058), with a moderate effect size for completers sample, but there was no significant difference between post-treatment to 3-month follow-up (p=0.157). There were no significant time effects at three-month follow-up for the intent to treat sample. |
| Hepburn et al. (2016) | Adapted group CBT for anxiety using videoconferencing (Telehealth Facing Your Fears - FYF) Intervention) vs Waitlist control | Primary outcome SCARED - parent report of youth's anxiety | Communication Accommodations     - Use of simple, written material and visual aids Structural or procedural adaptations - A structured and predictable approach  Intervention content adaptations - Use of role play or modelling  - Taking it slow | Feasibility - Attendance: Overall attendance was approximately 94%. Of the 16 families who participated fully, 8 attended 100% of the sessions, 2 missed 2 sessions, and 6 missed 1 session. 16/17 families completed the 10-session treatment course.  - Drop-out: 1/17 (5.9%) families did not complete the 10-session treatment.  - Usability of technology: Of the 138 videoconferencing sessions conducted to consent, complete assessments, or engage in intervention, 8 (5.8%) were significantly impacted by technical glitches, resulting in one or more families calling in by telephone to participate. Moderate technical glitches (defined as a brief disconnection with successful re-joining within a few minutes) were more common, with an average occurrence rate of 0.87/session. Of 17 families, 7 (41%) were disconnected at least one time during the 10-session intervention. 2 families experienced recurrent connection problems during the first five sessions.  - Fidelity of implementation: Therapist fidelity to the critical elements of the FYF approach was 92.1%. Sessions involving graded exposure tended to be less faithful to the manualised FYF protocol. Acceptability - Satisfaction with care: mean satisfaction score for parents was 92.9%, suggesting high parent satisfaction with the intervention content, delivery method, and alliance with therapist. When asked if they would recommend the program to a friend, 100% of the parents responded affirmatively. The mean Satisfaction score for youth was 88.8%. Younger children tended to report higher levels of enjoyment in the activities than most of the participating adolescents. When asked if they would recommend the program to a friend, 11/14 (79%) of participating youth said ""yes"". | Primary outcome:  Mental health outcome using SCARED: - There was a statistically significant difference between the Telehealth FYF and Comparison Group over time on parent report of youth anxiety, F (1, 31) = 8.73; p = 0.006; Eta squared = 0.22. Telehealth FYF group: Pre-treatment M(SD): 30.94 (9.35); Post-treatment M(SD): 26.76 (8.50). Waitlist comparison: Pre-treatment M(SD): 28.25 (11.95); Post-treatment M(SD): 32.13 (12.99). |
| Bemmer et al. (2021) | Adapted group CBT for social anxiety. | Primary outcome measures  LSAS-SR - self report measure of anxiety and avoidance of social situation.  Secondary outcome measures  DASS-21 - self-report measure of symptom severity of depression, anxiety, and stress.  K10 - self-report measure of psychological distress.  SIAS - self-report measure of social anxiety.  SPS - self-report measure of social anxiety.  Tolerability measures - self-report at the mid-point of treatment, assessing expectations of, and engagement with the intervention, and potential barriers. | Intervention content adaptations - Simplified content  - Integration of cognitive-behavioural approaches  - Integration of social skills training | Feasibility  - Attendance rate: 78/84 completed intervention  - Drop-out rate: 6/84 (8%) participants dropped out.  Acceptability  - Experience of care at mid-point (quantitative): 96% of 28 participants agreed or strongly agreed that they are enjoying the group.  - Experience of care at mid-point (qualitative): Participants indicated that making friends, being able to talk and ask questions without judgement, feeling understood by others and having practical help and support were working well within the group. Participants indicated that making phone calls, feeling like their anxiety was hindering their learning, and finding the groups either too long or too short were difficulties with the group. Participants generally reported positive engagement with other group members, though some reported difficulties with a group member who was perceived as too talkative and disruptive. | Effect of time (no comparison group)  Primary outcomes  Mental health outcome using the LSAS-SR:  - Significant improvements in anxiety and avoidance of social situations (total score) from pre (M = 79.78, SD = 27.36), to post (M = 70.17, SD = 31.04), M change = 9.61 (SD = 20.41), p < .001.  Secondary outcomes  Mental health outcomes using DASS-21; K10; SIAS; & SPS:  - Significant improvements in social anxiety related to initiating and maintaining conversations, depression, anxiety, and stress from pre- to post-intervention, but not in social anxiety related to fears of being observed or evaluated in daily activities and psychological distress. |
| Langdon et al. (2016) | Bespoke group CBT for anxiety vs waiting list | Primary Outcome Measure  HAM-A - clinician-rated scale of anxiety symptoms.  Secondary Outcome Measures  SPI - self-report measure of behavioural, physiological, and cognitive symptoms associated with social phobia.  LSAS - self-report measure of fear and avoidance throughout 24 listed situations likely to elicit social anxiety.  Social and Emotional Functioning Interview (Informant and Subject Versions) - semi-structured clinician-rated assessment of everyday social and psychiatric functioning.  SIAS - self-report measure of anxiety as experienced in social situations associated with social anxiety and social phobia.  Fear Questionnaire - self-report assessing individual perception of fears and phobias.  HAM-D - clinician-rated interview assessing depression symptom severity.  Interviews about experiences of care: Following the completion of the trial, participants were interviewed, and asked to rate nine questions on a 5-point Likert Scale about their experience of receiving therapy. Participants were also asked (1)‘What were you hoping for by taking part in this research study?’, (2)‘What was best about the group?’, (3)‘What was worst about the group?’, (4)‘What advice would you give for the next group?’ and (5)‘Were there any difficulties you feel that the group did not address?’ | Not applicable. | Feasibility  - Intervention attendance rate: M = 13.3 treatment sessions, SD = 7.17  - Intervention drop-out rate: During the trial, seven participants were lost (5 from intervention arm and 2 from control arm), representing an attrition rate of 13%.  Acceptability  - Experience of receiving care (quantitative): 53% of the participants agreed or strongly agreed that the individual sessions that were initially offered helped prepare them for the group sessions. 59% of the participants agreed or strongly agreed that they now knew how to reduce their feelings of anxiety following treatment. However, 38% of participants thought there was insufficient time during sessions and 41% thought there were too few sessions. 79% of participants agreed or strongly agreed that they found listening to the problems of others helpful, while nearly 80% agreed or strongly agreed that they felt supported by other group members. 56% agreed or strongly agreed that therapy reduced their anxiety, while 44% were neutral, disagreed, or strongly disagreed on this. 73% of participants agreed or strongly agreed that they would recommend therapy to others, and 73% agreed or strongly agreed that therapy was helpful.  - Experience of care (qualitative): 1) ‘Motivation to take part’. Participants described taking part in the trial to access help for their mental health problems, while others had hoped that they might form new relationships with other people with ASDs. 2) ‘Positive experiences’. Participants described that they enjoyed 'interacting with the others'. 3) ‘Negative experiences’. Many participants were clear that they wanted to have had longer sessions. Others spoke about issues around the dynamics of being in a group, with one participant stating, ‘the group could be easily hijacked’. Several spoke about needing more continuity and greater focus on making sure the sessions flowed more effectively, while there were a few participants who commented that they found taking part in a group very difficult and thought the whole experience was negative. 4) ‘Further adaptations’. Participants indicated they may benefit from more individual sessions, and the suggestion to alternate between blocks of both group and individual sessions might improve treatment efficacy. This would also help to ensure that clients are afforded sufficient time to address their difficulties. Participants asked for more innovative homework options, using technology. 5) ‘Pragmatic issues’. Participants told us that there were sometimes issues with public transport, travelling, the timings of the group, heating in the rooms and difficulties with parking, all of which they did not like." | Primary outcome  Mental health outcome using HAM-A:  HAM-A mean scores significantly improved over time, regardless of arm, and regardless of baseline scores, p < 0.001. There was no significant difference between the treatment and waitlist arms at either follow-up 1 (after initial 24 weeks of treatment) or 2 (after further 24 weeks of treatment) on the HAM-A, after controlling for baseline scores.  Secondary outcomes  Mental health outcomes using SPI; LSAS; SIAS; Fear Questionnaire; & HAM-D:  Controlling for baseline scores, there was no significant difference between the treatment and waitlist arms on any of the secondary outcomes at follow-up 1 or 2. There was a significant improvement over time, regardless of arm, and baseline scores, on the HAM-D, p = .008; Fear Questionnaire total phobia score, p = 0.019; LSAS Avoidance, p = 0.003; LSAS Fear/Anxiety, p < 0.002. There was a significant improvement over time, regardless of arm, and baseline scores on the SIAS, p < 0.001 and SPI, p = 0.007.  Social outcomes using Social and Emotional Functioning Interview (Informant and Subject Versions):  Controlling for baseline scores, there was no significant difference between the treatment and wait list arms on any of the secondary outcomes at follow-up 1 or 2. There was a significant improvement over time, regardless of arm, and baseline scores on the Social/Emotional Functioning Interview–Informant, p < .001; and Social/Emotional Functioning Interview–Subject Versions, p < .001. |
| Sofronoff et al. (2005) | Bespoke group CBT for anxiety (Exploring Feelings) (child only) vs brief CBT for anxiety (child + parent) vs Waitlist | Primary and secondary outcomes not specified.  James and the Maths Test' - child generates strategies for James to cope with anxiety in a situation, a therapist reported scores.  SCAS-P - parent-report measure assessing how likely a parent feels their child would feel anxious in a situation. SWQ-P - parent-report measure to gauge levels of social worry experienced by the child. | Not applicable. | Feasibility - Intervention drop-out rate: 5 children did not complete intervention (1 from CBT for anxiety - child only, 3 from waitlist and 1 from CBT for anxiety - child and parent)  - Attendance rate: 22/23 completed CBT-child only, and 24/25 completed CBT-child and parent. - Treatment fidelity: 25% of sessions were assessed by independent reviewers, of which 98% of material was found to have been covered accurately. Acceptability - Satisfaction with care: Qualitative reports by parents indicated that children developed friendships, were more confident in their day-to-day interactions and that the time spent with children similar to themselves and therapists had helped in this regard. Parents also reported that children became distressed more slowly and recovered more quickly when issues arose. Parents reported feeling competent with the content of the programme and therefore able to help their child, empowered by meeting parents with similar experiences, supported by the group and able to share problems and solutions. | Mental health outcomes using SCAS-P, James and the Maths test and SWQ: - There was a significant time x group interaction on SCAS-P, F (4,158) = 9.16, p < .001. Post-hoc comparisons showed that participants in CBT child only intervention group significantly improved from pre (M = 40.23, SD = 20.42) to six week follow-up (M = 29.42, SD = 15.3), p < .001, and participants in CBT child and parent intervention group significantly improved from pre (M = 35.25, SD = 16.44) to six week follow-up (M = 21.11, SD = 10.1), p < .001. There was also a significant difference between the two intervention groups at follow-up, with CBT child and parent showing greater improvement. SCAS-P subscales of separation anxiety, OCD, social phobia and generalised anxiety showed significant time x group interactions, with improvements seen in both intervention groups. - There was a significant time x group interaction on James and the Maths test, F (4, 158) = 28.31, p < .0001. Significantly more strategies were generated in both intervention groups compared to the waitlist group both at post and six-week follow-up, p < .001. There was also a significant difference between the two intervention groups at post and six-week follow-up, with CBT child and parent group generating significantly more strategies, p < .001.  - There was a significant time x group interaction on SWQ, F (4, 156) = 11.06, p < .0001. Post-hoc comparisons showed significant improvements in both CBT child only group (p < .001) and CBT child and parent group (p < .0001) from pre to six-week follow-up. There were significant differences between the two intervention groups and the waitlist group at six-week follow-up, but not between the two intervention groups. |
| McConachie et al. (2014) | Bespoke group CBT for anxiety (Exploring Feelings) immediate therapy vs delayed therapy | Primary outcomes measures: ADIS child version: structured clinical interview allowing primary diagnosis of an anxiety disorder. SCAS-C - self-report measure of anxiety in children. SCAS-P - parent-report measure of anxiety in children. CGI-I - Using all interview and questionnaire information from baseline and 3-month end point, clinicians rated how much the child’s anxiety had improved. Summary question: at 3-month end-point assessment, each parent and child separately answered the written question "Over the past 3 to 4 months do you think your [your child's] anxiety has reduced? (yes/no)".   Qualitative interviews to discuss experiences of therapy and questionnaire about information on other experiences. | Not applicable. | Feasibility - Intervention attendance rate: All intervention group families completed therapy (average attendance 91%).  - Drop-out rate: none from intervention group, and when the waitlist group were offered therapy, one child declined, one child as reported by their parent to longer be anxious, and one family was unable to participate. - Treatment fidelity: Child group leaders and senior observers rated adherence to the content of the 'Exploring Feelings' manual as high (97% and 93%, respectively).  Acceptability - Satisfaction with care: qualitative findings showed that all participants would recommend the group to others. Most considered the age range appropriate (i.e., 9-14), but having access to the group during secondary school transfer would be helpful. Participants found sharing experiences as helpful, felt accepted and valued alternative coping strategies to help manage anxiety. One child who previously had individual CBT felt frustrated that he had not learned new information. All parents except for one parent who had previously attend individual CBT reported that the group had a positive impact on their child's anxiety. | Primary outcomes Mental health outcomes using CGI-I, ADIS, SCAS-C, SCAS- P and summary question:  - Both children and parents in immediate therapy were more likely to report reduction in child anxiety than those in delayed therapy (child χ2(1) = 7.43, p = .006, parent χ2(1) = 4.01, p = .045). - No significant differences between immediate therapy and delayed therapy on CGI-I, χ(1) = 1.48, p = .224. 29.4% (n = 5) in immediate therapy were rated 'much' or 'very much improved' compared to 7.1% (n = 1) in delayed therapy. Three children were rated as having 'minimally worse' anxiety (immediate n = 1, delayed n = 1).  - 76% (n = 13) children in immediate therapy had some reduction in the severity of their primary anxiety disorder compared to 33% (n = 5) in delayed therapy, χ2(1) = 6.03, p = .014. - Significant differences in the rate of change on the SCAS-P between immediate therapy (rate of change -1.40 95% CI -2.24, -0.07) and delayed therapy (rate of change - 1.16 95% CI -2.24, -0.07).  - No significant differences in the rate of change on the SCAS-C between immediate therapy (rate of change -1.48 95% CI -2.88, -0.08) and delayed therapy (rate of change -1.30 95% CI -2.69, 0.10). |
| Reaven et al. (2009) | Bespoke group CBT (FYF programme) for anxiety vs Waitlist | Primary outcomes and secondary outcomes not specified.  SCARED - parent and child rated measure of anxiety and mood | Not applicable. | Feasibility  - Attendance rate: The attendance rate at sessions for those who completed treatment was 96%. 92% of families attended 90% or more of the 12-session treatment.  - Dropout rate: Low attrition rate (6%), two families did not complete treatment and did not attend the last two sessions dur to busy schedules and family crises. | Mental health outcomes using the SCARED: - A 2 (Group: Active Treatment vs. Wait List) ×2 (Time: Pretreatment vs. Posttreatment) repeated measures ANOVA of parent report of symptoms on the SCARED revealed a significant effect for Time, F (1, 30) =24.20, p=0.01; and a significant Group×Time interaction effect, F (1,30) =19.52, p=0.01. Thus, according to the parent report, children in the active treatment group experienced a significant decrease in severity of anxiety symptoms over time, whereas children in the Waitlist group did not. - In contrast to the parents’ ratings, when looking at child report on the SCARED, a 2 (Group: Active Treatment vs. Wait List) ×2 (Time: Pretreatment vs. Posttreatment) repeated measures ANOVA revealed no significant effects for Time, F(1, 27) =.67, p=0.43; no effects for Group F(1, 27) =2.63, p =0.12; and no interaction effects: F(1, 27) =.02, p =0.99. -There were no differences in clinical classification (% of children training scores within the clinically significant range by domain on the SCARED, with or without an anxiety disorder) between pre-treatment assessments and Wait List control assessments by either parent or child report. |
| J. Reaven, Blakeley-Smith, Culhane-Shelburne et al. (2012) | Bespoke group CBT for anxiety (FYF programme) vs TAU | Primary and secondary outcomes not reported.  SCARED - self- and parent-report questionnaire to determine symptoms of separation anxiety and social/or generalised anxiety. ADIS-P - clinician interviews, to assesses the presence of anxiety disorders and to determine CSRs. CGI-I - clinician-rated improvement from ADIS-P and SCARED data.  Satisfaction questionnaire. | Not applicable. | Feasibility  - Intervention attendance rate: 21 participants completed the intervention. 87.5% completion rate. Approximately 85% of children missed one session or less. - Intervention drop-out rate: 3 participants dropped-out of FYF. - Adherence to FYF intervention: Of the 144 intervention sessions conducted (12 groups, 12 sessions each), 42 sessions were coded for fidelity (i.e., 29.2%): 82% met criteria of including 85% or more of the required session elements and 97. 6% met criteria of including 80% or more of required session elements.  Acceptability  - Satisfaction of care: 34 children and 38 parents completed the measure within 2 weeks of finishing the intervention; participants in the TAU condition who completed the FYF intervention are also included. Parents rated 17 different activities and children rated 10 different activities; on average, 72% of respondents found the activities overall to be ‘very helpful’. Only an average of 4.7% parents and 9.9% children found the activities ‘not helpful’ overall | Mental health outcomes using ADIS-P, CGI-I and SCARED:  - In the intention-to-treat analyses, controlling for pre-intervention CSRs, significant differences between groups were found for post-intervention CSRs in all diagnoses: SEP, F(1, 40) = 4.21, p = 0.047; SOC, F(1, 40) = 6.04, p = 0.02; SpP, F(1, 40) = 14.45, p = 0.0001; and GAD, F(1, 40) = 8.11, p = 0.007. CSRs were lower in the FYF group post-intervention. Effect sizes ranged from medium to large.  - For the intention-to-treat sample, when controlling for baseline number of anxiety diagnoses, children in the FYF condition demonstrated a significant reduction in overall number of principal anxiety disorders, F (1, 42) = 5.39, p = 0.03. A significant reduction in diagnostic status did not occur in the TAU condition. There was a statistically significant reduction in GAD diagnoses in the FYF group, X2(1,42) =6.64, p=0.01; effect size = 0.85, but not in other diagnoses, p > 0.05. - For the intention-to-treat sample, 10/20 children (50%) in the FYF group and 2/3 children (8.7%) in the TAU group obtained a CGI-I reflecting a positive treatment response, which is a statistically significant difference by group, X2(1,42) = 9.07, p = 0.003, d=1.03. - All statistically significant group differences were maintained in completers analyses. - 3-month and 6-month follow-up (time effect of FYF group): For those returning the follow-up measures, post-intervention reductions on SCARED appeared to be maintained at both 3 and 6 months. |
| Keefer et al. (2017) - linked to Reaven et al. (2018) | Bespoke group CBT for anxiety (FYF programme) | Primary and secondary outcomes not specified but it is implied that IUS is the primary outcome  IUS - parent- and child-report assessing children’s tendency to experience negative emotions, behaviours, and cognitions to uncertain situations and events.  PSWQ - self-report worry scale in children and adolescents. SCARED-C/P - child- and parent-report screener for symptoms of anxiety | Not applicable. | Feasibility  - Intervention attendance: all participants (CYP-parent dyad) attended at least 11/14 sessions.  - Drop-out rate: 10/38 children and 8/36 parents who completed all pre-intervention study measures did not complete the corresponding post-intervention measures either due to study drop-out or non-response to requests for these measures. Therefore, 28 parent-child pairs provided pre- and post-intervention data.  - Treatment fidelity: Clinician fidelity to the intervention was high, ranging from 74% - 99% across sites when assessed. | Effect of time (no comparison group) Mental health outcomes on IUS, PSWQ and SCARED  - IUS: no significant differences pre- to post-intervention on parent version (mean diff = -2.89; 95% CI -7.22, 1.44); child version indicated a near significant change (mean diff = -5.7, 95% CI -11.9, 0.42). - PSWQ: no significant changes pre- to post-intervention (mean diff= -2.93; 95% CI -6.54, 0.7) - SCARED: significant reduction on parent (mean diff = -3.44; 95% CI -6.26, -0.62) but not child report mean diff= -2.13; 95% CI -6.25, 1.99) comparing pre- to post-intervention. |
| Reaven et al. (2015) | Bespoke group CBT for anxiety (FYF programme) | **Primary outcome measure:**  ADIS-P - clinicians conducted this measure with the parent to assess the presence of anxiety disorders and provide CSRs.  **Secondary outcome measure:**  CGI-I - clinician-rated improvement from ADIS-P and SCARED data.  Treatment fidelity measure - assessing fidelity to treatment.  Assessment of CBT knowledge.  Treatment acceptability - completed by clinicians, CYP and parent/guardian | Not applicable. | Feasibility  - Attendance rate: 94% of CYP completed the intervention, with no CYP missing more than 3 sessions.  - Dropout rate: 1 CYP did not complete the intervention.  - Treatment fidelity: ranged from 87-95%.  - Clinicians' CBT knowledge: significant improvement after training, that was maintained at follow up. Acceptability  - Experience of care: Clinicians, CYP and parent/guardians all endorsed high ratings of acceptability per session, with ratings highest amongst parent/guardians and clinicians facilitating their groups; acceptability ratings dropped slightly for one group intervention; positive ratings also given by CYP and clinicians facilitating their groups. Clinicians reported finding discussion and skills-building activities most helpful; CYP reported finding video-based activities most helpful; parent/guardian reported finding exploration and discussion-related activities most helpful | **Effect of time (no comparison group)**  **Primary outcomes**  Mental health outcomes using ADIS-P: - ADIS: Significant decreases in severity were found for the three primary anxiety diagnoses post-intervention: SOC, t(15) = 2.43, p = 0.03, d = .61; SEP, t(15) = 2.13, p = .05, d = .53; and GAD, t(15) = 3.03, p = 0.008, d = .76. Child participants demonstrated a significant reduction in overall number of anxiety disorders, t (15) = 2.78, p = 0.014, d = .70, post-intervention. In all, 40% of participants lost their principal anxiety diagnosis after the completion of the intervention program.  **Secondary outcomes**  Mental health outcome using CGI-I:  - CGI-I - 8 (53% of sample) had a clinically meaningful outcome post-intervention, 3 (20% of sample) were somewhat improved, and 4 (26.7% of sample) experienced no change. None of the children experienced a worsening of symptoms. |
| Reaven et al. (2018) | Bespoke group CBT for anxiety (FYF programme). 3 groups receiving intervention, each group had therapists trained in a different way: standard manual; workshop only, workshop plus. | Primary outcome measures ADIS-P - clinician interviews, to assesses the presence of anxiety and to determine CSRs. Secondary outcome measures  CGI-S - clinician-rated severity the child’s anxiety.  CGI-I - clinician-rated improvement of the child’s anxiety from ADIS-P data.  Treatment fidelity measure - assessing fidelity to treatment.  Assessment of CBT knowledge. | Not applicable. | Feasibility - Attendance rates: 84/91 completed the intervention.  - Drop-out rate: 7/91 did not complete treatment and additional 4 did not complete follow-up/post data.  - Treatment fidelity: ranged from 90-93%, with clinicians delivering the manualised approach having lower fidelity ratings.  - Clinicians' CBT knowledge: significant improvement after training for the Workshop and Workshop Plus modalities.  - Quality of training delivery was lower for clinicians delivering manualised rather than Workshop or Workshop Plus interventions. | Primary outcomes Mental health outcome using ADIS:  - Controlling for clinician's experience by site, there was a significant effect for Time, F(1, 59.37) = 76.02, p<.0001, ωp2 = .64, but no effect for the Time by Condition interaction or a main effect for Condition for GAD; a significant effect for time, F(1, 47.73) = 21.48, p <.0001, ωp2 = .29, but no Time by Condition interaction or main effect of Condition for SOC; a significant effect for time, F(1, 73.23) = 17.25, p < .0001, ωp2 = .18, but condition was not significant and Time by Condition interaction approached significance, F(2, 72.95) = 3.02, p = .06, ωp2 =.05 for SpP; effect of time approached significance, F(1, 21.95) = 3.57, p = .072, ωp2 = .10, but no other effects were observed for SEP. - 31/79 children (39.2%) no longer met criteria for GAD post-treatment and there were no differences among instructional conditions: χ2(2) = 2.94, p = 0.23. 22/77 children (28.6%) lost the SOC diagnosis post-treatment and there were no differences between instructional conditions, χ2(2) = 3.46, p = 0.18. 27/65 (41.5%) lost the SpP diagnosis following treatment, and there were no differences among conditions, χ2(2) = 5.1, p = 0.78. 12/23 (52.2%) lost the SEP diagnosis post-treatment and there were no differences between conditions, χ2(2) = 0.07, p = 0.97. Secondary outcomes Mental health outcomes using CGI-S-; CGI-I:  - There were no statistically significant differences in number of youths who met criteria for a clinically meaningful outcome across instructional conditions, χ2(2) = 1.63, p = .45: Workshop-Plus – 25.9%; Workshop – 44.4 %; and Manual – 29.6%. The following percentages reflected the number of youths in each instructional condition that “somewhat improved” following treatment: Workshop-plus – 40%; Workshop – 35.7%; and Manual – 37%. The following percentages represent the number of youths whose symptom severity showed no real change following treatment participation: Workshop-Plus – 32%; Workshop – 21.4%; and Manual – 33.3%. None of the children experienced a worsening of anxiety symptoms following treatment. |
| Walsh et al. (2018) - linked to Reaven et al. (2018) | Bespoke group CBT for anxiety (FYF programme). 3 groups receiving intervention, each group had therapists trained in a different way: standard manual; workshop only, workshop plus. | Primary outcomes and secondary outcome measures not specified. Treatment acceptability - completed by clinicians, CYP and parent/guardian.  SCARED - parent and adolescent rated measure of anxiety and mood. Satisfaction questionnaire. | Not applicable. | Feasibility  - Drop out - 7/91 who began treatment, dropped out of treatment before completion; 4/91 did not provide follow up data; thus, in total 11 were excluded.  - Attendance rate: 80/91 parent/youth dyads were treatment completers.  Acceptability Experience of care: Parent, youth, and clinician acceptability ratings were high across all instructional conditions (Manual, Workshop, Workshop-Plus). There were no differences in parent or youth acceptability ratings across any of the clinician training conditions (p’s range from .40 to .10). Paired sample t tests revealed a significant difference between parent and youth acceptability ratings of the overall treatment, were parents rated treatment acceptability higher than youth. Exposure techniques were rated more favourably than psychoeducation by parents (p=.039) and by youth (p=0.0001). There was a difference between youth clinician acceptability ratings across conditions. Significant differences were found in youth clinician acceptability ratings between the Manual (M=3.84, SD=.33) and Workshop conditions (M=4.25, SD=.36), p=.001, and between the Workshop (M=4.25, SD=.36) and Workshop-Plus (M=3.95, SD=.39) conditions, p=.026. There were significant differences in parent clinician acceptability ratings between the Manual (M=4.04, SD=.39) and Workshop conditions only (M=4.41, SD=.30), p=0.038. Thus, participation in a Workshop appears to be positively related to treatment acceptability, while the addition of bi-weekly feedback and consultation (at least for youth clinicians only), resulted in relatively lowered acceptability ratings. | Not applicable. |
| Pickard et al. (2020) - linked to Reaven et al. (2018) | Bespoke group CBT for anxiety (FYF programme) | Primary outcomes and secondary outcomes not specified.  Survey inquiring about overall intervention use, continued use of intervention activities, adaptations to intervention, barriers to intervention use, factors affecting sustained use, perceptions of intervention. | Not applicable. | Feasibility - Drop-out rate: 3/33 contacted participants did not complete follow-up survey.  - Attendance rate: 30/33 contacted participants completed follow-up surveys (90.91%).  - Qualitative data indicated clinicians find this a feasible intervention to implement and accessible for CYP and parent/guardian. Acceptability  - Sustainability: 23/30 clinicians (76.7%) continued to use the intervention due to effective anxiety reduction, FYF feasibility and acceptability, and fit of FYF with existing service. Sustained use of intervention was similar across training years and intervention modality. Most clinicians highly endorsed delivering the intervention to ages 8-14 years. 66.7% of clinicians reported delivering the programme in an individual format regardless of being trained to use it in a group delivery model. Clinicians used the range of activities that are part of the intervention with Stress-O-Meter, Deep breathing/Relaxation, Practice exposure in-session and Helpful thoughts/Active minds activities used most often. Clinicians who continued to use FYF reported making M 8.30 adaptations (SD=3.84). Adaptations incorporated in order of most frequent to least included tailoring, lengthening, removing, and shortening components. Clinicians reported making additions that would be considered fidelity-consistent, including additions to lengthen or supplement the existing FYF concepts or to enhance the way in which the program was taught. Adaptations were made in order to tailor the intervention to the learning needs of CYP and parent/guardian.  - Barriers to sustained use in order of most frequent to least were intervention no longer relevant to service, service unable to support use, clinician no longer working clinically, unable to obtain funding for intervention. Many clinicians reported that they experience barriers specific to the delivery of FYF in a group format due to insufficient staffing and difficulties with recruiting youth to participate in the group who are of the same age and level of functioning. | Not applicable. |
| Solish et al. (2020) | Bespoke group CBT for anxiety (FYF programme) | Primary outcomes and secondary outcomes not specified.  SCAS - child- and parent/guardian- rated measure of child anxiety  SCARED - child- and parent/guardian rated measure of child anxiety and mood. Group Questionnaires, Parent and Child versions - developed by authors to be used pre- and post-intervention, includes quantitative ratings (e.g., levels of anxiety and its interference), and qualitative responses (e.g., strategies a parent is currently using).  Satisfaction questionnaire parents’ satisfaction with the program was measured following intervention. | Not applicable. | Feasibility - Dropout rate: Of the 117 children enrolled, 12 did not complete group due to disruptive behaviour/ not being able to manage the group context (n= 6), missing > 3 sessions (n= 5), or change in family availability (n=1).  - Attendance rate: 105/117 children enrolled completed the intervention protocol with fewer than 3 missed appointments and were included in the analyses, yielding a retention rate of 89.74%.  - Treatment fidelity: fidelity to intervention delivery > 80%  Acceptability - Satisfaction of care: parent/guardian endorsed high rates of satisfaction (mean score 140.62, SD=15.43, maximum score 162); a general pattern of positive correlations between satisfaction and improvements in anxiety symptoms with moderately strong correlations between parent-reported anxiety and 1) parent perceived improvement of child's anxiety over time, 2) parent rated usefulness of information, 3) parent rated usefulness of exposure activities in session, and 4) parent-rated confidence to support CYP with anxiety (all p < 0.005). | Effect of time (no comparison group)  Mental health outcomes using SCAS; SCARED, group questionnaire:  - For the combined sample across sites, there were significant improvements from pre- to post-treatment on measures of anxiety (all p < 0.05), excluding on child-rated amount of worry and interference/distress.  - Within-site effects: Child- rated anxiety did not change significantly post-intervention in the community (p=0.11), but changes were significant for the tertiary hospital (p < 0.001) - Cross-site comparisons: controlling for age, there were no significant group differences across sites on change scores (calculated for all variables identified as changing significantly over time). |
| White et al. (2013) | Bespoke individual and group CBT for anxiety (The Multimodal Anxiety and Social Skill Intervention -MASSI) vs Waitlist | Primary outcome measure: CASI-Anx - parent-report inventory used to measure anxiety in autistic individuals PARS- clinician-assessed scale assessing the presence of anxiety symptoms in the past week.  Secondary outcome measures: CGI-I - clinician-assessed scale used in this study to assess global functioning. DD-CGAS: clinician-assessed scale of global functioning in children with developmental disabilities. Attendance rate, in-session participation, homework compliance and intervention fidelity recorded by therapists. Intervention satisfaction parent-assessed and adolescent-assessed rating scales. | Not applicable. | Feasibility  - Intervention attendance rate: 168/180 required individual therapy sessions (12 sessions x 15 participants) were attended. 10/15 participants received 13 individual sessions (M attendance 11.20, range 5-13). 7/15 participants attended all seven group sessions, 4/15 missed 1 session and 5/15 missed more than one session. - Homework compliance: M compliance was 58%, the module on functional assessment had the lowest rate of completion, as it required the greatest time investment by parents. M therapist rated in session client participation was 3.51 (SD = 0.66) (range 1 uninvolved to 4 actively involved). - Intervention drop-out rate: 2 in MASSI group due to decreases in teasing by his peers and decreased social anxiety after changing schools and worsening self-harm, recurring suicidal ideation and suicide attempts requiring multiple emergency room visits; and 3 in waitlist. - Intervention fidelity: Ranged for each module from 87.5-100%. 3/15 participants in MASSI accounted for 65% of the modules with less than 100% fidelity. All group sessions had 100% fidelity. Acceptability - Satisfaction with care: M parent satisfaction was 8.21 (SD = 2.49), scores ranged from 2 to 10 (1 not very helpful, 10 very helpful). M adolescent satisfaction was 7.47 (SD = 3.11), range 1-10. Individual therapy was rated as the most helpful by parents (M = 4, SD = 1.52) (from 1 least helpful to 5 most helpful). Group therapy (M = 3.50, SD = 1.40) was rated as the most helpful by adolescents followed closely by individual therapy (M = 3.36, SD = 1.34). | Primary outcomes Mental health outcomes using CASI-Anx, PARS:  - No significant differences between MASSI and waitlist on the CASI-Anx (t = 1.186, p = .22) and PARS (t = 0.997, p = .31) at post-treatment. 4/15 participants in the MASSI group showed clinically significant and reliable change (RCI > 1.96) on the CASI-Anx, compared to 2/15 in the waitlist.  - No significant change on the CASI-Anx (Z = -1.71, within-group d = 0.55) or the PARS from pre to post (Z = -.71, within-group d = 0.19). Secondary outcomes Social outcomes using DD-CGAS and CGI-I: - Significant differences between the MASSI group and waitlist (t = 2.280, p = .029) at post. Participants in the MASSI group had an average of 6-point increase (i.e., improvement), whereas participants in waitlist had an average increase of one-half point. - Significant pre-post improvement in the MASSI group (Z = -2.74, p < .01, d = 0.81).  - CGI-I scores indicated that 6/15 (40%) participants in the MASSI group were rated as responders, compared to 3/15 (20%) participants in the waitlist. |
| White et al. (2015) - linked to White et al. (2013) | Bespoke individual and group CBT for anxiety (The Multimodal Anxiety and Social Skill Intervention -MASSI) vs Waitlist | Primary and secondary outcomes not specified. CASI-Anx - parent-report inventory used to measure anxiety in autistic individuals Predictors: STAI-T assessing trait anxiety in parents, VIQ assessing verbal ability and SRS assessing severity of autism | Not applicable. | Feasibility - Intervention drop-out rate: 2/15 participants allocated to the MASSI group did not complete the full treatment (i.e., less than 12 individual sessions) due to changed schools and decreased anxiety or worsening self-harm and needing hospitalization. | Effect of time (no comparison group)  Mental health outcomes using CASI-Anx: - Significant decrease in CASI-Anx scores at post compared to pre, t(24) = 3.16, p = .004, at 3-month follow-up, t(18) = 5.15, p < .001 and at 1-year follow-up, t(14) = 2.77, p = .015. - Final model including severity of autism (SRS), parental anxiety (STAI-T) and verbal ability (VIQ) also showed significant decrease in CASI-Anx scores over the course of treatment (time 1 slope b = -6.90, p = .002), but a significant increase over 1-year follow-up (time 2 slope b = 0.74, p < .001). Anxiety did not return to the original level at pre-treatment, 1 year after completing the treatment χ2(1) = 10.25, p = .002. |
| Murphy et al. (2017) | Bespoke individual and group CBT for anxiety (The Multimodal Anxiety and Social Skill Intervention -MASSI) vs person-centred, non-directive counselling | Primary and secondary outcomes not specified. ADIS-C/P: clinician-administered interview to assess anxiety. CSRs ranging from 0 to 8 for diagnosis of anxiety. CASI-anx: parent-report inventory of anxiety in autistic individuals. Fidelity measures:  PCTPRS- to measure adherence to therapy. TPOCS-A - to measure therapeutic alliance via video-assessment of early sessions by blinded, independent raters. | Not applicable. | Feasibility - Intervention attendance rate: Significant difference in mean number of individual sessions attended between MASSI and Counselling (Mann-Whitney U = 32.5, p = .02 (MASSI M = 9.06, SD = 2.51; Counselling M = 11.71, SD = 1.06), with attendance at counselling sessions being significantly higher. - Intervention drop-out rate: 0% in both MASSI and Counselling. - Intervention adherence: Significant difference in the use of CBT techniques between MASSI (M = 25.88, SD = 11.98) and Counselling (M = 9.62, SD = 8.50), Mann-Whitney U = 25.5, p < .001. Significant difference in the use of counselling between MASSI (M = 15.76, SD = 6.32) and Counselling (M = 20.81, SD = 5.14), Mann-Whitney U = 63.5, p < .001, suggesting that the therapists were faithful to both interventions. Acceptability Therapeutic alliance: M scores for the five therapists ranged from 29.95 (SD = 5.42) to 39 (SD = 1.41). There were no significant differences between therapists (Kruskal-Wallis, H (4) = 7.25, p = .07). No significant difference in the mean alliance score for the coded CBT sessions (M = 31.17, SD = 7.12) and for the counselling sessions (M = 31.81, SD = 6.56), Mann-Whitney U = 120, p = .57. | Mental health outcomes using CASI-anx, ADIS-C/P: - No significant differences between MASSI and counselling on CASI-anx at post, F (1,35) = 1.88, p = .18, r = 0.25, and at 12-week follow-up, F (1, 35) = 0.95, p = .33, r = 0.18. - Significantly fewer participants in the counselling group met diagnosis of separation anxiety (ADIS-C/P) compared to the CBT group at post, p = .04, but not at 12-week follow-up, p = .11. No significant differences between groups in the number of participants that met diagnosis of social anxiety, specific phobia and generalised anxiety at both post and 12-week follow-up p > .05. - Significant differences between CBT and counselling in the severity of separation anxiety (CSRs) at post, F (1, 35) = 7.77, p = .01, r = 0.43, but not at 12-week follow-up F (1, 35) = 2.40, p = .13, r = 0.13. No significant differences between groups in the severity of social anxiety, specific phobia and generalised anxiety at post and at 12-week follow-up (all small effect sizes). |
| Maskey, McConachie, et al. (2019) | Bespoke individual CBT with virtual reality for fears and phobias (Blue Room using flat screen computer delivery of images) | Primary and secondary outcomes not specified. SCAS-P/C - child- and parent-report, to assess anxiety symptoms in children. Child confidence ratings - to assess child-rated confidence in tackling behavioural goal | Not applicable. | Feasibility - Intervention attendance rate: All children completed 4 sessions of 20 minutes of graded exposure. | Effect of time (no comparison group).  Mental health outcomes using confidence ratings, SCAS-P and SCAS-C: - Confidence: Seven (87.5%) children showed improved confidence from session 1 to session 4. One child stayed at the same level of confidence throughout the treatment, although this child was classified as a responder to the treatment. - Children classed as responders (n = 4) showed a decrease in SCAS-C scores at 6 months follow-up, which was maintained for two children (28.6%) at 12-month follow-up. None of the children classified as non-responders showed a reduction in SCAS-C scores. |
| Maskey, Rodgers et al. (2019) | Bespoke individual CBT with virtual reality for fears and phobias (Blue Room with VR) (immediately) vs delayed therapy | Primary and secondary outcomes not specified. SCAS-P/C - child- and parent-report measure assessing anxiety symptoms in children Fear survey schedule for children—revised FSSC-R - parent-report questionnaire with an overall intensity and fearfulness score CAPE - child-report of activities to measure any increase in participation in community activities. Confidence ratings - children rated their confidence at tackling their goal situation and parents rated their perception of their child's confidence | Not applicable. | Feasibility -Intervention attendance rate: All children in the immediate group completed all 4 sessions. 1 child did not receive treatment in the delayed group. - Intervention drop-out rate: No dropouts by 6-months. - Treatment fidelity: The average fidelity of treatment delivery across sessions was very high at 94.5% (range 91–96.5%). The overall mean fidelity for the eleven therapists ranged from 84.5-100%. | Mental health outcomes using SCAS-P, SCAS-C, FFSC-R and confidence ratings: - To compare groups’ mean questionnaire scores over time mixed factorial ANOVA were performed, with the group (control vs treatment) and time (baseline, 2 weeks post-treatment, 6 months post-treatment) entered as independent variables. All main effects of group and interactions between time and group were non-significant for all the questionnaires’ scores. Only 6-month outcome data presented in paper. - For the immediate treatment group, most child and parent confidence ratings for tackling the goal situation increased from session 1 to session 4.  Functional outcomes using CAPE:  - No significant differences between in groups in CAPE scores at 2-week post. No significant differences between groups in CAPE scores (formal activities diversity d = 0.2 95% CI -0.574 to 0.91, formal activities intensity d = 0.2 95% CI -0.585 to 0.899, informal activities diversity d = .4, 95% CI -0.703 to 0.779, informal activities intensity d = -0.3 95% CI -1.101 to 0.392) at 6-month follow-up. |
| Ekman et al. (2015) | Adapted individual CBT for anxiety | Primary and secondary outcome measures not specified.  Level of anxiety - self-report item-level rating on a scale from 0-3 for anxiety.  GAF - clinician-rated scale of global functioning. | Communication accommodations  - Use of simple written material and visual aids | Acceptability  - Satisfaction with care: Most clients reported finding the visualisation (i.e., visualised language on a whiteboard) helpful. Help from visualisation to remember the conversation with the therapist: Yes: 14/18; Not known: 1/18; No: 3/18; Chi square test p = .001. Visualisation useful in homework: Yes: 13/18; Not known: 2/18; No: 3/18; Chi square test p = .005. | Effect of time (no comparison group).  Mental health outcome using Level of anxiety:  - Anxiety related to behaviour excess: a statistically significant reduction (p < .001). In post hoc tests (Bonferroni) this was explained by a significant decrease in anxiety from both pre-measurements to post-treatment (p < .05). Anxiety related to behavioural avoidance: a statistically significant reduction (p < .0001). In post hoc tests (Bonferroni), this was explained by a significant decrease in anxiety from both pre-measurements to post-treatment (p < .005). Anxiety related to cognitive excess: a significant main term (p < .001). In post-hoc tests (Bonferroni), this was explained by a significant decrease in anxiety from both pre-measurements to midpoint (p < .05) and post-treatment (p < .05). Anxiety related to cognitive avoidance: no significant differences.  Social outcomes using GAF:  - Significant improvement in clients’ psychological, social and occupational ability to function from pre-treatment (M = 55.72, SE = 2.19) to post-treatment (M= 73.17, SE = 2.71), p < .0001. |
| Storch et al. (2013) | Adapted individual CBT for anxiety (Behavioural Interventions for Anxiety in Children with Autism - BIACA) vs TAU | Primary outcome measures  PARS - clinician-rated measure of child anxiety symptoms, presence and severity.  ADIS-C/P - clinician-rated measure assessing parent and child endorsement of disorder.  CGI-Severity and CGI-Improvement - clinician-ratings of anxiety severity and treatment-related improvement of anxiety.  Secondary outcome measures MASC-P - parent-rated measure assesses parental perception of symptoms in general, social and separation anxiety in children.  RCMAS - child-reported measure assessing anxiety symptoms. CIS-P - parent-rated measure assesses levels of impairment in children e.g., interpersonal, social and academic. | Accommodate individual differences - Tailor practice to individual needs and preferences     Intervention content adaptations - Consider the role of autism Involving wider support network - Parental involvement | Feasibility - Intervention attendance rate: 22 attended all sessions for CBT - Intervention dropout rate: 2/24 dropped out prematurely from CBT arm, none from TAU arm. 3 lost to follow up at 3 months for CBT, thus only 15 participants were assessed at 3-month follow-up. Intention-to-treat analysis included all 24 participants initially allocated to CBT group and all 21 allocated to TAU. | Primary outcomes Mental health outcomes using PARS, ADIS -C/P, CGI: - PARS: 29% reduction in anxiety symptoms at post-treatment in CBT group [baseline M 16.33 (SD 1.93); post-treatment M 11.58 (SD 3.15)] vs. 9% reduction in TAU [baseline M 17.62 (SD 2.04); post-treatment M 16.05 (SD 3.22)]. Treatment effect of d=1.03, p<0.01.  - ADIS: large treatment effects in favour of CBT over TAU at post-treatment (d=0.84, p<0.01). CBT: baseline M 5.42 (SD 0.72), post-treatment M 3.38 (SD 1.81); TAU: baseline M 5.62 (SD 0.92), post-treatment M 4.90 (SD 1.51). 38% of children in CBT group achieved clinical remission (DIS-IV CSR of 3 or less for the primary anxiety diagnosis) at post-treatment vs. 5% TAU (p=0.01, d=1.37). - CGI: Large treatment effects in favour of CBT over TAU for post-treatment CGI-Severity (d=1.06, p<0.01). CBT: baseline M 3.50 (SD 0.72), post-treatment M 2.67 (SD 0.48); TAU: baseline M 4.00 (SD 0.63), post-treatment M 3.57 (SD 0.87). More children in CBT arm were treatment responders relative to TAU [75% (18/24) vs 14% (3/21), p<0.01, d=1.59].  - 3-month follow-up (effect of time): no significant changes from post-treatment were observed among CBT responders on the PARS, CGI-Severity or ADIS at 3-month follow-up (p>0.05).  Secondary outcomes Mental health outcomes using MASC-P; RCMAS: - MASC-P: no significant group differences (p>0.05) - RCMAS: only the anxious arousal subscale showed a difference between groups at post-treatment [CBT: baseline M 2.50 (SD 1.69), post-treatment M 2.29 (SD 1.43); TAU: baseline M 3.24 (SD 1.87), post-treatment M 3.48 (1.63); p<0.05]. Dysphoric mood, oversensitivity and worry showed no significant group differences post-treatment (p>0.05).  - 3-month follow-up (effect of time): Significant changes from post-treatment to 3-month follow-up were observed among CBT responders on dysphoric mood and oversensitivity RCMAS scales (p<0.05). No significant changes from post-treatment were observed among CBT responders on the MASC-P and worry and anxious arousal RCMAS scales at 3-month follow-up (p>0.05). Social outcomes using CIS-P: - CBT arm showed greater improvements relative to TAU on CIS-P (d=0.73; p<.01). CBT: baseline M 21.13 (SD 9.51), post-treatment M 15.54 (SD 6.88); TAU: baseline M 24.71 (SD 10.35), post-treatment M 23.90 (SD 10.25).  - 3-month follow-up (effect of time): No significant changes from post-treatment were observed among CBT responders on the CIS-P at 3-month follow-up (p>0.05). |
| Ehrenreich-May et al. (2014) | Adapted individual CBT for anxiety (developmentally modified version of Behavioural Interventions for Anxiety in Children with Autism - BIACA) | Primary outcome measures PARS - clinician-administered checklist of anxiety symptoms in children and adolescents. ADIS-C/P - clinician-rated measure assessing all DSM-IV anxiety disorders in children and adolescents. CGI-S - clinician-rated measure of overall diagnostic severity. CGI-I - clinician-rated measure of overall diagnostic improvement. Secondary outcome measures CBCL- Anxious/Depressed - parent report measuring anxiety symptoms. RCADS - self-report measure of child/adolescent anxiety and depressive symptoms. | Communication accommodations - Use of clear, simple and preferred language | Feasibility - Intervention attendance: 17/20 participants were treatment completers.  - Intervention drop-out rate: 3/20 participants dropped out of the study prior to the post-treatment assessment and were lost to further follow-up (15% attrition). - Integrity and calibre of treatment: 20 sessions (10 sessions from each treatment site) were randomly selected for adherence rating. Agreement between raters was excellent (k = 0.89), suggesting a high degree of treatment adherence among therapists at both treatment sites. | Effect of time (no comparison group) Primary outcome Mental health outcome using PARS; ADIS; CGI-S; CGI-I: - Mean PARS total scores at post-treatment (M = 10.94, SD = 3.33) were significantly lower compared with baseline (M = 16.50, SD = 3.00), p < .001, d = 1.76. There was no significant difference between post-treatment and 1-month follow-up PARS scores, indicating that treatment gains were maintained. - A significant reduction in ADIS principal CSR values from baseline (M = 5.28, SD = 0.92) to post-treatment (M = 4.35, SD = 1.58), p < .001, d = 0.73 (medium effect). However, among treatment completers (n = 17), only three participants (17.65%) achieved remission of their principal anxiety disorder (i.e., were assigned CSRs ≤3). There was no significant change in principal CSR scores from post-treatment to 1-month follow-up, indicating that treatment gains were maintained. At 1-month follow-up, four subjects (23.53%) no longer met criteria for their principal anxiety disorder. - CGI-S scores were significantly lower at post-treatment (M = 3.06, SD = 1.20), as compared with baseline (M = 4.29, SD = 0.99), p < .001, d = 1.12. There was no significant difference between CGI-S scores at post-treatment and 1-month follow-up, indicating that treatment gains were maintained. - Among treatment completers (n = 17), while only 3 participants (16.67%) had achieved responder status by mid-treatment as indicated by ratings on the CGI-I, 13 participants (76.47%) achieved responder status at post-treatment. Among those completing 1-month follow-up (n = 11), 8 participants (72.73%) were classified as treatment responders. Secondary outcome Mental health outcome using CBCL- Anxious/Depressed; RCADS: - Significant decreases from baseline to post-treatment were observed in CBCL Anxiety/Depression scores (pre-treatment M 10.13, SD 3.67; post-treatment M 7.5, SD 3.43, p < .001, d = 0.74) and treatment gains were maintained at 1-month follow-up (M 5.56, SD 2.70). - There were no significant changes in adolescent-rated RCADS Total scores. |
| Wood et al. (2015) | Adapted individual CBT for anxiety (developmentally modified version of Behavioural Interventions for Anxiety in Children with Autism-BIACA) vs Waitlist | Primary outcome measure  PARS - clinician-rated measure of child anxiety symptoms, presence and severity.  Secondary outcome measure ADIS-C/P - clinician-rated measure assessing all DSM-IV anxiety disorders in children and adolescents.  CGI-Improvement - clinician-rated measure of overall diagnostic improvement. MASC-P - parent-rated measure assesses parental perception of symptoms in general, social and separation anxiety in children.  RCADS - child-reported measure assessing anxiety and depressive symptoms. MASS - child-report measure assessing treatment satisfaction and perceived improvement in therapy.  SACA - standardised interview assesses mental health services obtained by child. | Accommodate individual differences - Tailor practice to individual needs and preferences     Intervention content adaptations - Use of a rewards system Involving wider support network - Parental involvement | Feasibility - Intervention dropout rate: low dropout rate, 1/19 dropouts, 2/19 protocol violation during intervention (beginning excluded treatments or changing medication dose during CBT). Intent-to-treat analysis included 19 participants originally randomized to intervention group. Acceptability - Satisfaction with care: Both parents (M = 2.49, SD = 0.57) and youths (M = 2.21, SD = 0.46) in CBT reported satisfaction with treatment via the MASS at the post-treatment assessment in the CBT group. | Primary outcome Mental health outcomes using PARS:  - A significant difference was observed at post-treatment/post-waitlist for the PARS (p = .04, ES = 0.74), with the CBT group showing lower anxiety scores after controlling for baseline relative to the waiting list (WL) group. CBT group: baseline M 17.16 (SD 2.69), post-treatment M 11.62 (SD 3.26); WL group baseline M 18.29 (SD 2.09), post-treatment M 14.04 (SD 2.64).  Secondary outcomes Mental health outcomes using ADIS, CGI, MASC-P, RCADS: - Regarding CGI-Improvement criteria for positive treatment response, 79% (15/19) of those in the CBT condition experienced treatment response compared to 28.6% (4/14) of those in the WL at post-waitlist (p < 0.01, odds ratio = 9.38).  - ADIS: When considering positive diagnostic status at post-treatment (defined as a child still meeting criteria for their primary anxiety disorder diagnosis with an ADIS-C/PCSR score ≥ 4), in the CBT condition, 6/19 (32%) participants did not meet criteria for their principal anxiety disorder diagnosis at post-treatment, in comparison to 3/14 (21%) of participants in the WL condition. However, this difference was not found to be statistically significant (p = 0.70, odds ratio = 1.69), nor were group differences found for ADIS CSR changes for principal anxiety diagnoses [CBT group: baseline M 5.68 (SD 0.89), post-treatment M 3.28 (SD 2.10); WL group: baseline M 5.50 (SD 0.65), post-treatment M 4.04 (SD 1.88), p = 0.25, ES = 0.39].  - Group differences for parent-reported anxiety (MASC-P) were only marginally statistically significant (p = 0.10, ES = 0.59). CBT group: baseline M 60.58 (SD 16.01), post-treatment M 50.21 (SD 12.71); WL group baseline M 62.49 (SD 14.48), post-treatment M 58.44 (SD 9.01).  - Group differences for youth-reported anxiety (RCADS) were nonsignificant (p = .93, ES = 0.02). CBT group: baseline M 28.09 (SD 20.27), post-treatment M 19.58 (SD 14.65); WL group baseline M 24.65 (SD 17.31), post-treatment M 18.31 (SD 14.09).  - 1-month follow-up (effect of time): 13 adolescents from the CBT group completed 1-month follow-ups. At this time point, there was no significant change in clinician-rated anxiety between the post and 1-month assessment findings on the PARS (p= 0.79, ES = 0.07), and 10/13 participants-maintained treatment responder status (a statistically nonsignificant decline; p=0.22). Additionally, 8/13 (62%) children showed remission of their principal diagnosis at follow-up (a significant improvement from posttreatment; p= 0.02). In this group of 13 youth, there was a significant reduction in youth-reported anxiety from post-treatment to 1-month follow-up (using the RCADS; p= 0.02, ES = 0.95), with a marginal effect for parents (using the MASC-P; p=0.07, ES=.74). Service use using SACA (treatment completers only):  - Group differences were not detected in the rate of service use (p = 0.06). All youth randomized to the waitlist condition received some intervention services intended to target anxiety and/or autism symptoms (e.g., social skill deficits) including social skills training (n = 2; 18.2%), psychiatric medication management (n = 7; 63.6%), or special education services (n = 6; 54.5%). Seven waitlist participants (63.6%) received one service and 4 participants (36.4%) received two services. Most youth randomized to CBT (n = 11; 68.8%) received services during the acute phase, including medication management visits (n = 7; 43.8%), counselling or therapy in school (n = 3; 18.8%), special education services (n = 5; 31.3%), religious counselling (n = 1; 6.3%), case management (n = 1; 6.3%), individual supportive psychotherapy (n = 2; 12.5%), or was seen in the emergency room for emotional or behavioural problems (n = 1; 6.3%). Regarding the number of services, 5 (31.3%) children received no services, 5 (31.3%) children received one service, and 6 (37.5%) youth received two or more services. Except for the emergency room visit, these services had been initiated prior to the onset of the study. |
| Storch et al. (2015) | Adapted individual CBT for anxiety (developmentally modified version of Behavioural Interventions for Anxiety in Children with Autism - BIACA) vs TAU | Primary and secondary outcome measures not specified.  PARS- clinician-rated measure of child anxiety symptoms presence and severity.  ADIS-C/P - clinician-rated measure assessing parent and child endorsement of disorder.  CGI-Severity and CGI-Improvement - clinician-ratings of anxiety severity and treatment-related improvement of anxiety.  MASC-P - parent-rated measure assesses parental perception of symptoms in general, social and separation anxiety in children.  RCADS - child-reported measure assessing anxiety and depressive symptoms.  CIS-P - parent-rated measure assesses levels of impairment in children e.g., interpersonal, social and academic. | Accommodate individual differences - Tailor practice to individual needs and preferences     Intervention content adaptations - Use of a rewards system Involving wider support network - Parental involvement | Feasibility - Intervention attendance rate: All (n=16) attended all CBT sessions. - Intervention dropout rate: No drop out from CBT or TAU at post-treatment. Only 9 participants from CBT group were assessed at 1-month follow-up, 2 lost to follow up. | Mental health outcomes using PARS, ADIS -C/P, CGI, MASC-P; RCADS: - PARS: significant group differences at post-treatment were observed (d=0.79, p <0.05). CBT: baseline M 15.81 (SD 1.64), post-treatment M 10.94 (SD 3.91); TAU baseline M 15.67 (SD 2.50), post-treatment M.13.93 (SD 3.56). - ADIS: significant group differences at post-treatment were observed (d=1.30, p <0.01). CBT: baseline M 5.50 (SD 1.03), post-treatment M 3.69 (SD 1.35); TAU baseline M 5.73 (SD 0.80), post-treatment M 5.33 (SD 0.90). 37.5% of those in the CBT condition no longer met diagnostic criteria for their primary diagnosis at post-treatment, whereas no participants in the TAU condition met criteria for diagnostic remission (p=0.02). - CGI: significant group differences at post-treatment were observed in CGI-Severity scores (d=0.94, p <0.05). CBT: baseline M 3.69 (SD 0.60), post-treatment M 2.88 (SD 0.72); TAU baseline M 3.93 (SD 0.80), post-treatment M 3.67 (SD 0.82). 68.8% of those in the CBT group were treatment responders, compared to 26.7% of those in the TAU group (p=0.03). - MASC-P & RCADS: no significant differences were detected (p>0.05). - 3-month follow-up (effect of time): No significant changes from post-treatment were observed among CBT responders on the PARS, ADIS -C/P, CGI, MASC-P; RCADS at 3-month follow-up (p>0.05). Social outcomes using CIS-P: - Significant improvements in the CBT versus TAU arm were observed regarding overall functional impairment (d=0.59, p<0.05). CBT: baseline M 23.52 (SD 7.51), post-treatment M 17.24 (SD 9.75); TAU baseline M 25.67 (SD 7.45), post-treatment M 24.96 (SD 8.91). - 3-month follow-up (effect of time): No significant changes from post-treatment were observed among CBT responders on the CIS-P at 3-month follow-up (p>0.05). |
| Oerbeck et al. (2021) | Adapted individual CBT for anxiety ('Less Stress' program) | Primary and secondary outcome measures not specified.  K-SADS-PL - mother-reported measure assessing child comorbidity and school refusal. RCADS: child anxiety symptoms rated by parents and children.  CGAS: clinician-rated tool to indicate lowest overall level of the child’s psychosocial functioning (at home, at school, with peers). ILC: child-rated measure of quality of life. | Communication accommodations - Use of simple written material and visual aids - Use of technology Intervention content adaptations - Use of role play or modelling | Feasibility - Homework compliance: When the therapists rated to what extent the child had carried out the home assignments, the mean scores were considered satisfactory. The mean rating was 3.5 (SD =1.6) (6 indicates to a very large extent) and increased in the second half of the program (mean > 4) compared to the first half (mean > 3). The mean among the eight completers was higher (4.2, SD 0.5) than the general mean and very low in the two non-completers (0.8, SD 1.2). - Intervention adherence and therapist competence: The therapist-rated adherence to “Less stress” (e.g., adherence to specified session structure, therapeutic process and relationship skills, focus on the goals set for the session) and their perceived competence was considered satisfactory. The therapists found the manual easy to use but adaptations were considered necessary in some instances. - Intervention attendance rate: 8/10 (80%) participating children completed the “Less stress” program. Baseline values showed better functioning on important measures for the non-completers at baseline. Acceptability - Treatment satisfaction: Both parents and children expressed an overall contentment with “Less stress”. In the open-ended questions, most parents said “Less stress” was hard work but that they had learned methods to continue to use in helping their children to master anxiety. Many children expressed that they liked the program because they had become less anxious, had mastered assignments and got nice rewards. Not much was indicated as disliked, but one child noted there was too much talking, and one was not satisfied with the professor in the PowerPoint presentation. The parents in general put weight on the systematic work overtime, and the concrete goals to work on from session to session. However, one child said he disliked having to do “what he didn’t want to do” and his parents pointed out that although they were given many home assignments, they were not concrete enough and/or they lost control of what to work with at home. | Effect of time (no comparison group) Mental health outcomes using K-SADS; RCADS; CGAS: - K-SADS: After treatment, 7/8 (87.5%) completers benefited from the program. 5/7 (71.43%) children did not meet criteria for any anxiety disorders and two had fewer anxiety disorders. Furthermore, the three diagnosed Depressive episodes and school refusal were no longer present, with no change in the diagnoses of ADHD (7/10 children) and Enuresis nocturna (1/10 children).  - RCADS mother: baseline M 94.6 (SD 11.0), after treatment: M 73.2 (SD 20.1), p< 0.01. RCADS father: baseline M 96.2 (SD 16.3), after treatment: M 75.6 (SD 17.9), p=0.04. RCADS child: baseline M 67.0 (SD 16.0), after treatment M 60.0 (SD 12.7), p=0.01. Thus, significant improvement over time.  Social outcomes using CGAS: - CGAS, clinician-rated: baseline M 42.4 (SD 6.9); after treatment M 48.3 (SD 3.8), p < 0.01. Thus, significant improvement over time. Quality of life using  - ILC child, total score: baseline M 16.6 (SD 4.4), after treatment M 18.9 (SD 3.6), p=0.07. ILC, child, problem score: baseline M 3.6 (SD 1.5), after treatment M 3.0 (SD 1.7), p= 0.16. No significant improvement over time. |
| Wise et al. (2019) | Adapted individual CBT for anxiety (Treatment of Anxiety in Late Adolescents with Autism - TALAA) | Primary and secondary outcome measures not specified.  HAM-A - clinician-rated measure assessing anxiety symptoms.  HAM-D - clinician-rated measure of depressive symptoms.  CGI-S and CGI-I - clinician-rated measures of symptom severity and improvement  BAI - self-report measure assessing the presence and severity of anxiety. | Communication accommodations  - Use of simple, written material and visual aids  Accommodate individual differences  - Encourage individual's hobbies and interests  Intervention content adaptations - Use of a rewards system - Integration of emotion-focused strategies  - Integration of social skills training  Involving wider support network - Parental involvement | Feasibility  - Intervention drop-out rate: One participant dropped out at the mid-assessment point. | Effect of time (no comparison group).  Mental health outcomes using HAM-A; HAM-D; CGI-S; CGI-I; & BAI:  - CGI-S scores were significantly lower from baseline (M = 4.86, SD = 0.69) to post-treatment (M = 2.71, SD = 1.11), p < .001.  - Among participants (n = 7), including the one participant only completing treatment until the mid-assessment, four of the participants were classified as treatment responders (57%) based on report on the CGI-I at post-assessment.  - Results indicated a significant reduction in clinician-rated symptoms of anxiety on the HAM-A from baseline (M = 22.14, SD = 3.02) to post-treatment (M = 14.57, SD = 5.06), p < .006, (large effect).  - Clinician-rated symptoms of depression (HAM-D) did not reveal clinically significant reductions from pre-treatment (M = 6.71, SD = 5.61) to post-treatment (M = 5.71, SD = 3.40), p = .60.  - Analysis of participant reports of anxiety (BAI) did not reveal significant differences from pre-treatment (M = 12.33, SD = 7.29) to post-treatment (M = 9.33, SD = 5.85), p = .28. |
| Fujii et al. (2013) | Adapted individual CBT for anxiety (Behavioural Interventions for Anxiety in Children with Autism - BIACA) vs TAU | Primary and secondary outcome measures not specified.  ADIS-C/P - clinician-administered semi-structured diagnostic interview that generates diagnoses as well as CSR scores. | Structural or procedural adaptations - Format of the intervention Intervention content adaptations - Integration of social skills training Involving the wider support network - Involving school | Feasibility - Intervention drop out: Sixteen participants completed the intake assessment and were randomized for inclusion the study. Four of those participants (three CBT and one TAU) did not complete treatment or their post-assessments due to the family’s inability to consistently attend treatment sessions. - Service use during treatment: Children within the CBT condition did not receive any anxiety-specific services outside of our CBT program. However, two out of the seven children in CBT were on stable dosages of a psychotropic medication; four received speech therapy; and six had received social skills training in the year prior to enrolment in the current study. | Mental health outcome using ADIS-C/P: - Five of seven children (71.4 %) in CBT no longer met diagnostic criteria for their primary anxiety disorder at post treatment (32 weeks), whereas all five children in the TAU condition still met diagnostic criteria for their primary anxiety disorder (CSR => 4) at 16 weeks. (x2 = 6.12, p = .013).  - Mean (SD) highest anxiety disorder CSR scores at baseline for the CBT and TAU conditions were 5.57 (0.54) and 5.60 (0.55), respectively. Post CBT/TAU scores where highest anxiety disorder CSR ratings were 3.86 (0.90) and 5.60 (0.55). The CSR ratings significantly differed by treatment group at post-IT/post-TAU, F (2,12) = 6.62, p=.017. On average, children assigned to CBT had lower anxiety severity scores after 32 weeks of CBT when compared to children assigned to TAU for 16 weeks. |
| A. J. Russell et al. (2013) | Adapted individual CBT for OCD vs adapted Anxiety Management | Primary outcome  YBOCS - clinician-rated measure assessing severity of OCD symptoms.  Secondary outcomes  CGI-S and CGI-I – clinician-assessed scale measuring symptom severity and improvement.  D-YBOCS - semi structured interview to ascertain the presence and severity of OCD symptoms.  OCI-R – self-report scale measuring obsessive compulsive symptoms.  BDI - self-report measure of depression.  BAI & SCAS - self-report measures of anxiety.  LSAS - self-report assessing anxiety about and avoidance of a range of social situations.  WSAS – self-report scale measuring severity of impairment.  PR-CHOCI-R: parental/carer report assessing obsessive-compulsive symptoms in children.  Treatment satisfaction: a self-rated 8-point visual analogue scale. | Communication accommodations  - Use of clear, simple and preferred language  - Use of simple, written material and visual aids  Structural or procedural adaptations  - A structured and predictable approach  Intervention content adaptations - Integration of emotion-focused strategies  - Integration of cognitive-behavioural approaches | Feasibility  - Intervention drop-out rate: In the CBT arm, 1 participant was lost to follow-up due to major depressive episode and 2 discontinued the intervention due to acute medical admission and withdrawal from treatment, and in the anxiety management arm 3 participant discontinued the intervention due to depression, withdrawn assent and unknown reason.  - Intervention attendance rate: 20/23 people in each arm completed treatment. The mean number of treatment sessions was marginally greater in the CBT (M = 17.43, SD = 4.3) group than in the control group (M = 14.43, SD = 5.3, t = -2.022, p = .03; 95% CI -5.98 to -0.006). The M number of weeks between pre-treatment end-of-treatment ratings were AM group 23.74 (SD = 10.37); CBT for OCD 27.06 (SD = 10.27). The most usual length of treatment in weeks was 25.  Acceptability  - Treatment satisfaction: There were no differences between the two treatment groups in their reports of treatment satisfaction. AM M satisfaction score = 5.60 (SD = 2.131); CBT M satisfaction score = 4.9 (SD = 2.3), p = .425.  - Cross-over cases: Nine (39%) participants in the anxiety management comparison group, compared with three (13%) participants in the CBT group, asked to "crossover" or try the other treatment either at or after the 1-month follow-up point (χ2 = 4.05, p = .044). 8/9 participants originally randomised to anxiety management who "crossed over" to CBT completed the second treatment and attended for symptom ratings | Primary Outcome  Mental health primary outcome using YBOCS:  - There was no significant difference between treatment groups on YBOCS total severity scores (primary outcome measure) at the end of treatment when controlling for pre-treatment YBOCS severity ratings (p = .295). There were significant changes in YBOCS total severity ratings from pre-treatment to end of treatment in both the CBT group (p = .001) and AM group (p < .0001). The YBOCS total severity scores effect size was large and clinically meaningful in the CBT group (d = 1.15) and medium in the AM group (d = 0.6).  - There were more treatment responders (i.e., had a >25% reduction in YBOCS total severity ratings) in the CBT group as compared to the AM group (9/20 [45%] versus 5/20 [20%], respectively). However, this difference in response rate was not statistically significant (χ2 = 1.72, p = .160). When a more stringent rating of treatment response (a CGI “much or very much improved” combined with a >35% reduction in YBOCS total severity ratings) was considered, 6/20 (30%) of the CBT group achieved treatment response compared with 2/20 (10%) of the AM group. The groups did not differ significantly in the proportion of treatment responders. Slightly more participants in the CBT group were classified as remitted cases (i.e., with a YBOCS total severity rating of ≤12 1 week after treatment ended) as compared with the AM group (5/20 [20%] versus 3/20 [15%]) but this difference was not statistically significant.  - Cohen’s d effect sizes were 0.4 for the YBOCS total severity rating, 0.4 for YBOCS obsessions severity, 0.2 for YBOCS compulsions, and 0.3 for CGI, all indicating a small advantage for CBT over the AM group after treatment.  -In the CBT group, changes in YBOCS total severity scores compared to pre-treatment remained significant at 1-month follow-up (p = .005), 3-month follow-up (n = 10, p = .008), 6-month follow-up (n = 12, p = .007), and 12-month follow-up (n = 11; p = .011).  Secondary outcome  Mental health secondary outcomes using CGI-S; CGI-I; D-YBOCS; OCI-R; BDI; BAI; LSAS; SCAS; PR-CHOCI-R:  - Although clinician ratings of CGI-S changed significantly between pre- and post-treatment (p < 0.001), this did not vary by treatment group (p = 0.140).  - The treatment groups differ significantly in terms of the proportion of participants in each group rated as "much" or "very much improved" on the CGI-I scale (CBT n = 11; AM n = 5 (χ2 = 3.886 (df = 1), p = .050).  - D-YBOCS results not reported  - Neither of the groups showed significant differences between pre-, post-, and 1-month follow-up mean scores on any of the self-ratings (OCI-R; BDI; SCAS; BAI; LSAS).  - PR-CHOCI-R: Informant ratings differed significantly between pre- and post-treatment (improvement) only for the anxiety management comparison group (p < .05)  Social outcome using WSAS:  - Neither of the groups showed significant differences in social functioning (WSAS) between pre-, post-, and 1-month follow-up mean scores |
| Storch et al. (2020) | Adapted individual Family-based exposure-focused treatment - FET for anxiety vs TAU | Primary outcome measures  PARS - clinician-rated measure of child anxiety symptoms presence and severity.  ADISC/P - clinician-rated measure assessing parent and child endorsement of disorder.  CGIS/CGII - clinician-reported measure of overall symptoms severity and improvement of anxiety symptomatology. Secondary outcome measures MASC-P - parent-rated measure of child's anxiety. CAISP - parent-rated measure of impact of anxiety symptoms in a child's life. | Communication accommodations - Use of clear, simple and preferred language Intervention content adaptations - Use of a rewards system Involving wider support network - Parental involvement | Feasibility - Intervention attendance rate: 14/14 received FET intervention, 3/14 discontinued the intervention.  - Intervention dropout rate: 3/14 discontinued the intervention, thus 11/14 were assessed at post-treatment. 1/11 lost to 2-month follow-up. 7 assessed at 2-month follow-up. | Primary outcomes Mental health outcomes using PARS; ADIS; CGI: - There were significantly larger reductions in the FET group compared to TAU in the primary anxiety severity measures, with large effects observed for the PARS (FET: baseline M 14.07, SD=2.34; post-treatment M 7.79, SD=5.37. TAU: baseline M 16.22, SD 2.67; post-treatment M 15.06, SD 3.21. d=1.01, p<0.01), ADIS-C/P CSR (FET: baseline M 4.50, SD=0.52; post-treatment M 2.05, SD=1.85. TAU: baseline M 5.33, SD 0.84; post-treatment M 5.00, SD 0.77. d=1.11, p<0.01) and CGI-S (FET: baseline M 3.29, SD=0.73; post-treatment M 2.25, SD=0.87. TAU: baseline M 3.67, SD 0.59; post-treatment M 3.67, SD 0.69. d=1.22, p<0.01). - FET outperformed TAU for CGI-I (78.6% vs. 0% treatment responders), ADIS-CRS (85.7% vs. 0% clinical remission) measures of remission.  - No significant changes in FET group from post-treatment to 2 months follow up. In considering follow up response and remission status, 100% of treatment responders retained treatment responder and symptom remission status on all measures (i.e., the CGI-I, CGI-S, and ADIS-IV-C/P CSR) status at 2-month follow up. Secondary outcomes Mental health outcomes using MASCP: - No significant group effect was found for change in MASC-P anxiety symptoms. Social outcomes using CAISP: - FET associated with significantly larger reductions in CAIS-P total scores compared to TAU (FET: baseline M 33.90, SD=16.89; post-treatment M 16.49, SD=14.95. TAU: baseline M 29.58, SD 15.23; post-treatment M 32.10, SD 16.11. d=1.04, p<0.01).  - No significant changes in FET group from post-treatment to 2 months follow up. |
| Ollendick et al. (2021) | Adapted individual One-Session Treatment - OST for specific phobia | Primary and secondary measures not specified. ADISP/C- clinician-administered measure to assess the presence and severity of specific phobias.  Fear rating for child's primary specific phobia - parent-report. BAT - child-reported, percentage of steps completed was used as a measure of BAT completion. SUDs were also reported before and after completion of each task. Parent and Child Treatment Satisfaction- parent/child self-report of treatment satisfaction. Parents and children also rated fear, avoidance, and coping for phobic objects as compared to peers. Perceived interference, helpfulness of treatment, and willingness to recommend treatment to children with similar problems were also rated. | Communication accommodations - Use of simple written material and visual aids Accommodate individual differences - Encourage individual's hobbies and interests Involving wider support network - Parental involvement | Feasibility - Intervention drop out: After completion of initial assessments, one family opted not to participate in the study and one family was referred out due to the presence of severe comorbid psychopathology resulting in a total of 9 participants in the treatment protocol. 100% of the 9 families completed the treatment and 1 week follow-up, 3/9 families (33.33%) did not complete 6-month follow-up. Two families did not respond to the clinic after three attempts were made to contact them for the 6-month follow-up and one family indicated it was impossible to get their child to leave the house due to (non-phobia related) increased psychopathology. Acceptability -Treatment satisfaction: Parents reported a high level of satisfaction with child's improvement following treatment. Seven of the nine families (78%) indicated that they strongly agreed with the statement that treatment was helpful for their child, while two families (22%) indicated that they agreed with this statement. | Effect of time (no comparison group) Mental health outcomes using ADIS-P/C; BAT; fear rating: - ADIS-P: Significant and a large effect size reduction in symptoms on the ADIS-P overall (χ2(2) =10.38, p=.006, W=0.58). ADIS-P CSR means changed from 5.89 at pre-treatment, to 2.33 at post-treatment, and 2.17 at follow-up. Significant reductions in phobia severity from pre to post (p=.007), but not from post to follow-up (p=.590). Reliable change index scores calculated from Specific Phobia CSR scores on the ADIS-P to measure clinically significant changes: 6/9 (67%) children exceeded the clinical cutoff (>1.96) for change in specific phobia scores from pre to post and 6/9 (67%) were in the non-clinical range at post-assessment (CSR<4). 4/6 (67%) who completed the 6-month follow-up fell in the non-clinical range at post-assessment (CSR<4). For the pre to 6-month follow-up, four (67%) of the six children who completed follow-up assessment exceeded the clinical cutoff (>1.96) for change between their pre and 6-month CSR scores, and four (67%) of the six children who completed the 6-month follow-up fell in the non-clinical range at post-assessment (CSR<4) as well. - ADIS-C: Significant reduction in symptoms on the ADIS-C overall with a large effect size (χ2(2) =9.29, p<.01, W=0.53). ADIS-C CSR means changed from 5.67 at pre, to 2.50 at post-treatment, and 3.17 at follow-up. Significant reductions in phobia severity from pre to post (p=.011), but not from post-treatment to follow-up (p=.180). Reliable change index scores calculated for Specific Phobia CSR scores on the ADIS-C to measure clinically significant changes: 7/9 (78%) exceeded the clinical cutoff (>1.96) for change in specific phobia scores from pre to post and 7/9 (78%) were in the non-clinical range at post-assessment (CSR<4). 3/6 (50%) who completed 6 months follow-up assessment exceeded the clinical cutoff (>1.96) for change between their pre and 6-month CSR scores, with 4/6 (67%) who completed the 6-month follow-up in the non-clinical range at post-assessment (CSR<4). - BAT: At pre (p=.223), post (p=.414), and follow-up (p=.157), there were no significant differences between the percentages of BAT steps completed alone vs. with a parent, although in general more steps were completed in the parent present condition. Changes in the percentage of steps completed alone at the pre (M=.60 [60%], SD=.37), post (M=.74 [74%], SD=.33), and follow-up (M=.85 [85%], SD=.27) assessments. There was no significant reduction in avoidance without a parent present between pre-treatment assessment to follow-up (χ2(2) =.353, p=.838). 5/9 participants (55.55%) completed 70% or more of the steps at post-treatment, and at follow-up. There was no significant reduction in avoidance with a parent present between pre-treatment assessment to follow-up (χ2(2) =2.00, p=.368). With parent present, 7/9 participants (77.77%) completed 70% or more of the steps at post-treatment and 5/6 participants (83.33%) that remained at follow-up were able to complete 70% or more of steps at follow-up. - Significant reduction in parent fear ratings overall (χ2(2) =7.90, p=.019). Parent fear rating means changed from 6.98 at pre, to 3.22 at post-treatment, and 3.33 at follow-up. Significant reductions in parent fear ratings from pre to post (Z=− 2.67, p=.008), but not from post to follow-up (Z=− .14, p=.891). |
| Driscoll et al. (2020) | Adapted individual CBT for anxiety for young children (aged 4-8) ('Being Brave' programme) | Primary outcome measure CGI-A Improvement score - clinician-administered measure of global improvement of anxiety (i.e., treatment response).  Secondary outcome measure KSADS-E - clinician-administered measure to assess anxiety and other psychiatric disorders. PARS - clinician-administered measuring severity of anxiety symptoms. CQ - clinician-administered and parent-rated measure assessing changes in a child’s ability to manage specific anxiety-provoking situations. SPAS for 3- to 5-year-olds or SCAS-Parent version for children over five - parent-reported measure of their child's anxiety symptoms.  CBCL-Anxious Depressed - parent report measuring child anxiety symptoms. KFQ - child-report measure of the intensity of their fear of fear-provoking stimuli illustrated with pictures. FLIS - parent-reported measure of the extent to which child behaviour limited participation in activities typical of families with young children. | Communication accommodations - Use of simple, written material and visual aids    Accommodate individual differences - Tailor practice to individual needs and preferences  Structural or procedural adaptations - Format of intervention Intervention content adaptations - Integration of emotion-focused strategies - Integration of social skills training Involving wider support network  - Parental involvement | Feasibility - Intervention drop-out rate: 2/16 dropped out before intervention was complete (1 after Week 3 because of parental separation; 1 after Week 6 because the child had shown improvement, and the family did not want to keep coming), 1/16 declined post-treatment assessment, and further 4 declined follow-up assessment because of busy schedule, thus 11/16 completed 4-month follow-up. - Intervention attendance rate: Sixteen children enrolled in treatment, and 14/16 children and families completed at least 10 sessions (M 14.1, SD 3.89). One family had difficulty attending given parental separation during the study, and another family withdrew after six sessions because the child had become less anxious and busy schedules made continued treatment a lower priority. | Effect of time (no comparison group) Primary outcome Mental health outcome using CGI-A Improvement: - 81% of children were rated “much improved” or “very much improved” (CGI-I’s 1 or 2) post-treatment. 10 children continued to be rated "much-" or "very much improved" at 4 months follow-up. Secondary outcomes Mental health outcomes using KSADS-E; PARS; CQ; SPAS; SCAS; CBCL- Anxious/Depressed; KFQ: - PARS: Significant improvement in clinician ratings of child anxiety on PARS from baseline M 21.9 (SD 4.2) to post-treatment M 12.9 (SD 3.3) (p = 0.001), as well as from baseline to follow-up M 13.1 (SD 3.4) (p = 0.003). At baseline, 14/16 children (87.5%) scored above the clinical cut-off established on the PARS for typically developing children of 17.5, whereas at post-treatment, 1/15 only (6.7%) were above this cut-off, and at follow-up 1/11 (9.1%). - SPAS/SCAS: Significant improvement in parent ratings of child's anxiety on SPAS from baseline M 42.1 (SD 9.9) to post-treatment M 26.4 (SD 14.9) (p = 0.02), and from SPAS baseline to SPAS follow-up: 22.4 (16.6) (p = 0.04). Significant improvement in parent ratings of child's anxiety on SCAS from baseline M 30.3 (SD 6.7) to post-treatment M 20.5 (SD 11.2) (p = 0.04); but not between baseline to follow-up M 18.3 (SD 6.3) (p = 0.06). On the SPAS/SCAS, 15/16 (93.8%) of children were within the clinical range at baseline, 5/15 (33.3%) at post-treatment, and 2/10 (20%) at follow-up. - Differences on measures of anxiety (PARS, SCAS, SPAS), had effect sizes ranging from medium to very large post-treatment, which were maintained, with similar effect sizes, at follow-up. - Non-significant change for CBCL-Anxious/Depressed scores from baseline M 64.1 (SD 4.8) to post-treatment M 59.2 (SD 6.5), p=0.13; and from baseline to follow-up M 60.2 (SD 5.8), p=0.14. - KSADS-E: Children had a baseline mean of 2.25 (SD 1.0) anxiety disorders, while the post-treatment mean in completers was 0.87 (SD 1.06), representing a significant decrease in the number of anxiety disorders (p = 0.003). The rate of multiple anxiety disorders in completers declined from 80% to 13%. The one disorder that did not decline by a third or more was specific phobia. As a result, the number of children who were completely free of any anxiety disorder at post-treatment was only 40%. The number of anxiety disorders remained low at follow-up (mean = 0.36, SD = 0.92), representing a significant change from baseline (p = 0.0044). 82% of children had lost all anxiety diagnoses at follow-up, accounted for by a reduction in specific phobias. - Significant improvement on CQ scores from baseline M 2.43 (SD=0.82) to post-treatment M 4.99 (SD=0.86) (p = 0.001), and baseline to follow-up M 5.00 (1.18) (p = 0.01). - Non-significant improvement between KFQ scores from baseline M 1.08 (SD 0.37) to post-treatment M 0.95 (SD 0.27) (p = 0.066). KFQ was not administered at follow-up as authors felt children misunderstood the measure. Social outcomes using FLIS: - Families who completed treatment reported significant improvement in global family function from baseline M 0.46 (SD 0.42) to post-treatment M 0.33 (SD 0.36) (p=0.03), and baseline to follow-up M 0.32 (SD 0.28) (p=0.011); and significant reduction in the need to restrict family activities including socializing, traveling, and going new places from baseline M 0.37 (0.45) to post-treatment M 0.23 (0.37) (p=0.049), and baseline to follow-up M 0.20 (SD 0.21) (p=0.028). Global family impairment and restrictions on family activities were further improved at follow-up from baseline, with medium to large effect sizes. |
| **Interventions targeting emotional regulation** | | | | | |
| Factor et al. (2019) | Adapted group CBT for emotional regulation (Stress and Anger Management Program -STAMP) vs waitlist (delayed-treatment control group) | Primary and secondary outcome measures not specified.  ERC - parent-report measure assessing two dimensions of emotional regulation, regulation and liability/negativity. Parent confidence ratings - parent-rated measure of their perceived self-confidence in managing their child's anxiety and anger and their perceived confidence in their child's skills level in managing their own anxiety and anger (only the later reported in this review). | Intervention content adaptations - Simplified content | Feasibility - Intervention dropout rate: 4/27 families not to partake before intervention started, thus analyses is on 23 parent-child dyads. - Intervention attendance rate: All 23 participants completed more than 50% of sessions. | **Effect over time reported, even though there is a comparison group.**  Mental health outcomes using ERC; parent confidence ratings:  - A paired samples t-test for the treatment group revealed a significant difference in the scores for ERC-L/N pretreatment versus posttreatment, p = 0.03, with decreased ERC-L/N scores at posttreatment. For the waitlist group, there was no significant difference in the scores for ERC-L/N for pretreatment/intake versus pretreatment (same timepoint as post treatment for intervention group), p = 0.34. A paired samples t-test for ERC-ER from pretreatment to posttreatment for the treatment group, p = 1.0, and from intake to pre-treatment for the waitlist group, p = 0.49, revealed no significant differences.  - When treatment gains for all participants were analysed, a paired samples t-test for the whole sample combined revealed a significant difference in the scores for ERC-L/N from pretreatment (Time 1 for treatment group and Time 2 for waitlist group combined; M = 39.28, SD = 5.34) to posttreatment (Time 2 for treatment group and Time 3 for waitlist group combined; M = 35.11, SD = 4.46), p < 0.01, Cohen’s d= 0.77, suggesting a medium effect size. Although the ERC-ER increased from pretreatment (M = 22.22, SD = 2.21) to post-treatment (M = 22.78, SD = 2.67), these changes were not significant, p = 0.19. The increase in scores did, however, represent a small effect, Cohen’s d = 0.33. - Follow-up (treatment group only): no significant changes from pretreatment to follow-up on ERC-L/N (p=0.096) and ERC-ER (p=0.03). - A significant difference in parent confidence ratings for pretreatment versus posttreatment for parent perception of child’s ability to manage anger, p = .02, with increased parent confidence ratings for treatment group at posttreatment. There was also a significant difference in the treatment group from pretreatment to post-treatment with perception of child’s ability to manage anxiety, p = .01, with increased parent confidence ratings at Time 2. - For the whole group there was a significant difference in parent confidence ratings for pretreatment versus posttreatment for parent perception of child’s ability to manage anger, p < .01, and perception of child’s ability to manage anxiety, p < .01, with increased parent confidence ratings at posttreatment. |
| Scarpa et al. (2011) | Adapted group CBT for emotional regulation (Stress and Anger Management Program - STAMP) for young children (aged 4-8) vs waitlist (delayed-treatment control group) | Primary and secondary outcomes not specified. Ben and the Bullies and James and the Reading Group Vignettes - A child self-report of use of emotion regulation strategies in response to vignettes. ERC - Parental report on an emotion regulation scale. Behavioural Monitoring Sheet - Parental observation of anxiety/anger episodes Parent confidence ratings- parent-rated measure of their perceived self-confidence in managing their child's anxiety and anger and their perceived confidence in their child's skills level in managing their own anxiety and anger (only the later reported in this review). | Intervention content adaptations - Creative outlets and activities - Taking it slow Involving wider support network  - Parental involvement | Not reported. | Mental health outcomes using child-report of emotional regulation strategies; Emotion Regulation Checklist; Behavioural Monitoring Sheet; parent confidence ratings: - Emotion Regulation Checklist: no significant differences between groups in post-treatment scores. - Behavioural Monitoring Sheet: Frequency of anger/anxiety episodes per hour was significantly lower in intervention group (M = 0.16, SD = 0.05) compared to the waitlist group (M = 0.17, SD = 0.12), p < .05, effect size d=0.05. Duration in minutes per episode was marginally lower in intervention group (M = 2.57, SD = 2.10) compared to the waitlist group (M = 7.74, SD = 7.18), p < 0.10, d = 0.46.  - Use of emotion regulation strategies in response to vignettes: Children in the experimental group responded with a significantly greater average number of strategies than the delayed-treatment control group in response to vignettes (M = 4, SD = 2.45) compared to waitlist group (M = 1.29, SD = .95), p < 0.05, d = .65. - Parents in the experimental group reported significantly higher levels of confidence in their child’s ability to deal with anger (experimental group at Time 2 M 6.80 (SD=0.84); delayed treatment group at Time 2 M 3.43 (SD 0.98); p<0.05; d=0.89) and anxiety (experimental group at Time 2 M 6.60 (SD=0.89); delayed treatment group at Time 2 M 2.7 (SD 1.50); p<0.05; d=0.85) than in the delayed-treatment control group. |
| Swain et al. (2019) | Adapted group CBT for emotional regulation for young children (aged 4-8) (Stress and Anger Management Program - STAMP) | Primary and secondary outcomes not specified. Lability/Negativity subscale of ERC - parent-report measure of child's negative affect/ emotional states that may be displayed in childhood such as outbursts, anger and sadness.  Behavioural Monitoring Sheet - Parental observation of anxiety/anger outbursts. Questionnaire on perceived satisfaction and usefulness of the treatment - parent-report. Parent confidence ratings - parent-rated measure of their perceived self-confidence in managing their child's anxiety and anger and their perceived confidence in their child's skills level in managing their own anxiety and anger (only the later reported in this review). | Intervention content adaptations - Creative outlets and activities - Integration of emotion-focused strategies Involving wider support network  - Parental involvement | Acceptability - Treatment satisfaction: There was no significant difference between the treatment responder group and the non-responder group in rating of satisfaction, p=0.25. To open-ended questions, parents in the treatment responder group provided enthusiastic findings regarding changes they had seen in their child. Two parents in the treatment responder group found ways to bridge the gap between the therapy room and the classroom, whereas this was not noted in the treatment non-responder group. Parents in the treatment responder group made multiple references to the parent training component of STAMP, describing the psychoeducation, support and skills training components as helpful. Seven parents in the treatment responder group specifically mentioned the coping tools as being the most helpful part of the program, while one parent in the treatment non-responder group noted that ‘identifying tools’ was the least helpful part of the program for her child. Parents in the treatment non-responder group noted specific difficulties with generalization of skills and one indicated only ‘occasional’ use of the coping strategies by the child. When parents were asked to describe potential improvements, a mother in the treatment non-responder group indicated that the program should be more tailored to each child’s individual needs. In contrast, four parents in the treatment responder group asked for follow-up sessions, more time for discussion, or longer sessions. Parents in this group also seemed to answer the satisfaction surveys more completely, which could suggest conscientiousness or more dedication to the program, especially in terms of homework completion. | Effect of time (no comparison group) Mental Health Outcomes using Liability/Negativity ERC subscale; Behavioural Monitoring Sheet; Parent confidence ratings: - 12 child participants were classified as treatment responders*. Within the treatment responder group, patterns emerged in the behavioural monitoring data suggesting parallel decreases in duration and intensity of outbursts, while decreases in frequency were not associated with changes in intensity or duration. *A child was classified as a ‘treatment responder’ if he or she met at least two of the following four criteria: (1) statistically significant post-treatment decrease in Liability/Negativity, using the Reliable Change Index; (2) greater than 20% decrease in average intensity of outbursts; (3) greater than 20% decrease in frequency of outbursts; or (4) greater than 20% decrease in average duration of outbursts. - In the treatment responder group, there was a statistically significant pre (M 2.67 (SD 1.61)) to post (M 6.00 (SD 2.09)) increase in parent-reported confidence in their child’s ability to manage anger, p=0.002, and anxiety (pre: M 2.58 (SD=1.38); post: M 6.17 (SD2.33), p=0.002). In contrast, the treatment non-responder group showed no significant change (i.e., after Bonferroni correction) in parental confidence in their child’s ability to manage anger (pre: M 3.00 (SD=0.89); post: M 3.50 (SD 1.52), p=0.41), or anxiety (pre: M 2.83 (SD=1.47); post: M 4.17 (SD 1.33), p=0.07). |
| Sofronoff et al. (2017) | Adapted group parent-delivered cognitive behavioural emotional and social skills self-directed intervention (Secret Agent Society-SAS) with weekly therapist support to parents provided via Skype | Primary and secondary outcomes not specified.  ERSSQ - parent-rated measure of emotional regulation and social skills. SCAS - parent-rated measure of symptoms of anxiety in children. | Adjustments to the physical environment - Encourage the use of sensory resources and stimming Intervention content adaptations - Use of role play or modelling - Use of a rewards system - Taking it slow - Integration of cognitive behavioural approaches Involving wider support network - Involving parents in the intervention  - Involving school | Feasibility - Intervention drop-out rate: Several participants (n = 13/41 child participants; n = 12/38 parent participants) withdrew from the program for various reasons. Thus, 28/41 child participants and 26/38 parent participants completed the SAS program and post-intervention outcomes. At 6-week follow-up, only 18 children and 17 parents completed questionnaires. This is likely due to the timing of the follow-up period falling in the December to January holiday period in Australia. A self-directed format may not be suitable for all families as the study had a high attrition rate of approximately 32%, with some parents struggling to implement the program. Importantly, attrition analyses indicated significant differences between parents who discontinued and those who remained in the study. Parents who discontinued had a lower level of education and higher autism traits and were younger in age than those who remained in the study. Reasons provided by parents for discontinuing included other life events (e.g., marriage, illness, and re-location), time constraints, and difficulties engaging and or motivating their child to complete the program. - Intervention attendance rate: 28/41 child participants and 26/38 parent participants completed the SAS program. | Effect of time (no comparison group) Mental health outcomes using ERSSQ; SCAS: - The results showed a significant effect of Time for total SCAS score, F (2, 39) = 4.14, p = .023; Wilks’s lambda = .82; ηp2 = .17, indicating a large effect size; observed power = .70. The total SCAS scores significantly decreased from Time 1 (M = 25.56, SD = 15.18; pre-treatment) to Time 3, p = .020 (M = 21.53, SD = 12.85; post treatment). There was no significant difference between the scores on the SCAS at Time 1 (pre-treatment) and Time 2 (baseline period), p = .64, and there was no significant difference between scores on the SCAS at Time 2 (baseline period) and Time 3 (post-intervention), p = .10. There was a significant effect for time for total SCAS at 6-week follow-up compared to pre-intervention, Wilks’s lambda = .733, F (1, 17) = 6.18, p = .024; ηp2= .27; observed power = .65. This finding demonstrates significant reductions in child anxiety were reported at 6-week follow-up (M=17.33, SD=12.98) when compared with pre-intervention (M=23.44, SD=12.08). - A significant effect of Time for ERSSQ, F (1.46, 58.35) = 20.37, p = .000, ηp2 = .34; observed power = .99. The mean score for Time 3 on the ERSSQ (M 55.71, SD=14.31) was significantly different from Time 1 (M=45.27; SD=8.99), p = .0005 and Time 2 (M=46.58, SD=10.27), p = .0005, with scores on Time 3 significantly higher, indicating improved social and emotional regulation skills. There was no significant difference between the scores on the ERSSQ at Time 1 and Time 2. A significant effect of time for ERSSQ at 6-week follow-up compared to pre-intervention, F (1, 17) = 26.34, p < .001; ηp2 = .61; observed power = .99. The mean scores for ERSSQ at Time 4 (M=63.55, SD=13.54) were significantly higher when compared with Time 2 (M=46.44, SD=11/16), p < .001, indicating improved social skills when compared with pre-intervention. |
| Drusedau et al. (2022) | Bespoke group mindfulness-based intervention (The Tübinger Training for Autism Spectrum Disorders-TüTASS) for emotional regulation | Primary and secondary outcomes not specified. ILK - child- and parent-report measure of quality of life DIKJ - parent-report measure of depressive symptoms Child- and parent- report questionnaires specifically designed to assess feasibility and quality of the group training | Not applicable. | Feasibility Intervention attendance rate: 4 failed to attend all sessions Intervention drop-out rate: None of the participants dropped out.  Acceptability Satisfaction with care: Children enjoyed the intervention and profited very much. Both children and parents reported that the session focussing on emotions and body and the session focussing on strength and weakness were particularly helpful. Few negative comments about having homework and the weekly rhythm. Children (M = 4, SD = 1.22) and parents (M = 3.71, SD = 1.26) were well motivated to take part in the sessions (scale of 1 no to 5 yes totally). Children (M = 3.35, SD = 1.27) and parents (M = 3.72, SD = 0.89) reported that the training was helpful (scale of 1 very bad to 5 very good). Children (M = 4.35, SD = 0.86) and parents (M = 4.67, SD = 0.59) reported that it was good to have a group therapy setting (scale of 1 very bad to 5 very good). Parents (M = 4.5, SD = 0.51) reported that the overall concept was very good (scale of 1 not at all sensible to 5 very sensible). | Effect of time (no comparison group).  Mental health outcomes using DIKJ:  - No significant difference from pre (M = 52.87, SD = 11.59) to post (M = 54.04, SD = 11.96), t = 0.51, p = .616, d = 0.09. Quality of life using ILK P and ILK C: - No significant difference in ILK P scores from pre (M = 17.40, SD = 3.14) to post (M = 17.68, SD = 3.05), t = 0.50, p = .624, d = 0.10.  - No significant difference in ILK C scores from pre (M = 20.40, SD = 3.20) to post (M = 20.28, SD = 3.58), z = -0.08, p = .950, d = 0.03. |
| **CBT for various mental health needs** | | | | | |
| McGillivray et al. (2014) | Bespoke group CBT for anxiety, stress and depression vs Waitlist | Primary outcomes and secondary outcomes not specified.  DASS - self-report measure of depression, anxiety and stress.  ATQ - self-report measure of frequency of cognitive self-statements associated with depressed mood.  ASSQ - self-report measure of frequency of cognitive self-statements associated with anxiety. Self-report assessment questionnaire asking about any treatments for anxiety or depression they had received during the past 3 months. | Not applicable. | Feasibility  - Intervention attendance rate: 6/16 waitlist control group completed the programme, but it is unclear how many in the treatment group completed treatment.  - Intervention drop-out rate: All participants (n = 42: CBT n = 26, waiting list n = 16) completed pre- and post-treatment assessments. Participants in the waitlist group were then invited to take part in the intervention program. 6 of these participants subsequently completed the program, making the total of 32 people who finished the intervention program. Of the remaining participants from the waitlist group, 4 started the program but dropped out and 6 reported that they were no longer interested in participating due to changed personal circumstances. Participants were invited to return for a follow-up group session to undertake repeat assessments at 3 and 9 months after completion of the program. 27 participants completed the 3- and 9-month follow-up indicating a small attrition rate. | Mental health outcomes using DASS; ATQ; ASSQ:  - DASS overall: There was no significant effect for Group X Time interaction (p > .05). Participants improved over time regardless of whether they were in the treatment group or on the waitlist.  - ASSQ & ATQ: no significant effect for group X time interaction (p > .05).  - A subsequent analysis was undertaken with only those people who scored above the normal range on the depression (> 9), anxiety (> 7) and stress (> 14) subscales of the DASS, the ATQ > 60 and the ASSQ > 64. For the depression DASS subscale, a significant effect for Group X Time interaction (p < .05) was found for these participants. Significant effect for Group x Time interaction (p < .05) was seen for the DASS stress subscale, but not for anxiety DASS subscale (p > .05).  - For people who scored over 60 on the ATQ, there was no significant effect for Group X Time interaction (p > .05).  - For those who scored over 64 on the ASSQ, there was no significant effect for Group X Time interaction (p > .05)  - In order to determine whether the reduction of scores in the DASS depression and stress subscales was sustained over time, repeated measures analysis was conducted. Participants who were symptomatic and who had completed the treatment program and post group assessments were included. A significant main effect for time was found for the DASS depression subscale, p < .01; and the DASS stress subscale, p < .01. Follow up comparisons found that significant differences occurred between pre-group and post-group assessments, with no differences evident between scores at the 3- and 9-month follow-ups. Scores at the 9-month follow-up remained significantly lower than those obtained at the pre-program assessment.  Service use using a self-report assessment questionnaire:  - Most participants (78%) reported no change in their receipt of additional mental health treatments during the course of the intervention. During the study period, additional treatment for depression and for anxiety had been initiated for two (6.3%) and three participants (9.4%) respectively, while additional treatment for depression and for anxiety had been discontinued for five (15.6 %) and four participants (12.5 %) respectively. Treatment outside of the group CBT intervention was thus relatively stable over the study period. |
| Santomauro et al. (2016) | Bespoke group CBT for depression (‘Exploring depression’ programme) vs Waitlist | Primary and secondary outcomes not reported.  BDI-II: self-report, to measure symptoms of depression DASS-21: self-report, the depression scale was used as an additional measure for symptoms of depression. ERQ: self-report, to assess participants’ emotion regulation via use of cognitive reappraisal and expressive suppression. | Not applicable. | Feasibility - Intervention attendance rate: 100% attendance (including 23 one-on-one catch-up sessions). - Intervention drop-out rate: 1 out of 19 participants who started the intervention dropped out. Acceptability - Satisfaction with care: Of adolescents asked, 14/15 (93.3%) reported they enjoyed the programme. The group setting was considered by most adolescents to be the most helpful element. Adolescents generally found the tools helpful, but there were individual differences in which strategies were endorsed. | Mental health outcomes using BDI-II; DASS-21; ERQ:  - There was no significant group x time interaction on BDI-II scores, F(1,18) = 0.02, p = .893, η2 < .01 or significant main effect of condition or time. - There was no significant group x time interaction on DASS-21 depression subscale scores, F(1,18) = 3.86, p = .065, η2 = .17, or significant main effect of condition or time. Further analysis showed that the simple effect of time for the intervention group was significant, F (1,9) = 6.11, p = .035, η2 = .40, showing a significant improvement from pre (M = 24.20, SD = 8.97) to post (M = 17.20, SD = 8.95). No significant simple effect of time for the waitlist group. - Secondary analysis revealed significant improvement on BDI-II scores from week five (M = 28.38, SD = 16.39) and post (M = 22.99, SD = 15.76), t(58) = 3.16, p = .006, which was maintained at the booster session (M = 19.95, SD = 15), t(68) = 3.55, p = .002, but no significant differences between pre and after 5 weeks p = .868. Significant improvements were also seen on BDI-II scores from pre to booster session, t (68) = 3.59, p = .002. However, at 3-month follow-up BDI scores increased (M = 28.27, SD = 15.50), t (68) = 3.48, p = .003.  - Secondary analysis revealed significant improvements on DASS-21 depression subscale scores from pre (M = 23.33, SD = 9.87) to post (M = 16.33, SD = 12.02), t (51) = 2.48, p = .024, which were maintained at booster session (M = 15.05, SD = 7.69), t (51) = .66, p = .518. Significant improvements were seen on DASS-21 depression subscale scores from pre to booster session, t(51) = 3.51, p = .003, but levels of depression significantly increased at 3-month follow-up, t(51) = 3.07, p = .008. - Regarding emotion regulation, there was no significant group x time interaction on the use of cognitive appraisal, F (1,18) = 14, p = .719, η2 = .01 and on the use of expressive suppression, F (1,18) = .22, p = .646, η2 = .01. |
| Cooper at al. (2018) | Adapted individual CBT for various mental health needs | Primary and secondary outcomes not specified.  Short survey developed for this study, which sought information such as knowledge/use of adaptations of CBT as outlined in the NICE guidance. | Communication accommodations  - Use of clear, simple and preferred language  - Use of simple, written material and visual aids  Accommodate individual differences  - Encourage individual's hobbies and interests  Structural or procedural adaptations  - Format of intervention  - A structured and predictable approach  Intervention content  - Simplified and structured content  - Integration of emotion-focused strategies  - Integration of cognitive-behavioural approaches  Involving wider support network  - Parental involvement | Feasibility  % of therapists endorsing the following adaptations:  - Behavioural strategies to introduce change (n = 37, 74%); Using plain English more than with other clients (n = 35, 70%); A more structured and concrete approach to therapeutic work (n = 35, 70%); Psychoeducation about emotions (n = 34, 68%); More written and visual information than I usually use (n = 30, 60%); Discussing individual hobbies and interests as part of therapy (n = 29, 58%); Involving a family member or partner in sessions (n = 24, 48%); Avoiding metaphors in therapy (n = 20, 40%); Shorter sessions (n = 14, 28%); Cognitive strategies to introduce change (n = 14, 28%); Other (n = 5, 10%); Longer sessions (n = 1, 2%).  Acceptability  - Satisfaction with care: Most respondents favoured a cognitive behavioural approach with autistic clients, rating it 7.17/10 for helpfulness, and 74% having ever used this approach. 16% of responders used systemic approaches, giving it an average rating of 6.75/10 for helpfulness. 14% of respondents used eclectic approaches, giving it an average of 6.29/10 for helpfulness. 4% of responders used psychodynamic approaches giving it a 3.1/10 rating for helpfulness. | Not applicable. |
| **EMDR for PTSD** | | | | | |
| Fisher at al. (2022) | Adapted EMDR for PTSD | **Primary and secondary outcome measures not specified.**  Round 2 survey comprised of statements generated by thematically analysing responses to Round 1. Statements rated on a 5-point scale (1 = I always do this, 2 = I often do this, 3 = I sometimes do this, depending on the client, 4 = I never do this, 5 = I think this should not be done at all). | Increase knowledge and detection of autism  - Clinician training and skills  Adjustments to the physical environment  - Provide environmental and practical adjustments  - Encourage the use of sensory resources and stimming   Communication accommodations  - Use of clear, simple and preferred language        - Use of simple, written material and visual aids   Accommodate individual differences  - Evaluate individual needs and preferences  - Encourage individual's hobbies and interests  - Tailor practice to individual needs and preferences  Structural or procedural adaptations - A structured and predictable approach   Intervention content  - Taking it slow  - Consider the role of autism  - Integration of emotion-focused strategies  - Integration of cognitive-behavioural approaches  Involving wider support network - Parental involvement | Feasibility  Elements of EMDR that ≥ 80% of therapists often or always incorporate in therapy with autistic clients: 88% normalise experiences, 86% use very clear language, 91% be flexible and creative, 84% be aware of how they communicate their level of arousal through behaviour and use this information to evaluate how they are coping during sessions, 91% take time to understand the language they use around thoughts and emotions, and mirror this, 91% respond to the person in front of you, 95% be open to learning from the client and celebrate each person's uniqueness, 86% consider the role of autism within the conceptualisation, 98% consider small ‘t’s and big ‘T’s as possible targets, 88% use their background and history to identify resource possibilities, 84% try a range of different types of bilateral stimulation, 80% be particularly mindful of language as people may be very sensitive to failure and 'getting it wrong', 83% use their own language to describe emotions, 80% make time for the person to debrief about their week.  Elements of EMDR that ≥ 80% of therapists sometimes incorporate in therapy: 93% assess sensory preferences and sensitivities, 100% be more directive in style, 86% use visual aids, 93% avoid metaphors, 98% ask about and include special interests throughout, 100% always offer sessions at the same time and place, 100% take graduated/ progressive approach towards full trauma processing, 81% use flash, 93% focus on quality of life and functioning, 98% share a plan in advance for each session to manage expectations, 95% change the environment to reduce sensory demands, 100% provide extra psychoeducation around trauma, arousal and feeling physiologically overwhelmed, 98% slow down every phase, 88% use storytelling, 100% focus on building a positive self-image and coping strategies rather than pathologizing and eliminating symptoms, 100% prioritise the therapeutic relationship above everything else, 95% don't insist on or encourage eye contact, 81% use visual or simplified version of ratings scales, 100% be ready to reformulate throughout the therapy and to shift the focus, 93% give explicit permission to ask questions, 86% obtain information from other people as well as the person themselves, 100% expect to add to history taking throughout the therapy as new information emerges, 93% focus first on strengths and interests and then move onto problems and history, 86% vary the way you work, 88% create a visual timeline, 95% spell things out in black and white and be more directive than usual, 98% use an image of a place rather than imaginal calm place, 95% think in terms of a 'positive engaging focus' rather than necessarily a 'calm place', 93% be creative with the calm place, 81% encourage them to use stimming behaviour as self-soothing if it works, 95% use fantasy figures as resources in preparation stage, 93% use their special interests and how they feel when engaged in it as a resource, 81% include exercises to facilitate accessing emotions, 81% include props to help them identify emotions , 84% install a positive self-view as a resource, 91% be very clear with clients what the preparation phase is about and why it is necessary, 100% ask for all the elements but if they cannot provide information, go with whatever is given in assessment phase, 100% assure you are well tuned in to the client before starting, 80% use a progressive approach to processing, starting with the 'tip of the finger', 88% use any sensory modality as a target, not necessarily an image, 90% use a present-day target first, 88% use a literal description. Ask the person to explain what we would see if looking at a photo or a still of a movie, 95% proceed without an image if they struggle with finding an image, 98% offer alternatives, prompts and suggestions for cognitions, 93% skip negative cognition altogether if it causes problems, 88% allow the positive cognition to emerge during processing rather than identifying it beforehand, 95% use more prompts and suggestions to find positive cognition, 93% use softeners for the positive cognition, 100% be aware of the possibility of sensory overload, 98% use more directive interweaves than usual, 98% let the client choose and control length of sets in desensitisation phase, 93% use shorter sets and a more frequent return to target, 90% do not expect generalisation, 85% repeat their feedback to them to aid processing, 80% use more physical movement, 90% start with short sets and build up tolerance from there, 85% do more cognitive work at this stage if necessary to identify a positive cognition, 98% take longer to close down and leave extra time for a debrief, 95% end with a relaxing and positive activity, 100% offer clear guidance on what to do after the session and what they might experience after the session , 100% include your own thoughts as part of the debrief, 100% focus on quality of life and functioning to assess progress, 100% offer your own observations of what has changed, 95% don't emphasise keeping logs between sessions if difficult for the client. | Not applicable. |

*Service-level strategies*

| **Authors** | **Strategy vs comparison** | **Outcomes/measures** | **Adaptation categories and sub-categories** | **Acceptability/Feasibility findings** | **Effectiveness findings** |
| --- | --- | --- | --- | --- | --- |
| Detection of autism in mental health care | | | | | |
| Ford et al. (2019) | Identification of autism using the DAWBA | Primary and secondary outcome measures not specified. DAWBA: parent/teacher reported diagnostic assessment tool to determine autism diagnosis. Referral letters to extract presenting problems and classifying into independent categories coded present or absent including autism. Practitioner autism diagnosis and psychiatric disorders were collected via a brief questionnaire. | Increase knowledge and detection of autism - Introduction of screening tools for the detection of autism | Not applicable. | Autism detection using research diagnosis (DAWBA), presenting problems on referral and practitioner diagnosis: - Moderate agreement between referrer and research diagnosis (Kappa = 0.51) at baseline, which reduced over two years. - Moderate agreement between practitioner's and research diagnosis (Kendall's tau = 0.60) at baseline, which reduced over two years. - Fair agreement between referrer and practitioner's diagnosis (Kendall's tau = 0.36) at baseline. - Highest diagnostic instability occurred among children who practitioners considered to be potentially autistic, but who did not meet research criteria. |
| Hollocks et al. (2019) | Identification of autism using the SCQ | Primary and secondary outcome measures not specified. SCQ lifetime form: parent-report questionnaire of developmental history in the domains of social communication and language development  SCQ current form - teacher-report questionnaire assessing child's behaviour over the past 3 months. ADOS 2nd edition modules 3 and 4 - diagnostic assessment of autism | Increase knowledge and detection of autism - Introduction of screening tools for the detection of autism | Not applicable. | Autism detection using SCQ: - Parent/caregiver-report SCQ correctly classified 53% of participants (AUC - 0.52) and had a cut-off score of 15, with sensitivity of 83.7% and specificity of 12.5% - Teacher-report SCQ correctly classified 56% of participants (AUC - 0.56) and had a cut-off score of 15, with sensitivity of 72.9% and specificity of 35.5% - Non-significant relationship between parent and teacher reported autism traits (r = -0.07, p = .58). |
| Stadnick et al. (2015) | Identification of autism using the ADOS | Primary and secondary outcome measures not specified. Presence and type of DSM-IV autism diagnosis were extracted from evaluation reports. ADOS: clinician-administered assessment to assist in the diagnosis of autism SRS: parent-report measure that examines the severity of autistic traits  SCQ: parent-report measure that examines the presence of autistic traits. | Increase knowledge and detection of autism - Introduction of screening tools for the detection of autism | Not applicable. | Autism detection using ADOS, SCQ and SRS: - 35 children received a DSM-IV ASD diagnosis, of which 37% received a diagnosis of autistic disorder, 29% a diagnosis of Asperger’s disorder and 34% a diagnosis of PDD-NOS. 75% (n = 18/35) scored within the 'severe' range of concern on the SRS and 89% (n = 23/35) scored above the autism cutoff on the SCQ. - Of the 27 children who did not receive a final autism diagnosis, 70% were classified as "non-spectrum" on the ADOS. The remaining 8 children scored in the "autism" or "ASD" range on the ADOS but did not receive a final diagnosis. Of those with available data, 3 scored above the autism cutoff on the SCQ and four scored within the "severe" range on the SRS.  - 7/8 cases attributed elevated ADOS scores to symptoms of anxiety disorder or symptoms of an ADHD diagnosis. |
| **General adaptations to standard practice** | | | | | |
| Petty et al. (2021) | Adaptations in general and specialist mental health services for autistic people | Freelisting interview guide was developed, which included questions in relation to 1) adaptations made prior to or within a client appointment 2) service-level adaptations 3) adaptations made in the physical environment and 4) adaptations relating to a person's gender. | Increase knowledge and detection of autism  - Clinician training and skills  Adjustments to the physical environment - Provide environmental and practical adjustments  - Encourage the use of sensory resources and stimming  Communication accommodations  - Use of clear, simple and preferred language  - Use of simple, written material and visual aids  Accommodate individual differences  - Evaluate individual needs and preferences  - Tailor practice to individual needs and preferences  Structural or procedural adaptations - A structured and predictable approach | Feasibility Adaptations before attending an appointment (Salient items are considered to be consensus responses to the question):  - Check the suitability of the sensory environment - listed by 60% of staff, M rank = 6.2, salience = .334 (salient)  - Find out about the client in advance - listed by 33% of staff, M rank = 1.6, salience = .306 (salient)  - Be clear in communication - listed by 33% of staff, M rank = 4.6, salience = .206 (salient)  - Check suitability of lighting - listed by 33% of staff, M rank = 7.6, salience = .179 (salient)  - Ensure the client is prepared for what will happen - listed by 20% of staff, M rank = 3.0, salience = .168 (salient)  - Find out if the client has sensory needs - listed by 27% of staff, M rank = 6.3, salience = .141 (not salient)  - Be prepared to adjust communication - listed by 20% of staff, M rank = 4.3, salience = .141 (not salient)  - Find out about the client in advance from significant people - listed by 20% of staff, M rank = 5.3, salience = .139 (not salient)  - Find out about the client in advance from case notes - listed by 13% of staff, M rank = 1.5, salience = .129 (not salient)  - Ensure the client is prepared about the purpose of the appointment - listed by 13% of staff, M rank = 3.0, salience = .117 (not salient)   Service-level adaptations:  - Provide a sensory friendly environment - listed by 53% of staff, M rank = 2.6, salience = .448 (salient)  - Make information available for clients about the service - listed by 46% of staff, M rank = 4.1, salience = .346 (salient)  - Ensure suitable noise levels - listed by 53% of staff, M rank = 4.4, salience = .334 (salient)  - Adapt communication - listed by 40% of staff, M rank = 4.7, salience = .244 (salient)  - Keep to plain design - listed by 33% of staff, M rank = 3.4, salience = .230 (salient)  - Offer flexible session timings - listed by 33% of staff, M rank = 3.2, salience = .203 (salient)  - Maintain the specialist skillset of staff - listed by 27% of staff, M rank = 2.8, salience = .181 (salient)  - Adapt written correspondence - listed by 20% of staff, M rank = 2.0, salience = .181 (salient)  - Utilise a protected building or space - listed by 20% of staff, M rank = 2.0, salience = .167 (salient)  - Use signs up to modify the environment - listed by 20% of staff, M rank = 3.3, salience = .134 (not salient)    Adaptations within a client appointment:  - Communicate clearly - listed by 60% of staff, M rank = 2.0, salience = .536 (salient)  - Avoid ambiguity - listed by 27% of staff, M rank = 2.5, salience = .231 (salient)  - Offer a flexible and individualised approach - listed by 33% of staff, M rank = 5.0, salience = .204 (salient)  - Check for understanding - listed by 27% of staff, M rank = 4.8, salience = .180 (not salient)  - Agree etiquette for making eye contact - listed by 27% of staff, M rank = 3.5, salience = .177 (not salient)  - Slow the pace of communication - listed by 20% of staff, M rank = 2.7, salience = .169 (not salient)  - Avoid idioms - listed by 20% of staff, mean rank, M rank = 3.3, salience = .159 (not salient)  - Monitor own communication - listed by 20% of staff, M rank = 3.0, salience = .156 (not salient)  - Consider the room seating arrangement - listed by 20% of staff, M rank = 3.3, salience = 1.52 (not salient)  - Use agendas - listed by 27%, M rank = 6.5, salience = .151 (not salient)   Gender:  - Know how someone identifies - listed by 60% of staff, M rank = 1.9, salience = .444 (salient)  - Maintain awareness of gender differences - listed by 60% of staff, M rank = 2.4, salience = 4.06 (salient)  - Do not make assumptions - listed by 33% of staff, M rank = 2.6, salience = .243 (salient)  - Know pronoun or name preferences -listed by 40% of staff, M rank = 2.7, salience = .230 (salient)  - Use a preference notifications system - listed by 13% of staff, M rank = 1.5, salience = .126 (not salient)  - Maintain awareness of gendered socialisation - listed by 13% of staff, M rank = 2.5, salience = .100 (not salient)  - Check suitability of clinician gender - listed by 13% of staff, M rank = 3.0, salience = .083 (not salient)  - Adapt questioning for female representation - listed by 13% of staff, M rank = 3.0, salience = .067 (not salient)  - Offer gender appropriate resources - listed by 7% of staff, M rank = 1.0, salience = .067 (not salient)  Adaptations to the physical environment:  - Reduce noise - listed by 80% of staff, M rank = 3.8, salience .559 (salient)  - Provide adjustable lighting - listed by 60% of staff, M rank = 3.6, salience = .442 (salient)  - Neutralise decor - listed by 47% of staff, M rank = 4.4, salience = .306 (salient)  - Offer space - listed by 40% of staff, M rank = 2.7, salience = .279 (salient)  - Reduce scents - listed by 33% of staff, M rank = 5.4, salience = .200 (not salient)  - Neutralise all sensory demands - listed by 27% of staff, M rank = 4.0, salience = .172 (not salient)  - Control outside noise - listed by 33% of staff, M rank = 6.2, salience = .162 (not salient)  - Avoid patterns - listed by 20% of staff, M rank = 3.3, salience = .144 (not salient)  - Reduce the number of items in the environment - listed by 20% of staff, M rank = 4.3, salience = .138 (not salient)  - Provide sensory resources - listed by 27% of staff, M rank = 6.0, salience = .129 (not salient) | Not applicable. |
| Jones et al. (2021) | Adaptations in psychiatric in-patient care for autistic people | Survey covering a range of domains including adaptations within the settings. | Increase knowledge and detection of autism  - Introduction of screening tools for the detection of autism   Adjustments to the physical environment - Provide environmental and practical adjustments  - Encourage the use of sensory resources and stimming  Communication accommodations  - Use of simple, written material and visual aids  - Provide communication support  Accommodate individual differences  - Evaluate individual needs and preferences  - Tailor practice to individual needs and preferences | Feasibility  Prevalence of adaptations across participating clinics: 81% of in-patient units (64/79 respondents) have specific assessments on ‘likes and dislikes’ of patients with autism; 82% of in-patient units (65/79 respondents) have assessments of coping strategies; Care plans based on individual needs specific to people with autism were available in 71% of units (53/75 of respondents); Only two-third of units (66%, 52/79 respondents) provided communication passports; 62% (49/79 respondents) a bespoke sensory assessment; The presence of a standardised protocol for people with autism (specific protocol for admission, assessment and management of people with autism) was available only in a fifth of the respondent’s units (21%, 17/79 respondents); Of all units 63% provided visual signage or orientation tools; 76% were able to provide visual timetables; 74% units were able to provide visual help/cue cards; 60% units were able to provide social stories; One of seven units (14%) reported being unable to provide any extra adaptations beyond communication support for people with autism; Open access low-stimulus area (52% of units providing this, 41/79 respondents); On request low-stimulus area (42%, 33/79); Scheduled access low-stimulus area (15%, 12/79); Lighting adaptations (23%, 18/79); Ability to adapt meal plans to sensory requirements (51%, 40/79); Noise adaptations (14%, 11/79); Other adaptations (4%, 3/79); No adaptations provided (15%, 12/79).  - The survey looked at the assessments in place for in-patient services to support people with autism in a person-centred manner as per current good practice. 90% (71/79 respondents) of units reported offering autism assessment.  - Other adaptations (i.e., tools/strategies designed for specifically supporting autistic people) mentioned in the free text as made available in the respondent's in-patient units included ear defenders, weighted blankets, stress ball and relaxing music (no proportions provided in paper). | Not applicable. |
| Spain et al. (2017) | Adaptations to standard inpatient and outpatient care for autistic people | A topic guide was used consisting of pre-specified, semi-structured questions including any adaptations were made to their standard clinical approach. | Adjustments to the physical environment - Provide environmental and practical adjustments  Communication accommodations  - Use of clear, simple and preferred language Accommodate individual differences  - Tailor practice to individual needs and preferences | Feasibility The following adaptations were made by professionals to their standard practice: - Ensure that appointments are offered at a convenient time. - Ensure that the clinical environment is not overly stimulating. - Use of didactic questions - Use of Socratic style  - Little reliance on metaphors or colloquialisms, which may prove difficult for autistic individuals to understand - Encourage people to be 'active participants' whereby their views about the pace and content of clinical work are sought, as they may have had limited opportunities to develop assertiveness skills. This means encouraging people to feel confident to say what they think. | Not applicable. |
| Strategies for improving clinicians’ skills and autism knowledge | | | | | |
| Dreiling et al. (2022) | Extension for community healthcare outcomes project (Project ECHO) - a tele-mentoring platform to support mental health professionals when working with autistic people with co-occurring mental health difficulties. | Primary and secondary outcome measures not specified.  PCASE Survey: self-report measuring mental health provider level of confidence in effectively enacting the treatment strategy described.  Knowledge test - self-report measure of mental health provider autism knowledge.  Problem-solving scenarios: self-report measure to assess changes in provider’s clinical problem-solving skills using evidence-based strategies that were discussed during ECHO Autism sessions.  Satisfaction survey: self-report. Participants were also invited to share their thoughts and suggestions for improvement in free-response sections. | Increase knowledge and detection of autism  - Clinician training and skills | Feasibility  - Attendance rate: 88.2% (n = 45) of participants attended at least 80% of ECHO sessions (average attendance was 8.78/10 sessions). Providers who attended less than 60% of ECHO sessions (n = 4), who attended more than 60% of sessions but failed to complete post-assessment measures (n = 1), or who failed to meet both attendance and pre-post questionnaire completion (n = 30) were excluded from the analysis, yielding a final sample size of 51.  Acceptability  - Satisfaction (quantitative) (M = 1.32, range 1-2) was rated highly (5-point scale, with “1” indicating the highest degree of satisfaction).  - Satisfaction (qualitative): responses were positive, suggestions for improvement included ideas for additional didactic topics (e.g., gender and ASD) and recommendations for future ECHO Autism cohorts made up of previous participants to gain advanced-level training including additional opportunities for case presentations and feedback. | Effect of time (no comparison group).  Outcomes on the way to improving care:  Clinicians’ self-efficacy using PCASE Survey; autism knowledge using the Knowledge test; problem-solving skills using Problem-solving scenarios:  - Significant improvements in self-efficacy from pre (M = 64.90, SD = 13.36), to post (M = 85.29, SD = 11.10), p < .001.  - Significant improvements in autism knowledge from pre (M = 11.06, SD = 2.77), to post (M = 14.31, SD = 2.56), p < .001.  - Significant improvements in awareness in best-practice treatment considerations for autistic individuals from pre (M = 0.92, SD = 0.23), to post (M = 1.34, SD = 0.18), p < .001. |
| Helverschou et al. (2021) | Autism, Intellectual Disability and Psychiatric Disorder (AUP) Network consisting of meetings and seminars to guide mental health professionals in providing specialised mental health care to autistic people with ID. | Primary and secondary outcome measures not specified.  PAC: caregiver-report of mental health. | Increase knowledge and detection of autism  - Clinician training and skills | Not applicable. | Effect of time (no comparison group).  Psychiatric assessment using PAC:  - Significant improvements in proportion with psychiatric disorders from referral to after 12 months p < .001, but not from after 12 months to 24-27 months. |
| Kuriakose et al. (2018) - linked to Cervantes et al. (2019) | ASD-CP Pathway - Comparing pre-implementation and post-implementation | **Primary and secondary outcome measures not specified.**  Service use data were extracted from medical records. | Increase knowledge and detection of autism  - Clinician training and skills | Not applicable. | Effect of time (no comparison group).  Service use extracted from medical records: - There was a significantly fewer youth with holds/restraints in Children's Comprehensive Psychiatric Emergency Program (CCPEP) and inpatient units in the 18 months post implementation group compared to the pre implementation group (fisher's exact = .050, p = .039). - There were no significant differences in inpatient length of stay (p = .10), total length of stay (p = .07), brief-stabilisation unit holds/restraints (p = .06), inpatient holds/restraints (p = .29), total holds/restraints (p = .19), total intramuscular medication (p = .26) between the pre implementation group and the 18 months post implementation group. |
| Cervantes et al. (2019) | ASD-CP Pathway - Comparing pre-implementation, post-implementation and follow-up | **Primary and secondary outcome measures not specified.**  Service use data were extracted from medical records. | Increase knowledge and detection of autism  - Clinician training and skills | Not applicable. | Effect of time (no comparison group).  Service use extracted from medical records: - There were significantly fewer brief-stabilisation unit holds/restraints (p = .013, r = -0.44), total holds/restraints (p = .012, r = - 0.45) and total intramuscular medication (p = .004, r = - 0.51) in the 18 months follow up group compared to the pre implementation group, but not between the 18 months post implementation group and the pre implementation group. - There were no significant differences in inpatient length of stay, total length of stay and inpatient holds/restraints between the pre implementation group and the 18 months post implementation group and between the pre implementation group and the 18 months follow up group (p > .05).  - Significantly smaller proportion of participants in the follow up group experienced a hold/restraint than in the pre implementation group (χ2(1) = 7.04, p = .001) and in the 18-month post implementation group compared to the pre implementation group (χ2(1) =4.36, p=0.04). No significant differences in the proportion of youth experiencing any holds/ restraints between the 18 months post implementation group and the 18 months follow up group (p = .39). |

*Note.* **ADIS-C/P** = Anxiety diagnostic interview schedule - child and parent version, **AM** = Anxiety management, **ASD-CP** = Autism Spectrum Disorder Care Pathway, **ASSQ** = Anxious self-statements questionnaire, **ATQ** = Automatic thoughts questionnaire, **BAI** = Beck Anxiety Inventory; **BAT** = Behavioural Approach Task, **BAY-I** = Beck Youth Anxiety – Inventory, **BDI** = Beck Depression Inventory, **BDI-II** = Beck Depression Inventory-II, **BIACA** = Behavioural Interventions for Anxiety in Children with Autism, **CAISP** = Childhood Anxiety Impact ScaleParent, **CALIS** = Child Anxiety Life Inference Scale, **CAPE** = Children’s Assessment of Participation and Enjoyment, **CASI-Anx** = Child and Adolescent symptom inventory-4 autism anxiety scale, **CATS** = Children's Automatic Thoughts Scale, **CBCL** = Child Behaviour Checklist, **CBCL-Anxious/Depressed** = Child Behavioural Checklist Anxious/Depressed subscale, **CBCL-Anx** = The Child Behaviour Checklist anxiety subscale, **CBT** = Cognitive behavioural therapy, **CGAS** = Children’s Global Assessment Scale, **CGI-A** = Clinician Global Impression of Anxiety, **CGI-I** = The Clinical Global Impression – Improvement, **CGI-S** = Clinical Global Impression – Severity, **CI** = Confidence intervals, **CIS-P** = Columbia impairment scale - parent version, **CQ** = Coping Questionnaire, **CSR** = Clinical severity rating, **CSRs** = Clinician severity ratings, **CYP** = Children and young people, **DASS** = Depression anxiety stress scale, **DASS-21** = Depression anxiety stress scale – 21, **DAWBA** = Development and Well-Being Assessment, **DD-CGAS** = Developmental disabled children's global assessment scale, **DIKJ** = Depression inventory for children and adolescents, **D-YBOCS** = Dimensional Yale-Brown Obsessive Compulsive Scale, **ECHO** = Extension for community healthcare outcomes, **EMDR** = Eye Movement Desensitisation and Reprocessing, **ERC** = Emotion Regulation Checklist, **ERQ** = Emotion Regulation Questionnaire, **ERSSQ** = Emotion Regulation and Social Skills Questionnaire, **ESQ** = The Experience of Service Questionnaire, **FET** = Family-based exposure-focused treatment, **FLIS** = Family Life Impairment Scale, **FSSC-R** = Fear survey schedule for children—revised, **FYF** = Facing Your Fears, **GAD** = Generalised anxiety, **GAF** = Global assessment of functioning, **HAM-A** = Hamilton Rating Scale for Anxiety, **HAM-D** = Hamilton rating scale for depression, **ILC** = Inventory of Life Quality in Children, **ILK** = Inventory for assessment of quality of life in children and adolescents, **IUS** = Modified Intolerance of Uncertainty Scale, **K10** = Kessler Psychological Distress Scale, **KFQ** = Koala Fear Questionnaire, **KSADS-E** = Kiddie Schedule for Affective Disorders and Schizophrenia for DSM-IV, Epidemiologic Version, **K-SADS-PL** = The Schedule for Affective Disorders and Schizophrenia for School-Age Children–Present and Lifetime version, **LSAS** = Liebowitz Social Anxiety Scale, **LSAS-SR** = Liebowitz Social Anxiety Scale—Self-Report, **M** = Mean, **MASC-P** = Multidimensional anxiety scale for children-parent version, **MASS** = Multidimensional Adolescent Satisfaction Scale, **MASSI** = Multimodal Anxiety and Social Skills Intervention, **OCD** = obsessive compulsive disorder, **OCI-R** = Obsessive compulsive inventory-revised, **OST** = One-Session Treatment, **PAC** = Psychopathology in Autism Checklist, **PARS** = Paediatric anxiety rating scale, **PAS** = Preschool Anxiety Scale, **PCASE** = Primary Care Autism Self-Efficacy Survey, **PCTPRS** = Primary Care Therapy Process Rating Scale, **PR-CHOCI-R** = Children's Obsessive-Compulsive Inventory-Parent Version, **PSWQ** = Modified Peen State Worry Questionnaire, **PTSD** = Post traumatic stress disorder, **RCADS** = Revised Child Anxiety and Depression Scale, **RCMAS** = Revised children's manifest anxiety scale, **SACA** = Service assessment for children and adolescents, **SAS** = Secret Agent Society, **SCARED** = The Screening for Childhood Anxiety and Related Emotional Disorders, **SCAS-C/P** = Spence Children's Anxiety Scale - child/parent report, **SCQ** = Social Communication Questionnaire, **SD** = standard deviation, **SEP** = Separation anxiety, **SIAS** = Social Interaction Anxiety Scale, **SOC** = Social anxiety, **SPAS** = Spence Preschool Anxiety Scale, **SPI** = Social Phobia Inventory, **SpP** = Specific phobia, **SPS** = Social Phobia Scale, **SRS** = Social Responsiveness Scale, **STAI-T** = State-trait anxiety inventory - trait scale, **STAMP** = Stress and Anger Management Programme, **SUDs** = Subjective Units of Distress, **SWQ-P** = Social Worries Questionnaire-Parent, **T** = timepoint, **TALAA** = Treatment of Anxiety in Late Adolescents with Autism, **TAU** = Treatment as usual, **TPOCS-A** = Therapy Process Observational Coding System - Alliance scale, **VIQ** = Verbal Intelligence Quotient, **WL** = Waitlist, **WSAS** = Work and social adjustment scale, **YBOCS** = Yale-Brown Obsessive Compulsive Scale.

**Table S13**. Meta-regression analyses

|  | **Moderators** | **Coefficient** | **95% CI** | **SE** | ***Z* value** | ***p* value** |
| --- | --- | --- | --- | --- | --- | --- |
| **Parent** | **Type of CBT** | | | | | |
|  | Intercept: Adapted (n = 8) | 0.83 | 1.45 to 0.33 | 0.32 | 2.60 | p = .009** |
|  | Bespoke (n = 4) | -0.74 | -1.80 to 0.33 | 0.55 | -1.35 | *p* = .176 |
|  | Test of moderators: *QM*(1) = 1.83, *p* = .176, *R*^2^ = 0% | | | | | |
|  | Test of heterogeneity: *QE*(10) = 64.45, *p < .*001***; *I*^2^ = 84.49% | | | | | |
|  | **Format of CBT** | | | | | |
|  | Intercept: Combined (n = 2) | -0.04 | -1.33 to 1.25 | 0.66 | -0.06 | *p* = .950 |
|  | Group (n = 5) | 1.10 | -0.43 to 2.63 | 0.78 | 1.41 | *p* = .158 |
|  | Individual (n = 5) | 0.40 | -1.12 to 1.93 | 0.78 | 0.52 | *p* = .602 |
|  | Test of moderators: *QM*(2) = 2.47, *p* = .291, *R*^2^ = 0% | | | | | |
|  | Test of heterogeneity: *QE*(9) = 63.03, *p < .*001***; *I*^2^ = 85.72% | | | | | |
| **Clinician** | **Type of CBT** | | | | | |
|  | Intercept: Adapted (n = 6) | 0.84 | 0.44 to 1.24 | 0.20 | 4.16 | *p* < .001*** |
|  | Bespoke (n = 6) | -0.72 | -1.27 to -1.78 | 0.28 | -2.60 | *p* = .009** |
|  | Test of moderators: *QM*(1) = 6.75, *p* = .009**, *R*^2^ = 49.39% | | | | | |
|  | Test of heterogeneity: *QE*(10) = 21.04, *p* = .020**; *I*^2^ = 52.48% | | | | | |
|  | **Format of CBT** | | | | | |
|  | Intercept: Combined (n = 2) | 0.09 | -0.63 to 0.80 | 0.36 | 0.24 | *p* = .812 |
|  | Intercept: Group (n = 5) | 0.11 | -0.72 to 0.95 | 0.42 | 0.27 | *p* = .790 |
|  | Individual (n = 5) | 0.84 | -0.01 to 1.69 | 0.43 | 1.94 | *p* = .052 |
|  | Test of moderators: *QM*(2) = 6.48, *p* = .039*, *R*^2^ = 39.47% | | | | | |
|  | Test of heterogeneity: *QE*(9) = 20.97, *p* = .012*; *I*^2^ = 57.09% | | | | | |

*Note*. **CBT** = Cognitive Behavioural Therapy. **p* < .05, **p <.01, ****p* < .001

**Table S14.** Predictors of outcome (*N* = 5)

| **Authors (year)** | **Description of strategy** | **Adaptation (sub)categories** | **Predictors of outcome** |
| --- | --- | --- | --- |
| Bemmer et al. (2021) | Adapted group CBT for social anxiety. | **Intervention content**  - Simplified and structured content  - Integration of cognitive-behavioural approaches  - Integration of social skills training | **Demographics** (age, sex, IQ estimate, and autism symptomatology) and **pre-intervention social functioning** did not statistically significantly predict changes on the Liebowitz Social Anxiety Scale—Self-Report total score. |
| A. J. Russell et al. (2013) | Adapted individual CBT for OCD | **Communication accommodations**  - Use of simple and preferred language  - Use of simple, written material and visual aids  **Intervention content**  - Simplified and structured content  - Integration of emotion-focused strategies  - Integration of cognitive-behavioural approaches | **Age, verbal IQ, and autism symptomatology** were not statistically significantly associated with treatment outcome (the percentage change in total obsessive compulsive disorder severity scores). |
| Solish et al. (2020) | Bespoke group CBT for anxiety (‘FYF’ programme) | Not applicable. | Although correlation analysis suggested that child’s baseline **IQ** was negatively correlated with parent-rated improvement in anxiety (r= -0.32, p= 0.008), the linear regression models showed that this was not a significant predictor. The correlations with age and autism symptomatology did not reach significance. The linear regression models found that baseline **IQ, age, and autism symptomology** did not predict levels of change in anxious symptoms (parent- and child-report) over time, nor did the child’s **gender** (using one-way ANOVAs). |
| Swain et al. (2019) | Adapted group CBT for young children (aged 4-8) (‘STAMP’ programme) | **Intervention content adaptations**  - Creative outlets and activities  - Integration of emotion-focused strategies  **Involving wider support network**  - Parental involvement | The treatment responder and treatment non-responder groups did not significantly differ in **age**, **cognitive ability, autism symptomatology.** |
| White et al. (2015) | Bespoke individual and group CBT for anxiety (‘MASSI’ programme) | Not applicable. | Higher **verbal ability** predicted more anxiety improvement during treatment. Higher **severity of autism** predicted more severe anxiety pre-treatment and greater improvement post-treatment. The only significant predictor of change during the follow-up period was higher **trait anxiety in parents** at baseline, with children of more anxious parents experiencing less of an increase in anxiety post-treatment. |

*Note*. **CBT** = Cognitive Behavioural Therapy, **IQ** = Intelligence Quotient.
